# Multi-Omic Analyses Reveal Bidirectional Genetic Links and Convergent Inflammatory-Neuronal Signatures Between Frailty, Chronic Pain, and Rheumatoid Arthritis

**DOI:** 10.64898/2026.06.30.26356940

**Authors:** Jonny P Flint, Brittany L. Mitchell, Stephanie M. Heyworth, Hannah M. Smith, Isabelle F. Foote, Simon R. Cox, Michelle Luciano, Michelle K. Lupton

## Abstract

**Background:** Frailty reflects reduced physiological resilience and increased vulnerability to stressors, functioning as a key biological marker of ageing. Yet, the molecular and causal mechanisms linking frailty to chronic inflammatory conditions remain unclear.

**Methods:** We integrate Mendelian randomisation (MR), pairwise genome-wide association (PW-GWAS), and epigenetic-protein prediction to dissect bidirectional biological pathways between frailty, chronic pain (CP), and rheumatoid arthritis (RA).

**Results:** Across nine genetically defined frailty phenotypes- six domain-specific factors, a general frailty factor (GF), and two cumulative indices (Frailty Index – FI, Fried Frailty Score – FFS) – MR revealed widespread causal effects of CP and RA on frailty. Genetic liability to higher chronic pain robustly increased frailty severity across the cumulative frailty measures (FI and FFS) and the disability-related frailty domain (F6), with the strongest effect observed for the Frailty Index (β = 0.70, p = 7.9 × 10⁻⁵²), while cumulative frailty also elevated pain risk (β = 0.37, *p* = 2.6 × 10⁻¹³), supporting a reciprocal feedback model. RA exerted more selective effects, increasing cumulative frailty (FI and FFS) and disability-related frailty (F6), whereas the general frailty factor increased RA susceptibility (OR = 4.59, p = 9.5×10⁻⁴). Notably, the multimorbidity-related frailty domain showed an inverse effect on RA risk (OR = 0.65, p = 0.0078), suggesting immune adaptation or exhaustion within advanced frailty states. PW-GWAS and fine-mapping revealed shared causal loci linking inflammation, metabolism, and neurostructure – *SLC39A8, NLGN1, IL2RA, ERBB3*, and *MAGI3* – highlighting neuroimmune and synaptic processes as convergent ageing pathways. Epigenetic-proteomic analyses further identified two opposing axes: inflammatory and complement proteins (*CRP, C5, CCL18, STC1*) positively associated with frailty, versus neuronal adhesion and Extracellular Matrix (ECM) proteins (*NCAM1, CNTN4, NTRK3, ADAMTS13*) conferring resilience.

**Conclusion:** Together, these findings redefine frailty as a biologically dynamic interface between inflammation and structural maintenance, where opposing molecular pathways shape vulnerability and resilience. This integrative framework highlights frailty as both a biomarker and potential intervention target – linking inflammatory dysregulation to modifiable trajectories of functional ageing.

## Introduction

Frailty is a multidimensional clinical trait characterised by diminished physiological resilience and an increased susceptibility to stressors (^1^). Beyond its clinical relevance, as a measure to identify individuals at higher risk of both functional and cognitive decline (^2,3^), frailty has emerged as a biological marker of accelerated ageing, integrating multisystem decline and chronic low-grade inflammation (“inflammation”) (^4,5^). Despite its predictive value for morbidity and mortality, the genetic and biological mechanisms underpinning frailty remain incompletely understood. Although numerous biological pathways have been associated with frailty, it remains unclear whether these processes represent causes, consequences, or correlates of frailty (^6^). At present two principal frameworks dominate frailty measurement: the multidimensional Frailty Index (FI) (^7^),which quantifies the accumulation of health deficits across multiple physiological, psychological, and cognitive systems, including deficits such as cardiovascular disease, difficulty walking, memory problems, and depression; and the Fried Frailty Score (FFS) (^8^), which assesses weight loss, exhaustion, low physical activity, slow gait speed, and grip strength to capture physical frailty. Both capture facets of accelerated ageing but differ in scope and composition.

Recent advances in molecular genetics, particularly Genome-Wide Association Studies (GWAS), have enabled the identification of genetic variants associated with frailty risk and provided new insights into its underlying biological architecture (^9^). Genetic studies have recently elucidated frailty’s molecular architecture – highlighting its complexity. The first well-powered GWAS of Frailty, on FI, revealed loci linked to inflammatory and metabolic pathways (^9^), while a later GWAS of the FFS identified mitochondrial and neuromuscular components (^10^). However, these GWAS’ applied traditional univariate approaches conflating distinct biological processes embedded within broad frailty scores. To overcome this, genomic structural equation modelling (Genomic SEM) was applied to 30 frailty deficits, uncovering a general frailty factor of shared genetics between all deficits and six orthogonal genetically distinct domains, that reflect discrete clusters of frailty related pathways between deficits (^11^). This multivariate framework provides a new way to test multiple pathways of frailty and explore how these pathways are related to frailty related outcomes, including directionality of effect.

One example is whether chronic inflammation drives frailty or whether frailty amplifies inflammation, and/or how both interact bidirectionally. Cross-sectional and longitudinal data are inconsistent; with research showing elevated Interleukin 6 and C-reactive protein (CRP) preceding frailty onset, while others suggest that frailty status itself predicts later inflammatory activation (^12–14^). Mendelian randomisation (MR) studies have provided partial clarity but report conflicting conclusions. For example, a study found a causal effect of CRP on frailty, whereas proteome-wide MR analyses identified bidirectional effects mediated by metabolic and immune proteins (^15,16^). These discrepancies reinforce that frailty’s inflammatory correlates cannot be fully explained by single-pathway models. Such studies were conducted using broad frailty phenotypes, such as the Frailty Index, rather than genetically distinct frailty domains.

Two inflammation-related conditions – chronic pain (CP) and rheumatoid arthritis (RA) – exemplify the complex and bidirectional relationships observed between inflammatory pathology and frailty (^17–20^). Both syndromes are highly prevalent in ageing populations and show consistent epidemiological associations with frailty, yet their biological interconnections remain underexplored. Genome-wide association studies of CP and RA highlight shared molecular mechanisms: the GWAS of the chronic pain identified 39 risk loci implicated in neuroimmune and nervous system signalling pathways (^21^), while the GWAS of RA identified 101 risk loci enriched for cytokine, T-cell, and broader immune signalling pathways (^22^).

Together, these findings suggest overlapping inflammatory-ageing mechanisms linking frailty, CP, and Rebuilding on this, recent two-sample Mendelian randomisation studies have specifically examined the causal relationships between CP, RA, and frailty, although all have relied on aggregate measures of frailty. Two MR analyses reported bidirectional effects between chronic pain and frailty, with genetic liability to pain increasing frailty risk and, conversely, frailty liability modestly increasing chronic pain susceptibility (^23,24^). Similarly, MR analyses of RA and frailty have shown that genetically predicted RA increases frailty risk, partially mediated through circulating inflammatory cytokines (^25^), while broader autoimmune liability also elevates frailty susceptibility (^26^). Yet, because these studies operationalise frailty as an aggregate construct, they may obscure domain-specific biological processes-for example, neuroinflammatory versus metabolic pathways-that differentially contribute to frailty in the context of chronic inflammatory disease. Consequently, the direction and magnitude of the frailty-inflammation relationship remains unresolved, warranting analyses that integrate both latent-domain and cumulative measures to clarify whether these connections reflect shared biology, causal feedback, or distinct but parallel processes of ageing. Though clearly, the direction and magnitude of these associations remain underexplored – some studies infer frailty as a downstream effect of inflammatory disease (^27,28^), while others indicate that higher frailty predisposes individuals to immune dysregulation and chronic pain (^29,30^). Our recent work demonstrated that polygenic liability to chronic pain increased frailty and was threefold predictive of frailty status compared to polygenic liability for frailty itself, while genetic predisposition to rheumatoid arthritis predicted lower frailty status (^31^). These opposing effects illustrate frailty’s polygenic and pleiotropic complexity, where multiple interacting pathways modulate vulnerability in ageing.

To determine whether the observed associations reflect shared biological mechanisms, bidirectional causal relationships, or distinct but parallel ageing processes, we implemented an integrative five-step framework combining causal inference, genomic architecture mapping, and longitudinal molecular profiling. We first used two-sample Mendelian randomisation to test bidirectional causal relationships between nine genetically defined frailty phenotypes (six latent frailty subdomains, a general frailty latent factor, the Frailty Index, and the Fried Frailty Score) and two inflammation-related outcomes (chronic pain and rheumatoid arthritis). To dissect the underlying mechanisms, pairwise GWAS, fine-mapping, and colocalisation analyses were then applied to distinguish shared from distinct causal variants across frailty domains. We next examined 84 validated protein-based EpiScores in two independent ageing cohorts to identify circulating molecular signatures associated with frailty both cross-sectionally and across time. Building on this, we assessed whether polygenic liabilities for frailty, chronic pain, and RA translated into measurable proteomic profiles through integrated PRS-EpiScore networks. Finally, longitudinal growth models identified proteomic trajectories predictive of frailty progression. Collectively, this study provides an integrated, multi-omic framework to position frailty as a biological consequence, as well as a precursor and parallel expression of chronic inflammatory processes in ageing.

## Methods

### GWAS Datasets

We performed two-sample Mendelian randomisation (MR) to test causal relationships between nine frailty phenotypes: six latent frailty subdomains identified using Genomic Structural Equation Modelling (Genomic SEM) – F1 (limited social support), F2 (unhealthy lifestyle), F3 (multimorbidity), F4 (metabolic problems), F5 (poorer cognition), and F6 (disability) – together with a latent general frailty factor (GF), representing genetic influences shared across all frailty deficits (Figure 1) (^11^), the aggregate Frailty Index (FI) (^9^), and the Fried Frailty Score (FFS) (^10^), and two outcomes: chronic pain (CP) (^21^) and rheumatoid arthritis (RA)(^22^).

**Figure 1.**
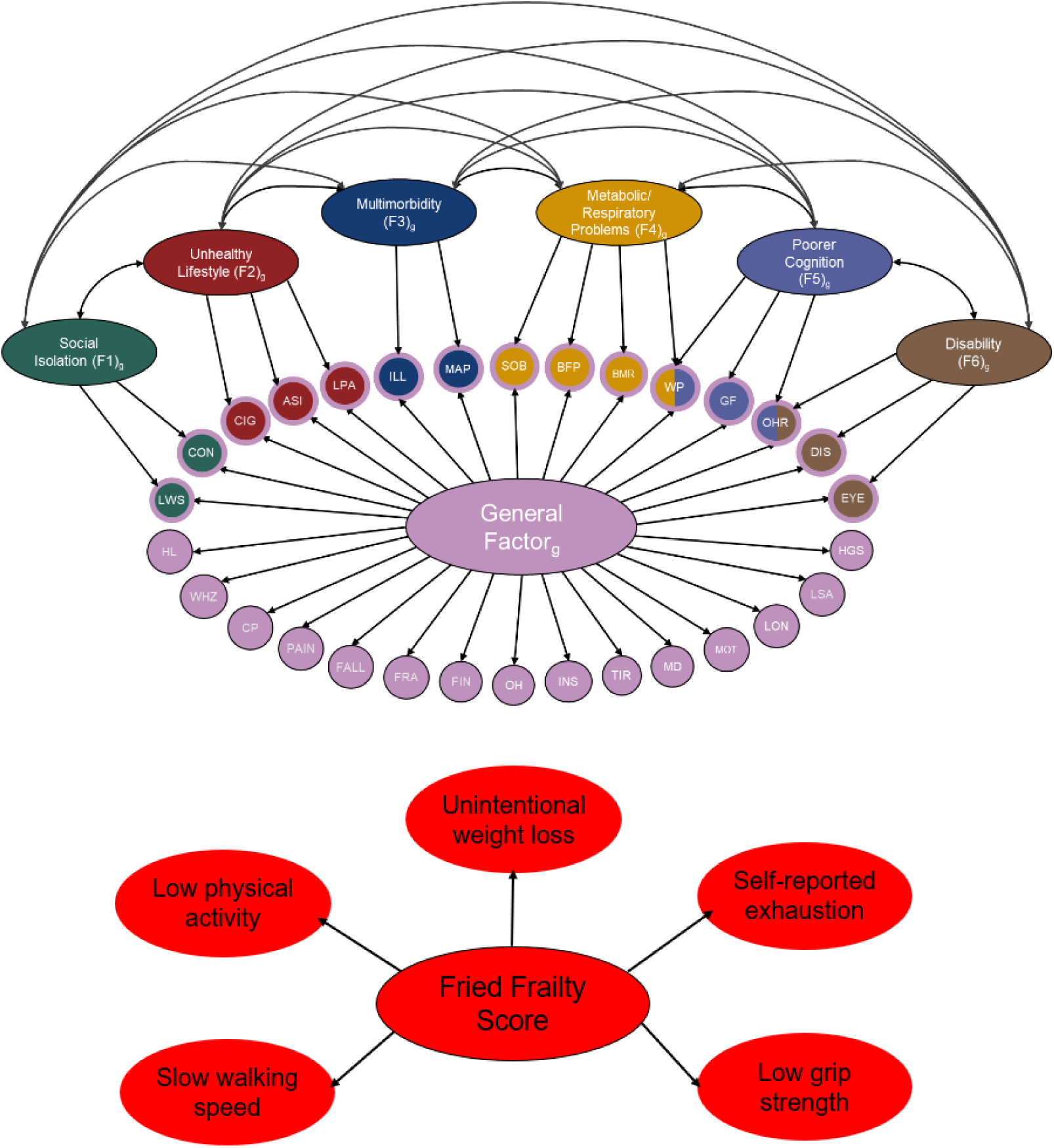
Panel A shows a path diagram of the multivariate frailty model (adapted from Foote et al., 2025), in which a general frailty factor (GF)-conceptually analogous to a frailty index-is modelled alongside six orthogonal domain-specific factors (F1-F6). The diagram is presented without factor loadings to emphasise the overall structure. The general factor captures shared variance across 30 frailty indicators, while domain-specific factors represent distinct components of frailty. Indicator abbreviations are as follows: ASI, pulse wave arterial stiffness index; BFP, body fat percentage; BMR, basal metabolic rate; CIG, cigarettes per day; CON, unable to confide; CP, chest pain; DIS, long-standing illness, disability or infirmity; EYE, eye disorder; FALL, falls in past year; FIN, financial difficulties; FRA, fracture; GF, low fluid intelligence; HGS, low hand grip strength; HL, hearing loss; ILL, number of non-cancer illnesses; INS, insomnia; LON, loneliness; LPA, physical inactivity; LSA, low social/leisure activity; LWS, not living with partner; MAP, mean arterial pressure; MDD, major depressive disorder; MOT, low motivation; OH, poor oral health; OHR, poor overall health rating; PAIN, pain; SOB, shortness of breath; TIR, tiredness; WHZ, wheeze; WP, slow walking pace. Panel B shows a path diagram of the Fried Frailty Score phenotype, defined by five components: unintentional weight loss, self-reported exhaustion, low physical activity, slow walking speed, and low grip strength.

Each dataset is described in Table 1, with extensive details on phenotypes and participants in Supplementary Table 1. In brief, all analyses used publicly available European-ancestry GWAS summary statistics harmonised to the GRCh37/hg19 build. The frailty phenotypes and chronic pain were analysed as quantitative traits, whereas rheumatoid arthritis was analysed as a binary case-control trait. The latent frailty measures (F1-F6 and GF) were derived from a previously published multivariate GWAS that had used Genomic Structural Equation Modelling (Genomics SEM) to estimate the factor model (^11^). The FI GWAS measure, somewhat akin to the orthogonal General Frailty factor from the multivariate GWAS, was a 49-item accumulation-of-deficits approach previously applied in UK Biobank (^9^). Items spanned physical, clinical, and psychological domains, and were coded as deficits to produce a proportional FI score. On the other hand, the FFS classifies individuals according to five criteria – unintentional weight loss, exhaustion, low physical activity, slow walking speed, and weak grip strength – yielding a score from 0 to 5. Although often analysed as a binary phenotype (frail ≥3), the FFS in the GWAS was treated as an ordinal variable to maximise statistical power (^10^). Figure 1 demonstrates the structure of the Genomic SEM frailty model and the aggregate FFS measure.

**Table 1.** All genome-wide association studies (GWAS) used across analyses. For frailty, alongside cumulative measures, we include the general latent factor and six orthogonal residual factors (F1-F6), each representing distinct domains – sample sizes reflect effective N (N ^) as estimated by Genomic SEM. UK Biobank was the primary source of data for most frailty indicators, but several traits included data from large consortia.

| Trait / GWAS | Phenotype | Sample Size | Cohorts | Reference |
| --- | --- | --- | --- | --- |
| GF | General factor across 30 indicators | $\hat{N} = 2,105,711$ | UK Biobank + Consortia | Foote et al. (2025) |
| F1 | Limited social support | $\hat{N} = 385,176$ | | |
| F2 | Unhealthy lifestyle | $\hat{N} = 559,200$ | | |
| F3 | Multimorbidity | $\hat{N} = 755,865$ | | |
| F4 | Metabolic problems | $\hat{N} = 377,810$ | | |
| F5 | Poorer cognition | $\hat{N} = 327,699$ | | |
| F6 | Disability | $\hat{N} = 699,030$ | | |
| Frailty Index | Accumulation of health deficits | $N = 164,610$ | UK Biobank | Atkins et al. (2021) |
| Fried Frailty Score | Physical frailty phenotype | $N = 386,565$ | UK Biobank | Ye et al. (2023) |
| Chronic pain | Self-reported chronic pain | $N = 380,089$ | UK Biobank | Johnston et al. (2019) |
| Rheumatoid arthritis | Clinical diagnosis | $N = 14,361$ cases / $43,923$ controls | EIRA, NARAC, WTCCC | Okada et al. (2014) |

### Mendelian Randomisation (MR) Analyses

#### Instrument selection and pre-processing

Mendelian randomisation (MR) leverages genetic variants as instrumental variables to infer causal relationships between exposures and outcomes under three core assumptions: that instruments are robustly associated with the exposure (relevance), influence the outcome only through the exposure (exclusion restriction), and are independent of exposure-outcome confounders (independence) (^32^). All analyses were conducted in accordance with the STROBE-MR reporting guidelines (^33^).

For each exposure, genome-wide significant single-nucleotide polymorphisms (SNPs; P < 5 × 10⁻⁸) were selected as instruments and clumped for linkage disequilibrium using PLINK v1.9 with a 1000 Genomes European reference panel (r² < 0.001, 10 Mb window). SNPs were retained only if present in both the LD reference and outcome datasets, and variants failing allele matching were excluded. Instrument strength was assessed using F-statistics (F = β²/SE²), with all retained instruments exceeding the conventional threshold for strong instruments (F > 10), indicating no evidence of weak instrument bias. Across all analyses, between 2 and 202 instruments were retained, and the minimum instrument F-statistic in every analysis exceeded 22.38, indicating consistently strong instruments (Supplementary Table 2).

#### Harmonisation and directionality filtering

Exposure and outcome datasets were harmonised using the TwoSampleMR framework (^33^), including strand alignment, removal of palindromic variants with intermediate allele frequencies, and exclusion of SNPs that could not be unambiguously aligned. Steiger directionality testing was applied to retain variants explaining more variance in the exposure than in the outcome, thereby reducing the risk of reverse causation. This procedure was implemented for both forward and reverse analyses, resulting in high SNP retention for forward models and more stringent filtering in the reverse direction.

#### MR estimation and sensitivity analyses

Primary causal estimates were obtained using inverse-variance weighted (IVW) MR, with MR-Egger, weighted median, simple mode, and weighted mode estimators included as sensitivity analyses. Effect estimates were reported with standard errors and P-values, and IVW results were corrected for multiple testing using the Benjamini-Hochberg false discovery rate (FDR). Sensitivity analyses included Cochran’s Q statistic to assess heterogeneity, the MR-Egger intercept test to evaluate directional horizontal pleiotropy, leave-one-out analyses to identify influential instruments, and MR-PRESSO to detect potential outlier variants where applicable (^34^). All analyses were conducted in R (version 4.3.1) using the TwoSampleMR and MR-PRESSO packages (^34–37^).

### Pairwise GWAS(PW-GWAS), Fine Mapping and Colocalisation

We applied pairwise GWAS (PW-GWAS; https://github.com/joepickrell/gwas-pw, (^38^)) to identify genomic regions jointly associated with each frailty phenotype and either CP or RA, and to assess whether overlaps were due to shared or distinct causal variants. PW-GWAS models two GWAS jointly across 1,703 independent LD blocks and estimates posterior probabilities (PPAs) for four mutually exclusive models: Model 1: association with frailty only, Model 2: association with CP or RA only, Model 3: association with both Frailty and CP or Frailty and RA due to a shared causal variant, Model 4: association with Frailty and CP or Frailty and RA via distinct causal variants. Genome-wide patterns of shared architecture were visualised using the Ritchie Lab PhenoGram tool (http://visualization.ritchielab.org/phenograms/plot). A suggestive threshold of PPA₃ ≥ 0.50 was used to highlight broad patterns of overlap. A stringent PPA₃ ≥ 0.80 threshold identified high-confidence shared regions that progressed to fine-mapping and colocalisation. Although PPA thresholds are inherently arbitrary, this strict criterion ensured that only credible shared blocks were retained for downstream analyses, as seen in other PW-GWAS studies (^39^). Blocks overlapping the extended MHC (chr6: 25-35 Mb) regions were excluded due to complex LD structure.

Secondly, for selected blocks (PPA₃ ≥ 0.80) under PW-GWAS Model 3 we performed Bayesian fine-mapping, using BEDtools (^40^), to obtain 90% credible sets of shared causal variants. Fine-mapping summaries included: genomic span and number of variants in each credible set; and, maximum variant-level posterior probability of being the shared causal variant (variant-level PPA₃). Variants were mapped to nearby transcripts using gene annotation files, and genes were prioritised based on the highest variant-level PPA₃ among their assigned variants. These summaries informed downstream block- and gene-level visualisations. Thirdly, to thoroughly distinguish true shared causal variants from distinct signals, we performed variant-level colocalisation (coloc v5.2.3, (^41^)) for all blocks with PPA₃ ≥ 0.90 (shared) or PPA₄ ≥ 0.90 (distinct). Analyses used ±250 kb windows around PW-GWAS block boundaries. Quality Control was conducted to ensure post allele harmonisation, and filtering to MAF ≥ 1% and p < 0.05. Consistent with the underlying GWAS designs, quantitative trait priors were used for chronic pain and the frailty phenotypes, whereas rheumatoid arthritis was modelled as a case-control trait using the reported case proportion from Okada et al. (2014). Opposite to PW-GWAS, in posterior probabilities PP.H₄ (shared variant) and PP.H₃ (distinct variants) are the assigned signals from the colocalisation output.

#### Epi-Protein-Score Analyses Cohorts

Participants were drawn from the Lothian Birth Cohort 1936 (LBC1936), a longitudinal study of community-dwelling Scottish older adults born in 1936, of which most took a general cognitive ability test at ∼11 years old in Scotland and were subsequently followed up at age ∼70 (^42^). Once restricted to participants with complete frailty, methylation, polygenic and covariate data, the analytical sample consisted of 895 individuals at Wave 1 (453 female, 442 male, age ∼70 range 67.61-71.30), 792 at Wave 2 (417 female, 375 male, age ∼73 range 70.91-74.15), 611 at Wave 3 (319 female, 292 male, age ∼76 range 74.68-77.70), and 499 at Wave 4 (262 female, 237 male, age ∼79, range 77.96 - 80.87). Frailty increased across waves, with mean Frailty Index (FI) values of 0.16 (range 0-0.49) at Wave 1, 0.18 (range 0-0.55) at Wave 2, 0.20 (range 0.02-0.65) at Wave 3, and 0.21 (range 0.03 - 0.63) at Wave 4. The same analytic framework was then applied to an independent sample of 339 Australian adults from the Prospective Imaging Study of Ageing (PISA) (^43^), aged 42-78 years (mean = 58.9), measured at one time-point. PISA participants’ FI ranged from 0.00 to 0.45 (mean = 0.11), and the sample comprised 104 males and 235 females. Full details for cohorts, methylation and genomic data construction can be found in Supplementary Materials.

#### Frailty Index construction

A Frailty Index (FI) was constructed separately within the independent cohorts (LBC1936, PISA), drawing on a set of deficits spanning physical, biological, social, psychological, and cognitive domains.

Deficits were coded as binary (0 = absent, 1 = present), with an intermediate value of 0.5 used where a deficit was partially present, or as continuous measures scaled between 0 and 1. For each participant, the FI was calculated as the sum of all deficit values divided by the total number of deficits assessed, yielding a proportional score ranging from 0 to 1. Higher FI values indicate greater frailty. The full list of deficits used in LBC1936 and PISA are in Supplementary Tables 3 and 4, respectively.

#### Proteins and EpiScores

The training and validation of the 84 protein-based EpiScores used in this study have been reported previously (^44^). In brief, each EpiScore was generated using a separate penalised regression model that identified a weighted combination of CpG sites optimally predicting circulating levels of its target protein. All 84 EpiScores satisfied the predefined performance criteria (Pearson r > 0.1 and p < 0.05) when evaluated in an independent subset of the Generation Scotland cohort (STRADL; N = 778) by comparing predicted and measured protein concentrations. For each EpiScore, the accompanying annotation table reports the platform-specific assay identifier, the proteomic measurement panel used, the HGNC gene symbol encoding the protein, the intended protein target of the assay (the protein targeted by the aptamer or Olink antibody), the UniProt accession ID, and the UniProt-recommended full protein name (a descriptive biological name of the protein encoded by the gene, as previously described (^44,45^).

These EpiScores were subsequently computed from DNAm beta values in the LBC cohorts (LBC1936: n = 895, followed by adjustment for technical factors using linear regression. DNAm processing procedures are described in the Supplementary Methods. In the present study, LBC1936 EpiScores were projected into Wave 1, 2, 3 and 4 and were corrected for set, array, and hybridisation date at each wave. Residuals from these models were then extracted and carried forward for all downstream analyses. In PISA, the same 84 EpiScores were projected into DNAm data and corrected for batch effects using the equivalent set of technical covariates. A full list of the 84 EpiScores can be found in Supplementary Table 5.

#### Polygenic Scores

PRSs for RA, CP, GF, F1-F6, FI, and FFS, from the same GWAS used for MR (Table 1), were generated using SBayesR with recommended defaults. SBayesR jointly estimates SNP effects under mixture-normal priors and incorporates genome-wide LD structure (^46^). PRSs were standardised within cohort/wave.

#### Statistical Modelling

For PRS-protein and PRS-EpiScore associations regression models included age, sex, and the first four genetic ancestry principal components. Linear models were fitted separately for each protein/EpiScore, excluding models with <50 observations. Coefficients and p-values were extracted, and false discovery rate (FDR) correction was applied across all protein (or EpiScore) associations for each PRS. Frailty index was regressed on individual EpiScores in LBC1936 and PISA, adjusting for age and sex. EpiScores passing FDR < 0.05 were classified as harmful (β > 0) or protective (β < 0).

#### Network Analyses

Directed PRS-protein-frailty networks were constructed for each wave using significant PRS-protein and protein-frailty edges. Edge weights corresponded to regression coefficients. Network layouts were generated using force-directed algorithms in igraph(^47^). PISA networks used PRS-EpiScore-frailty relationships. Two additional LBC1936 networks were generated: Protein-protein correlation networks (|r| > 0.30); and, combined protein + FI networks linking significant Epiproteins directly to FI. Harmful/protective biomarkers were colour-coded with blue indicating protective direct.

#### Longitudinal Modelling of Protein and Frailty Trajectories

Proteomic trajectories (Waves 1-4) and frailty trajectories (Waves 1-5) were modelled using linear mixed-effects growth models of the form protein ∼ age + sex + (1 + age | ID) and FI ∼ age + sex + (1 + age | ID). Participants contributed a total of 3,635 observations (N = 1,091 individuals) across 5 waves of data (mean = 3 waves). Baseline ages ranged from 67.6 to 83.1 years (mean = 74.5, range 67.61 - 83.12), and frailty index (FI) scores ranged from 0 to 0. 65. Frailty progression showed meaningful inter-individual variability (FI slope: mean = 0.076, SD = 0.027; range = −0.053 to 0.278). Protein-specific slopes were extracted for 84 proteins with sufficient repeated measurements; 22 proteins exhibited minimal between-person slope variability (SD < 0.01) and were excluded, leaving 62 proteins for analysis, a list of the remaining proteins is available in Supplementary Table 6. Empirical Bayes estimates of random intercepts and slopes were combined with baseline covariates (age, sex), and all slope terms were standardised within protein. Longitudinal associations were then tested using the model FI slope ∼ protein slope + protein intercept + baseline age + sex, with false-discovery rate (FDR) correction applied across proteins. Proteins were classified as “harmful” or “protective” based on the sign of the regression coefficient (β) for protein slope, where positive β values (steeper increases in protein levels) predicted faster frailty progression and negative β values predicted slower progression.

## Results

### Bidirectional Mendelian Randomisation

Primary MR results (Figure 2) are presented following exclusion of the extended major histocompatibility complex (MHC) region (chr6:25-35 Mb). Analyses demonstrated that excluding the MHC region yielded highly similar results to analyses including the MHC region (Supplementary Tables 7-8), with the overall pattern of bidirectional associations remaining unchanged. The principal differences were attenuation of the FI on RA and F6 on RA associations, suggesting partial influence of MHC-region variants. Leave-one-out analysis identified rs9275160 as the influential variant underlying the original FI on RA association.

**Figure 2.**
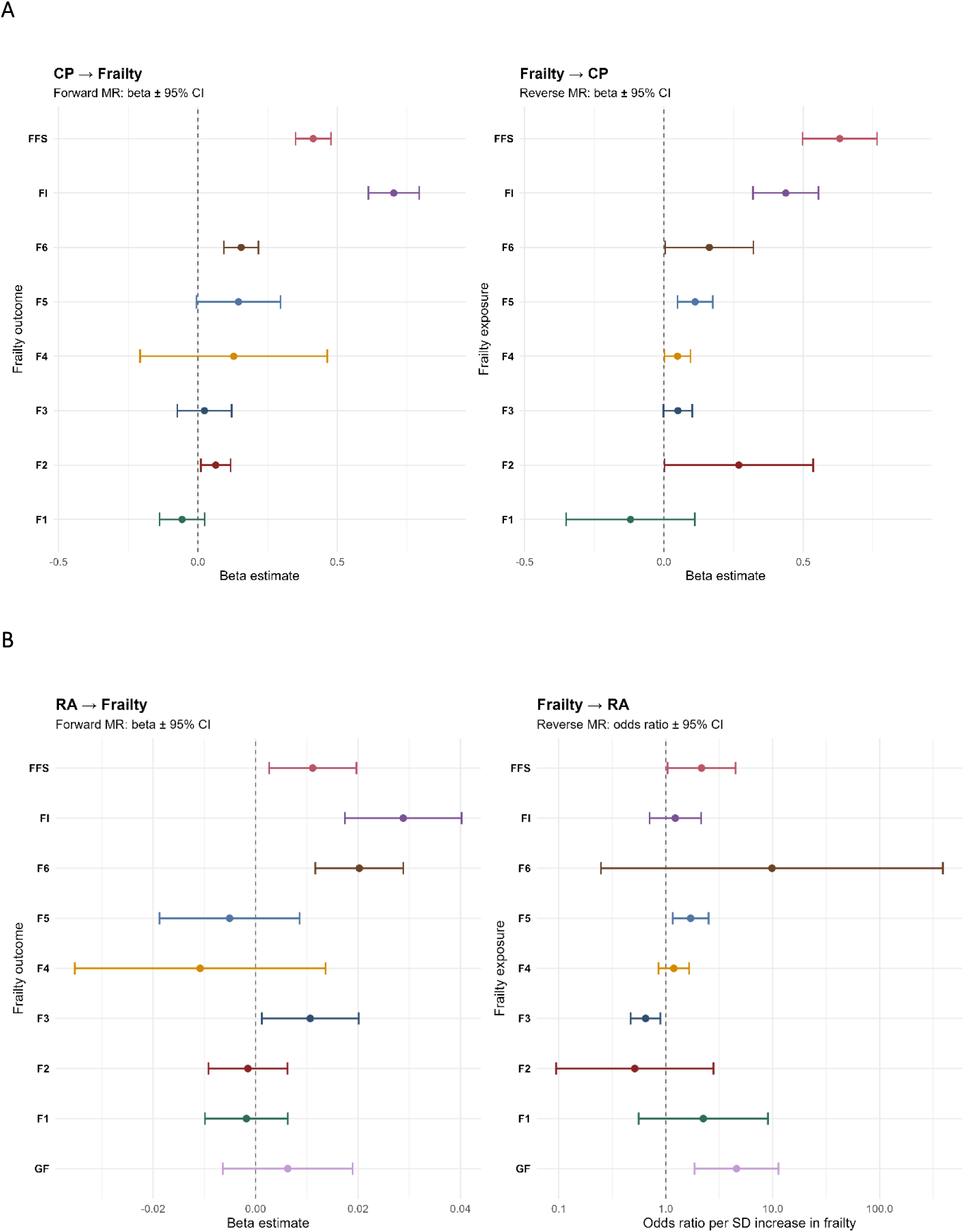
Bidirectional Mendelian randomisation between frailty, chronic pain, and rheumatoid arthritis. Forest plots show inverse-variance weighted (IVW) causal effect estimates (β) with 95% confidence intervals. Panel A displays bidirectional MR results for frailty measures and chronic pain (CP), and Panel B for frailty measures and rheumatoid arthritis (RA), following FDR. Positive β values indicate higher frailty or disease liability, whereas negative β values indicate protective associations. Frailty measures comprise the General Frailty factor (GF), six Genomic SEM frailty domains – F1 (limited social support), F2 (unhealthy lifestyle), F3 (multimorbidity), F4 (metabolic problems), F5 (poorer cognition), and F6 (disability) – together with the Frailty Index (FI), representing accumulation of health deficits, and the Fried Frailty Score (FFS), representing the Fried Frailty Score.

In the forward direction, MR analyses demonstrated that genetic liability to chronic pain (CP) and rheumatoid arthritis (RA) exerted widespread causal effects on multiple frailty constructs (Figure 2, Panel A). CP showed compelling evidence of increasing frailty severity across all global measures. For the Fried Frailty Score (FFS), the IVW analysis indicated a strong positive association of CP on FFS (IVW β = 0.41, 95% CI: 0.35-0.48, p = 1.7×10⁻³⁷, FDR = 7.5×10⁻³⁶) with consistent support from MR-Egger (β = 0.73, p = 3.0×10⁻^55^) and the weighted median estimator (β = 0.35, p = 1.0×10⁻²³). Similarly robust effects were observed for the Frailty Index (IVW β = 0.70, 95% CI: 0.61-0.79, p = 7.9×10⁻⁵², FDR = 4.6×10⁻⁵⁰). CP also increased domain-specific frailty factors, including disability-related frailty, F6 (F6; IVW β = 0.16, p = 8.2×10⁻⁷, FDR = 9.0×10⁻⁶. Following FDR correction, no evidence was observed for limited social support-related frailty (F1), unhealthy lifestyle-related frailty (F2), multimorbidity frailty (F3), metabolic frailty (F4), or poorer-cognition-related frailty (F5).

Reverse MR analyses for CP revealed a broader pattern of associations when frailty constructs were modelled as exposures (Figure 2, Panel A). Strong evidence was observed for global frailty measures, with both the Fried frailty score (IVW β = 0.63, 95% CI: 0.50-0.76, p = 1.1×10⁻²¹, FDR = 1.7×10⁻²⁰) and the Frailty Index (IVW β = 0.37, 95% CI: 0.27-0.47, p = 2.6×10⁻¹³, FDR = 3.2×10⁻¹²) showing robust positive effects on CP, supported by consistent estimates across sensitivity analyses. No convincing evidence was observed for the domain-specific frailty factors following FDR correction. Associations with general frailty (GF) and CP are presented in the Supplementary Results (Supplementary Tables 7 and 8), as this construct includes pain-related indicators and is therefore not fully independent of chronic pain.

RA (Figure 2, Panel B) also demonstrated reliable forward-direction effects on global frailty measures. RA on frailty effects is reported as beta coefficients, while frailty on RA effects are reported as odds ratios, reflecting the continuous versus binary nature of the outcomes. For the Fried Frailty Score (IVW β = 0.011, 95% CI: 0.003-0.020, p = 0.010, FDR = 0.041), RA showed a small but significant positive association, with concordant estimates from MR-Egger (β = 0.022, p = 0.030) and support from the weighted median estimator (β = 0.014, p = 5.0×10⁻⁴). Similarly robust effects were observed for the RA on Frailty Index on (IVW β = 0.029, 95% CI: 0.017-0.040, p = 6.9×10⁻⁷, FDR = 8.1×10⁻⁶), with concordant estimates from MR-Egger (β = 0.035, p = 0.014) and the weighted median estimator (β = 0.029, p = 1.5×10⁻⁵). RA also showed domain-specific effects, increasing disability-related frailty (IVW β = 0.020, 95% CI: 0.012-0.029, p = 3.7×10⁻⁶, FDR = 3.8×10⁻⁵) whereas no convincing evidence was observed for general frailty (GF), limited social support-related frailty (F1), unhealthy lifestyle-related frailty (F2), multimorbidity-related frailty (F3), metabolic-related frailty (F4), or poorer-cognition-related frailty (F5).

Reverse MR analyses revealed a more selective pattern of associations when frailty constructs were modelled as exposures and RA as the outcome. The General Frailty factor demonstrated strong evidence of increasing RA liability (IVW OR = 4.59, 95% CI: 1.86-11.32, p = 9.5×10⁻⁴, FDR = 0.0050), with a consistent estimate from the weighted median approach (OR = 4.63, p = 0.015). In contrast, multimorbidity frailty (F3) was inversely associated with RA risk (IVW OR = 0.65, 95% CI: 0.47-0.89, p = 0.0078, FDR = 0.033), with replication using the weighted median estimator (OR = 0.47, p = 3.3×10⁻⁴). Other frailty domains (F1, F2, F4, F5, F6), as well as FI and FFS, showed no convincing evidence of causal effects on RA, with effect estimates generally close to the null and wide confidence intervals across sensitivity analyses.

Collectively, bidirectional analyses showed that genetic liability to CP and RA exerted consistent causal effects on global frailty measures, with effects present on latent domains of frailty such as disability-related frailty (F6). In contrast, reverse effects of frailty on CP and RA were more selective, with robust effects of the Frailty Index (FI) and Fried Frailty Score (FFS) on CP, and significant associations of the General Frailty factor (GF) and multimorbidity frailty (F3) with RA. FDR correction was applied to the IVW analyses, while sensitivity analyses (MR-Egger, weighted median, leave-one-out and MR-PRESSO), together with Cochran’s Q heterogeneity tests, supported the robustness of the findings (Supplementary Tables 9-14, leave-one-out plots, Supplementary Materials: Supplementary Figures 1-70).

### GWAS Pairwise, Fine-Mapping and Colocalisation – Chronic Pain

All PW-GWAS findings for Frailty-Chronic Pain and Frailty-Rheumatoid Arthritis are in Supplementary Tables 15 and 16, respectively.

Figure 3 summarises LD blocks in which chronic pain and each frailty phenotype showed evidence of shared or distinct genetic architecture, defined by PPA₃ > 0.50 (shared causal variant) and PPA₄ > 0.50 (distinct causal variants). Narrower frailty traits showed modest overlap with CP, including F1 (4 shared, 2 distinct), F2 (4, 1), F3 (24, 30), and F4 (10, 9) showed modest overlap with CP, whereas F5 (24, 20) and F6 (12, 1) demonstrated stronger overlap. Global measures exhibited the strongest overlap, with FFS (97, 1), FI (118, 0), and GF (137, 0) blocks meeting the shared threshold and none exceeding the distinct threshold for FI and GF.

**Figure 3.**
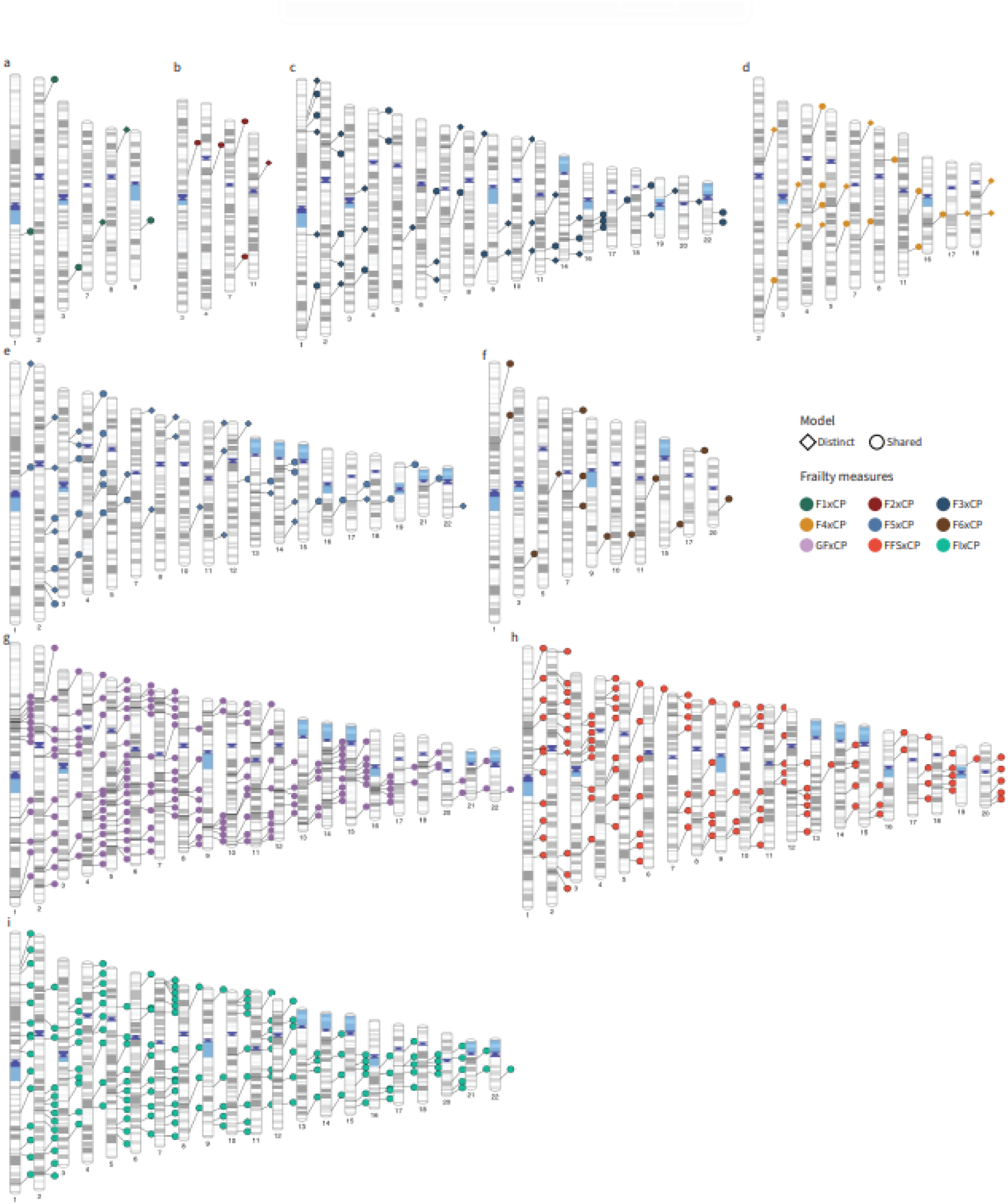
Ideogram of LD blocks identified as having evidence of shared genetic architecture between frailty and chronic pain, based on a posterior probability (PPA) > 50% for either model 3 (PPA₃ > 0.5), which assumes a single causal variant influencing both traits, or model 4 (PPA₄ > 0.5), which assumes separate causal variants for each trait within the LD block. Panels a-i correspond to F1 × CP (limited social support-related frailty), F2 × CP (unhealthy lifestyle-related frailty), F3 × CP (multimorbidity-related frailty), F4 × CP (metabolic-related frailty), F5 × CP (poorer cognition-related frailty), F6 × CP (disability-related frailty), GF × CP (General Frailty), FFS × CP (Fried Frailty Score), and FI × CP (Frailty Index), respectively.

**Figure 4.**
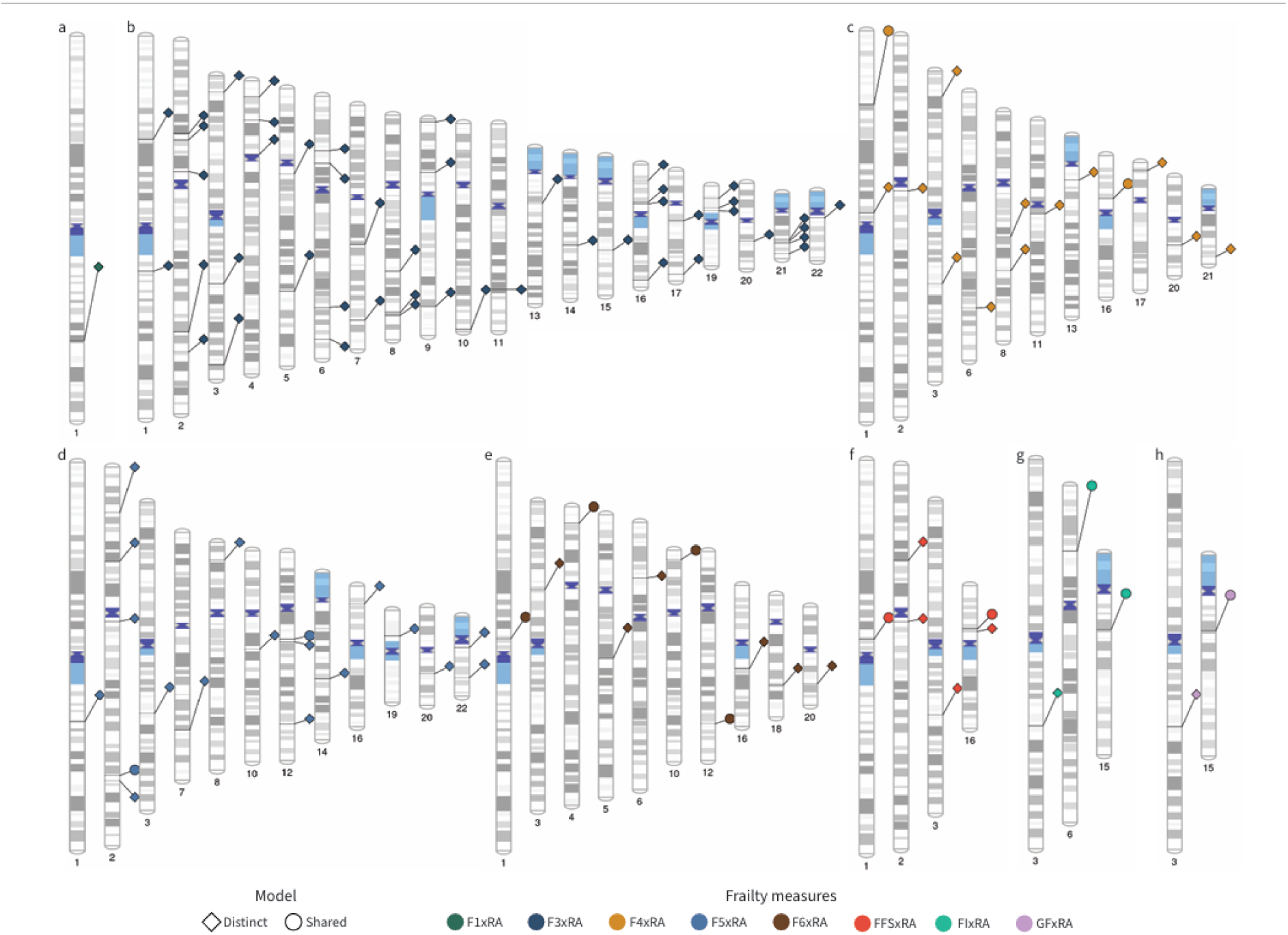
Ideogram of LD blocks identified as having evidence of shared genetic architecture between frailty and rheumatoid arthritis, based on a posterior probability (PPA) > 50% for either model 3 (PPA₃ > 0.5), which assumes a single causal variant influencing both traits, or model 4 (PPA₄ > 0.5), which assumes separate causal variants for each trait within the LD block. Panels a-h corresponds to F1 × RA (limited social support-related frailty), F3 × RA (multimorbidity-related frailty), F4 × RA (metabolic-related frailty), F5 × RA (poorer cognition-related frailty), F6 × RA (disability-related frailty), FFS × RA (Fried Frailty Score), FI × RA (Frailty Index), and GF × RA (General Frailty), respectively.

Building on these ideogram-level patterns, we next focused on regions showing the strongest evidence of shared genetic architecture (P₃ ≥ 0.80), to identify the specific LD blocks most likely to harbour true shared causal variants. For Frailty and Chronic Pain, all frailty measures yielded at least one LD block with qualified evidence for a shared causal signal (PPA₃ > 0.5).

Plots displaying the 90% credible sets for each shared block were generated, showing the number of variants per set alongside their variant-level PPA₃ values. Additional plots depicting the genes nearest to these credible-set variants, summarising the maximum variant-level PPA₃ assigned to each gene, were also produced for every frailty-CP pairing and are provided in the Supplementary Materials (Supplementary Figures 71-77), with corresponding gene-level plots shown in Figures 5 and 6.

**Figure 5.**
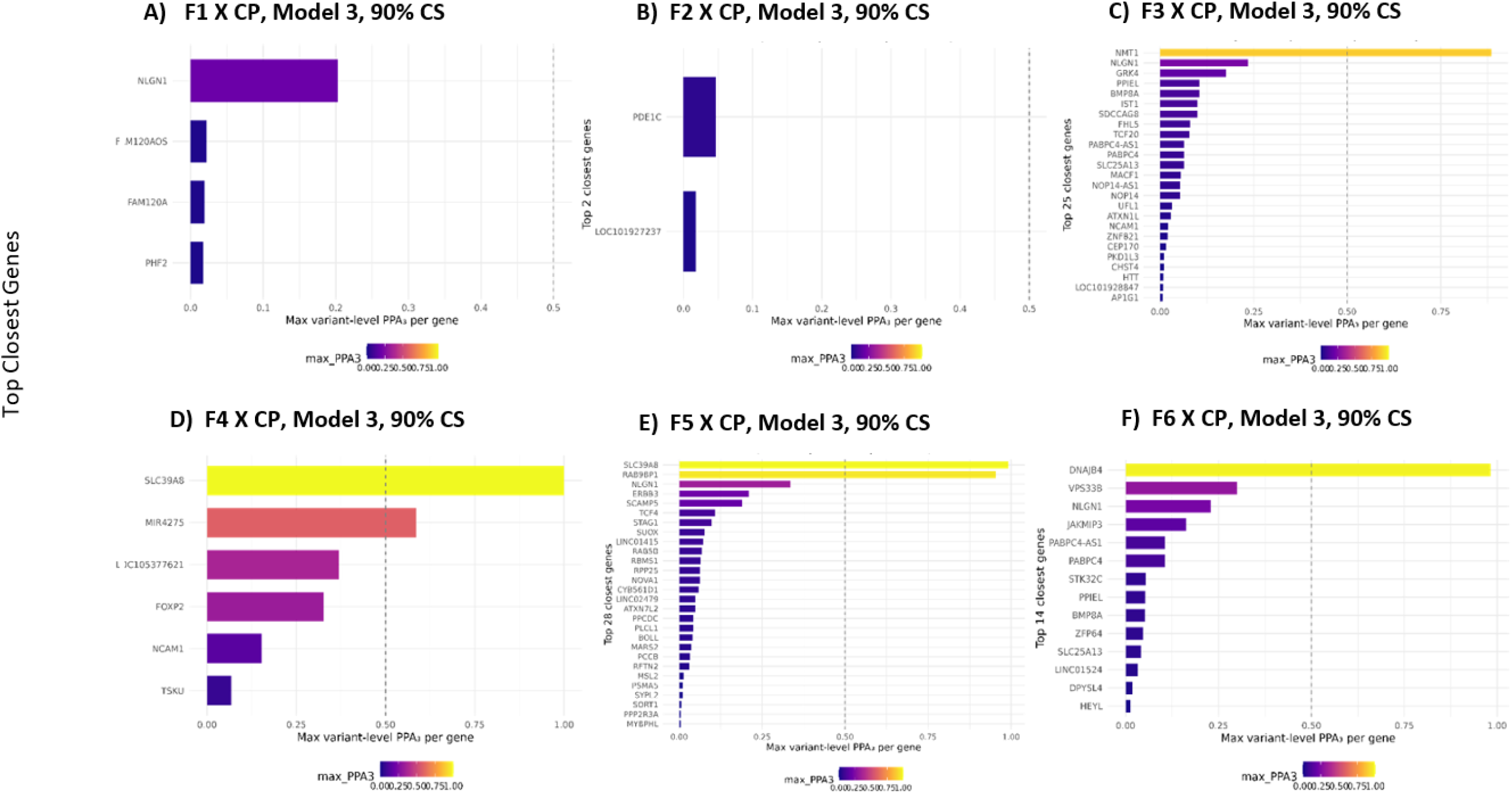
Panels A-F show the gene mapped plots, mapped by closest-gene annotation, summarised by their maximum variant level PPA.3 – each plot is one of six frailty domains (F1-F6) – limited social support, unhealthy lifestyle, multimorbidity, metabolic problems, poorer cognition, and disability-related frailty and chronic pain.

**Figure 6.**
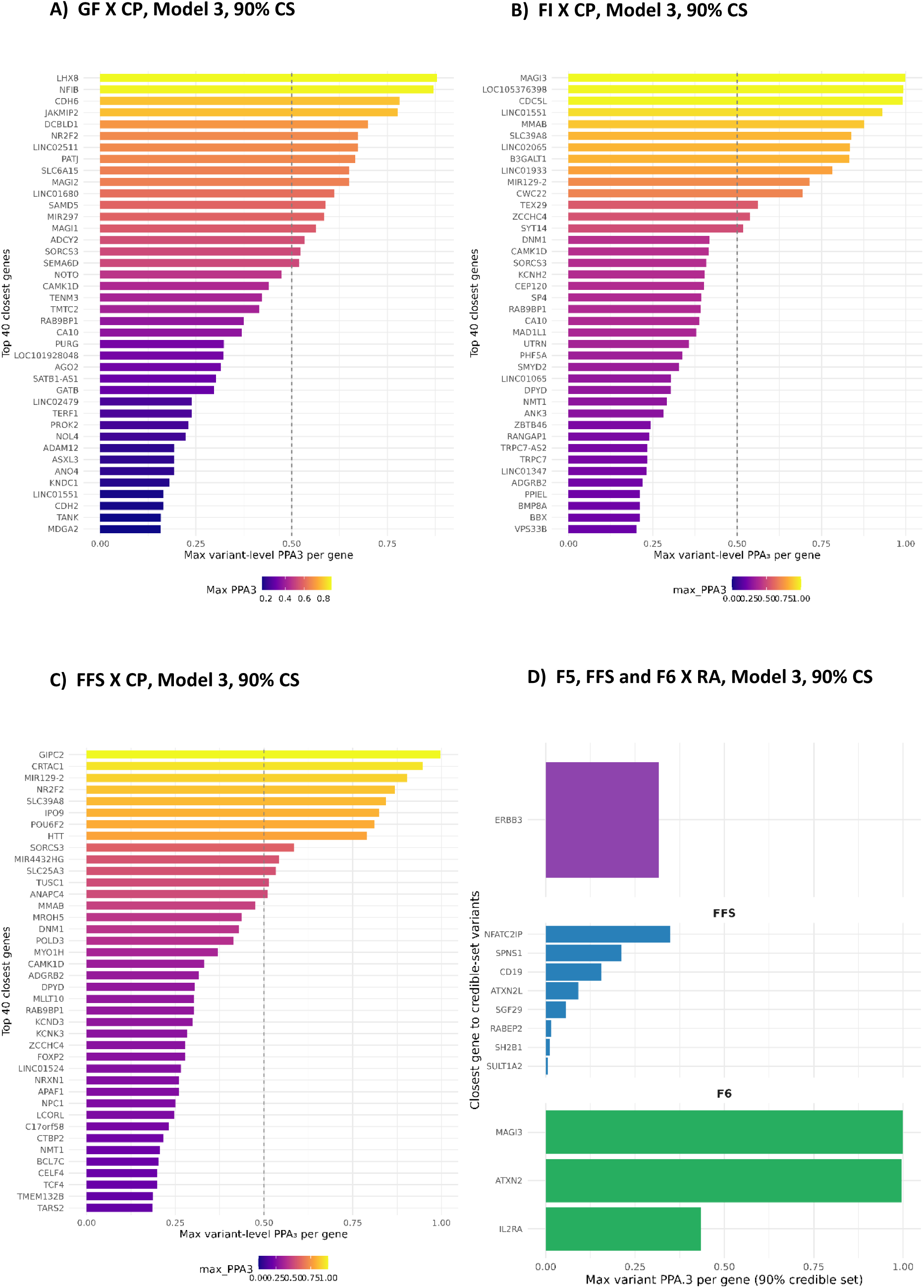
Panels A-D show the gene mapped plots, mapped by closest-gene annotation, summarised by their maximum variant level PPA.3 – Panels A-C are Frailty Index and chronic pain, General Frailty and chronic pain and Fried Frailty Score on chronic pain. Whilst Panel D is F5 (poorer cognition-related frailty), Fried Frailty Score and F6 (disability-related frailty) and rheumatoid arthritis.

#### Frailty Index

For the Frailty Index (FI), >80 blocks exceeded this threshold, and >70 contained ≥1 variant with non-zero PPA₃ in the CS90 sets. Four loci showed convergent support across GWAS-PW, fine-mapping, and colocalisation: block 581 (chr5; *RAB9BP1*) with P₃ = 0.99999 and PP.H₄ = 0.92; block 419 (chr4; ZCCHC4) with P₃ = 0.99998 and PP.H₄ = 0.85; block 601 (chr5; *TRPC7*) and block 1244 (chr12; *MMAB*) with P₃ ≥ 0.997 and PP.H₄ ≈ 0.80. Additional regions (blocks 23, 761, 864, 382) showed moderate colocalisation (PP.H₄ = 0.70-0.75) despite strong PW support, indicating shared mechanisms even without a single dominant causal SNP. Compared with the diffuse overlap observed for General Frailty (GF), FI-CP produced multiple discrete loci with variant-level convergence.

#### Fried Frailty Score

The Fried Frailty Score (FFS) exhibited extensive overlap with CP, with many blocks showing P₃ > 0.95 and >70 containing variants with PPA₃ ≥ 0.80. Strong fine-mapped signals included *GIPC2* (chr1, block 47; PPA₃ = 0.998), *CRTAC1* (chr10, block 1071; PPA₃ = 0.95), and *MIR129-2/TTC17* (chr11, block 1121; PPA₃ = 0.90). Additional high-confidence loci involved *NRXN1, FOXP2, SOX6, DCC, ROBO2, BPTF, NLGN1, SETD1A,* and *TRPS1*. Colocalisation identified shared causal variants at *SLC39A8* (block 466; PP.H₄ = 0.989), *DAPP1* (block 465; PP.H₄ = 0.989), *RNU1-1* (block 19; PP.H₄ = 0.905), and *MBD2* (block 1559; PP.H₄ = 0.903). Many other fine-mapped regions showed weaker colocalisation (PP.H₄ < 0.7), but overall FFS-CP yielded one of the clearest shared signatures across domains.

#### General Frailty

General Frailty (GF) also showed broad overlap, with 84 GWAS-PW regions showing P₃ > 0.90. Fine-mapping produced compact credible sets and colocalisation identified five high-confidence loci: *SORCS3* (chr10, block 820; PP.H₄ = 0.97), *ADCY20* (chr5, block 404; PP.H₄ = 0.96), *CAMK1D* (chr10, block 775; PP.H₄ = 0.91), *TNIP1/RAB9BP1* (chr5, block 449; PP.H₄ = 0.91), and *CTNNA2* (block 123; PP.H₄ = 0.79). Additional regions showed moderate support (PP.H₄ = 0.58-0.74), including *PROK2, CA10*, *MAML3,* and *SP4*, implicating neuroimmune, metabolic, and extracellular matrix pathways.

#### Domain-specific frailty factors

F6 (disability-related frailty) showed nine regions with shared architecture (median P₃ = 0.85), including four with P₃ ≥ 0.90. Fine-mapping identified seven CS90 sets, most notably *DNAJB4* (chr1:78.4 Mb; PPA₃ = 0.98; PP.H₄ = 0.86) and NLGN1 (chr3:172-173 Mb; P₃ = 0.99; lead PPA₃ = 0.23; PP.H₄ = 0.72). Other loci, including *VPS33B, JAKMIP3/STK32C, PABPC4, SLC25A13, ZFP64*, and *LINC01524*, had P₃ = 0.67-0.90 with weak colocalisation (PP.H₄ < 0.30), indicating diffuse polygenic overlap alongside clear variant-level signals.

F5 (poorer-cognition related frailty) showed broad shared architecture, with 13 LD blocks showing P₃ ≥ 0.80 and nine ≥ 0.90. Fine-mapping identified high-confidence signals at *SLC39A8* (chr4; blocks 464 and 465), where lead variants had PPA₃ = 0.99 and 0.68 with strong colocalisation (PP.H₄ = 0.994 for both). Additional loci included *RAB9BP1* (chr5; PPA₃ = 0.95; PP.H₄ = 0.61), *NOVA1* (chr14), and *ERBB3-RAB5B-SUOX* (chr12; PP.H₄ ≈ 0.41). Other loci (*NLGN1, LINC02479, RBMS1, TCF4, PLCL1*) showed strong PW signals (P₃ = 0.81-0.91) but weak colocalisation (PP.H₄ = 0.10-0.38), indicating regional rather than variant-level sharing.

F4 (metabolic-related frailty) showed six regions with strong shared architecture (median P₃ = 0.94), three with P₃ ≥ 0.97. Two adjacent loci on chr4 showed the clearest evidence for shared causality at *SLC39A8*: block 302 (PPA₃ = 0.9998; PP.H₄ = 0.83) and block 303 (lead PPA₃ = 0.37; PP.H₄ = 0.999). Other shared regions, including *FOXP2, MIR4275, NCAM1, and TSKU*, had P₃ = 0.80-0.98 with weaker colocalisation (PP.H₄ ≈ 0.15-0.57). Most effects were concordant, although *FOXP2* (block 528) increased F4 but decreased CP, while *NCAM1* (block 756) showed the opposite pattern.

F3 (multimorbidity-related frailty) showed five regions with shared architecture (median P₃ = 0.74), including two loci with strong evidence of a shared causal variant: *NLGN1* (block 373, chr3; P₃ ≈ 0.80; PP.H₄ = 0.94) and *NMT1* (block 1453, chr17; PP.H₄ = 0.94). Other loci, including *GRK4*, *SDCCAG8*, *FHL5*, *TCF20*, *PABPC4*, *SLC25A13*, *NCAM1*, and *ZNF821*, had moderate PW support (P₃ = 0.10-0.60) with weak colocalisation (PP.H₄ < 0.15), consistent with diffuse polygenic overlap rather than a single shared causal variant.

F2 (unhealthy-lifestyle-related frailty) showed three shared regions: *PDE1C* (block 767, chr7; P₃ = 0.98; PP.H₄ = 0.21), *LOC101927237* (block 444, chr4; P₃ = 0.94; PP.H₄ = 0.42), and *FOXP1* (block 324, chr3; P₃ = 0.74; PP.H₄ = 0.28). Fine-mapping additionally prioritised *MKRN1* (block 828, chr7), although no region reached PP.H₄ > 0.80, indicating broad behavioural-genomic overlap without clear causal convergence.

F1 (limited social support-related frailty) showed two high-confidence loci: *NLGN1* (block 382, chr3; P₃ = 0.99; PPA₃ up to 0.40; PP.H₄ = 0.76) and *FAM120AOS* (block 982, chr9; P₃ = 0.89; lead PPA₃ ≈ 0.02; PP.H₄ = 0.31). Additional loci, including LINC01884 (block 145, chr2) and *INTS3* (block 76, chr1), showed moderate PW support (P₃ ≈ 0.55-0.57) but weak colocalisation (PP.H₄ ≤ 0.20), consistent with diffuse regional overlap beyond the primary *NLGN1* signal.

Figures 5 (Panels A-F) and 6 (Panels A-C) show the strongest genes by variant-level PPA₃ between frailty and chronic pain.

### GWAS Pairwise, Fine-Mapping and Colocalisation – Rheumatoid Arthritis

The ideogram in Figure 4 applies the same LD block criteria (PPA₃ > 0.5 or PPA₄ > 0.5) to rheumatoid arthritis (RA), revealing a distinct pattern from chronic pain. Several domains showed exclusively distinct architecture-F1 (1 distinct) and F3 (47 distinct)-indicating phenotype-specific RA-frailty signals. Limited sharing was observed for F4 (2 shared, 12 distinct), F5 (2, 17), and FFS (2, 4), while F6 showed the strongest shared pattern (4 shared, 6 distinct), all outside regions. The FI exhibited 2 shared and 1 distinct block, including one shared region in the MHC (the only MHC involvement), while GF showed 1 shared and 1 distinct block, both outside these regions.

Only F6, F5, and FFS retained evidence of shared causal architecture with RA. F6-RA showed the strongest profile, with three LD blocks reaching P₃ = 1.00 (blocks 69, 1016, 1245). F5-RA and FFS-RA each showed a single shared block-block 1211 (P₃ = 0.88) and block 1451 (P₃ = 0.84), respectively-while all other domains (GF, FI, F1-F4) did not reach P₃ ≥ 0.80, instead showing weaker sharing (≤ 0.69) or stronger support for distinct variants (P₄ > P₃).

Fine-mapping of F6-RA identified three regions on chr1, chr10, and chr12 with credible sets of 13, 3, and 2 variants, mapping to *MAGI3* (block 69), *IL2RA* (block 1016), and *ATXN2* (block 1245). *MAGI3* and *IL2RA* showed concordant allelic effects, whereas *ATXN2* showed discordant effects, indicating shared variants with divergent downstream consequences. F5-RA produced a compact two-variant credible set at *ERBB3* (chr12; block 1211; lead PPA₃ = 0.83), with fully concordant effects, supporting a unified genetic influence.

In contrast, FFS-RA yielded a broader 32-variant credible set on chr16 (block 1451), spanning *SGF29*, *SPNS1*, *CD19*, and *NFATC2IP*. Top PPA₃ values (0.6-0.7) were lower than for F5 and F6, but the signal formed a single LD region. Direction-of-effect analysis showed mixed influences: *SGF29* and *ATXN2L* were concordant, whereas *NFATC2IP*, *SPNS1*, and *CD19* were discordant, suggesting opposing biological effects within the same locus.

Gene-level summaries (Figure 6, Panel D) showed that within F6, *MAGI3* and *ATXN2* had the strongest per-gene PPA₃ (>0.9), followed by *IL2RA* (≈0.45), linking synaptic, RNA metabolism, and immune pathways. In F5, *ERBB3* was the sole prioritised gene. In FFS, *SGF29*, *SPNS1*, and *NFATC2IP* carried the highest PPA₃, implicating chromatin regulation, autophagy, and T-cell signalling.

The discordant effects at *NFATC2IP, SPNS1*, and *CD19* underscore that even within shared loci, frailty and RA may diverge phenotypically depending on allele direction. Colocalisation analyses reinforced these findings. For F6-RA, *MAGI3*, *ATXN2*, and *IL2RA* showed strong support for a shared causal variant (PP.H₄ ≥ 0.9; H₃ < 0.05), consistent with true pleiotropy. The *ERBB3* locus (F5-RA) also colocalised strongly (PP.H₄ ≈ 0.93). The FFS-RA region (chr16) showed a more diffuse pattern (PP.H₄ ≈ 0.84; PP.H₃ ≈ 0.11), reflecting broader LD but still supporting partial colocalisation.

Most shared loci showed concordant allelic directions, indicating variants increasing liability to both frailty and RA. However, *ATXN2* (F6) and *NFATC2IP*, *SPNS1*, and *CD19* (FFS) showed discordant effects, demonstrating that shared causal variants can produce opposing phenotypic outcomes. Figure 6 (Panel D) show the strongest genes by variant-level PPA₃ between frailty and Rheumatoid Arthritis.

### Epigenetic profiles of Frailty and Genetic Risk

#### LBC1936 Results

##### Wave 1 (age ∼70)

At Wave 1, 44 EpiScores were significantly associated with the Frailty Index after FDR correction. The strongest positive associations involved inflammatory and complement-related markers, including *CRP* (β = 0.205, 95% CI: 0.141-0.269, FDR = 2.1×10⁻⁸), *C5a* (β = 0.178, 95% CI: 0.114-0.242, FDR = 1.4×10⁻⁶), and *STC1* (β = 0.268, 95% CI: 0.168-0.368, FDR = 3.0×10⁻⁶), with additional associations for *CCL18*, *PIGR*, *LGALS3BP*, *VCAM-1*, *MMP-12/9*, *CD163*, and *GZMA* (all FDR ≤ 0.0177). Negative associations were concentrated in neuronal adhesion and ECM-related markers, including *NCAM1* (β = −0.211, FDR = 1.35×10⁻⁸), *OMD* (β = −0.219, FDR = 5.9×10⁻⁸), *NTRK3*, *CNTN4*, *SLITRK5*, *SHBG* (β = −0.243), *IGFBP-1*, *SELL*, *Adiponectin*, *SEMA3E*, and *ADAMTS13*. Network structure showed two dense, internally correlated clusters (inflammatory vs neuronal/ECM) with the Frailty Index centrally positioned. PRS associations were limited: FFS-PRS (*WFIKKN2*, *SHBG*, *STC1*; FDR ≤ 0.033), GF-PRS (*ICAM5*), and nominal effects for CP-PRS and RA-PRS. Complete Wave 1 EpiScore and PRS association results are provided in Supplementary Tables 17 and 18.

The cross-sectional proteomic signature of frailty remained consistent across subsequent LBC1936 follow-up waves. Across Waves 2-4, between 35 and 47 EpiScores remained significantly associated with the Frailty Index after FDR correction. Positive associations consistently involved inflammatory, complement, endothelial, and proteolytic markers, including *CRP*, *C5/C5a*, *CCL18*, *PIGR*, *LGALS3BP*, *VCAM-1*, and *MMP* family proteins, whereas negative associations consistently involved neuronal adhesion and ECM markers, including *NCAM1*, *CNTN4*, *OMD*, *NTRK3*, *SLITRK5*, *SHBG*, *IGFBP-1*, and *SEMA3E*. Correlation network structure remained stable across follow-up, with inflammatory and neuronal/ECM proteins forming two opposing clusters centred on the Frailty Index. Because Wave 3 contained the greatest number of significant protein associations (47 EpiScores), Figure 7 demonstrates the integrated protein–frailty network for this time point. PRS-EpiScore associations showed modest temporal variation, with FFS-PRS predominating at earlier follow-up waves and F5-PRS becoming more prominent at Wave 3. Complete cross-sectional EpiScore and PRS results for Waves 2-4 are provided in Supplementary Tables 19-24.

**Figure 7.**
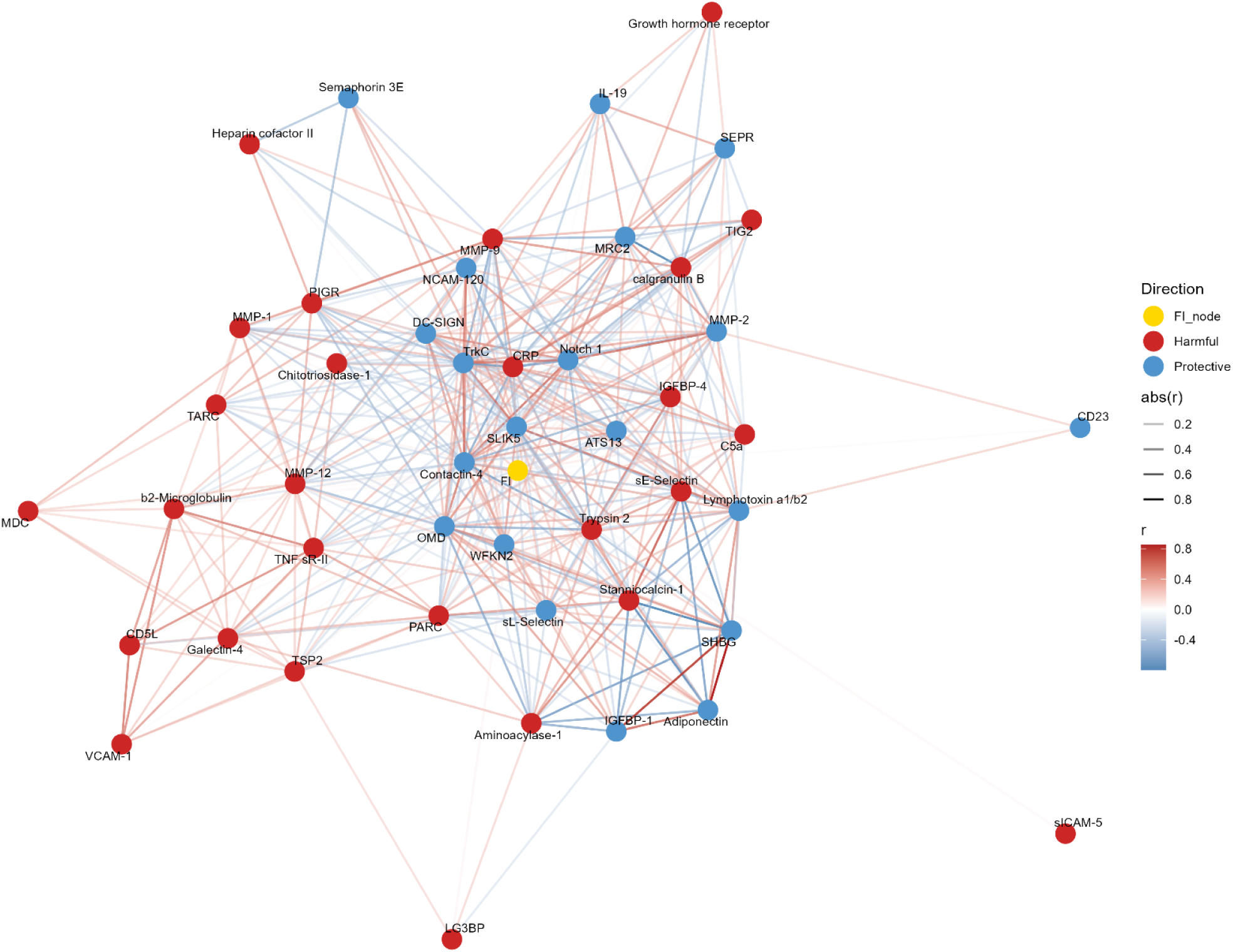
Integrated Protein-Frailty Network (LBC1936 Wave 3). An extended network incorporates direct protein-FI associations alongside the protein-protein correlation structure. In addition to the protein-protein edges (|r| > 0.30), each protein with a significant association with FI (FDR < 0.05) is connected directly to an FI node via an edge weighted by its regression coefficient (β). Edge colour again reflects effect direction (blue = protective, red = harmful), with transparency scaling to |β|.

##### Longitudinal Modelling: Protein Trajectories and Frailty Progression

In LBC1936 participants with repeated proteomic assessments, frailty showed meaningful inter-individual variation in its rate of change (FI slope: M = 0.075, SD = 0.027; range −0.053 to 0.278). Protein slopes displayed substantially greater heterogeneity (M = 0.046, SD = 0.424), although 22 proteins were excluded due to insufficient between-person variability. Mixed-effects models identified nine protein slopes that remained associated with frailty progression after FDR correction. Because both protein and frailty slopes were standardised within protein, β values reflect predictive strength rather than proportional effect size, and may exceed ±1 when slope variance is low or intercept-slope correlations are high; for example, a 1 SD increase in the *CRP* trajectory corresponds to a 2.9 SD difference in the standardised frailty slope, a scaling-dependent effect size.

Steeper increases in *CXCL10/IP-10* (β = 0.451, p = 8.9×10⁻⁶, FDR = 0.00056) and *C5/C5a* (β = 0.139, p = 5.65×10⁻⁵, FDR = 0.00166) predicted faster frailty worsening. The *CCL18/PARC* slope showed a similarly large effect (β = 1.024, p = 7.9×10⁻⁵, FDR = 0.00166). Additional significant harmful markers included *CD163* (β = 0.390, p = 0.00142, FDR = 0.0223), *CLEC11A/SCGF*-β (β = 0.199, p = 0.00462, FDR = 0.0443), *CRP* (β = 2.898, p = 0.00561, FDR = 0.0443), and *STC1* (β = 0.120, p = 0.00562, FDR = 0.0443). All FDR-significant positive associations were inflammatory, complement-related, or acute-phase markers. A smaller number of proteins showed protective longitudinal associations. Increases in *CNTN4* (β = −0.218, p = 0.00258, FDR = 0.0325) were associated with slower frailty progression, with a similar effect for *MRC2* (β = −0.963, p = 0.00849, FDR = 0.0594). These proteins predominantly reflected neuronal adhesion, ECM-processing, vascular, or immune-trafficking pathways. Complete longitudinal EpiScore-frailty trajectory results are provided in Supplementary Table 25. Complete cross-sectional and longitudinal analyses, including PRS-EpiScore associations across all four waves, are provided in Supplementary Tables 17-25.

##### PISA

In the PISA cohort, fourteen protein EpiScores were significantly associated with frailty after FDR correction, showing a pattern highly consistent with the associations observed across LBC1936. Positive associations (higher EpiScore corresponding to higher frailty) involved complement activation, proteolysis, chemotactic signalling, and related immune pathways.

These included *Aminoacylase-1* (β = 0.23, SE = 0.060, 95% CI = 0.12-0.35, FDR = 0.0083), *C5a* (β = 0.21, SE = 0.056, 95% CI = 0.10-0.32, FDR = 0.0083), *TECK* (β = 0.20, SE = 0.054, 95% CI = 0.09-0.31, FDR = 0.0083), *IGFBP-4* (β = 0.20, SE = 0.057, 95% CI = 0.09-0.33, FDR = 0.0087), *STC1* (β = 0.21, SE = 0.060, 95% CI = 0.09-0.33, FDR = 0.0087), *LGALS3BP* (β = 0.18, SE = 0.058, 95% CI = 0.07-0.30, FDR = 0.0156), *TSP2* (β = 0.17, SE = 0.058, 95% CI = 0.06-0.29, FDR = 0.0272), and *MMP-12* (β = 0.16, SE = 0.059, 95% CI = 0.05-0.27, FDR = 0.0381). These effects reflect involvement of the same complement, ECM-remodelling, and inflammatory processes that repeatedly associated with frailty in the LBC1936 waves.

Negative associations reflected metabolic, endocrine, neuronal-adhesion, and *ECM*-stability pathways, mirroring the protective patterns observed throughout LBC1936. These included *IGFBP-1* (β = −0.20, SE = 0.059, 95% CI = −0.33 to −0.07, FDR = 0.0098), *SHBG* (β = −0.19, SE = 0.055, 95% CI = −0.30 to −0.08, FDR = 0.0098), *Adiponectin* (β = −0.18, SE = 0.058, 95% CI = −0.29 to −0.06, FDR = 0.020), *NCAM-120* (β = −0.20, SE = 0.060, 95% CI = −0.32 to −0.08, FDR = 0.0138), *SLITRK5* (β = −0.18, SE = 0.063, 95% CI = −0.31 to −0.06, FDR = 0.0351), and *WFIKKN2* (β = −0.16, SE = 0.061, 95% CI = −0.28 to −0.04, FDR = 0.0417). The overlap in directional effects, molecular categories, and specific EpiScores indicates high reproducibility of both inflammation-associated and neuronal/*ECM*-linked markers across independent cohorts. The PISA protein-frailty network displayed a correlation structure similar to that observed in LBC1936, with positively associated inflammatory and complement-related proteins forming a correlated cluster, and negatively associated metabolic and neuronal-adhesion markers forming a second cluster adjacent to FI. Polygenic influences on protein EpiScores were more restricted than in LBC1936. After FDR correction, only F5-PRS demonstrated significant protein associations. Higher PRS5 was associated with elevated *MMP-12* (β = 0.15, SE = 0.044, 95% CI = 0.06-0.23, FDR = 0.041) and *STC1* (β = 0.14, SE = 0.046, 95% CI = 0.05-0.24, FDR = 0.050), and with reduced *Notch-1* (β = −0.18, SE = 0.051, 95% CI = −0.28 to −0.08, FDR = 0.040). These proteins correspond to pathways repeatedly associated with frailty across LBC1936-ECM-related proteases *(MMP-12*), stress-induced growth-factor signalling (*STC1)*, and neuronal/immune receptor regulation (Notch-1). Complete PISA EpiScore-Frailty Index and PRS-EpiScore results are presented in Supplementary Tables 26 and 27, respectively.

## Discussion

This study provides multi-omic evidence for bidirectional and domain-specific genetic links between frailty, chronic pain, and rheumatoid arthritis, bridging genetic architecture, and molecular biomarkers. Frailty, chronic pain and rheumatoid arthritis all involve persistent low-grade inflammation, metabolic alteration, and impaired resilience, yet the directionality and molecular overlap between the traits have been underexplored. The present study delineates the bidirectional and frailty domain-specific pathways linking frailty(s) with inflammation-related conditions. Through complementary analyses of GWAS, fine-mapping, colocalisation, and longitudinal epigenetic-proteomic data, we show that frailty is neither a singular nor downstream consequence of inflammation, but a composite, biologically embedded state reflecting the convergence of inflammatory, neuronal, and metabolic processes.

Across genetic layers, our results revealed strong bidirectional associations between frailty and chronic pain, supported by both Mendelian randomisation (MR) and shared genomic architecture. The forward direction indicated that genetic liability to chronic pain increases frailty burden across global measures and specific frailty domains, while reverse MR supported a causal pathway from frailty to elevated pain susceptibility. Pairwise GWAS and colocalisation analyses identified shared loci enriched for neuroimmune and extracellular-matrix (ECM) regulation, including *SLC39A8*, *NLGN1*, and *TRPC7*, suggesting intertwined pathways of nociceptive signalling, inflammation, and structural remodelling. In contrast, the relationship between frailty and RA was more confined to disability- and cognition-related frailty domains. Shared loci at IL2RA, *ERBB3*, and *MAGI3* implicated immune regulation and synaptic integrity as potential interfaces between chronic autoimmunity and functional decline. These findings align with epidemiological evidence that RA and frailty frequently co-occur but also highlight that the biological intersection between them is restricted to specific physiological domains, rather than a global frailty phenotype.

Our Mendelian randomisation analyses extend previous work by moving beyond cumulative frailty scores to examine causal relationships across genetically defined frailty domains. Prior MR studies linking frailty with chronic pain or rheumatoid arthritis have relied exclusively on deficit-based measures (that is the aggregate Frailty Index or Fried Frailty Score), which conflate multiple biological processes into a single composite trait (^23,24^). By contrast, our approach – using Genomic SEM-derived frailty domains – allows the direction and strength of associations to be resolved across specific physiological and behavioural axes of vulnerability. Within this framework, chronic pain emerged as a broad upstream driver of frailty, exerting significant causal effects across cumulative indices (FI, FFS) and distinct domains reflecting disability-related frailty (F6). These findings suggest that the impact of chronic pain extends beyond nociception, contributing to metabolic imbalance, inactivity, and physiological decline that accelerate frailty progression. Conversely, cumulative frailty measures (FI and FFS) increased chronic pain susceptibility, consistent with a feedback loop in which systemic vulnerability amplifies nociceptive sensitivity and inflammatory reactivity. This bidirectional but asymmetrical pattern provides mechanistic support for clinical observations that pain both drives and results from multisystem decline.

For rheumatoid arthritis, causal effects were more selective. Genetic liability to RA increased frailty primarily through disability-related frailty (F6), alongside robust effects on the cumulative frailty measures (FI and FFS), reflecting the contribution of chronic inflammation and mobility impairment to functional vulnerability. However, the reverse MR identified an inverse effect of the F3 domain on RA liability, possibly indicating immune exhaustion or compensatory downregulation of immune activity in highly frail individuals – a pattern reminiscent of the “inflammageing plateau” seen in late-life immune ageing (^4,5^).

Together, these domain-level results refine prior MR findings by demonstrating that frailty’s causal relationships with inflammatory conditions are multidimensional rather than unitary. Decomposing frailty into its genetic domains reveals distinct biological routes-metabolic, behavioural, immune, and physical-through which chronic pain and autoimmune processes interact with systemic ageing. This multidomain approach highlights frailty not merely as an endpoint of disease but as an active interface between chronic inflammation, lifestyle exposures, and physiological resilience.

The Pairwise GWAS and Colocalisation findings demonstrate how different frailty domains map onto partially overlapping but biologically distinct pathways. The cumulative frailty measures (FI, FFS, GF) showed extensive genomic overlap with chronic pain, implicating widespread polygenic sharing across immune, metabolic, and neuronal systems. In contrast, domain-level constructs revealed sharper mechanistic specificity. For example, poorer cognition-related frailty (F5) colocalised with *SLC39A8* and *ERBB3*-genes linking zinc transport, mitochondrial regulation, and neuroimmune signalling (^48, 49^) – whereas physical disability (F6) overlapped with *IL2RA* and *MAGI3*, central to T-cell activation and synaptic scaffolding (^50,51^). Rheumatoid arthritis shared causal variants with frailty domains through *IL2RA*, *MAGI3*, and *ATXN2*, suggesting convergence in immune and RNA metabolism pathways, whereas chronic pain shared variants at *NLGN1, FOXP2*, and *SORCS3*-genes associated with neuronal plasticity and central sensitisation (^52–55^). This domain-level resolution clarifies why aggregate frailty measures often yield diffuse biological associations: they may be conflating heterogeneous processes spanning neuroendocrine, vascular, immune, and behavioural axes. Parsing frailty into genetically distinct domains allows biological dissection of overlapping yet non-identical ageing trajectories, those governed predominantly by inflammation versus those reflecting metabolic or neuronal decline. Such separation is crucial for identifying modifiable targets within complex phenotypes.

The epigenetic-proteomic analyses revealed that frailty was characterised by two opposing epigenetic-proteomic signatures. One comprised positively associated inflammatory, complement, and acute-phase signatures, including *CRP*, *CCL18*, *C5*, and *MMPs*, consistent with chronic immune activation and tissue remodelling (^56–59^). The other axis comprised negatively associated signatures linked to neuronal adhesion, growth, and *ECM* stability, including *NCAM1, CNTN4, NTRK3, and ADAMTS1*3 (^60–63^). Higher predicted levels of these were observed in individuals with lower frailty burden, indicating preserved neurostructure and metabolic integrity. These associations should not be interpreted as causal protective effects, but rather as correlates of physiological preservation or resilience. Longitudinal analyses further demonstrated that increasing inflammatory and complement protein trajectories predicted faster frailty progression over time, whereas trajectories of structural and neuronal markers aligned with slower decline. Together, these results reveal that frailty’s molecular signature reflects a balance between vulnerability and resilience processes that evolve dynamically across ageing. The observed Epigenetic-proteomic dichotomy corresponds to the genetic architecture identified through PW-GWAS. Loci such as *SLC39A8, IL2RA, and ERBB3* map onto proteins involved in metal ion transport, cytokine signalling, and receptor regulation-all represented among the significant EpiScores (^64–66^). This genetic-to-epigenetic convergence supports the notion that frailty arises from integrated dysregulation across inflammatory, metabolic, and neuronal systems rather than from any single pathway.

These findings have implications for understanding inter-individual variability in ageing trajectories. The identification of resilience-related molecular profiles-especially those involving neuronal adhesion and ECM maintenance – suggests that preserved system integrity may buffer against inflammatory acceleration of frailty (^67–70^). Because frailty development is closely intertwined with lifestyle factors such as diet, physical activity, and stress, the present results highlight opportunities to integrate genetic and molecular stratification with behavioural or environmental interventions (^71,72^). Mapping how lifestyle modifies domain-specific pathways (inflammation versus structural preservation) may help identify modifiable leverage points to delay frailty progression or mitigate its systemic consequences. This aligns with the emerging view of frailty not solely as a clinical outcome, but as a measurable biological state that bridges ageing biology and modifiable risk (^73^).

Several methodological limitations should be acknowledged. First, the frailty measures used across GWAS and cohort analyses likely capture early or subclinical frailty rather than advanced states of physiological decline, reflecting a healthy volunteer bias inherent in large biobank samples (^74^). This may attenuate genetic associations and underrepresent the biology of more severe frailty phenotypes observed in clinical or institutionalised populations.

Second, Mendelian randomisation analyses, while designed to infer causality, remain sensitive to biases arising from pleiotropy, collider effects, and selection on survival-particularly in ageing cohorts where frailty itself influences participation and longevity (^74,75^). These factors may distort bidirectional effect estimates or obscure latent shared influences. Finally, the findings remain limited by reliance on European-ancestry datasets. Future studies incorporating more diverse ancestries, clinically enriched cohorts, and deeper phenotyping of frailty severity, especially as frailty may exhibit differently across populations and cultures, will be crucial to determine whether the identified pathways generalise across populations and frailty stages, and whether they capture causal biology or shared systemic vulnerability.

In summary, this study provides multi-layered evidence that frailty, chronic pain, and rheumatoid arthritis share convergent biological mechanisms that span immune activation, neurostructure preservation, and systemic resilience. Frailty emerges not as a passive consequence of disease, but as an active biological interface between chronic inflammation and system integrity. By combining genetic, molecular, and longitudinal data, our results reveal a bidirectional and domain-specific framework of ageing, offering new opportunities to target inflammation, neurostructure health, and lifestyle factors to preserve functional capacity.

## Data Availability

All data produced in the present study are available upon reasonable request to the authors

## Acknowledgments

Concept and Design: JP Flint, Drafting of the manuscript: JP Flint; Critical revision of the manuscript: JPF, BLM, SMH, HMS, IFF, SRC, ML, MKL.

This research was funded by the Legal & General Group (research grant to establish the independent Advanced Care Research Centre at the University of Edinburgh). The funder had no role in the conduct of the study, interpretation, or the decision to submit for publication. The views expressed are those of the authors and not necessarily those of legal and general. The authors declare no competing interests. Availability of data and material: Details and links for all summary statistics used are available in Supplementary Table 1.

## Notes

### Competing Interest Statement

The authors have declared no competing interest.

### Author Declarations

Lothian Birth Cohort 1936 (LBC1936). LBC1936 is a longitudinal study of community dwelling older adults living in and around Edinburgh, Scotland (United Kingdom). Individuals joined the study when they initially took part in the Scottish Mental Survey in 1947 and have thus far taken part in 5 waves of testing. The main analyses, when examining frailty outcomes, were performed on wave 1 (mean age = 69.60; SD = 0.83; N = 1005 (509 males, 496 females)). Ethics for LBC1936 was approved by the MultiCentre Research Ethics Committee for Scotland (Wave 1: MREC/01/0/56), the Lothian Research Ethics Committee (Wave 1: LREC/2003/2/29), and the Scotland A Research Ethics Committee (Waves 2, 3, 4 and 5: 07/MRE00/58) and all methods were performed in accordance with the relevant guidelines and regulations. Informed Written Consent was obtained from participants at each of the waves. The genotypes, which were collected via blood samples from the participants at Wave 1 were processed using stringent quality control measures. Genotyping was conducted using the Illumina Human610- Quadv1 chip, with 542,050 SNPs passing quality control. Out of 1,091 individuals in the LBC1936 cohort, genomic DNA from 1,071 individuals was successfully isolated using standard procedures at the Wellcome Trust Clinical Research Facility (WTCRF) Genetics Core. Twenty-nine samples did not meet quality control standards before genotyping. The remaining 1,042 blood-extracted samples were genotyped at the WTCRF Genetics Core with the Illumina Human610-Quadv1 chip, and SNPs were imputed to the 1000 Genomes reference panel (phase 1, version 3). After undergoing the described quality control procedures, samples from 1,005 individuals were retained for analysis. Prospective Imaging Study of Aging (PISA). PISA is a longitudinal cohort of ~3800 Australian adults aged 40-80 years. Since this cohort includes twin pairs, we selected one of each pair at random to be included in our current analyses. When the Frailty Index (FI) was derived and linked with genotypic data, 3265 participants (1034 males and 2231 females) remained for the main analysis, with a mean age of 60.34 (SD = 6.98). The PISA study protocol was approved by the Human Research Ethics Committee (HREC) at QIMR Berghofer Medical Research Institute. Participants are community dwelling and were recruited from a pool of participants who previously volunteered for research on risk factors for physical or psychiatric conditions and had genotypes already available. Genome-wide genotyping of participants was previously conducted using various genotyping arrays, including Illumina chips based on HapMap references (317 K, 370 K, 610 K, 660 K) and more recent Illumina arrays based on the 1KGP reference (Core + Exome, PsychArray, OmniExpress) as detailed in earlier studies. All datasets have been combined following rigorous quality control procedures and imputed using the Haplotype Reference Consortium (HRC) Release 1 reference panel.

### Summary of Updates

Authors may wish to note any significant differences between this version and previous versions of the manuscript.

## References

1 Clegg A, Young J, Iliffe S, Rikkert MO, Rockwood K. Frailty in elderly people. Lancet. 2013 Mar 2;381(9868):752–62.

2 Hubbard RE, Peel NM, Samanta M, Gray LC, Mitnitski A, Rockwood K. Frailty status at admission to hospital predicts multiple adverse outcomes. Age and ageing. 2017 Sep 1;46(5):801–6.

3 Ward DD, Flint JP, Littlejohns TJ, Foote IF, Canevelli M, Wallace LM, Gordon EH, Llewellyn DJ, Ranson JM, Hubbard RE, Rockwood K. Frailty trajectories preceding dementia in the US and UK. JAMA neurology. 2025 Jan 1;82(1):61–71.

4 Ferrucci L, Fabbri E. Inflammageing: chronic inflammation in ageing, cardiovascular disease, and frailty. Nature Reviews Cardiology. 2018 Sep;15(9):505–22.

5 Salaffi F, Di Matteo A, Farah S, Di Carlo M. Inflammaging and frailty in immune-mediated rheumatic diseases: how to address and score the issue. Clinical reviews in allergy & immunology. 2023 Apr;64(2):206–21.

6 Wang J, Maxwell CA, Yu F. Biological processes and biomarkers related to frailty in older adults: a state-of-the-science literature review. Biological research for nursing. 2019 Jan;21(1):80–106.

7 Searle SD, Mitnitski A, Gahbauer EA, Gill TM, Rockwood K. A standard procedure for creating a frailty index. BMC geriatrics. 2008 Sep 30;8(1):24.

8 Fried LP, Tangen CM, Walston J, Newman AB, Hirsch C, Gottdiener J, Seeman T, Tracy R, Kop WJ, Burke G, McBurnie MA. Frailty in older adults: evidence for a phenotype. The Journals of Gerontology Series A: Biological Sciences and Medical Sciences. 2001 Mar 1;56(3):M146–57.

9 Atkins JL, Jylhävä J, Pedersen NL, Magnusson PK, Lu Y, Wang Y, Hägg S, Melzer D, Williams DM, Pilling LC. A genome-wide association study of the frailty index highlights brain pathways in ageing. Aging cell. 2021 Sep;20(9):e13459.

10 Ye Y, Noche RB, Szejko N, Both CP, Acosta JN, Leasure AC, Brown SC, Sheth KN, Gill TM, Zhao H, Falcone GJ. A genome-wide association study of frailty identifies significant genetic correlation with neuropsychiatric, cardiovascular, and inflammation pathways. Geroscience. 2023 Aug;45(4):2511–23.

11 Foote IF, Flint JP, Fürtjes AE, Lawrence JM, Mullin DS, Fisk JD, Karakach TK, Rutenberg A, Martin NG, Lupton MK, Llewellyn DJ. Uncovering the multivariate genetic architecture of frailty with genomic structural equation modeling. Nature Genetics. 2025 Aug;57(8):1848–59.

12 Gale CR, Baylis D, Cooper C, Sayer AA. Inflammatory markers and incident frailty in men and women: the English Longitudinal Study of Ageing. Age. 2013 Dec;35(6):2493–501.

13 Bålsrud P, Ulven SM, Ottestad I, Retterstøl K, Schwab U, Holven KB. Association between inflammatory markers, body composition and frailty in home-dwelling elderly: an 8-year follow-up study. GeroScience. 2024 Dec;46(6):5629–41.

14 Soysal P, Stubbs B, Lucato P, Luchini C, Solmi M, Peluso R, Sergi G, Isik AT, Manzato E, Maggi S, Maggio M. Inflammation and frailty in the elderly: a systematic review and meta-analysis. Ageing research reviews. 2016 Nov 1;31:1–8.

15 Luo YF, Cheng ZJ, Wang YF, Jiang XY, Lei SF, Deng FY, Ren WY, Wu LF. Unraveling the relationship between high-sensitivity C-reactive protein and frailty: evidence from longitudinal cohort study and genetic analysis. BMC geriatrics. 2024 Mar 4;24(1):222.

16 Michaëlsson K, Zheng R, Baron JA, Fall T, Wolk A, Lind L, Höijer J, Brunius C, Lemming EW, Titova OE, Svennblad B. Cardio-metabolic-related plasma proteins reveal biological links between cardiovascular diseases and fragility fractures: a cohort and Mendelian randomisation investigation. EBioMedicine. 2025 Mar 1;113.

17 Lin T, Zhao Y, Xia X, Ge N, Yue J. Association between frailty and chronic pain among older adults: a systematic review and meta-analysis. European Geriatric Medicine. 2020 Dec;11(6):945–59.

18 Yang S, Wang H, Tao Y, Tian J, Wen Z, Cao J, Zhang W, Peng S, Zhang X. Association of chronic pain with frailty and pre-frailty in older adults: A systematic review and meta-analysis. Archives of Gerontology and Geriatrics. 2025 Feb 11:105784.

19 Abdullah GA, Akpan A, Phelan MM, Wright HL. The complex role of neutrophils in healthy aging, inflammaging, and frailty. Journal of leukocyte biology. 2025 Sep;117(9):qiaf117.

20 Bauer ME. Accelerated immunosenescence in rheumatoid arthritis: impact on clinical progression. Immunity & Ageing. 2020 Mar 9;17(1):6.

21 Johnston KJ, Adams MJ, Nicholl BI, Ward J, Strawbridge RJ, Ferguson A, McIntosh AM, Bailey ME, Smith DJ. Genome-wide association study of multisite chronic pain in UK Biobank. PLoS genetics. 2019 Jun 13;15(6):e1008164.

22 Okada Y, Wu D, Trynka G, Raj T, Terao C, Ikari K, Kochi Y, Ohmura K, Suzuki A, Yoshida S, Graham RR. Genetics of rheumatoid arthritis contributes to biology and drug discovery. Nature. 2014 Feb 20;506(7488):376–81.

23 Dai Z, Wu Y, Chen J, Huang S, Zheng H. Assessment of relationships between frailty and chronic pain: a bidirectional two-sample Mendelian randomisation study. Age and Ageing. 2024 Jan 1;53(1):afad256.

24 Wang S, Han J, Wang Q, Li Q, Cui Y. The Causal Relationship Between Chronic Pain and Frailty: A Two-Sample Mendelian Randomization Study. Biological research for nursing. 2025 Oct;27(4):630–9.

25 Wen L, Fan J, Shi X, Zhou H, Yang Y, Jia X. Causal association of rheumatoid arthritis with frailty and the mediation role of inflammatory cytokines: a Mendelian randomization study. Archives of Gerontology and Geriatrics. 2024 Jul 1;122:105348.

26 Zhou J, Zhang Y, Ni T, Li Y, Shao H, Wang F, Xu S, Huang Y, Zhang J, Zhao T. Does autoimmune diseases increase the risk of frailty? A Mendelian randomization study. Frontiers in Endocrinology. 2024 Aug 27;15:1364368.

27 Mitchell A, Malmgren L, Bartosch P, McGuigan FE, Akesson KE. Pro-inflammatory proteins associated with frailty and its progression-A longitudinal study in community-dwelling women. Journal of Bone and Mineral Research. 2023 Aug 1;38(8):1076–91.

28 Li C, Zeng N, Xue FS. Temporal association between chronic pain and frailty occurrence, and the modifiable role of a healthy lifestyle in Chinese middle-aged and older population: a community based, prospective cohort study. Aging Clinical and Experimental Research. 2025 Dec;37(1):1–1.

29 Chaplin WJ, McWilliams DF, Millar BS, Gladman JR, Walsh DA. The bidirectional relationship between chronic joint pain and frailty: data from the Investigating Musculoskeletal Health and Wellbeing cohort. BMC geriatrics. 2023 May 5;23(1):273.

30 Huang H, Ni L, Zhang L, Zhou J, Peng B. Longitudinal association between frailty and pain in three prospective cohorts of older population. The Journal of nutrition, health and aging. 2025 Jun 1;29(6):100537.

31 Flint JP, Welstead M, Cox SR, Russ TC, Marshall A, Luciano M. Multi-Polygenic prediction of Frailty and its Trajectories highlights Chronic Pain, Rheumatoid Arthritis, and Educational Attainment pathways. medRxiv. 2025 Jul 21:2024–05.

32 Zheng J, Baird D, Borges MC, Bowden J, Hemani G, Haycock P, et al. Recent developments in Mendelian randomization studies. Curr Epidemiol Rep. 2017 Nov 22;4(4):330–45.

33 Skrivankova VW, Richmond RC, Woolf BA, Yarmolinsky J, Davies NM, Swanson SA, VanderWeele TJ, Higgins JP, Timpson NJ, Dimou N, Langenberg C. Strengthening the reporting of observational studies in epidemiology using Mendelian randomization: the STROBE-MR statement. Jama. 2021 Oct 26;326(16):1614–21.

34 Hemani G, Zheng J, Elsworth B, Wade KH, Haberland V, Baird D, et al. The MR-Base platform supports systematic causal inference across the human phenome. Elife [Internet]. 2018 May 30;7. Available from: 10.7554/eLife.34408

35 Verbanck M, Chen CY, Neale B, Do R. Detection of widespread horizontal pleiotropy in causal relationships inferred from Mendelian randomization between complex traits and diseases. Nat Genet. 2018 May;50(5):693–8.

36 ) A Language and Environment for Statistical Computing_. R Foundation for Statistical Computing. A Language and Environment for Statistical Computing_ R Foundation for Statistical Computing.

37 Sanderson E, Spiller W, Bowden J. Testing and correcting for weak and pleiotropic instruments in two-sample multivariable Mendelian randomization. Stat Med. 2021 Nov 10;40(25):5434–52.

38 Pickrell JK, Berisa T, Liu JZ, Ségurel L, Tung JY, Hinds DA. Detection and interpretation of shared genetic influences on 42 human traits. Nature genetics. 2016 Jul;48(7):709–17.

39 Mitchell BL, Lupton MK, Rentería ME, Simpson MA, Reay WR. Genetic exploration of the relationship between liability to psychiatric disorders and acne vulgaris. European Journal of Human Genetics. 2026 Feb 4:1–9.

40 Quinlan AR, Hall IM. BEDTools: a flexible suite of utilities for comparing genomic features. Bioinformatics. 2010 Mar 15;26(6):841–2.

41 Giambartolomei C, Vukcevic D, Schadt EE, Franke L, Hingorani AD, Wallace C, Plagnol V. Bayesian test for colocalisation between pairs of genetic association studies using summary statistics. PLoS genetics. 2014 May 15;10(5):e1004383.

42 Deary IJ, Gow AJ, Taylor MD, Corley J, Brett C, Wilson V, Campbell H, Whalley LJ, Visscher PM, Porteous DJ, Starr JM. The Lothian Birth Cohort 1936: a study to examine influences on cognitive ageing from age 11 to age 70 and beyond. BMC geriatrics. 2007 Dec 5;7(1):28.

43 Lupton MK, Robinson GA, Adam RJ, Rose S, Byrne GJ, Salvado O, Pachana NA, Almeida OP, McAloney K, Gordon SD, Raniga P. A prospective cohort study of prodromal Alzheimer’s disease: Prospective Imaging Study of Ageing: Genes, Brain and Behaviour (PISA). NeuroImage: Clinical. 2021 Jan 1;29:102527.

44 Gadd DA, Hillary RF, McCartney DL, Zaghlool SB, Stevenson AJ, Cheng Y, Fawns-Ritchie C, Nangle C, Campbell A, Flaig R, Harris SE. Epigenetic scores for the circulating proteome as tools for disease prediction. Elife. 2022 Jan 13;11:e71802.

45 Smith HM, Moodie JE, Monterrubio-Gómez K, Gadd DA, Hillary RF, Chybowska AD, McCartney DL, Campbell A, Redmond P, Page D, Taylor A. Epigenetic scores of blood-based proteins as biomarkers of general cognitive function and brain health. Clinical Epigenetics. 2024 Mar 25;16(1):46.

46 Lloyd-Jones LR, Zeng J, Sidorenko J, Yengo L, Moser G, Kemper KE, Wang H, Zheng Z, Magi R, Esko T, Metspalu A. Improved polygenic prediction by Bayesian multiple regression on summary statistics. Nature communications. 2019 Nov 8;10(1):5086.

47 Csardi MG. Package ‘igraph’. Last accessed. 2013 Apr 7;3(09):2013.

48 Nebert DW, Liu Z. SLC39A8 gene encoding a metal ion transporter: discovery and bench to bedside. Human genomics. 2019 Sep 14;13(1):51.

49 Ding Y, Zhang Y, Zhang L. THE ROLE OF THE NEUREGULIN 1-ERBB4 SIGNALING PATHWAY IN NEUROLOGICAL DISORDERS. Journal of Physiology & Pharmacology. 2024 Dec 1;75(6).

50 Peerlings D, Mimpen M, Damoiseaux J. The IL-2-IL-2 receptor pathway: Key to understanding multiple sclerosis. Journal of translational autoimmunity. 2021 Jan 1;4:100123.

51 Kotelevets L, Chastre E. A new story of the three Magi: Scaffolding proteins and lncRNA suppressors of cancer. Cancers. 2021 Aug 24;13(17):4264.

52 Camporesi E, Lashley T, Gobom J, Lantero-Rodriguez J, Hansson O, Zetterberg H, Blennow K, Becker B. Neuroligin-1 in brain and CSF of neurodegenerative disorders: investigation for synaptic biomarkers. Acta Neuropathologica Communications. 2021 Feb 1;9(1):19.

53 Yu Q, Wang X, Yang Y, Chi P, Huang J, Qiu S, Zheng X, Chen X. Upregulated NLGN1 predicts poor survival in colorectal cancer. BMC cancer. 2021 Aug 2;21(1):884.

54 He BH, Yang YH, Hsiao BW, Lin WT, Chuang YF, Chen SY, Liu FC. Foxp2 is required for nucleus accumbens-mediated multifaceted limbic function. Neuroscience. 2024 Mar 26;542:33–46.

55 Kamran M, Laighneach A, Bibi F, Donohoe G, Ahmed N, Rehman AU, Morris DW. Independent associated SNPs at SORCS3 and its protein interactors for multiple brain-related disorders and traits. Genes. 2023 Feb 14;14(2):482.

56 Palamidas DA, Chatzis L, Papadaki M, Gissis I, Kambas K, Andreakos E, Goules AV, Tzioufas AG. Current insights into tissue injury of giant cell arteritis: from acute inflammatory responses towards inappropriate tissue remodeling. Cells. 2024 Feb 29;13(5):430.

57 McMahon M, Ye S, Pedrina J, Dlugolenski D, Stambas J. Extracellular matrix enzymes and immune cell biology. Frontiers in Molecular Biosciences. 2021 Aug 30;8:703868.

58 Wojciuk B, Frulenko I, Brodkiewicz A, Kita D, Baluta M, Jędrzejczyk F, Budkowska M, Turkiewicz K, Proia P, Ciechanowicz A, Kostrzewa-Nowak D. The Complement System as a Part of Immunometabolic Post-Exercise Response in Adipose and Muscle Tissue. International Journal of Molecular Sciences. 2024 Oct 29;25(21):11608.

59 Potempa M, Hart PC, Rajab IM, Potempa LA. Redefining CRP in tissue injury and repair: more than an acute pro-inflammatory mediator. Frontiers in Immunology. 2025 Feb 28;16:1564607.

60 Duncan BW, Murphy KE, Maness PF. Molecular mechanisms of L1 and NCAM adhesion molecules in synaptic pruning, plasticity, and stabilization. Frontiers in Cell and Developmental Biology. 2021 Jan 28;9:625340.

61 Bamford RA, Zuko A, Eve M, Sprengers JJ, Post H, Taggenbrock RL, Fäβler D, Mehr A, Jones OJ, Kudzinskas A, Gandawijaya J. CNTN4 modulates neural elongation through interplay with APP. Open Biology. 2024 May 15;14(5):240018.

62 Zeng S, Jiang K, Ge J, Tang M, Wen Y, Ma X, Liu H, Xiong X. NTRK fusion promotes tumor migration and invasion through epithelial-mesenchymal transition and closely interacts with ECM1 and NOVA1. BMC cancer. 2024 Dec 5;24(1):1502.

63 Satz-Jacobowitz B, Hubmacher D. The quest for substrates and binding partners: A critical barrier for understanding the role of ADAMTS proteases in musculoskeletal development and disease. Developmental Dynamics. 2021 Jan;250(1):8–26.

64 Li S, Liu S. SLC39A10 Expression is Associated with the Prognosis of Hepatocellular Carcinoma and the Tumor Immune Microenvironment. Med Discoveries. 2024;3(11):1223.

65 An N, Li M. Comprehensive analysis of characteristic genes of inflammation-related bronchopulmonary dysplasia based on bioinformatics methods.

66 Samuriwo T. Functional Characterisation of Copine III (CPNE3) in ERBB2 Overexpressing Breast Cancer (Doctoral dissertation, UCL (University College London)).

67 Vidović T, Ewald CY. Longevity-promoting pathways and transcription factors respond to and control extracellular matrix dynamics during aging and disease. Frontiers in aging. 2022 Jul 7;3:935220.

68 Statzer C, Park JY, Ewald CY. Extracellular matrix dynamics as an emerging yet understudied hallmark of aging and longevity. Aging and Disease. 2023 Jun 1;14(3):670.

69 Dzyubenko E, Hermann DM. Role of glia and extracellular matrix in controlling neuroplasticity in the central nervous system. InSeminars in Immunopathology 2023 May (Vol. 45, No. 3, pp. 377–387). Berlin/Heidelberg: Springer Berlin Heidelberg.

70 Ji F, Lee HS, Lee H, Kim JH. The impact of frailty syndrome on skeletal muscle histology: preventive effects of exercise. FEBS Open Bio. 2025 May 5.

71 Howlett SE, Rutenberg AD, Rockwood K. The degree of frailty as a translational measure of health in aging. Nature Aging. 2021 Aug;1(8):651–65.

72 Tang J, Ma Y, Hoogendijk EO, Chen J, Yue J, Wu C. Associations between healthy lifestyle and mortality across different social environments: a study among adults with frailty from the UK Biobank. European Journal of Public Health. 2024 Apr 1;34(2):218–24.

73 Sato R, Vatic M, Peixoto da Fonseca GW, Anker SD, von Haehling S. Biological basis and treatment of frailty and sarcopenia. Cardiovascular research. 2024 Jun;120(9):982–98.

74 van Alten S, Domingue BW, Faul J, Galama T, Marees AT. Reweighting UK Biobank corrects for pervasive selection bias due to volunteering. International journal of epidemiology. 2024 Jun 1;53(3):dyae054.

75 Sanderson E, Rosoff D, Palmer T, Tilling K, Smith GD, Hemani G. Bias from heritable confounding in Mendelian randomization studies. medRxiv. 2024 Sep 6:2024–09.

