## Supplementary Materials for "Multi-Omic Analyses Reveal Bidirectional Genetic Links and Convergent Inflammatory-Neuronal Signatures Between Frailty, Chronic Pain, and Rheumatoid Arthritis"

### Full information on the Independent Cohorts

#### *Lothian Birth Cohort 1936 (LBC1936)*

LBC1936 is a longitudinal study of community-dwelling older adults from Edinburgh and the surrounding areas in Scotland (UK) (1). Participants were originally recruited through the Scottish Mental Survey of 1947 and have since completed five waves of follow-up assessments. For the present analyses focusing on frailty outcomes, we used data from Wave 1, where the mean age was 69.60 years (SD = 0.83) and the sample comprised 1,005 individuals (509 men and 496 women). Ethical approval for LBC1936 was granted by the Multi-Centre Research Ethics Committee for Scotland (Wave 1: MREC/01/0/56), the Lothian Research Ethics Committee (Wave 1: LREC/2003/2/29), and the Scotland A Research Ethics Committee (Waves 2–5: 07/MRE00/58). All procedures adhered to relevant guidelines and regulations, and written informed consent was obtained from participants at every wave.

Genotyping was performed on blood samples collected at Wave 1 and underwent stringent quality-control procedures (2). DNA was extracted for 1,071 of the 1,091 cohort members using standard protocols at the Wellcome Trust Clinical Research Facility (WTCRF) Genetics Core. Twenty-nine samples failed quality control prior to genotyping. The remaining 1,042 blood-derived samples were genotyped on the Illumina Human610-Quadv1 array, yielding 542,050 SNPs that passed QC. SNPs were then imputed to the 1000 Genomes Phase 1, Version 3 reference panel. After all QC steps were applied, genotype data for 1,005 individuals were available for analysis. For methylation, DNA was extracted from whole-blood samples, and genome-wide DNA methylation (DNAm) was profiled using the Illumina 450K methylation array. Quality-control procedures for the DNAm data have been detailed previously (3–6). In summary, raw intensity files underwent background correction and control-based normalisation. Samples exhibiting technical issues—such as poor hybridisation or nucleotide extension, inadequate bisulfite conversion, or abnormal signal staining—were excluded manually. Probes failing quality thresholds were also removed, including those with detection rates below 95% or fewer than 450,000 detectable probes at $p < 0.01$. Additionally, samples with discrepancies between methylation-predicted and recorded sex, or inconsistencies between genotype data and SNP control probes, were excluded (3)

#### *Prospective Imaging Study of Ageing (PISA)*

PISA is a longitudinal cohort comprising approximately 3,800 Australian adults aged 40–80 years (6). Because the sample includes twin pairs, one twin from each pair was randomly selected to avoid relatedness in the present analyses. After deriving the Frailty Index (FI) and linking it to available genotype data, 3,265 individuals (1,034 men and 2,231 women) were retained for the primary analyses, with a mean age of 60.34 years (SD = 6.98). Ethical approval for PISA was granted by the Human Research Ethics Committee of the QIMR Berghofer Medical Research Institute.

Participants were community-dwelling adults recruited from an existing volunteer pool who had previously participated in studies of physical or psychiatric risk factors and already had genotype data available. Genome-wide genotyping had been performed using a range of Illumina platforms, including earlier HapMap-based arrays (317K, 370K, 610K, 660K) as well as more recent arrays based on the 1000 Genomes reference panel (Core + Exome, PsychArray, OmniExpress), as described in prior publications (7, 8). All genotyping datasets were harmonised and subjected to stringent quality-control procedures before being imputed to the Haplotype Reference Consortium (HRC) Release 1 reference panel (9).

To ensure that all analyses were based on a consistent and biologically meaningful sample, the PISA dataset was restricted to participants who had complete EpiScore measurements and matching frailty information. Because EpiScore data, frailty indices, and genetic scores and covariates were sourced from separate components of the PISA study, alignment by participant identifier (MLID) was essential to avoid mismatched or duplicated records. We therefore limited the analytic sample to individuals present in both the proteomic and frailty datasets, ensuring that each retained participant contributed a single, coherent set of observations – this resulted in a downstream sample of 339 participants.

Enter PISA methylation here -
For methylation, DNA was extracted from whole blood samples, and DNAm was profiled using the Illuminanfinium

MethylationEPIC v1.0 and v2.0. Data quality control and normalisation were conducted using the R package Meffil (version 1.3.8) (10). For quality control “blood gse35069 complete” was used as the cell type reference and the featureset defined as “epic:epic2”. Plink (version 1.90-b.7.7) was used to extract imputed genotype data to verify sample identity and mismatch detection. Quality control parameters used were as follows:   beadnum.samples.threshold = 0.1, detectionp.samples.threshold = 0.1, detectionp.cpgs.threshold = 0.1, beadnum.cpgs.threshold = 0.1, sex.outlier.sd = 5, snp.concordance.threshold = 0.95, sample.genotype.concordance.threshold = 0.8). Metyhlation data s werenormalised using the following parameters: control.pcs = 1:10, batch.pcs = 1:10, batch.threshold = 0.0. Only the 20,000 most variable CpG probe sites were used for normalisation.

Leave One Out Plots

Leave-one-out sensitivity analyses were performed to evaluate whether the overall Mendelian randomisation estimates were driven by individual genetic instruments. Each point represents the inverse-variance weighted (IVW) causal estimate obtained after excluding a single SNP, with horizontal lines indicating the corresponding 95% confidence interval. The red point ("All") represents the IVW estimate including all SNPs. Consistent estimates across iterations indicate that no single variant disproportionately influenced the overall causal effect estimate.

### Chronic Pain Exposure on Frailty Outcomes

CP and FI

Supplementary Figure 1. Leave-one-out sensitivity analysis for CP and FI.

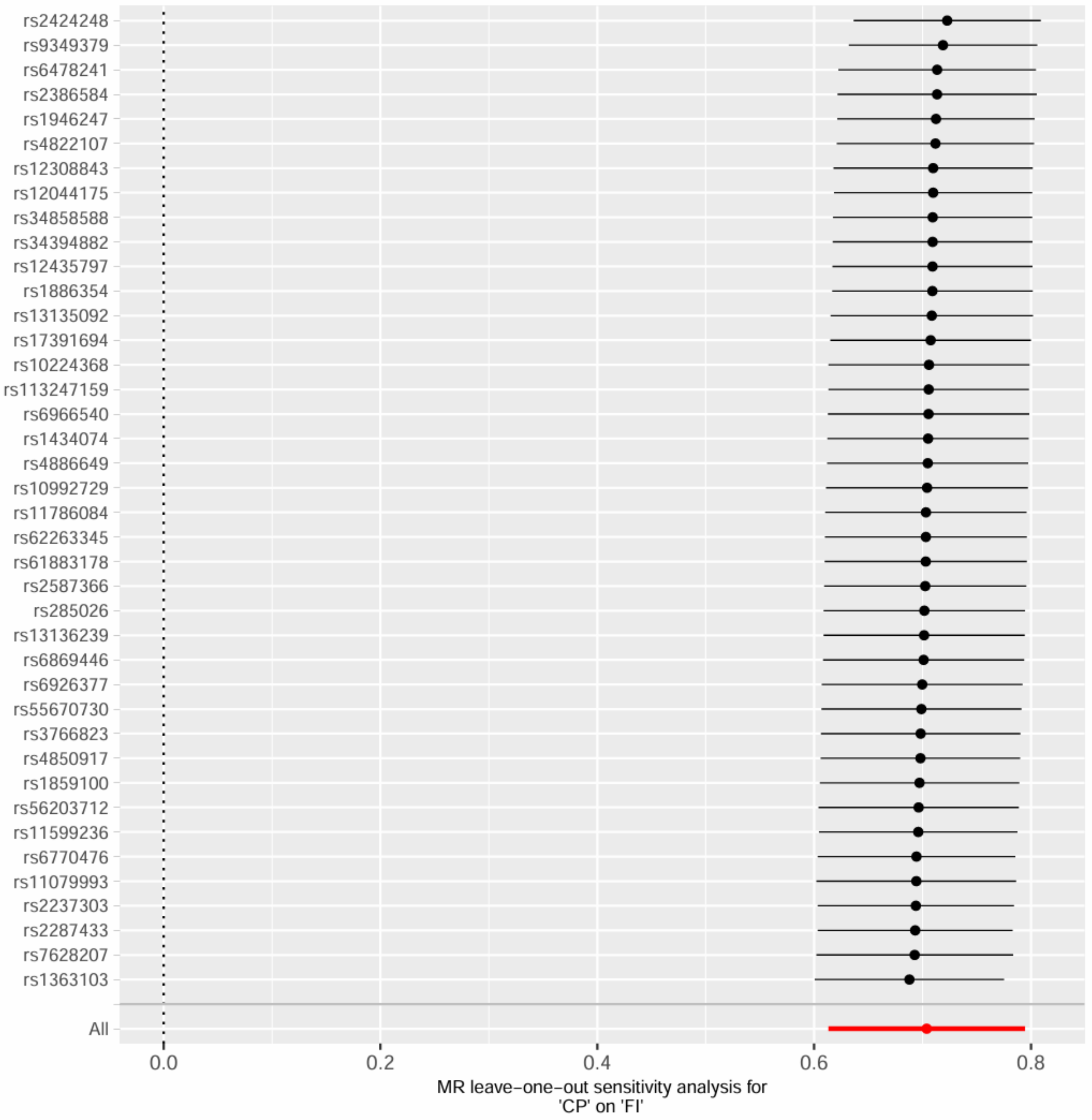


CP and FI, excluding MHC region

Supplementary Figure 2. Leave-one-out sensitivity analysis for CP and FI, excluding MHC region.

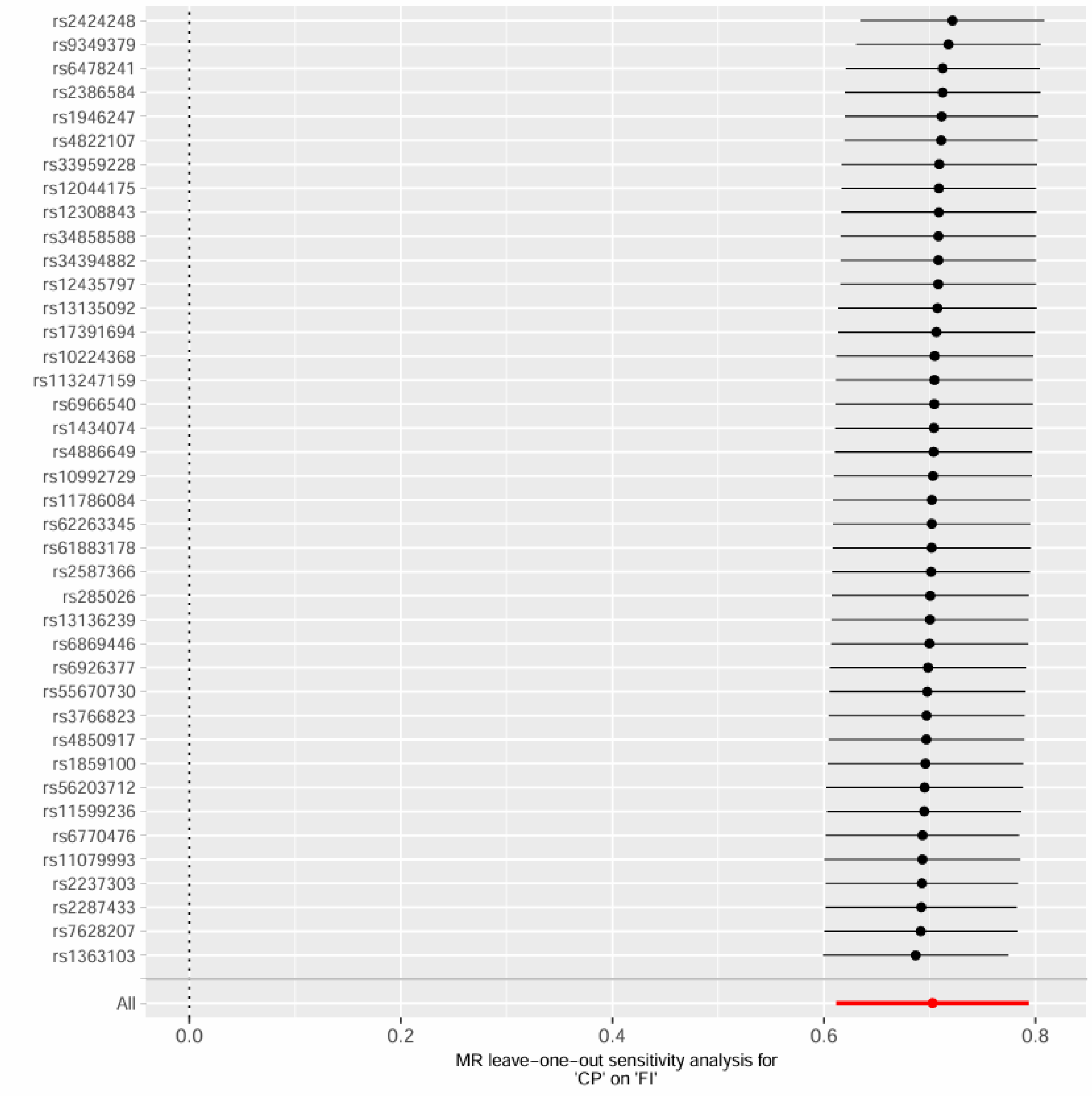

rs2424248
rs9349379
rs6478241
rs2386584
rs1946247
rs4822107
rs33959228
rs12044175
rs12308843
rs34858588
rs34394882
rs12435797
rs13135092
rs17391694
rs10224368
rs113247159
rs6966540
rs1434074
rs4886649
rs10992729
rs11786084
rs62263345
rs61883178
rs2587366
rs285026
rs13136239
rs6869446
rs6926377
rs55670730
rs3766823
rs4850917
rs1859100
rs56203712
rs11599236
rs6770476
rs11079993
rs2237303
rs2287433
rs7628207
rs1363103
All
0.0
0.2
0.4
0.6
0.8
MR leave-one-out sensitivity analysis for
'CP' on 'FI'


CP and FFS

Supplementary Figure 3. Leave-one-out sensitivity analysis for CP and FFS.

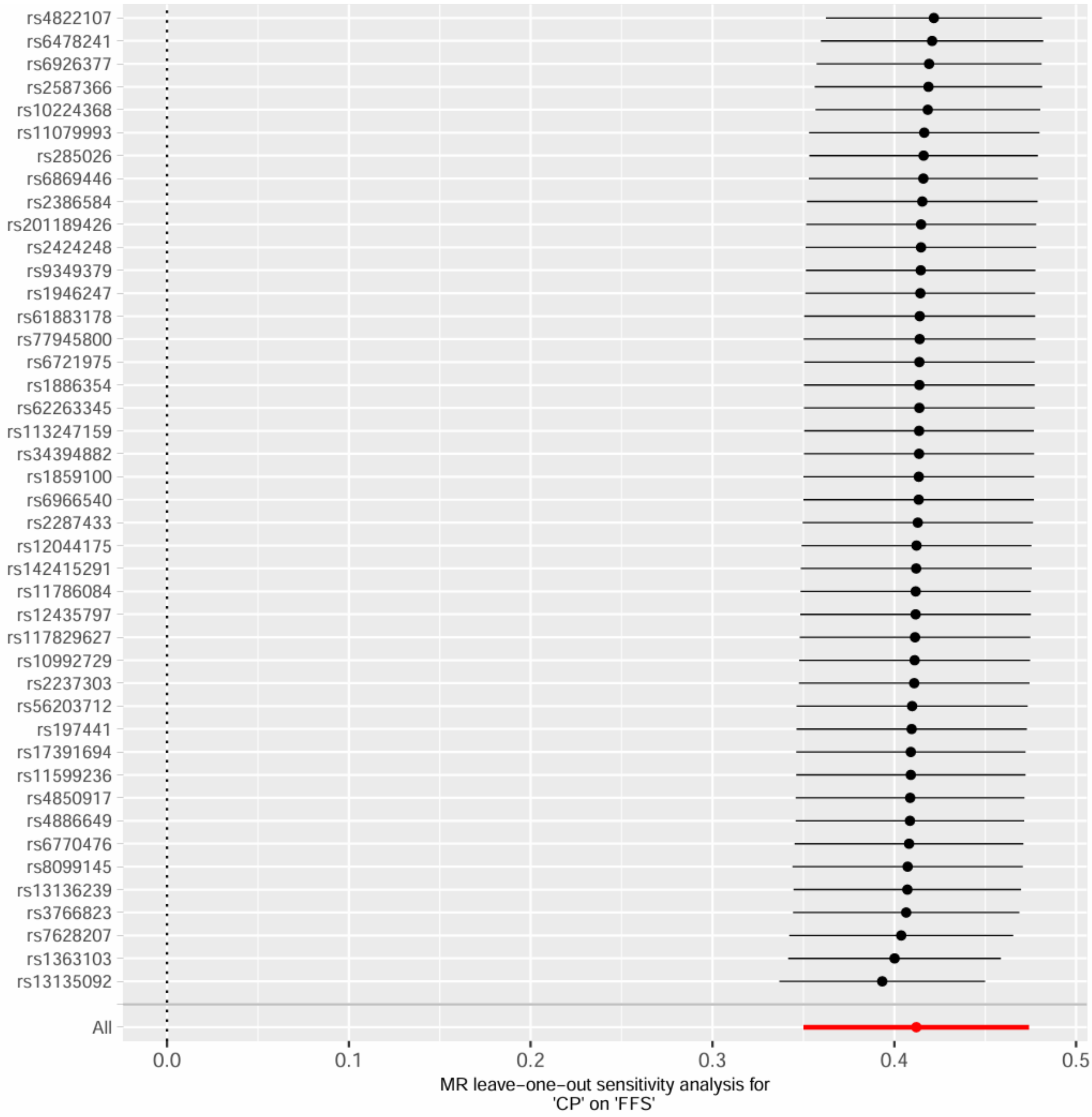

CP and FFS, excluding MHC region

Supplementary Figure 4. Leave-one-out sensitivity analysis for CP and FFS, excluding MHC region.

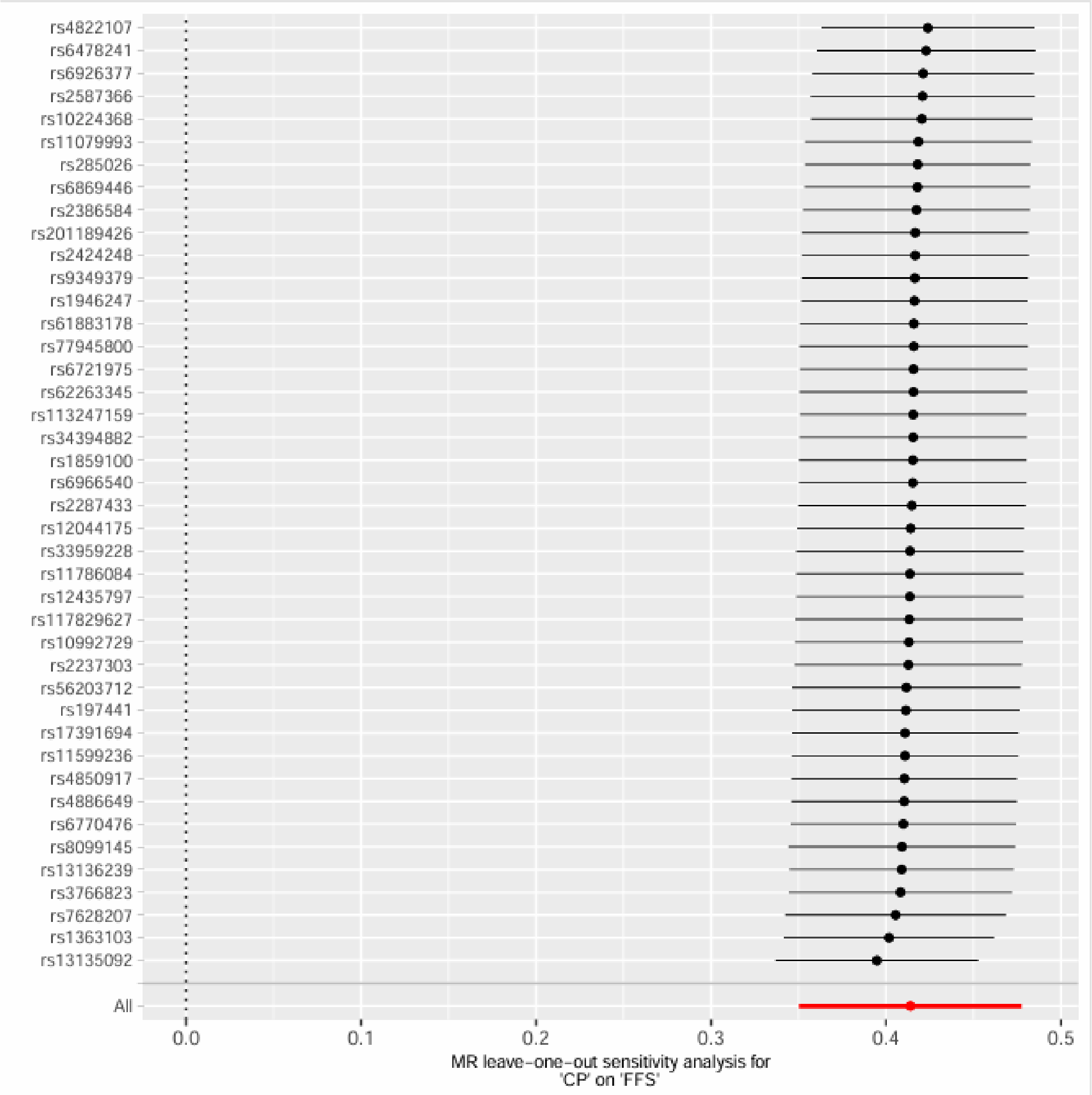

CP and GF

Supplementary Figure 5. Leave-one-out sensitivity analysis for CP and GF.

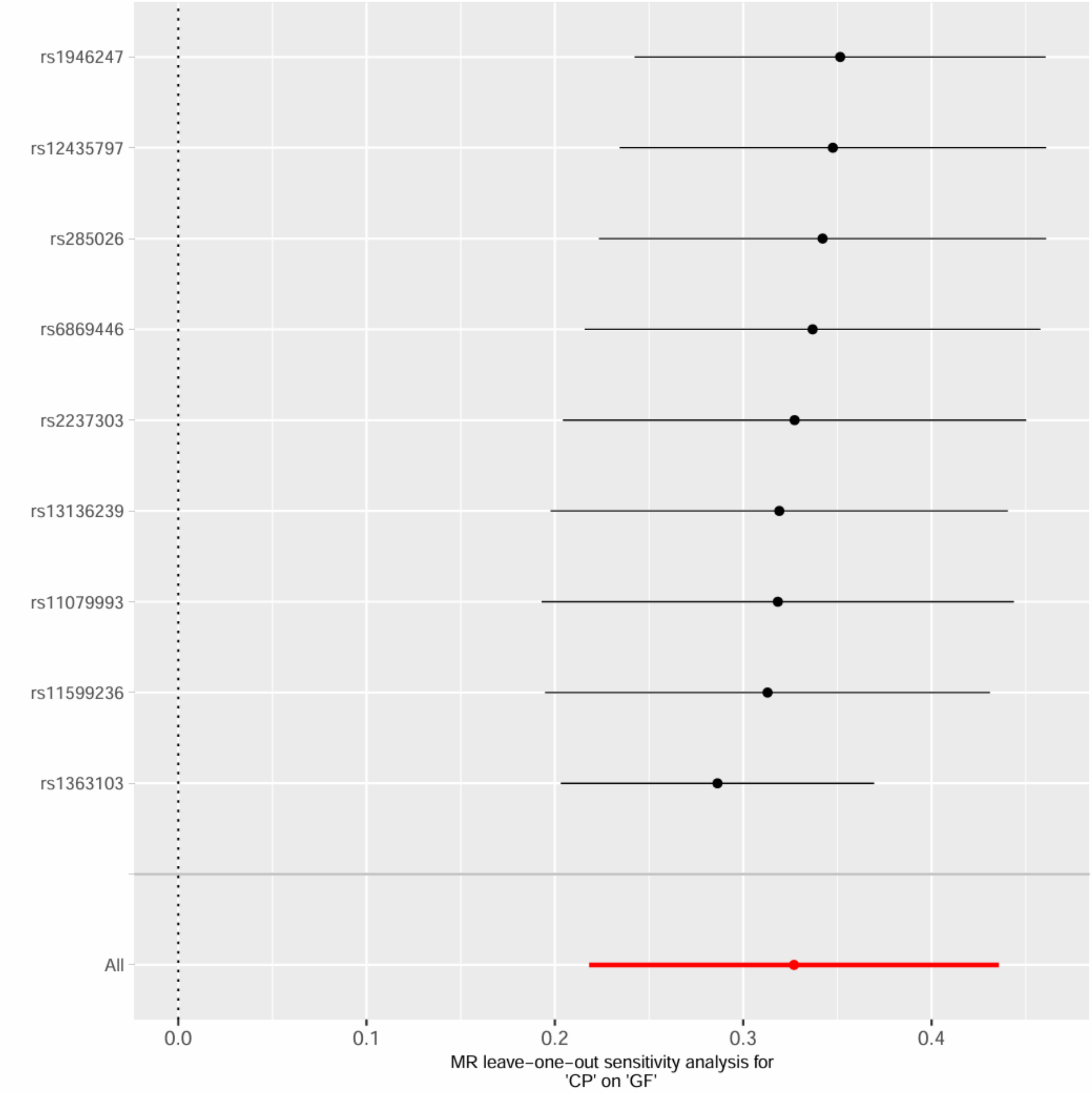

CP and GF, excluding MHC region

Supplementary Figure 6. Leave-one-out sensitivity analysis for CP and GF, excluding MHC region.

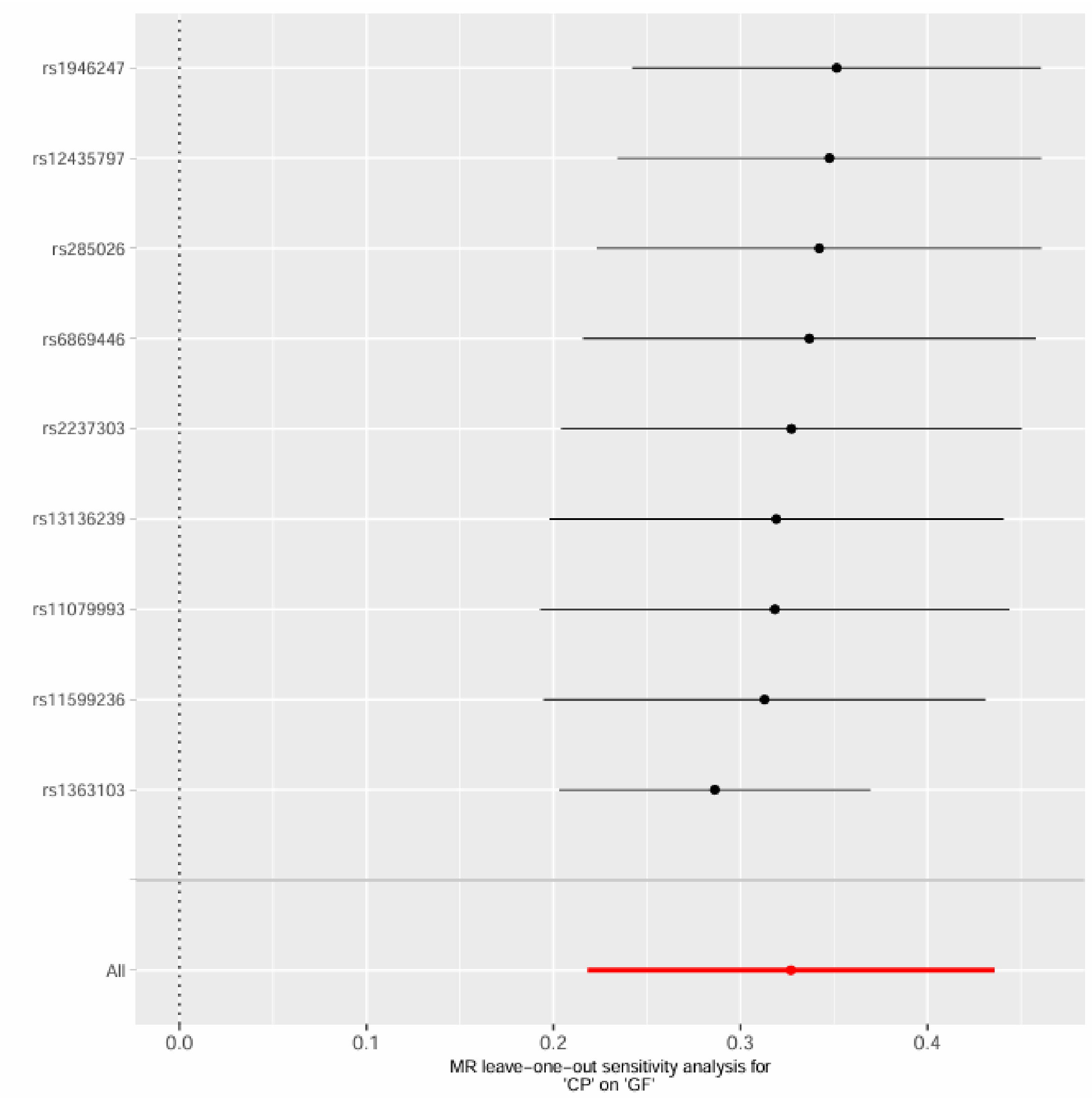


CP and F1

Supplementary Figure 7. Leave-one-out sensitivity analysis for CP and F1.

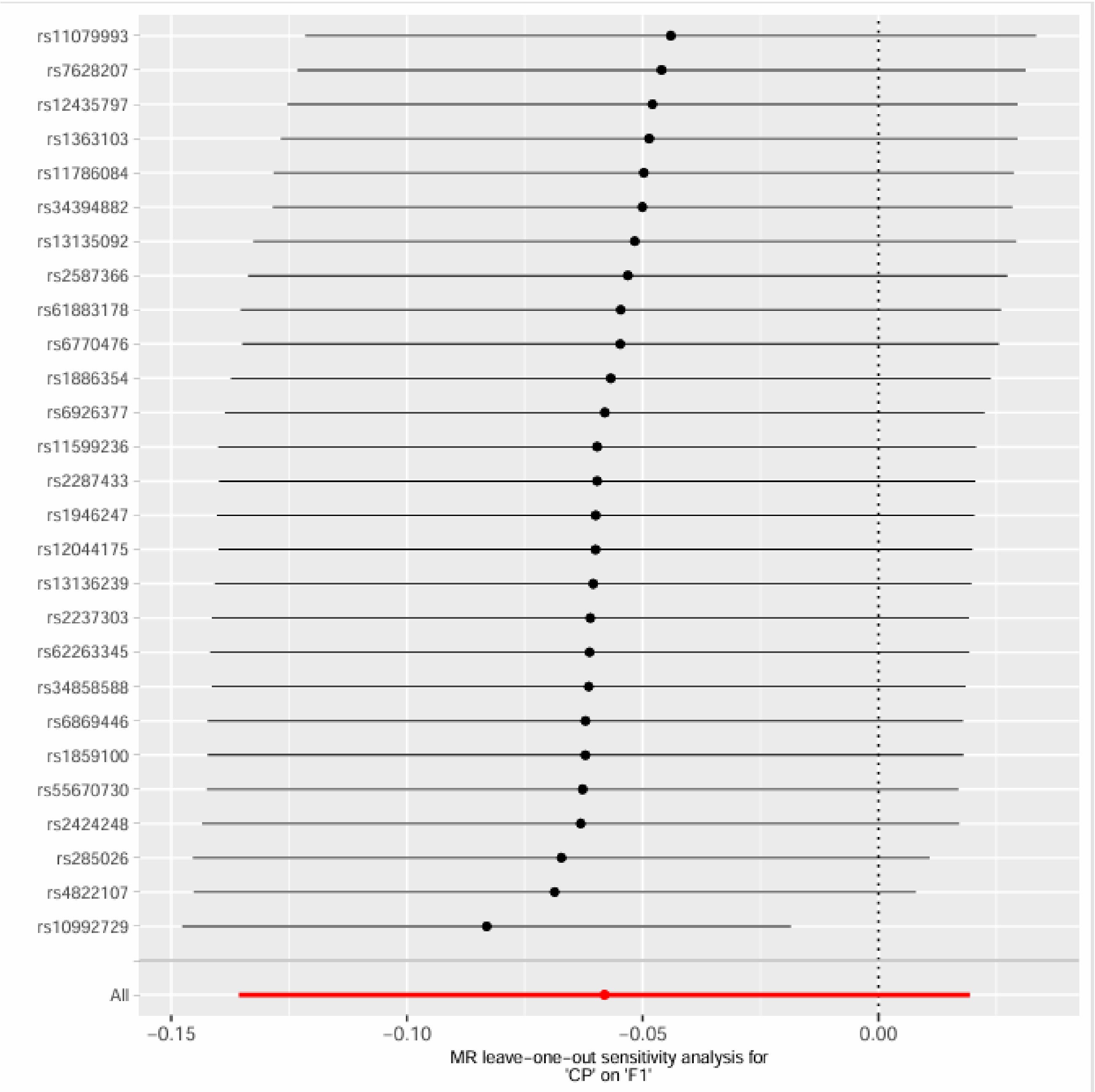

CP and F1, excluding MHC region

Supplementary Figure 8. Leave-one-out sensitivity analysis for CP and F1, excluding MHC region.

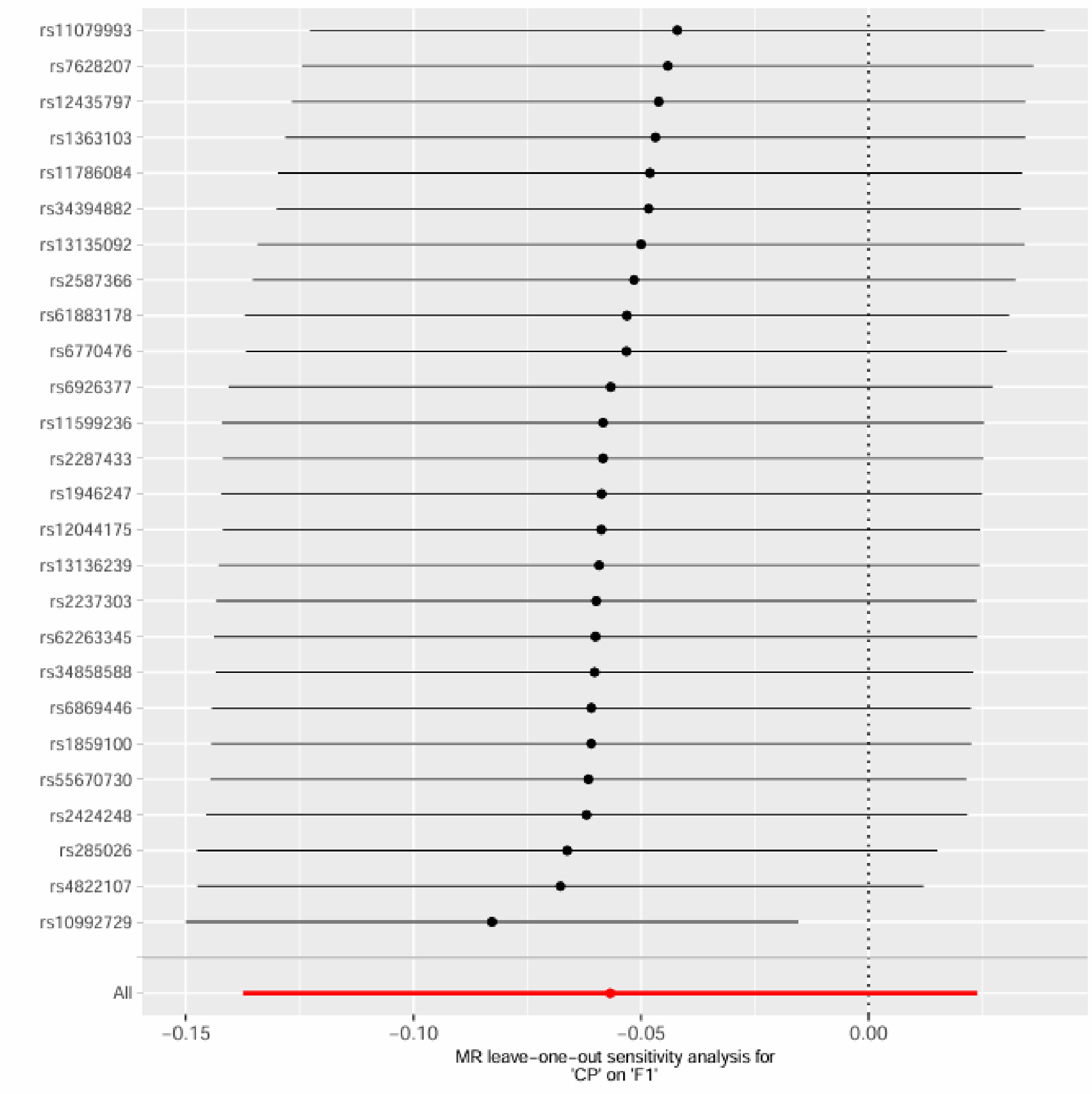

CP and F2

Supplementary Figure 9. Leave-one-out sensitivity analysis for CP and F2.

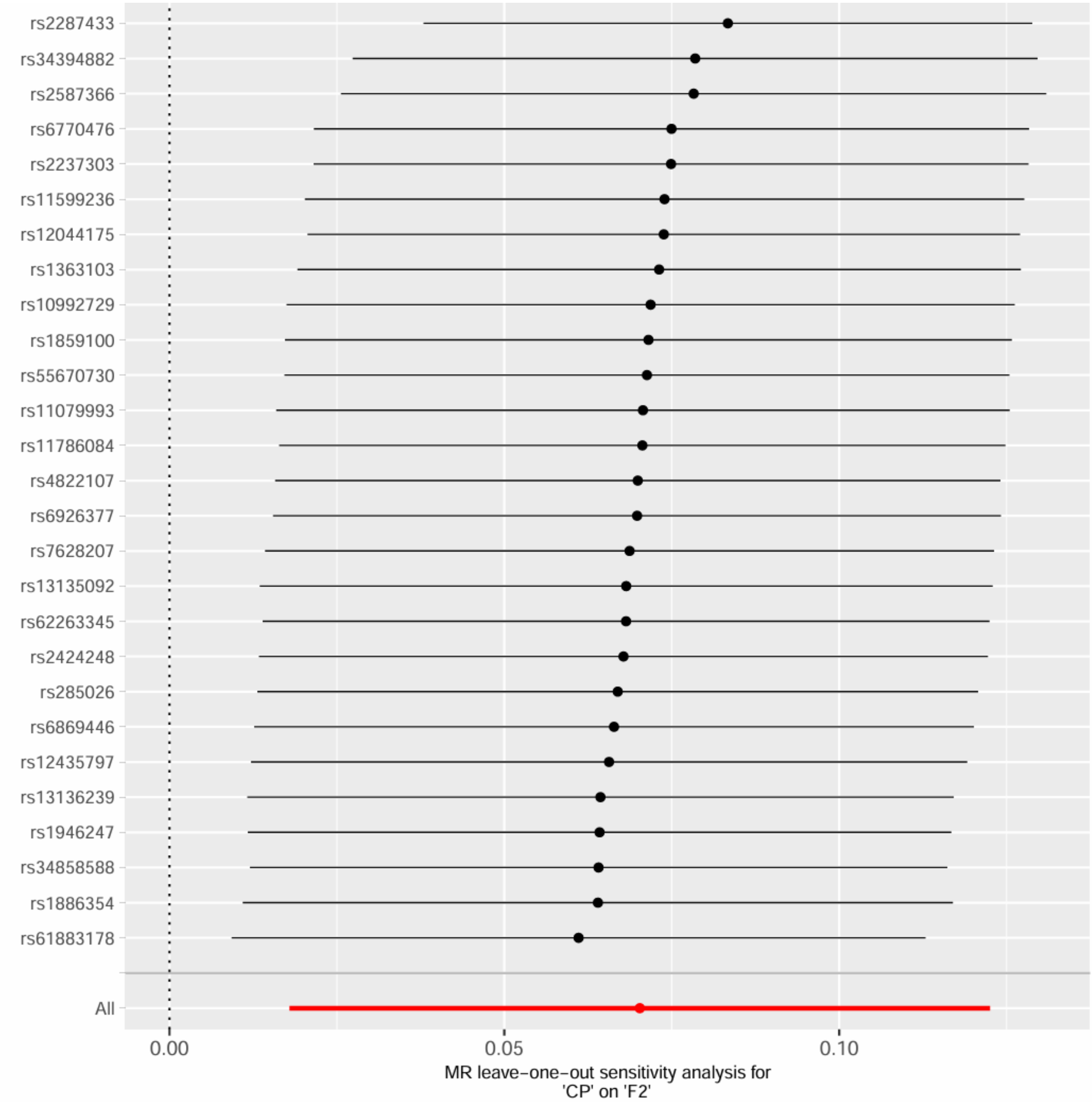

CP and F2, excluding MHC region

Supplementary Figure 10. Leave-one-out sensitivity analysis for CP and F2, excluding MHC region.

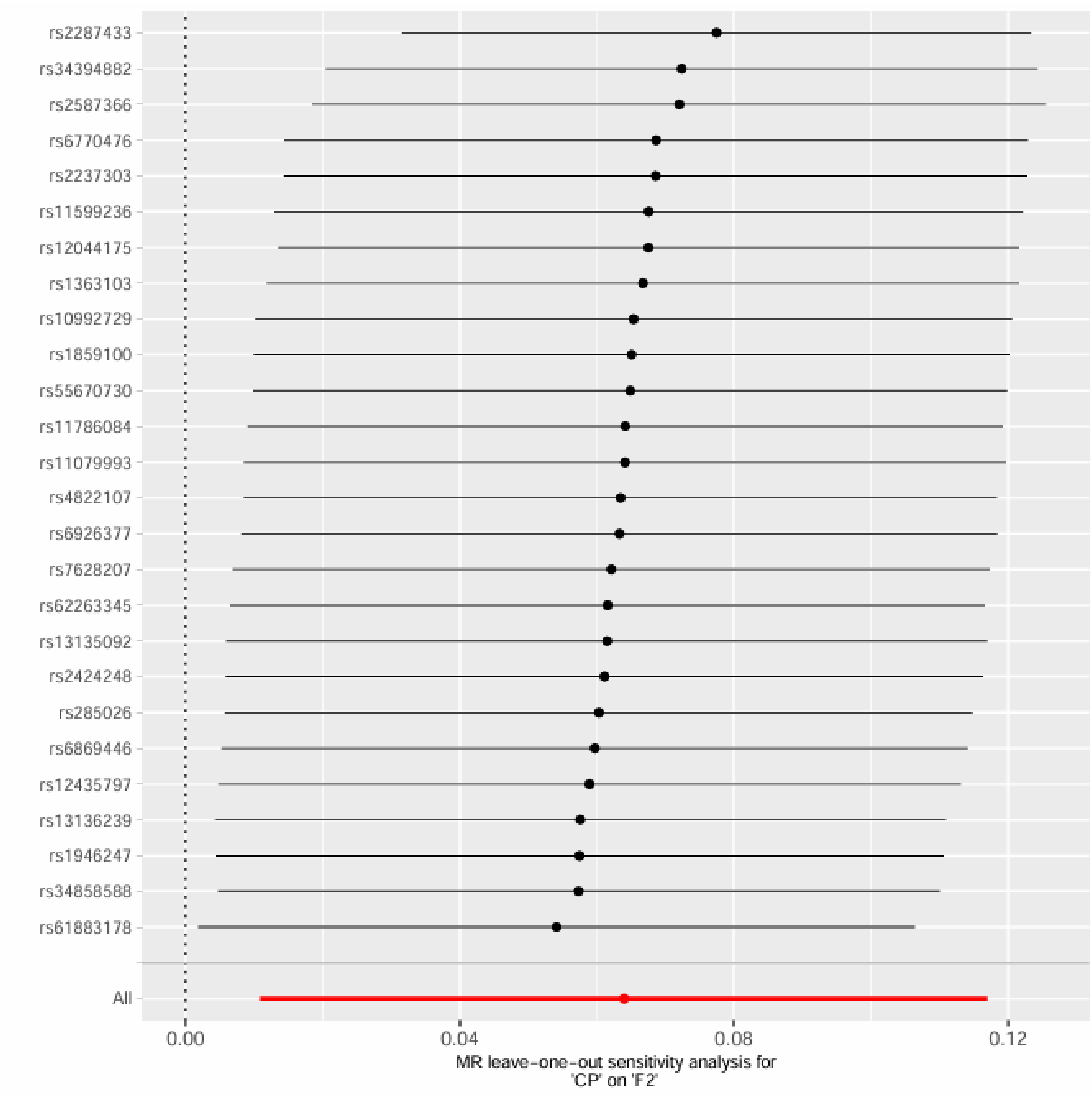

CP and F3

Supplementary Figure 11. Leave-one-out sensitivity analysis for CP and F3.

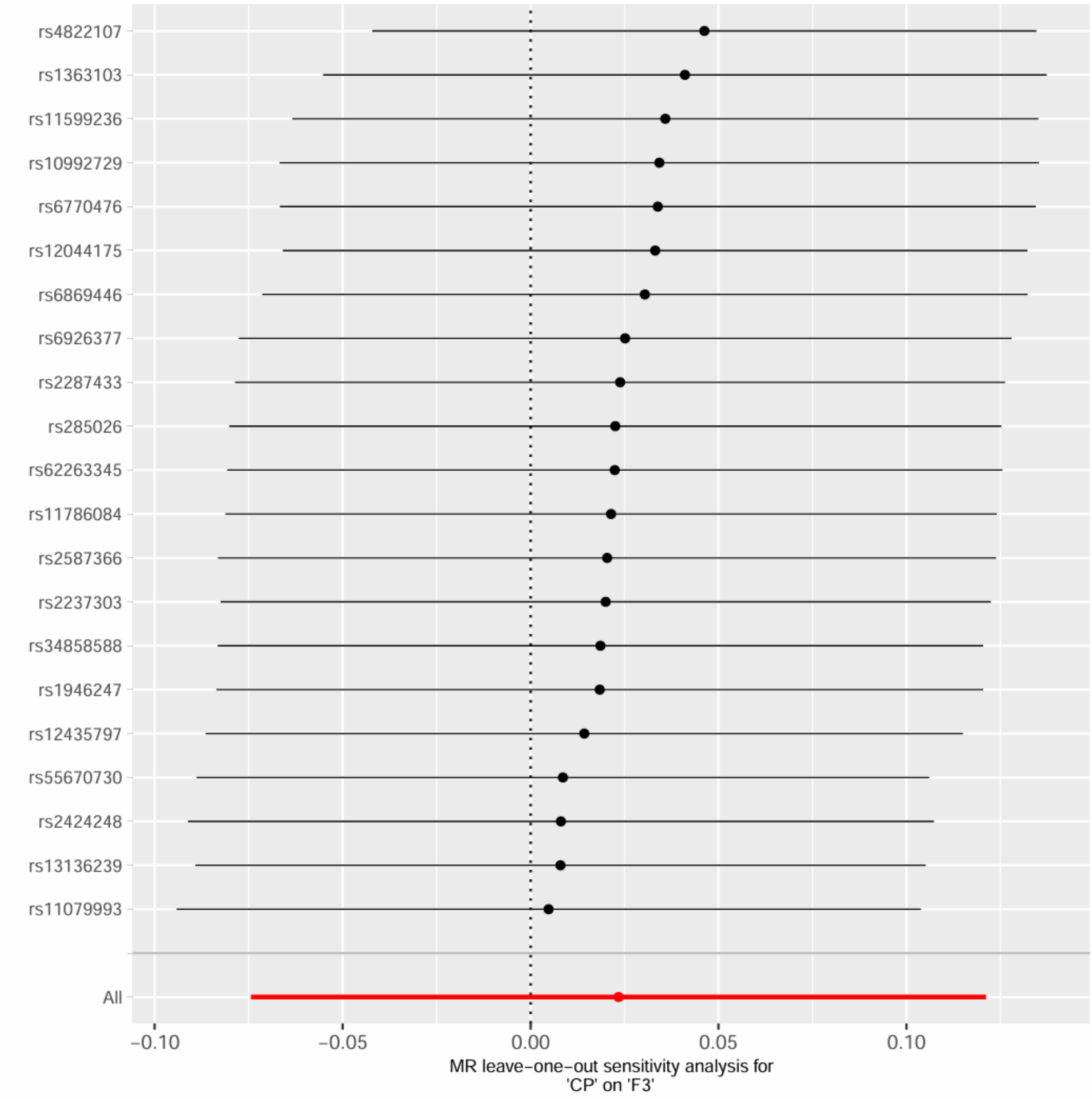

CP and F3, excluding MHC region

Supplementary Figure 12. Leave-one-out sensitivity analysis for CP and F3, excluding MHC region.

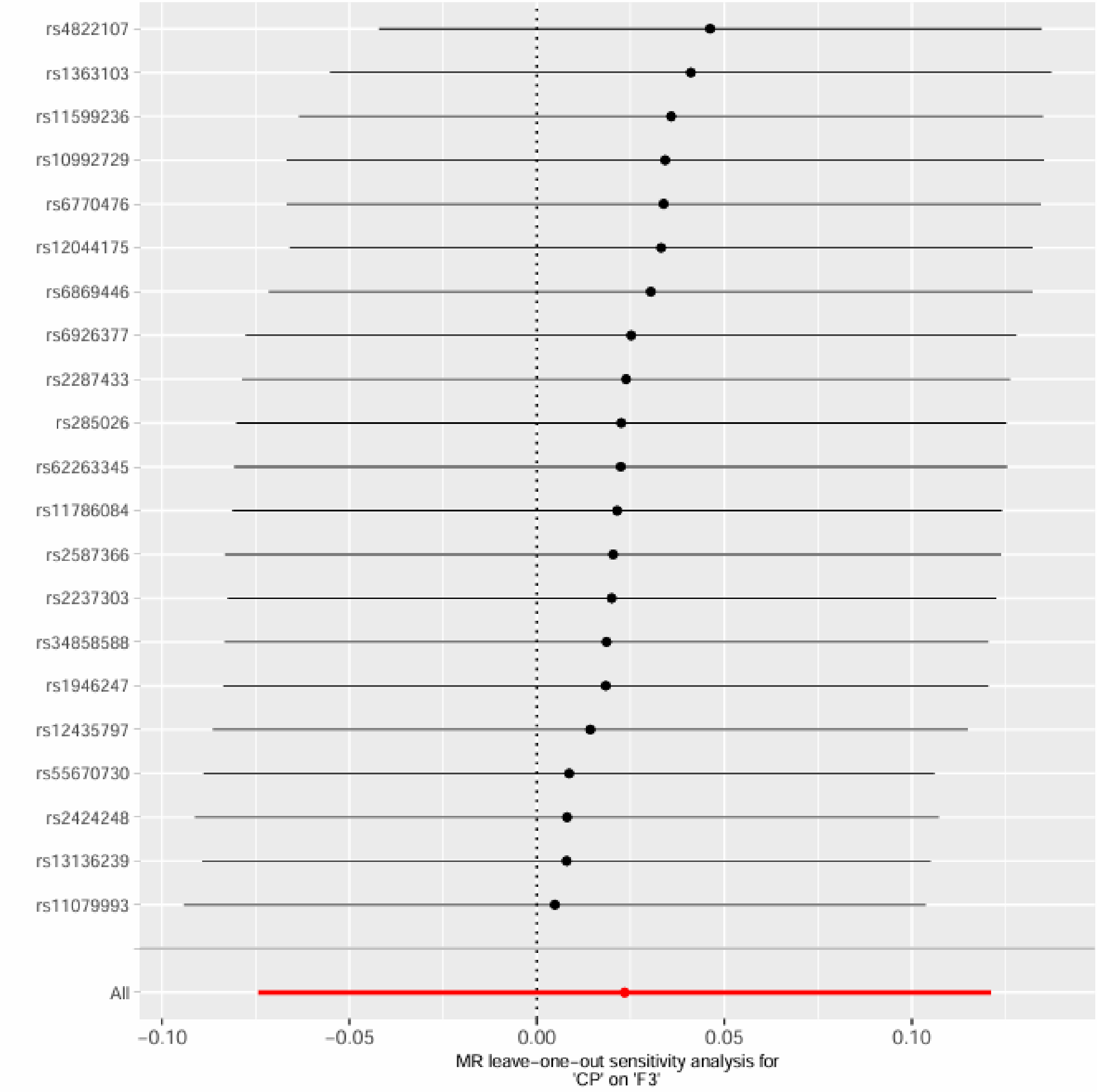

CP and F4

Supplementary Figure 13. Leave-one-out sensitivity analysis for CP and F4.

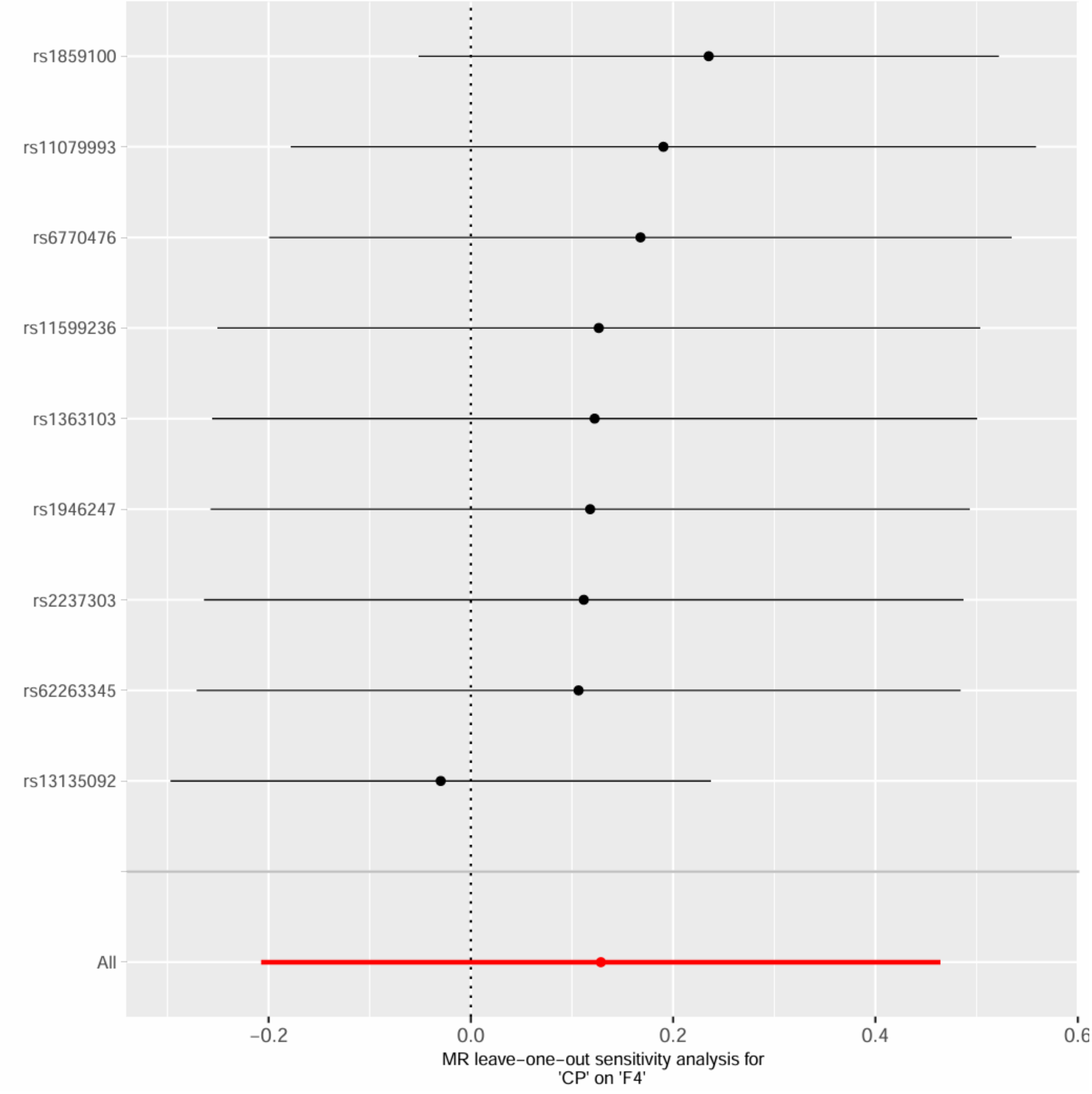

CP and F4, excluding MHC region

Supplementary Figure 14. Leave-one-out sensitivity analysis for CP and F4, excluding MHC region.

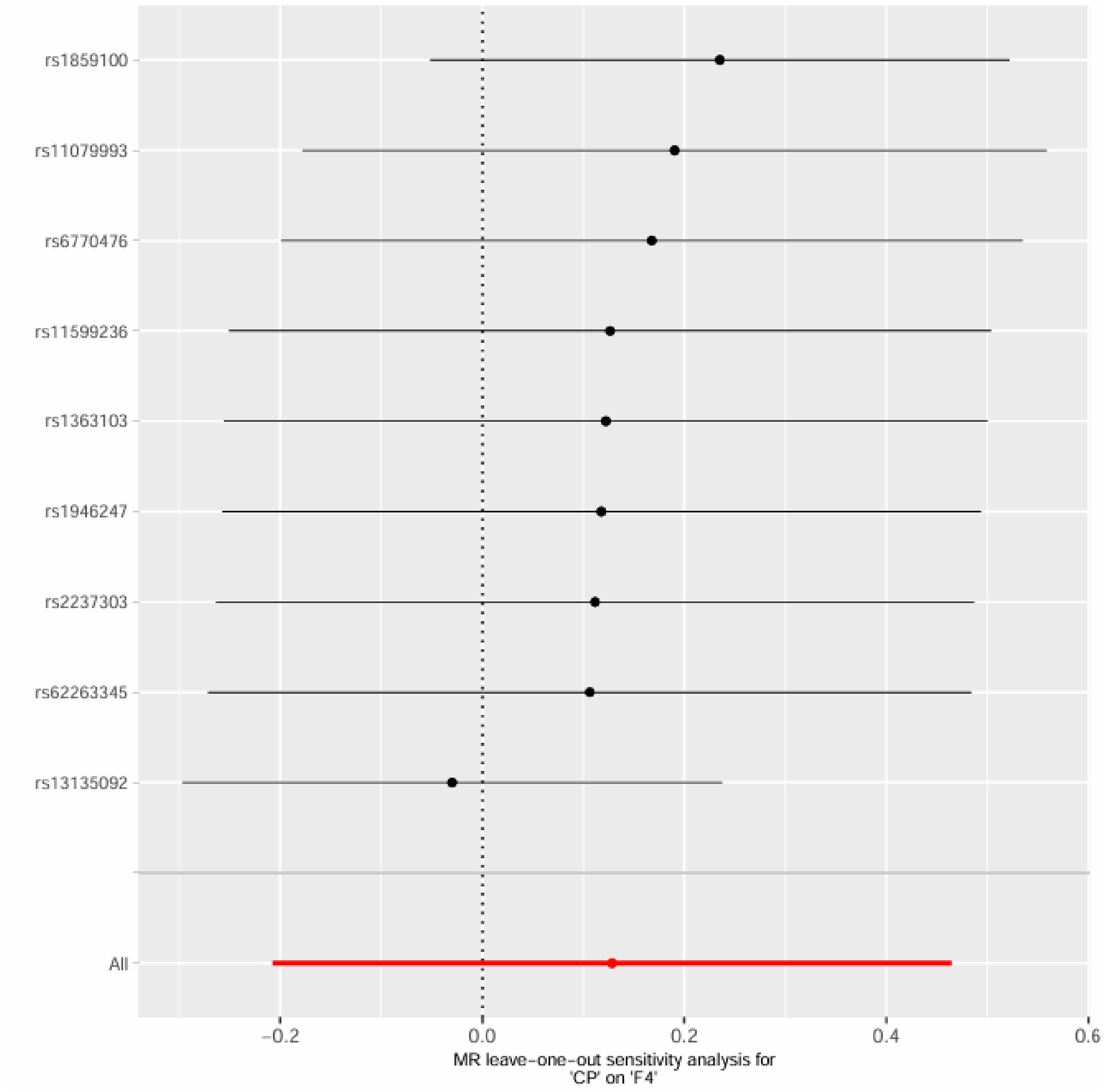

rs1859100
rs11079993
rs6770476
rs11599236
rs1363103
rs1946247
rs2237303
rs62263345
rs13135092
All
−0.2
0.0
0.2
0.4
0.6
MR leave−one−out sensitivity analysis for
'CP' on 'F4'

CP and F5

Supplementary Figure 15. Leave-one-out sensitivity analysis for CP and F5.

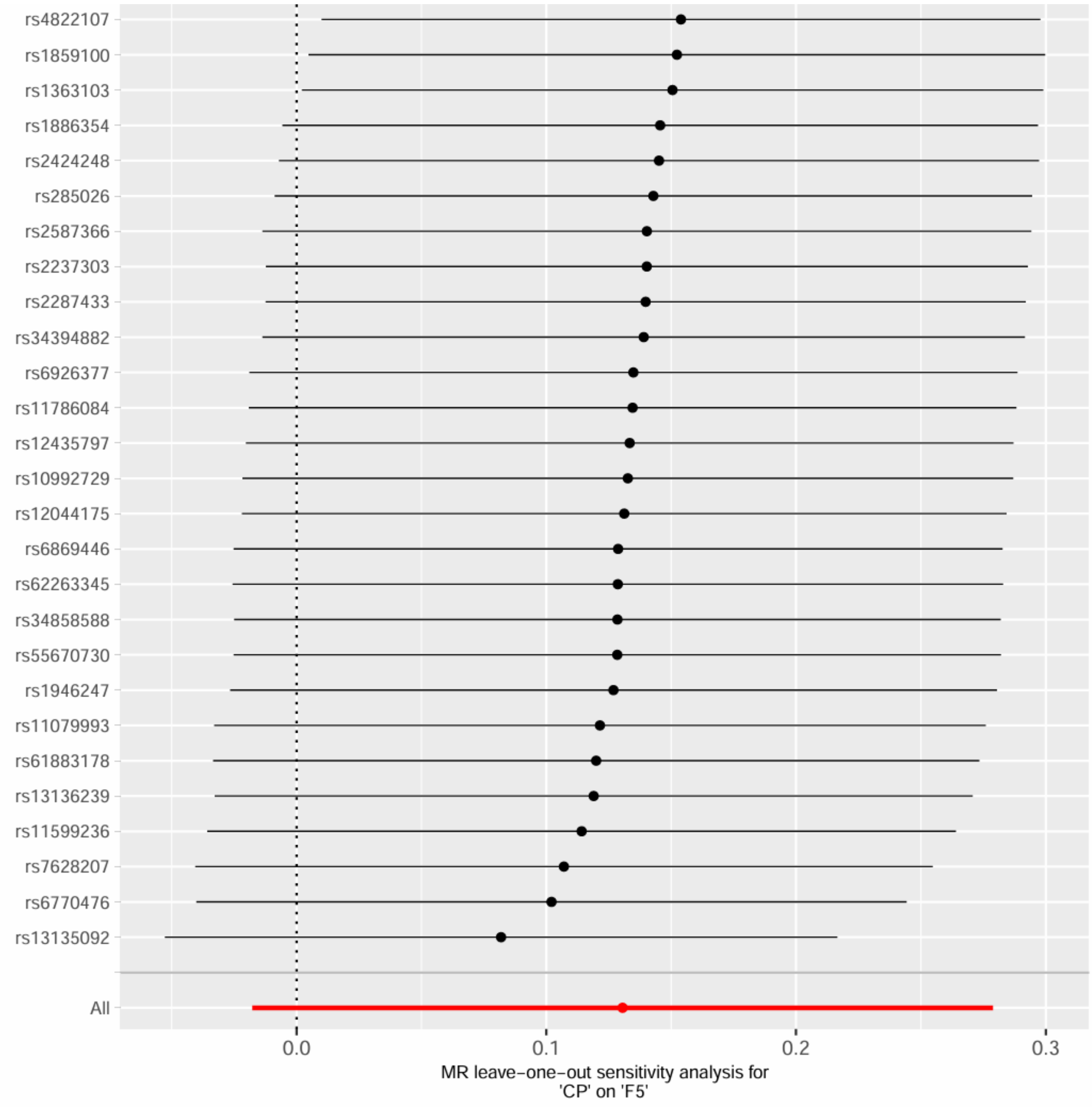

CP and F5, excluding MHC region

Supplementary Figure 16. Leave-one-out sensitivity analysis for CP and F5, excluding MHC region.

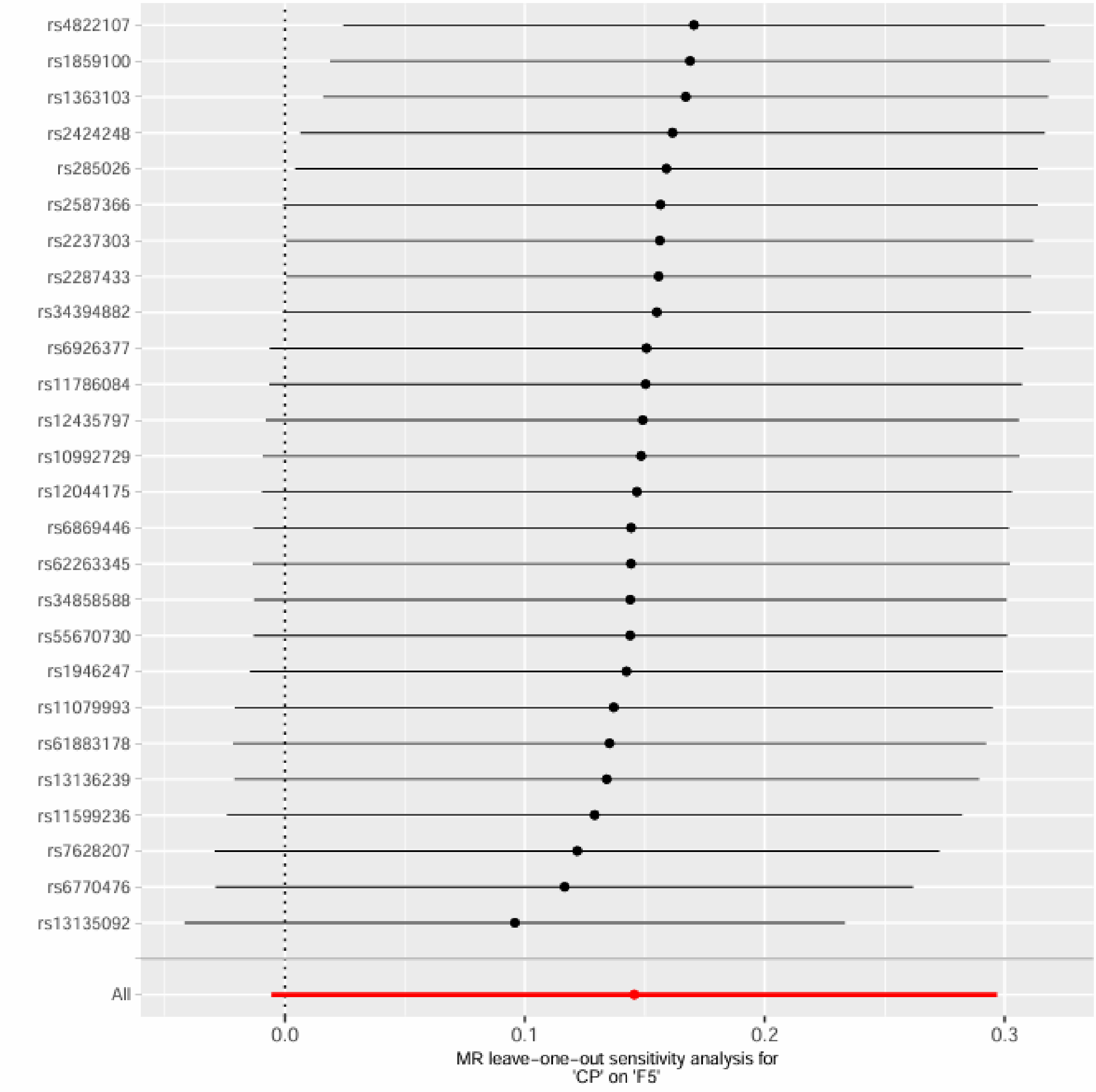

CP and F6

Supplementary Figure 17. Leave-one-out sensitivity analysis for CP and F6.

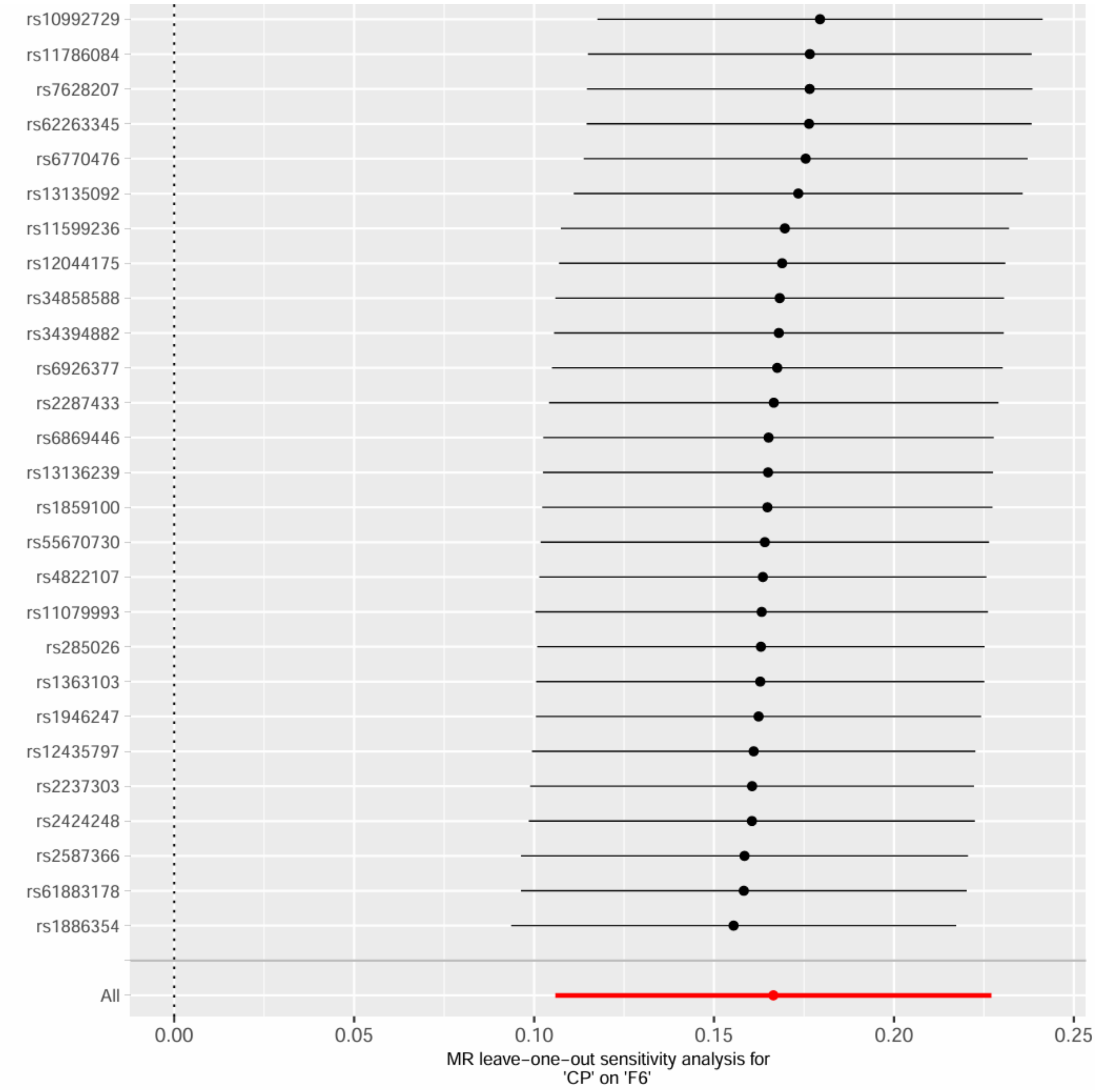

CP and F6, excluding MHC region

Supplementary Figure 18. Leave-one-out sensitivity analysis for CP and F6, excluding MHC region.

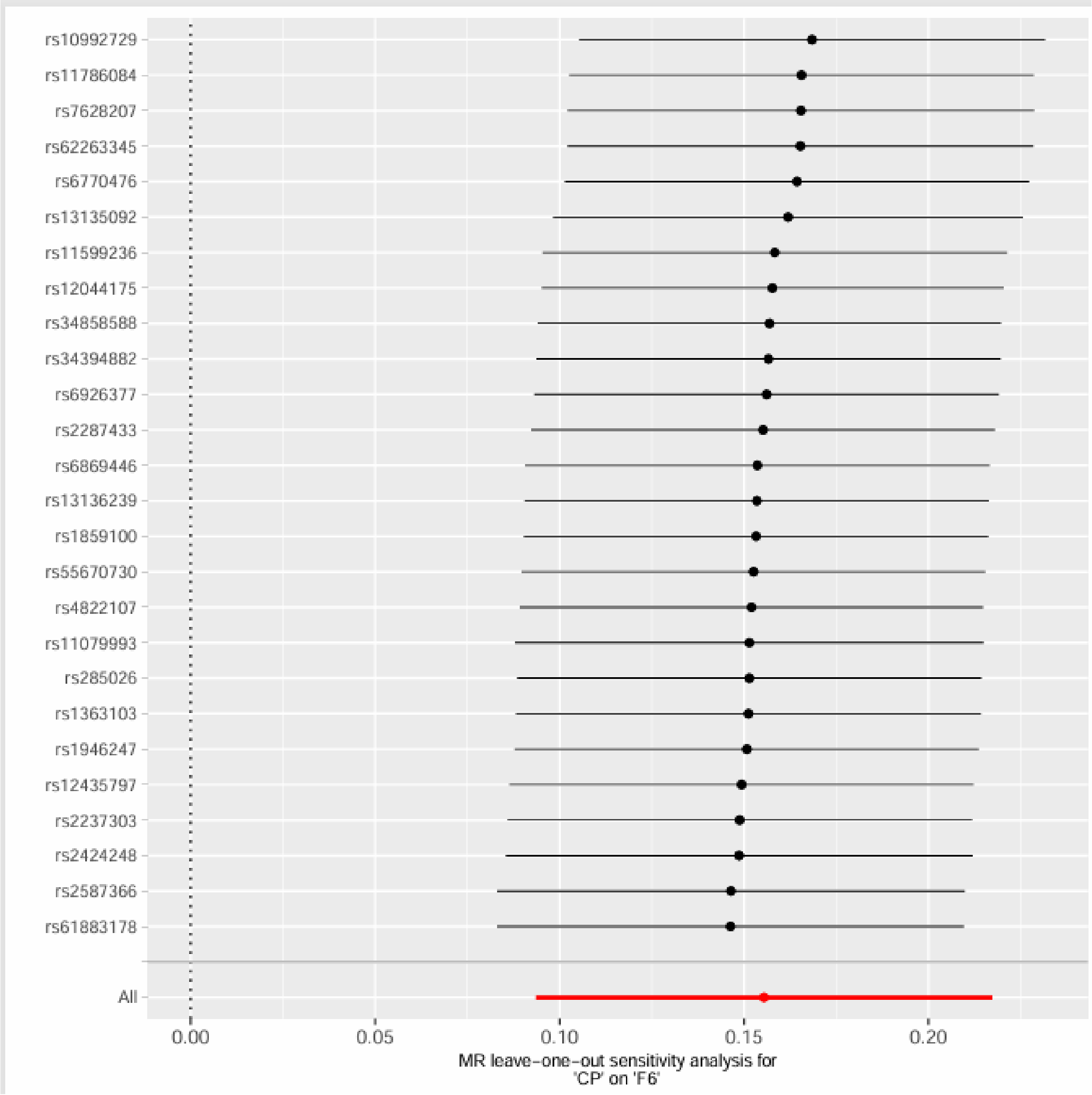

Rheumatoid Arthritis Exposure on Frailty Outcomes

RA and FI

Supplementary Figure 19. Leave-one-out sensitivity analysis for RA and FI.

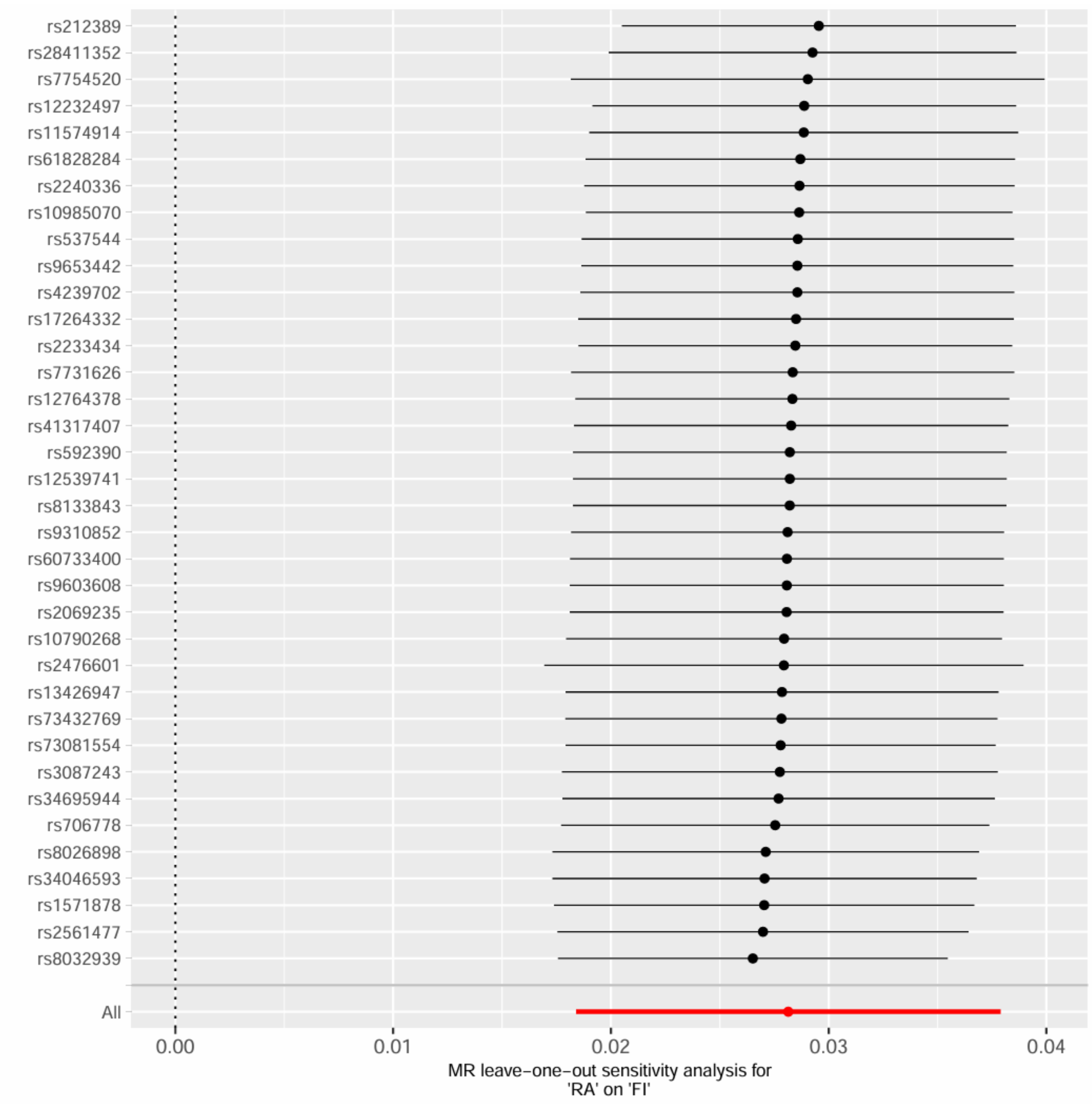

RA and FI, excluding MHC region

Supplementary Figure 20. Leave-one-out sensitivity analysis for RA and FI, excluding MHC region.

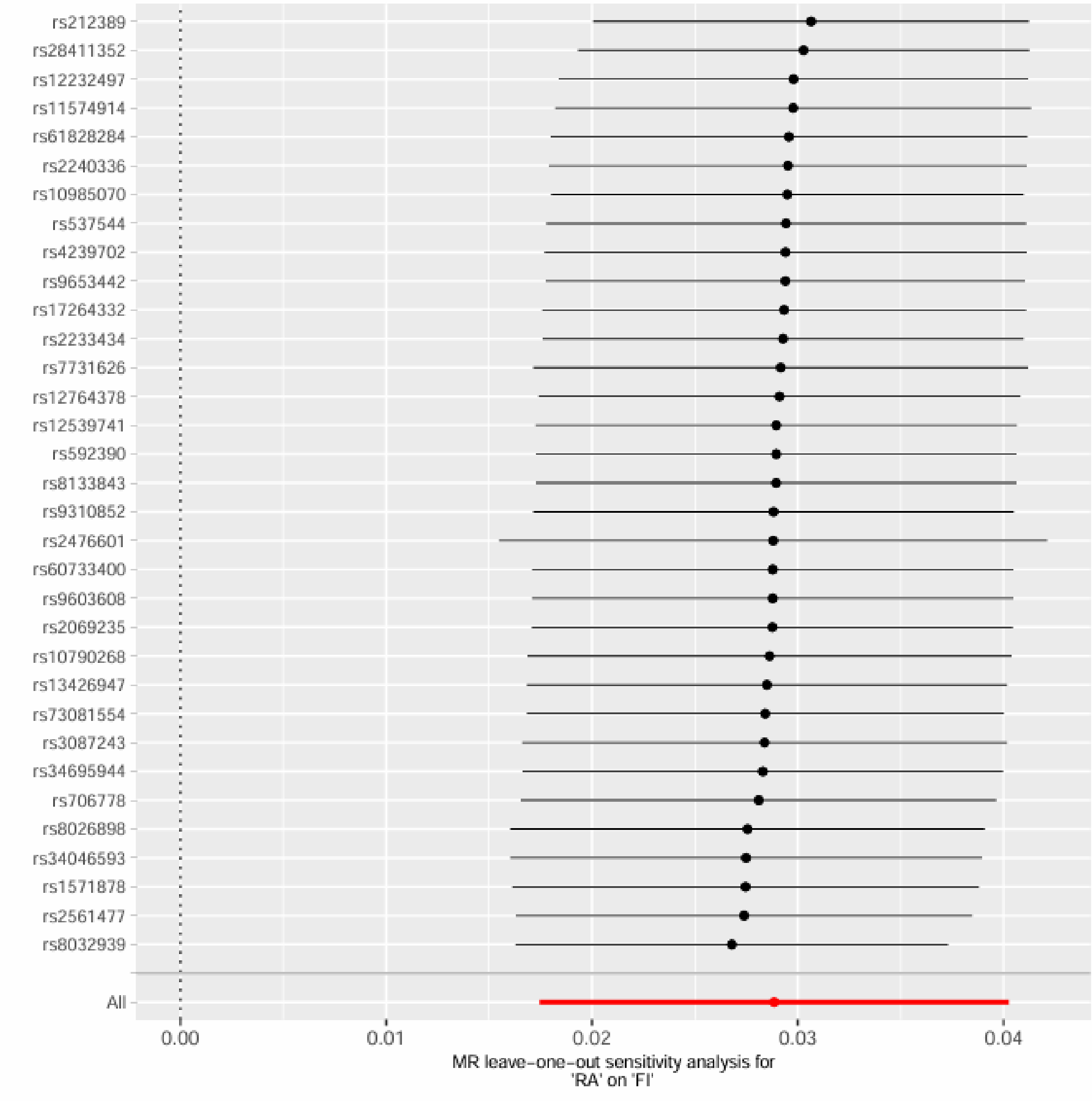

RA and FFS

Supplementary Figure 21. Leave-one-out sensitivity analysis for RA and FFS.

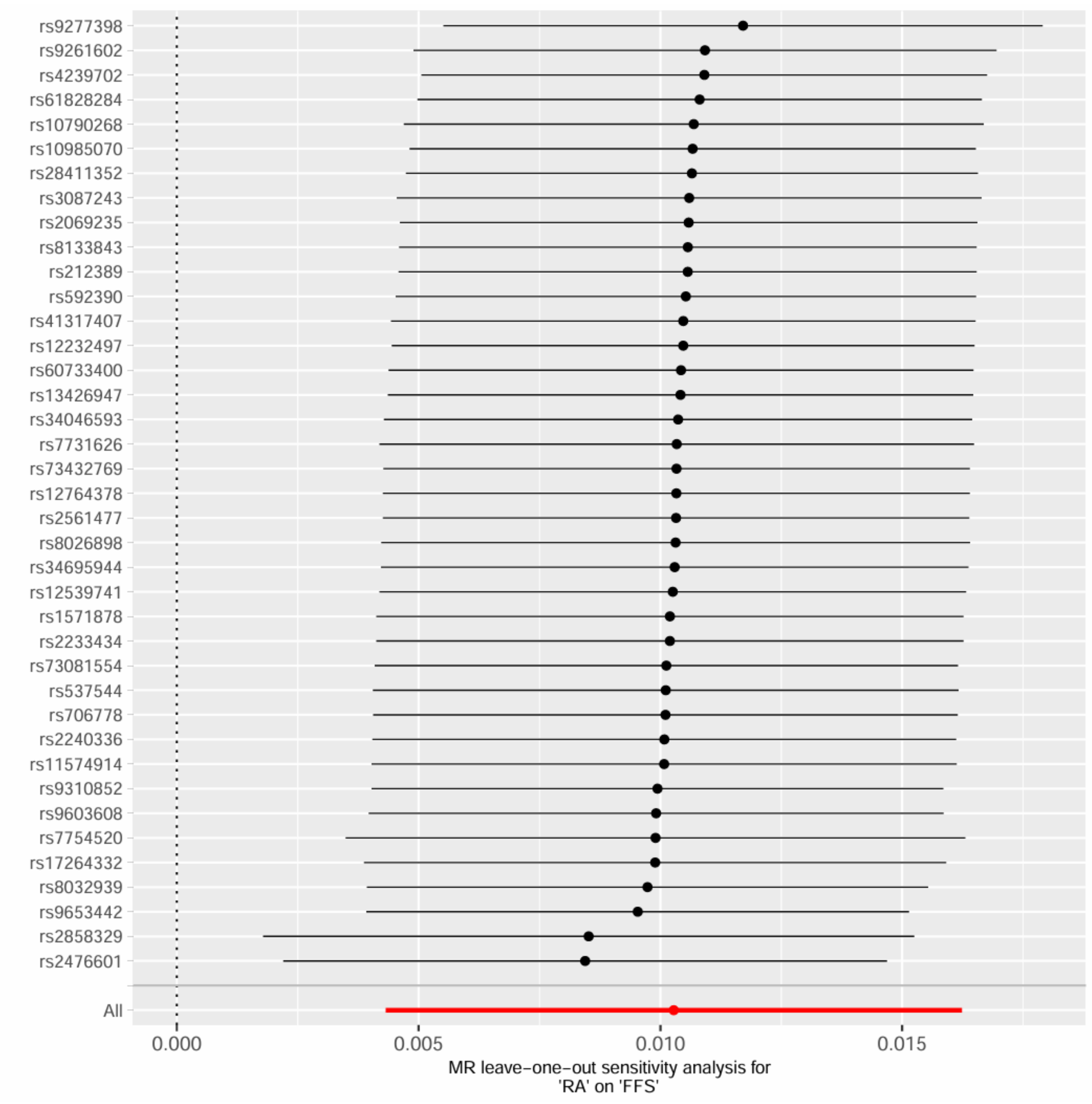

RA and FFS, excluding MHC region

Supplementary Figure 22. Leave-one-out sensitivity analysis for RA and FFS, excluding MHC region.

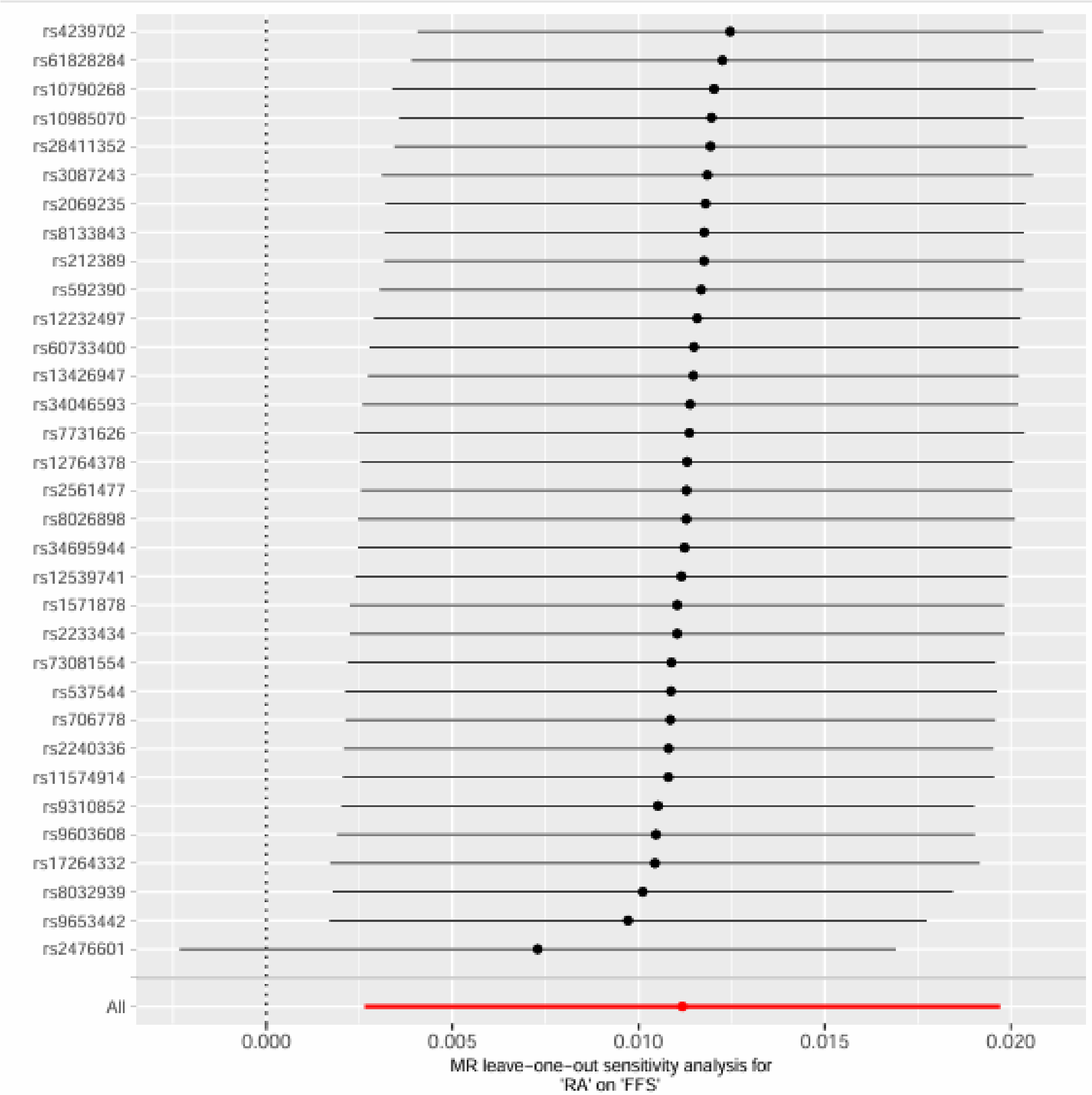

rs4239702
rs61828284
rs10790268
rs10985070
rs28411352
rs3087243
rs2069235
rs8133843
rs212389
rs592390
rs12232497
rs60733400
rs13426947
rs34046593
rs7731626
rs12764378
rs2561477
rs8026898
rs34695944
rs12539741
rs1571878
rs2233434
rs73081554
rs537544
rs706778
rs2240336
rs11574914
rs9310852
rs9603608
rs17264332
rs8032939
rs9653442
rs2476601
All
0.000
0.005
0.010
0.015
0.020
MR leave-one-out sensitivity analysis for
'RA' on 'FFS'

RA and GF

Supplementary Figure 23. Leave-one-out sensitivity analysis for RA and GF.

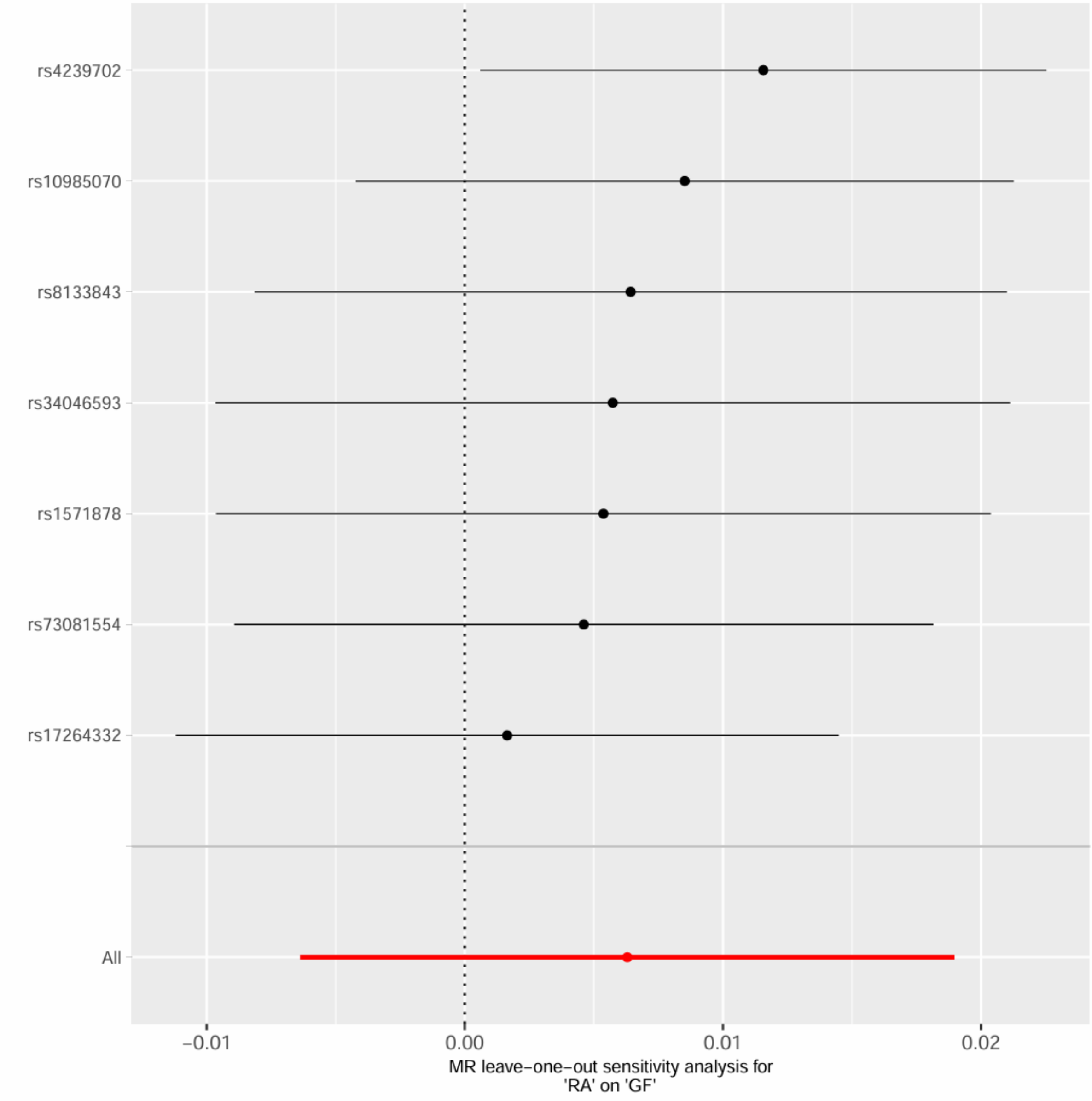

RA and GF, excluding MHC region

Supplementary Figure 24. Leave-one-out sensitivity analysis for RA and GF, excluding MHC region.

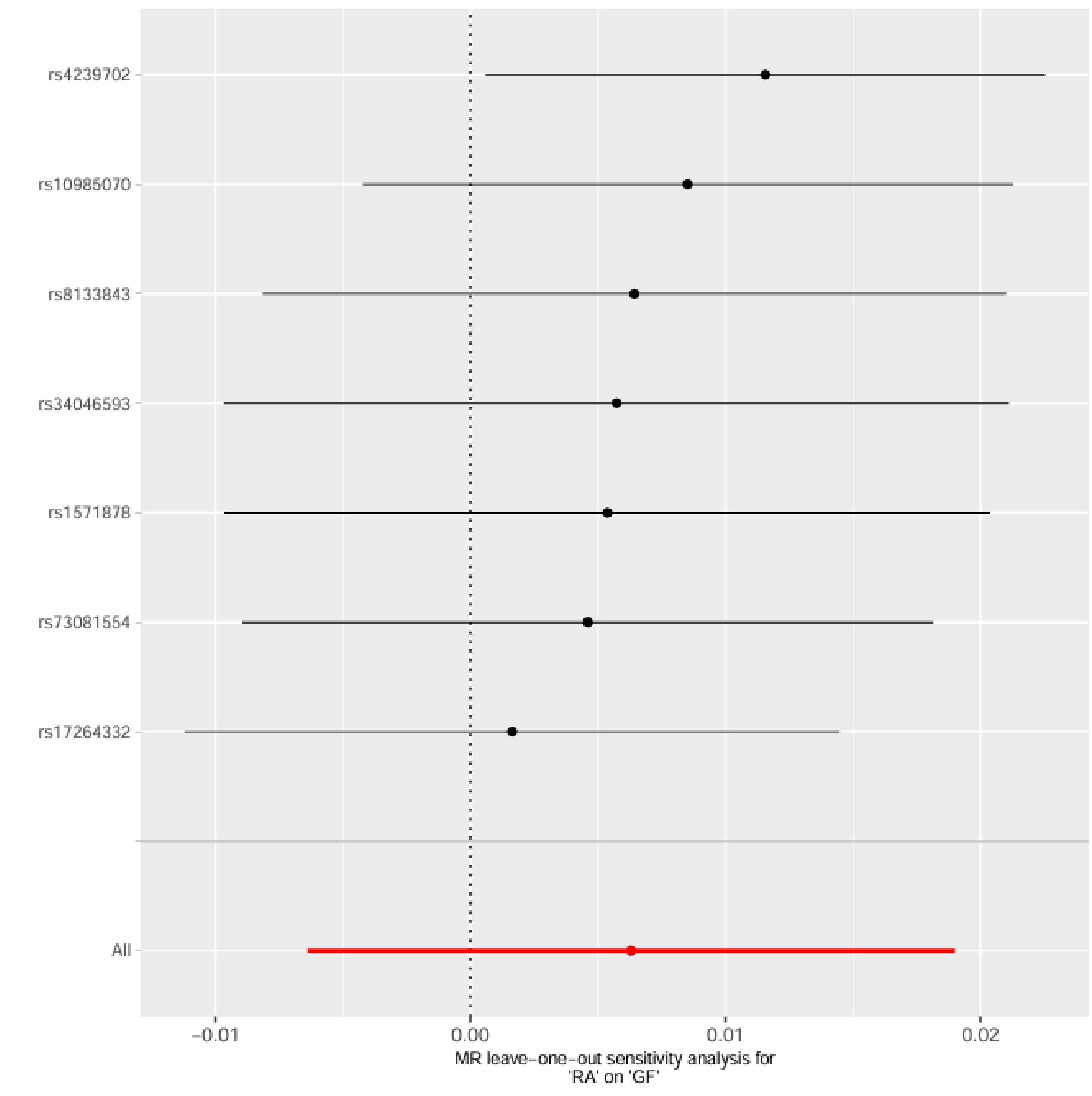

RA and F1

Supplementary Figure 25. Leave-one-out sensitivity analysis for RA and F1.

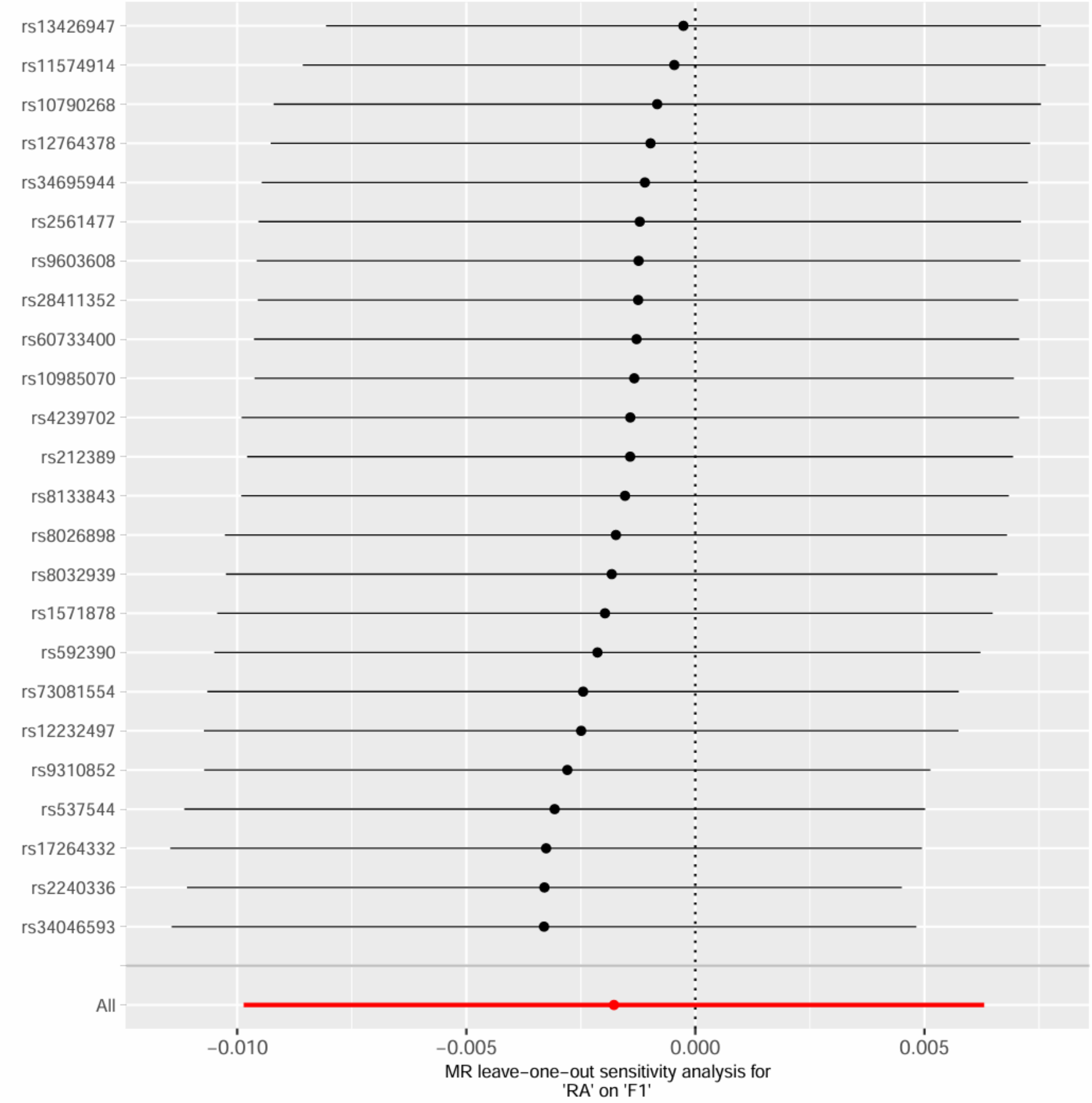

RA and F1, excluding MHC region

Supplementary Figure 26. Leave-one-out sensitivity analysis for RA and F1, excluding MHC region.

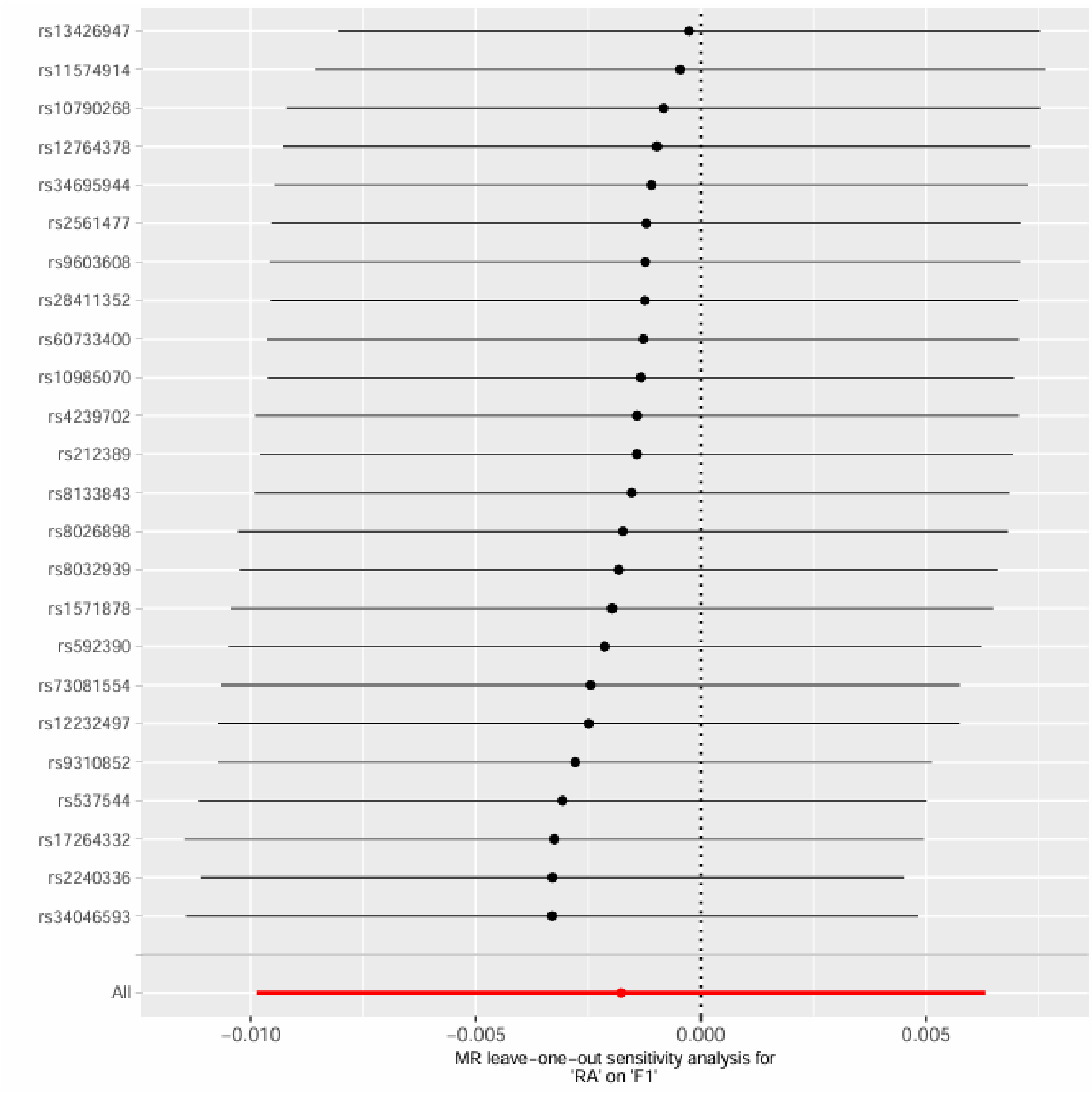

RA and F2

Supplementary Figure 27. Leave-one-out sensitivity analysis for RA and F2.

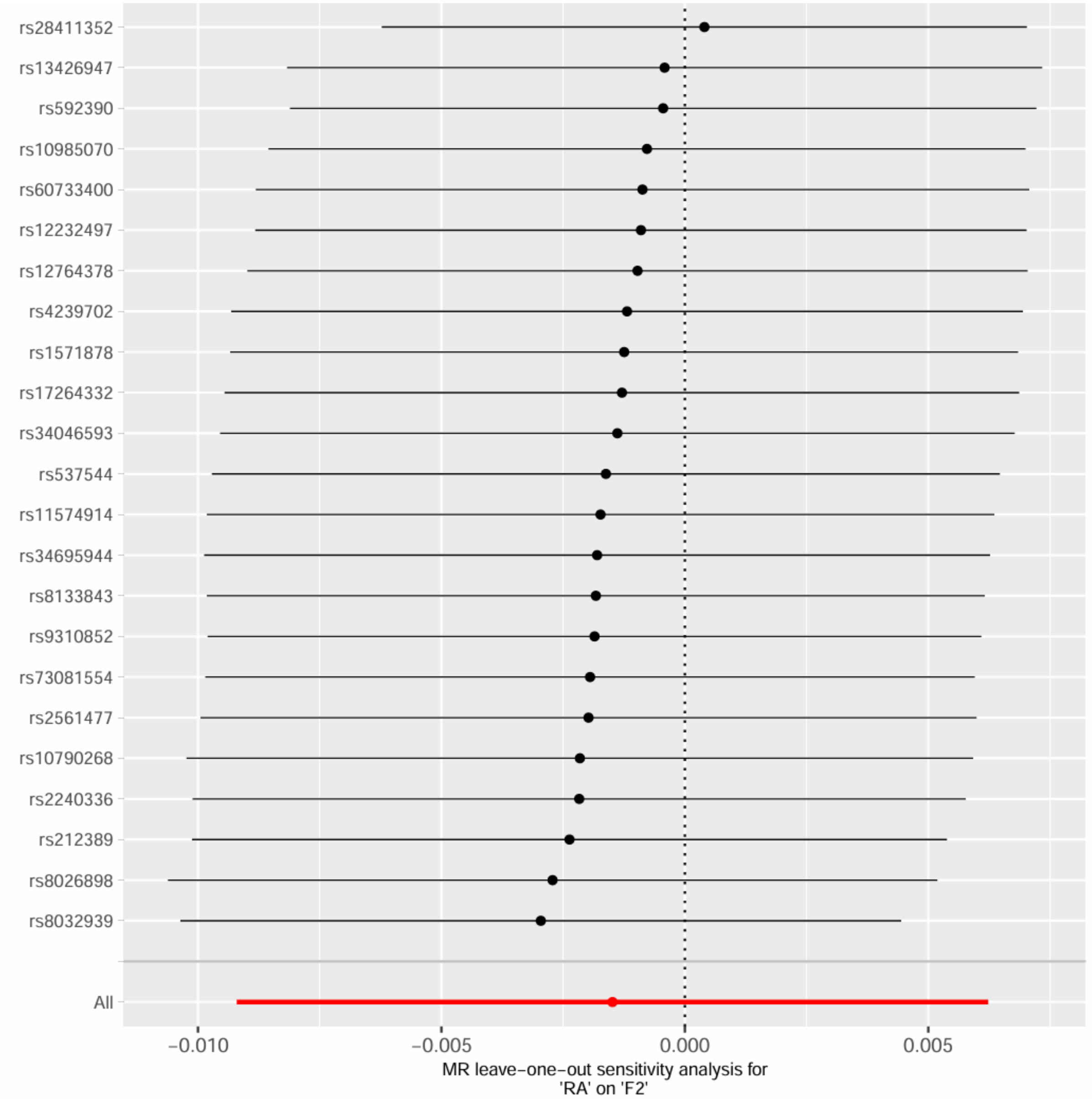

RA and F2, excluding MHC region

Supplementary Figure 28. Leave-one-out sensitivity analysis for RA and F2, excluding MHC region.

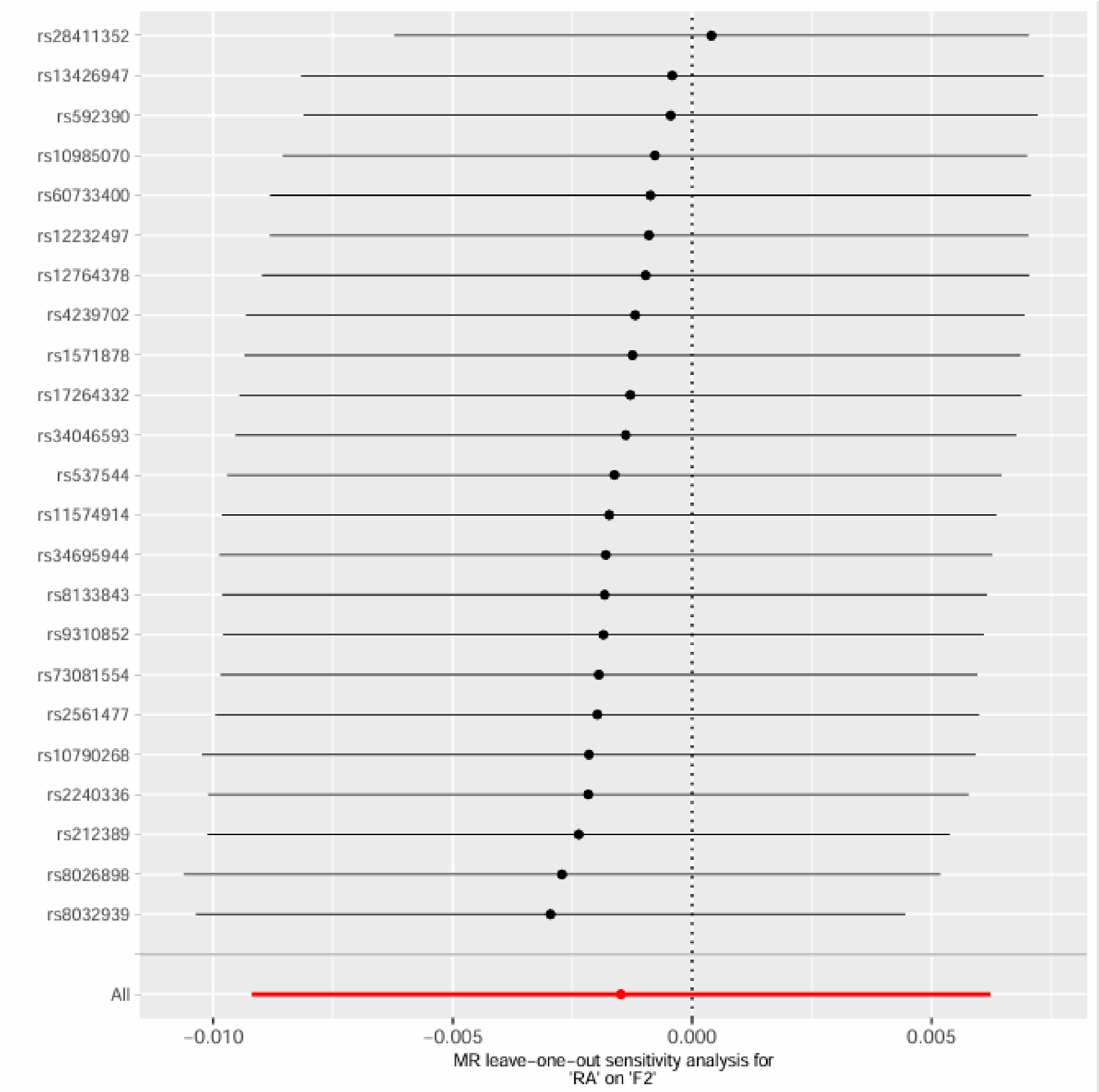

RA and F3

Supplementary Figure 29. Leave-one-out sensitivity analysis for RA and F3.

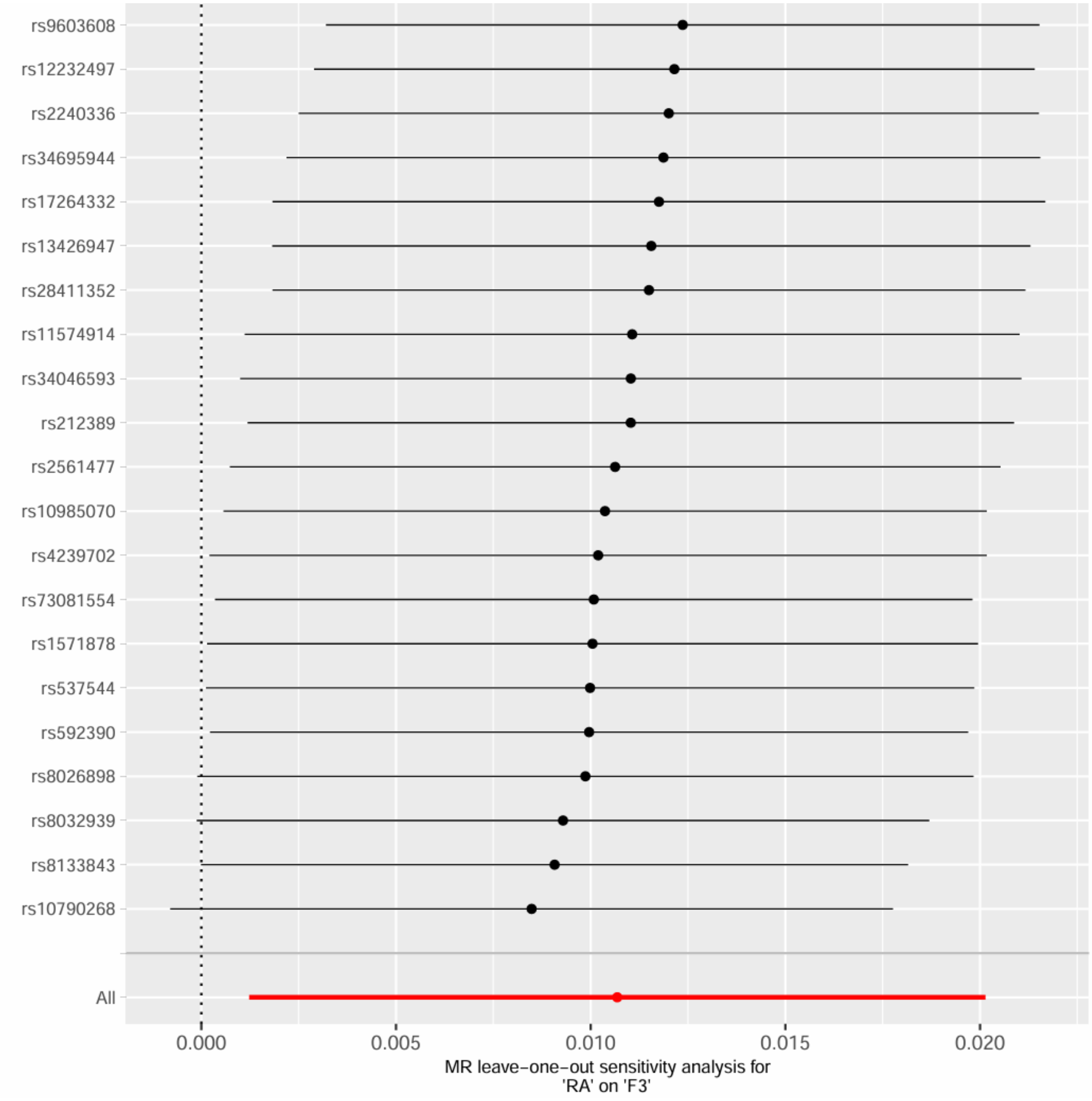

RA and F3, excluding MHC region

Supplementary Figure 30. Leave-one-out sensitivity analysis for RA and F3, excluding MHC region.

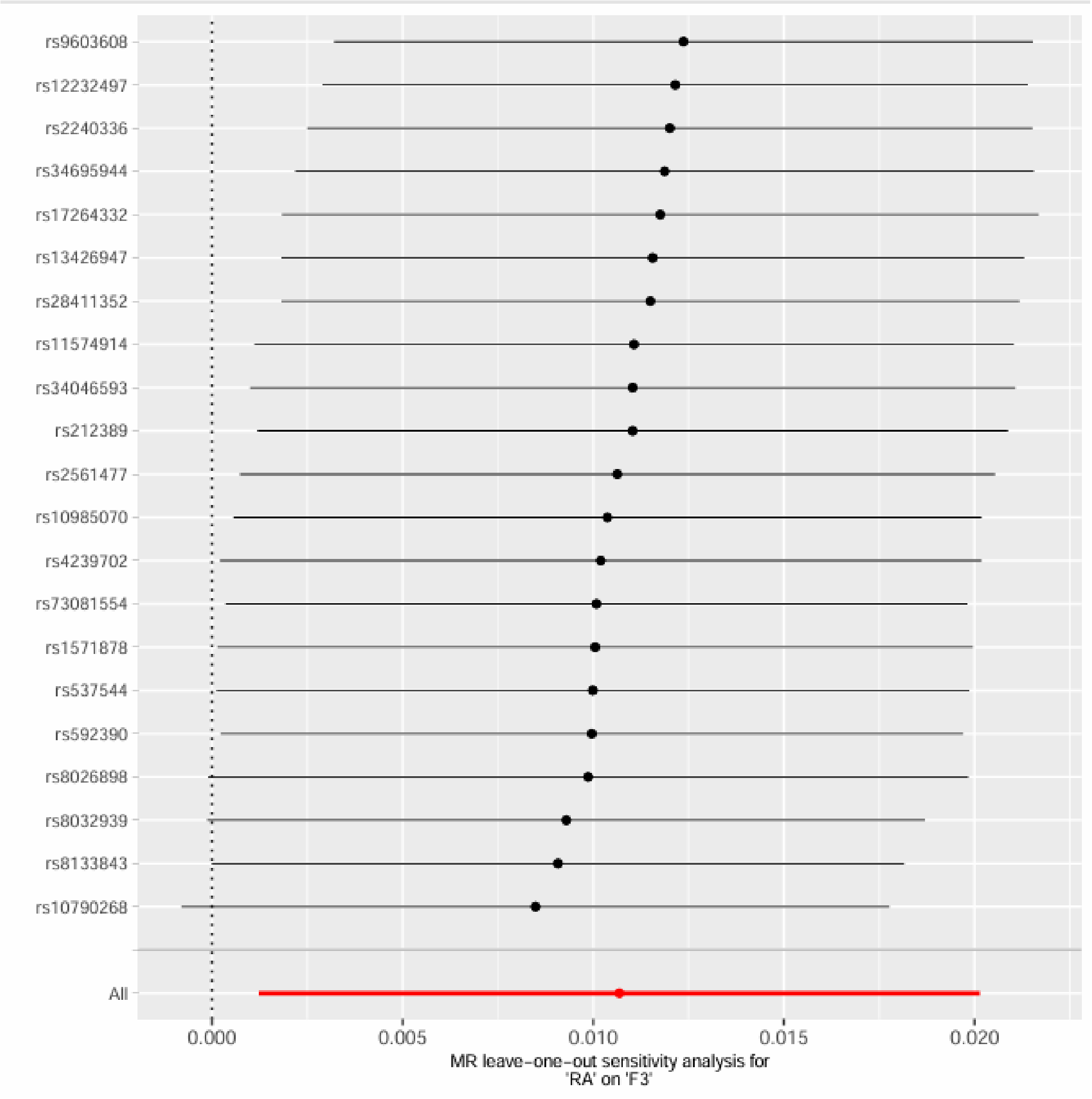

RA and F4 – Insufficient number of SNPs

RA and F5

Supplementary Figure 31. Leave-one-out sensitivity analysis for RA and F5.

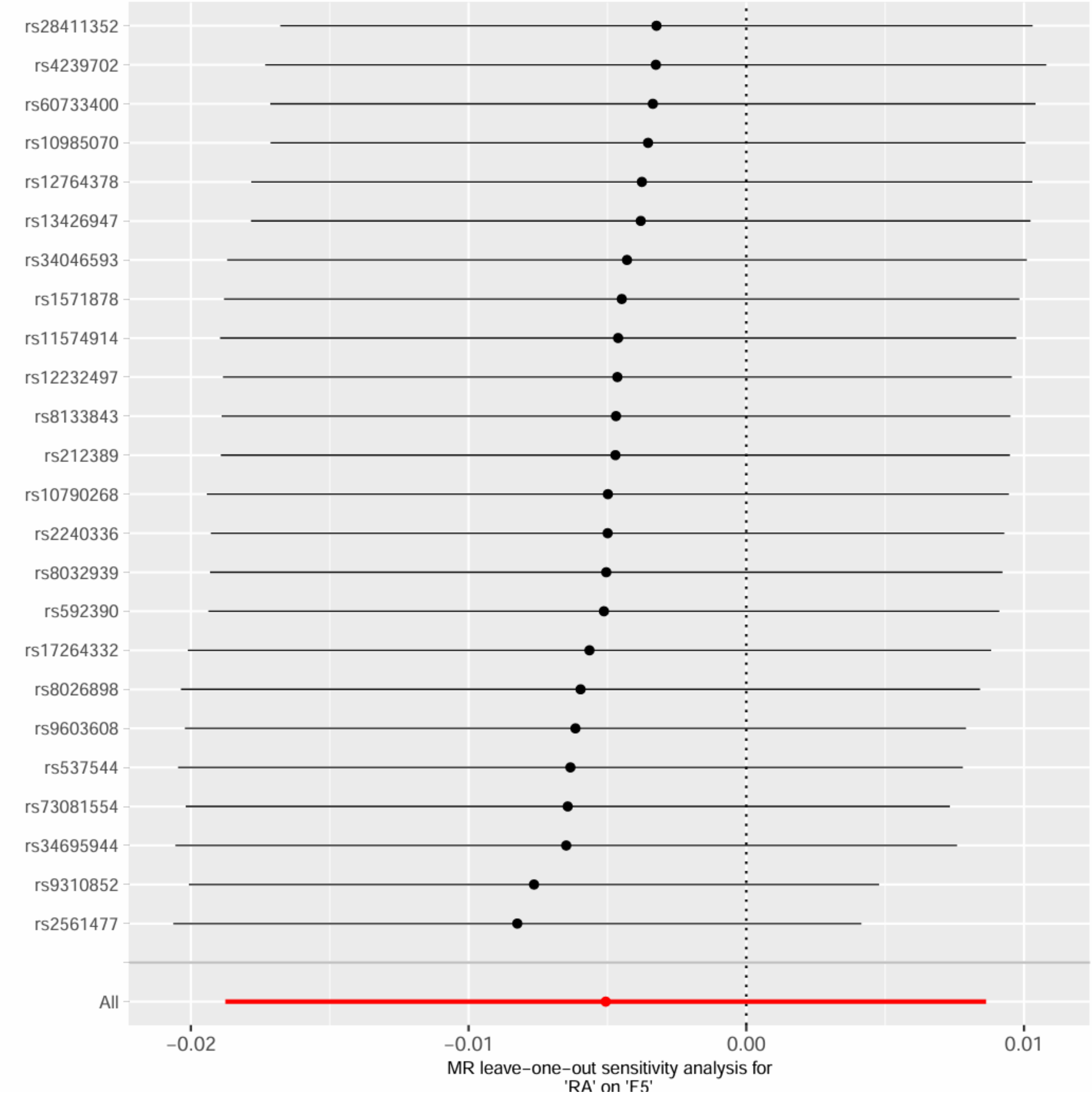


RA and F5, excluding MHC region

Supplementary Figure 32. Leave-one-out sensitivity analysis for RA and F5, excluding MHC region.

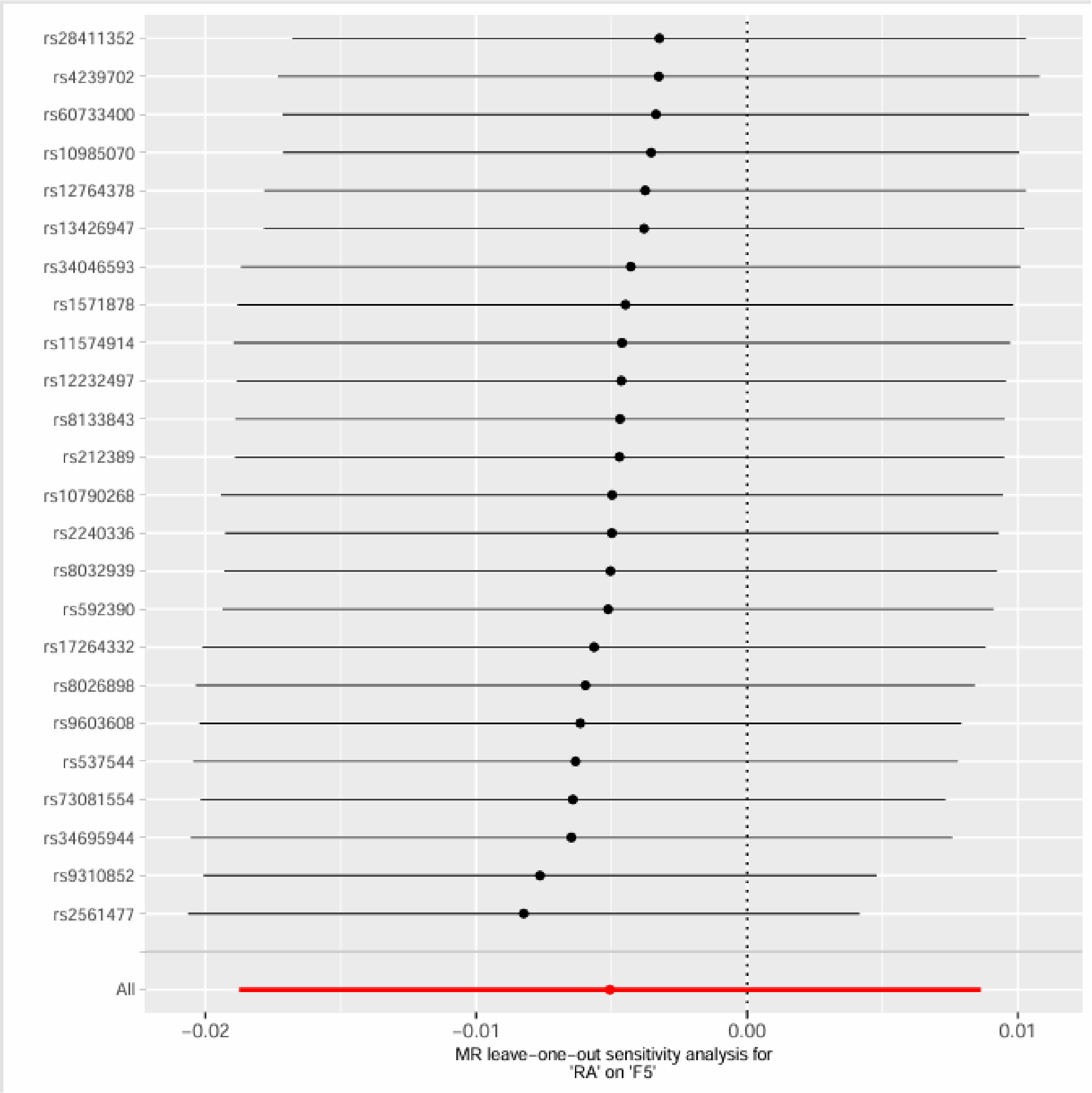
rs28411352
rs4239702
rs60733400
rs10985070
rs12764378
rs13426947
rs34046593
rs1571878
rs11574914
rs12232497
rs8133843
rs212389
rs10790268
rs2240336
rs8032939
rs592390
rs17264332
rs8026898
rs9603608
rs537544
rs73081554
rs34695944
rs9310852
rs2561477
All
−0.02
−0.01
0.00
0.01
MR leave−one−out sensitivity analysis for
'RA' on 'F5'

RA and F6

Supplementary Figure 33. Leave-one-out sensitivity analysis for RA and F6.

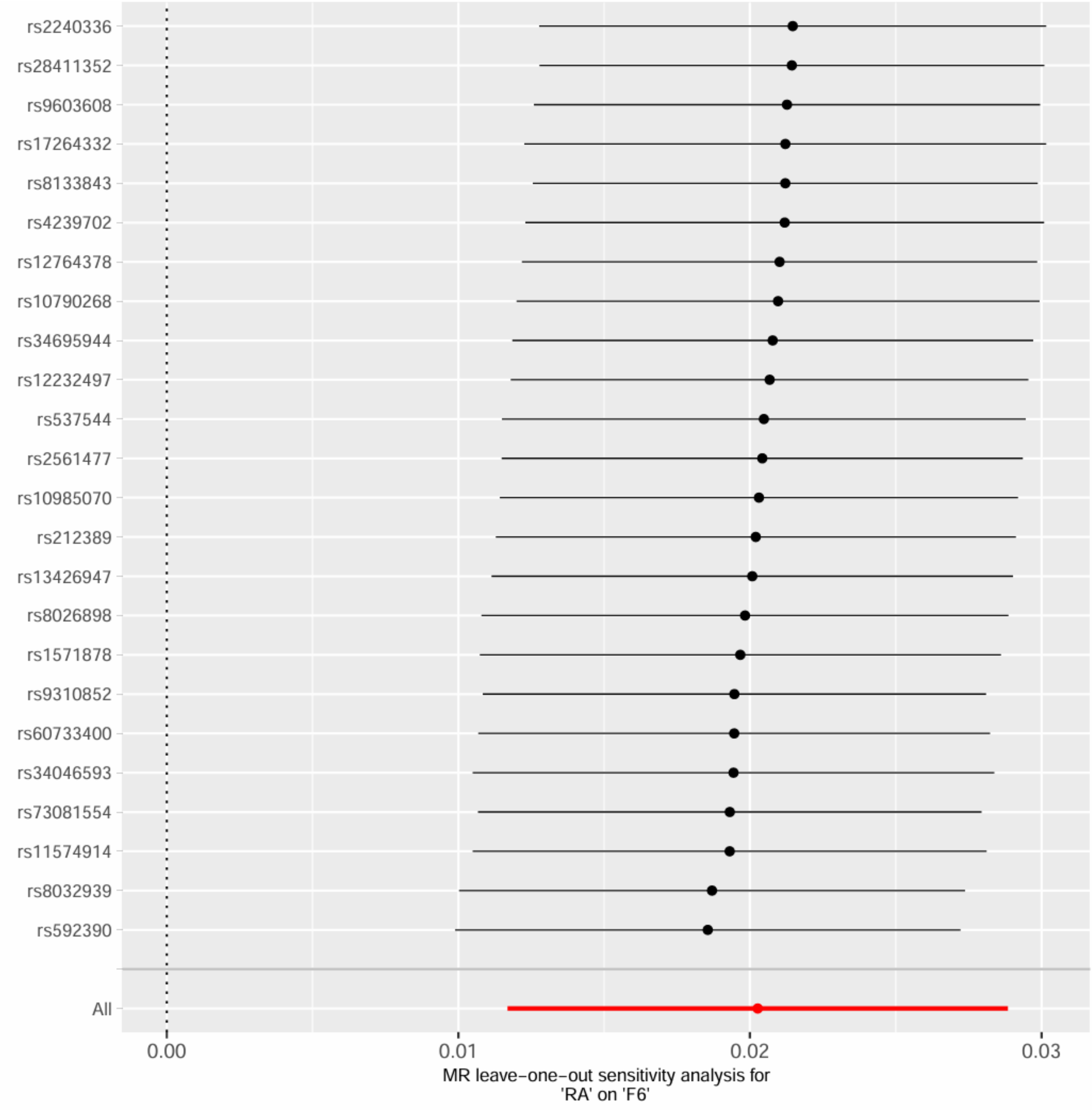

RA and F6, excluding MHC region

Supplementary Figure 34. Leave-one-out sensitivity analysis for RA and F6, excluding MHC region.

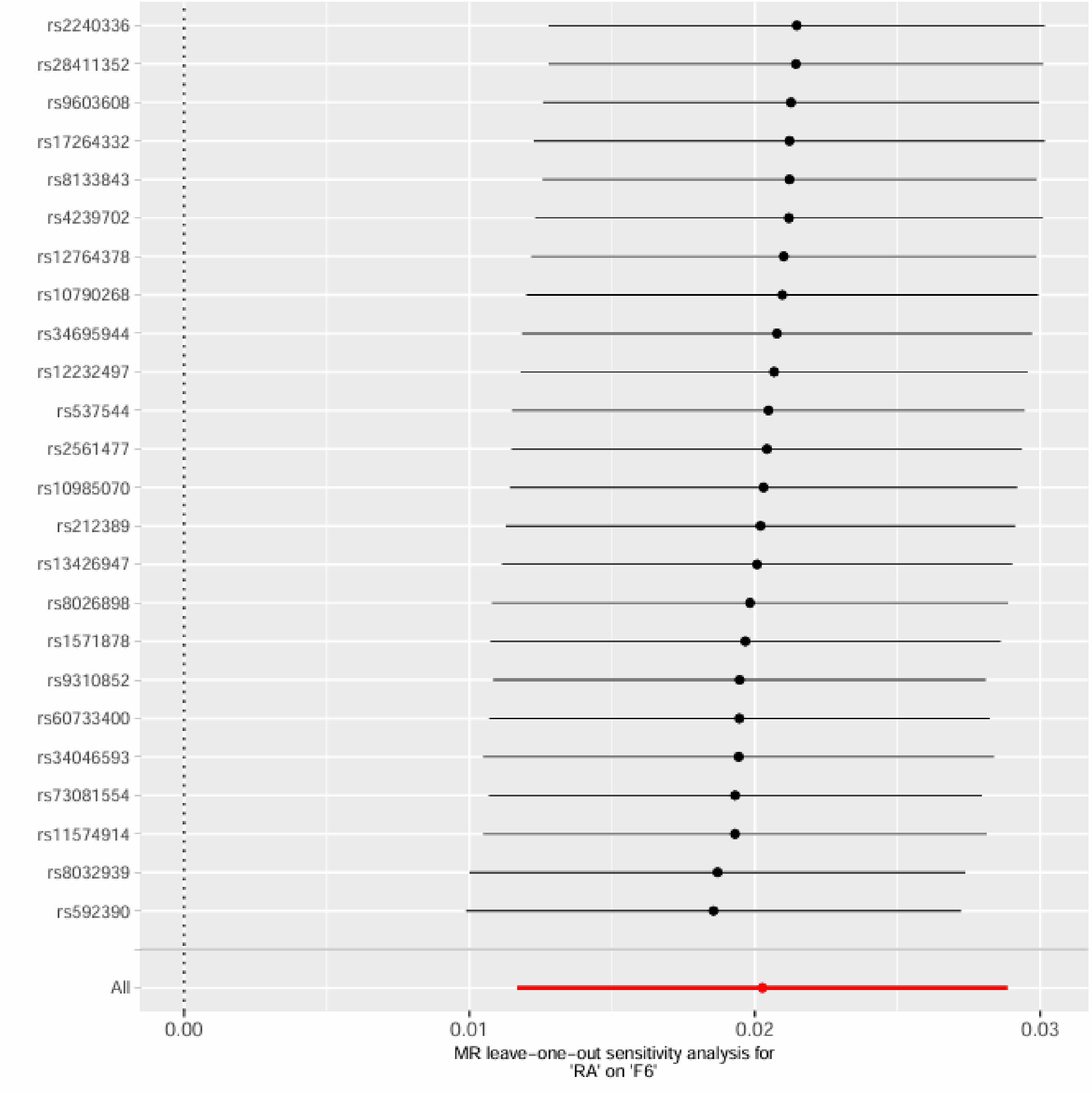

Frailty Exposures on Chronic Pain Outcome

FI on CP

Supplementary Figure 35. Leave-one-out sensitivity analysis for FI on CP.

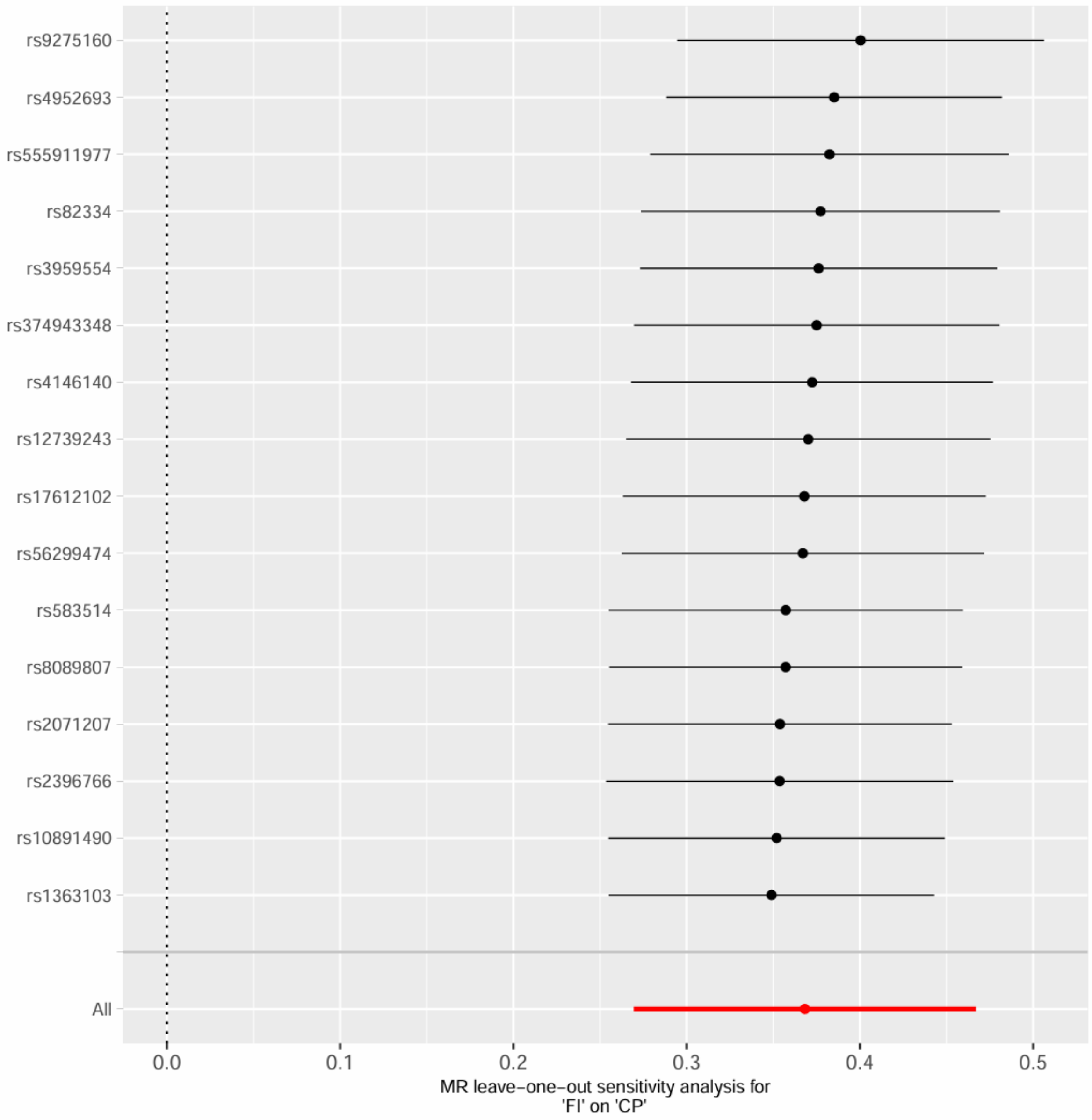

FI on CP, excluding MHC region

Supplementary Figure 36. Leave-one-out sensitivity analysis for FI on CP, excluding MHC region.

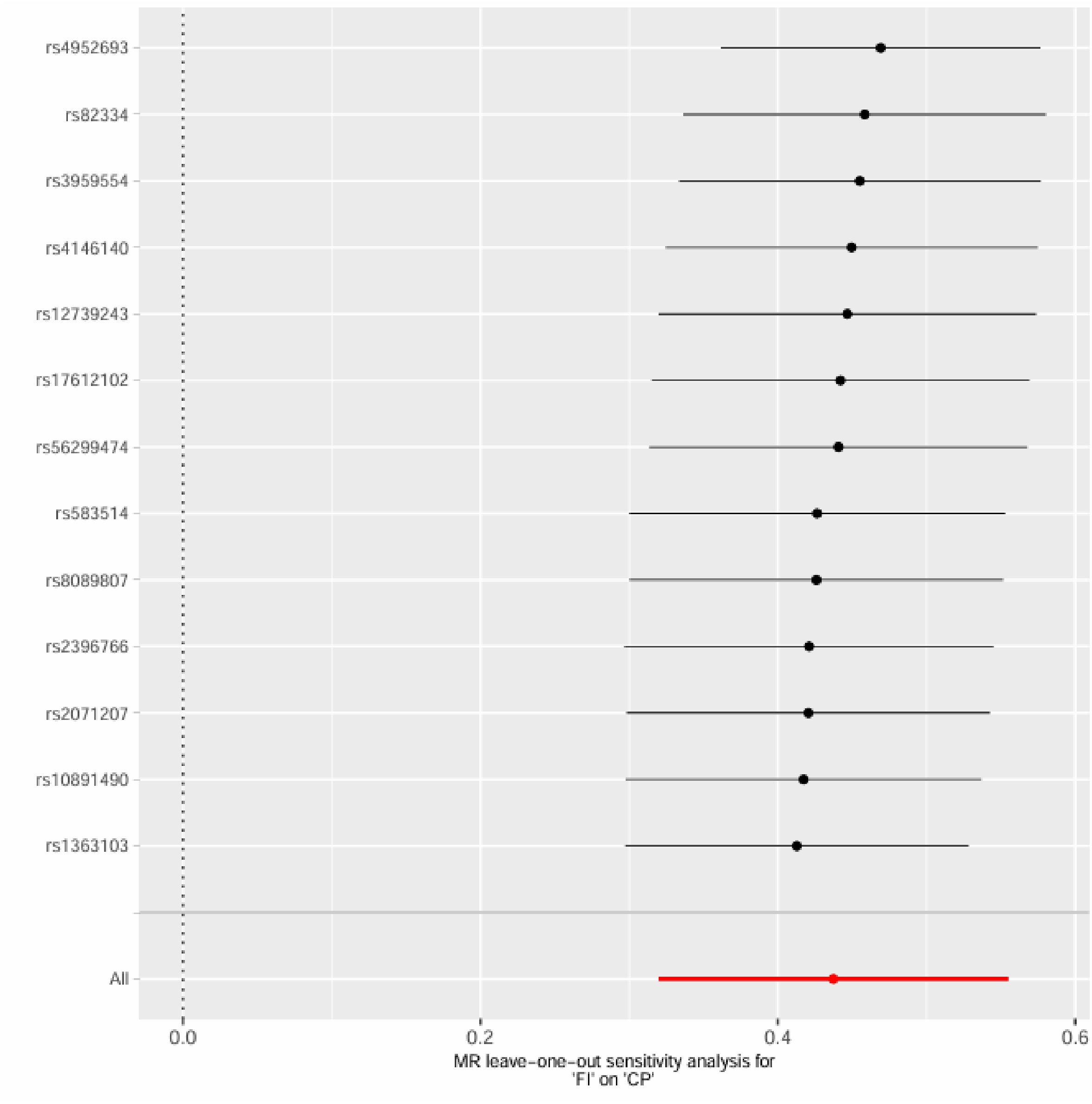

FFS on CP

Supplementary Figure 37. Leave-one-out sensitivity analysis for FFS on CP.

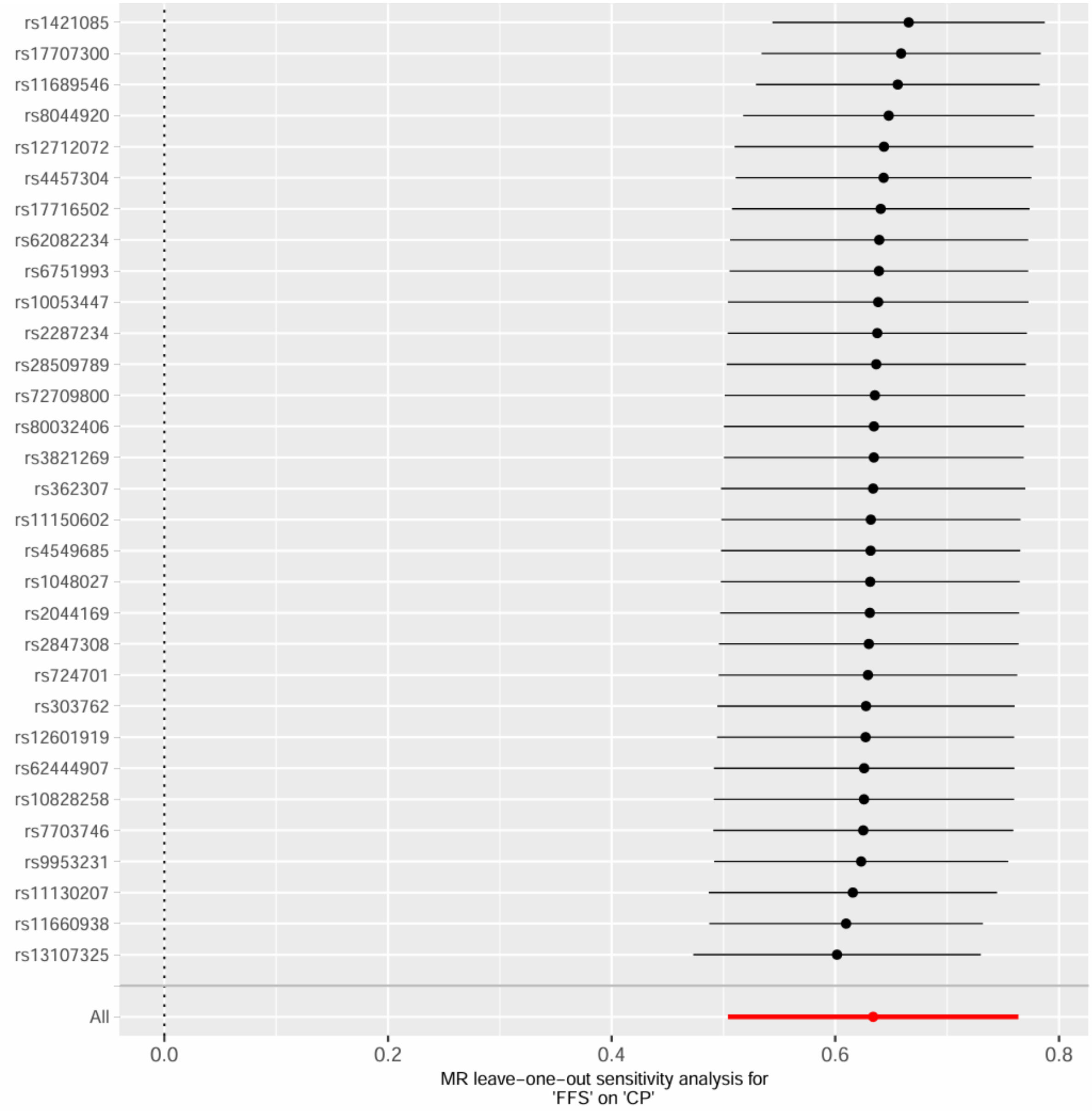

FFS on CP, excluding MHC region

Supplementary Figure 38. Leave-one-out sensitivity analysis for FFS on CP, excluding MHC region.

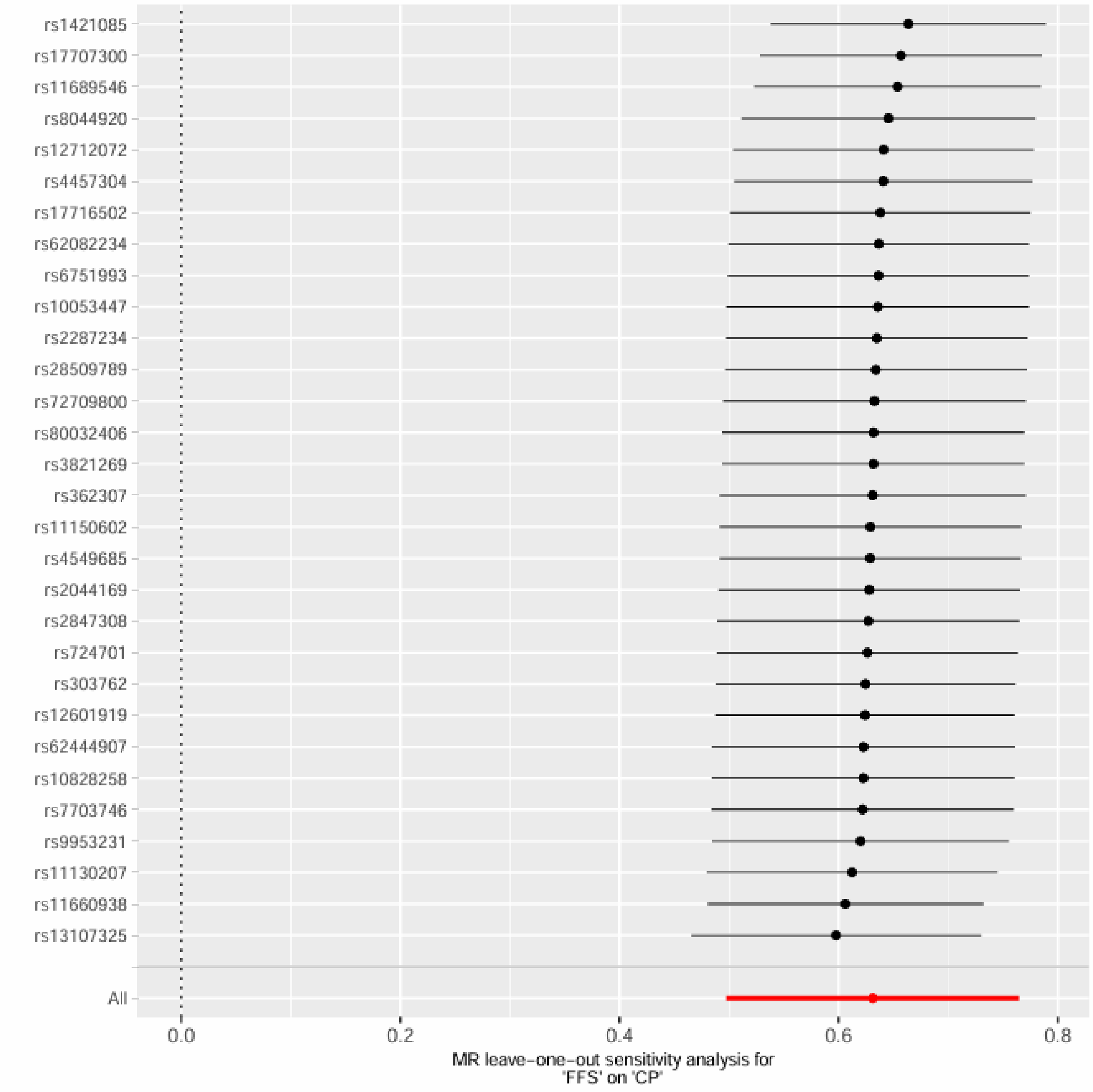

GF on CP

Supplementary Figure 39. Leave-one-out sensitivity analysis for GF on CP.

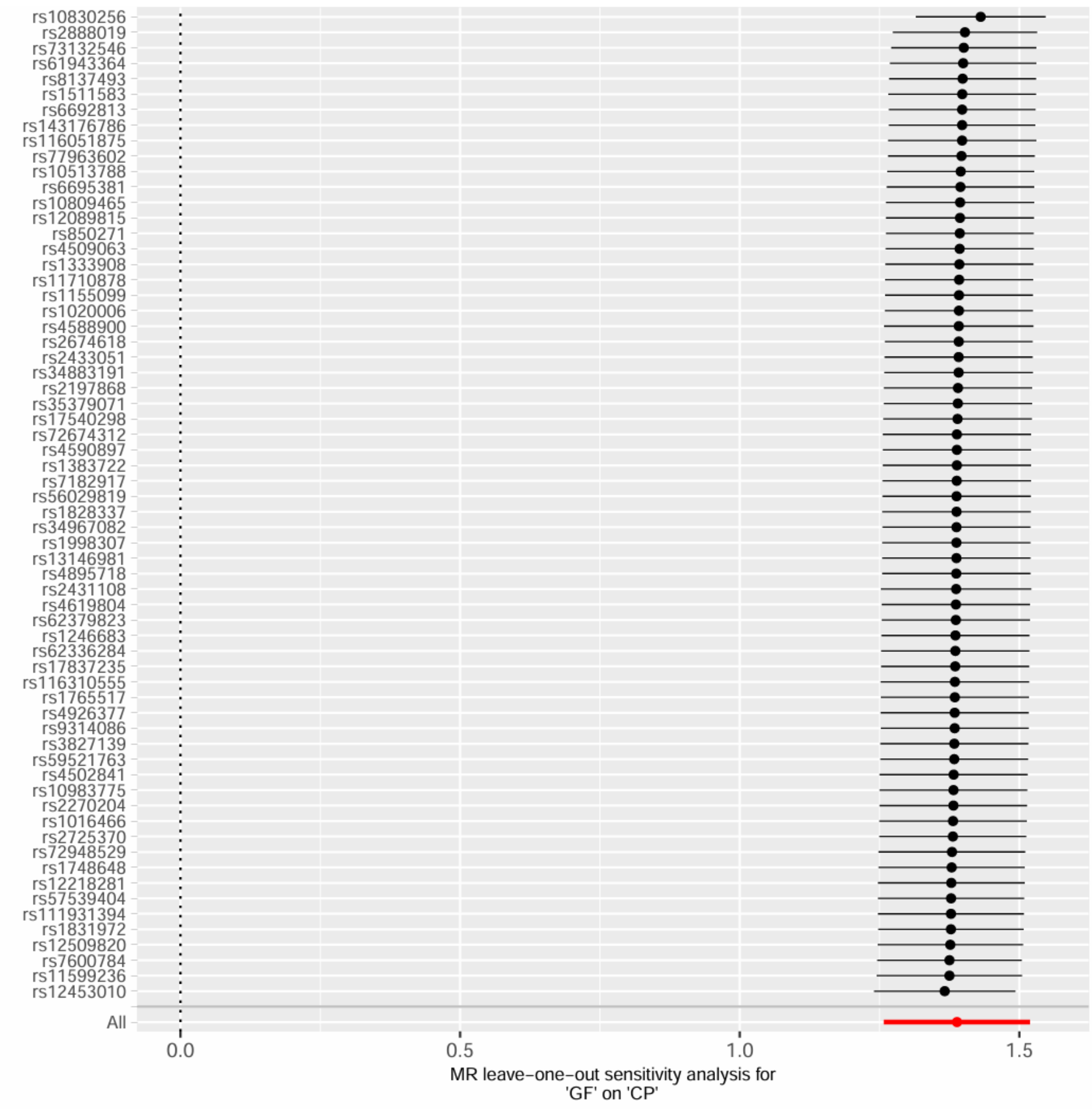


GF on CP, excluding MHC region

Supplementary Figure 40. Leave-one-out sensitivity analysis for GF on CP, excluding MHC region.

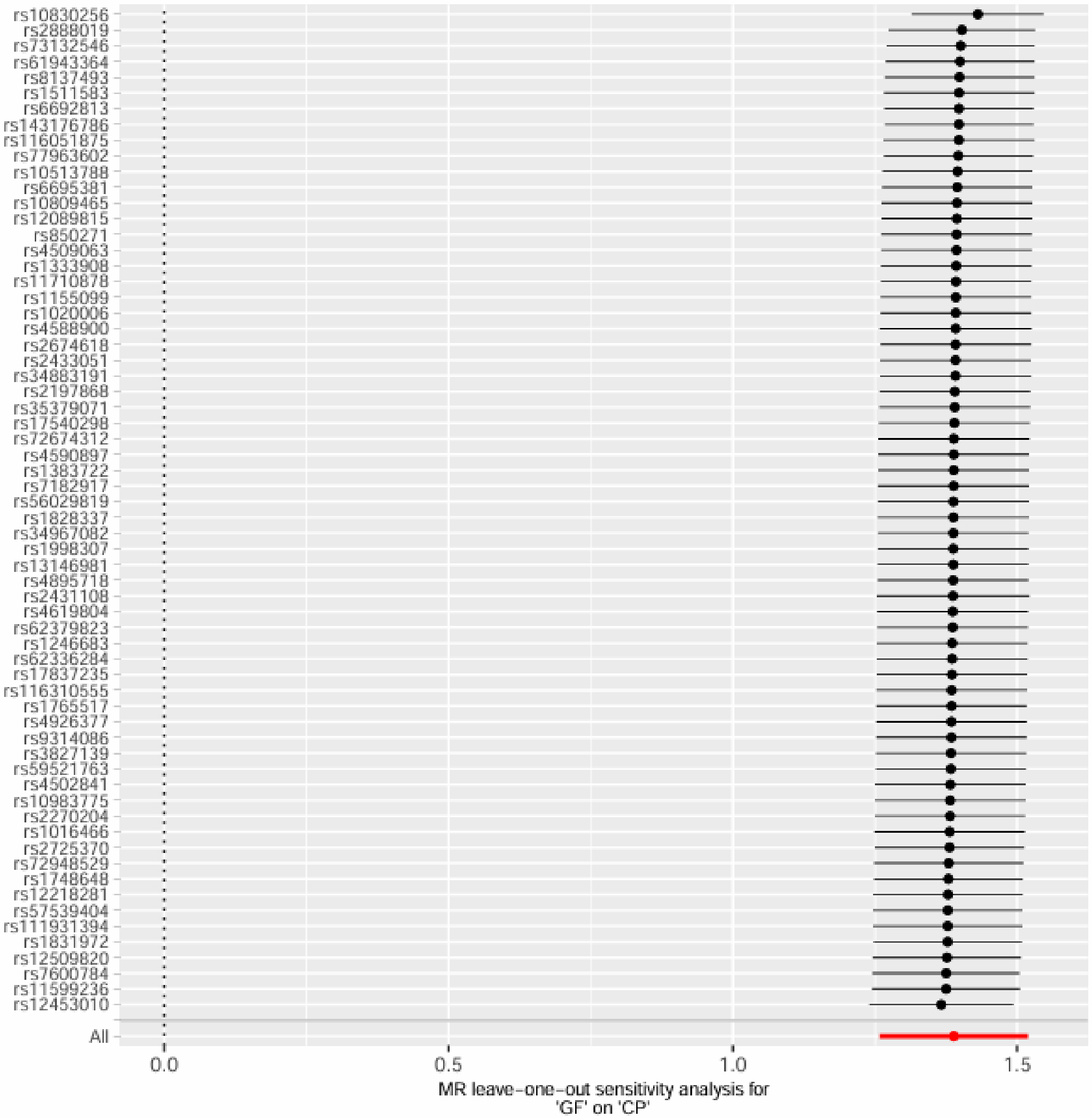
rs10830256
rs2888019
rs73132546
rs61943364
rs8137493
rs1511583
rs6692813
rs143176786
rs116051875
rs77963602
rs10513788
rs6695381
rs10809465
rs12089815
rs850271
rs4509063
rs1333908
rs11710878
rs1155099
rs1020006
rs4588900
rs2674618
rs2433051
rs34883191
rs2197868
rs35379071
rs17540298
rs72674312
rs4590897
rs1383722
rs7182917
rs56029819
rs1828337
rs34967082
rs1998307
rs13146981
rs4895718
rs2431108
rs4619804
rs62379823
rs1246683
rs62336284
rs17837235
rs116310555
rs1765517
rs4926377
rs9314086
rs3827139
rs59521763
rs4502841
rs10983775
rs2270204
rs1016466
rs2725370
rs72948529
rs1748648
rs12218281
rs57539404
rs111931394
rs1831972
rs12509820
rs7600784
rs11599236
rs12453010
All
0.0
0.5
1.0
1.5
MR leave−one−out sensitivity analysis for
'GF' on 'CP'

F1 on CP

Supplementary Figure 41. Leave-one-out sensitivity analysis for F1 on CP.

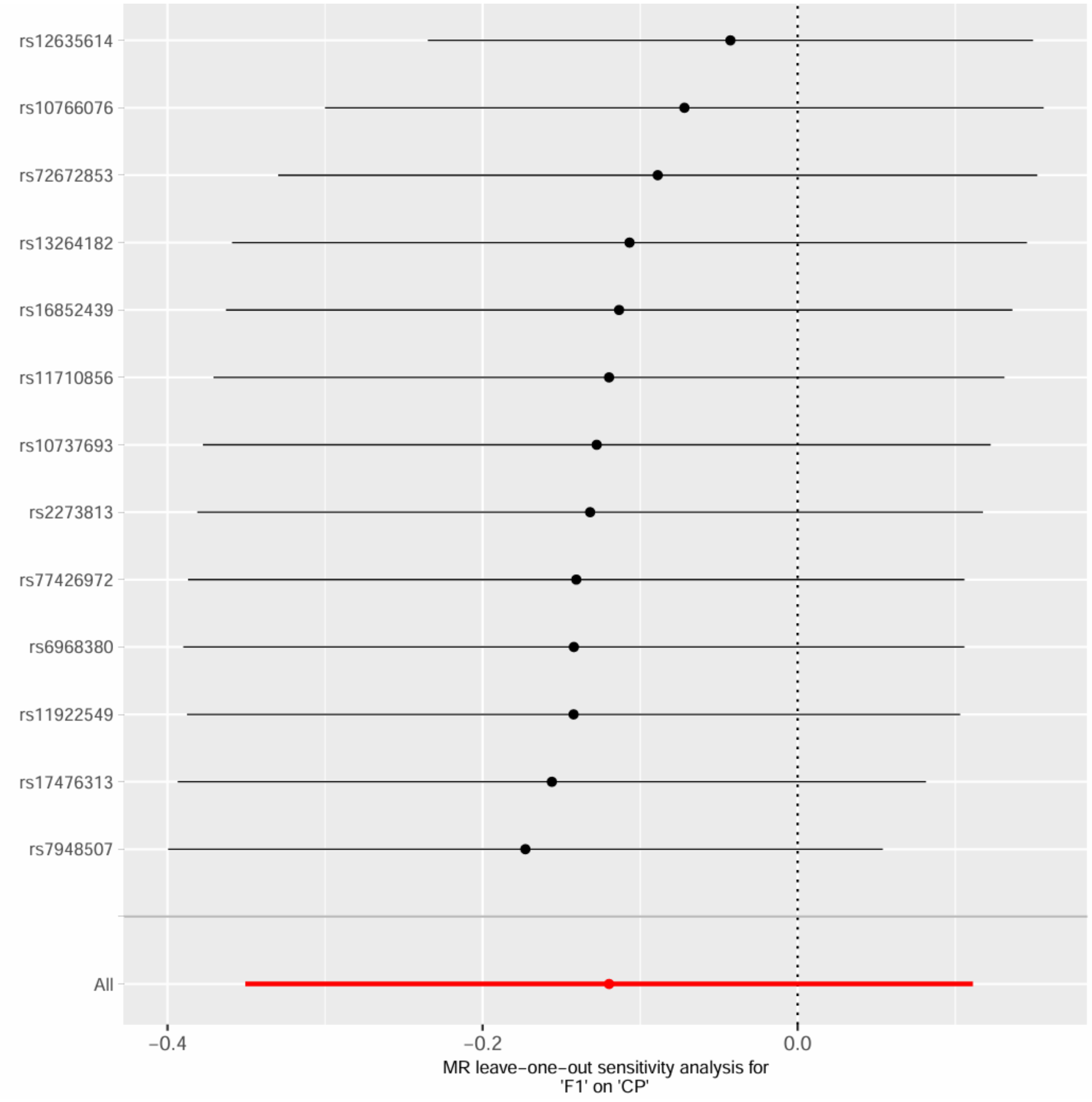

FI on CP, excluding MHC region

Supplementary Figure 42. Leave-one-out sensitivity analysis for FI on CP, excluding MHC region.

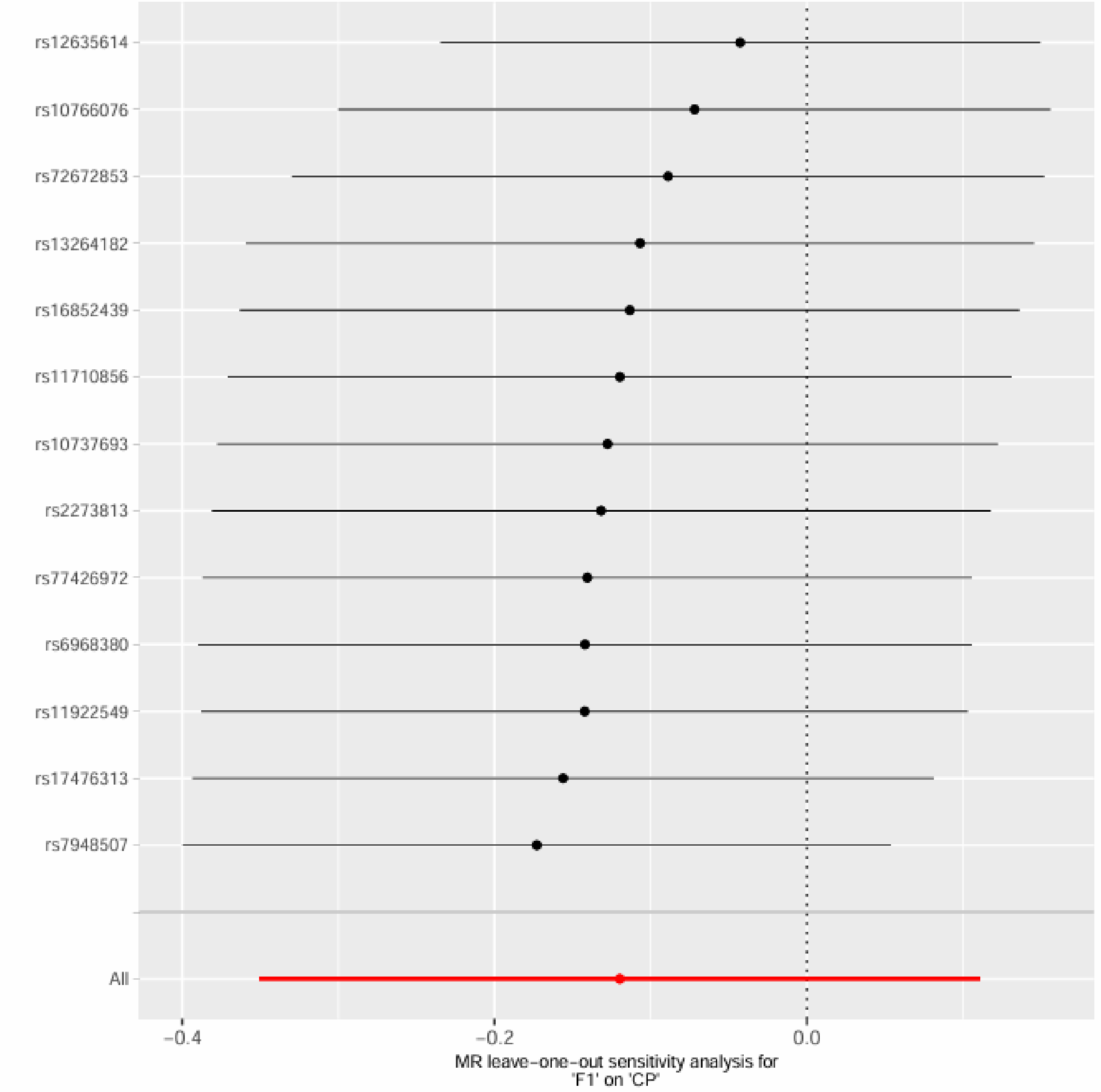

F2 on CP

Supplementary Figure 43. Leave-one-out sensitivity analysis for F2 on CP.

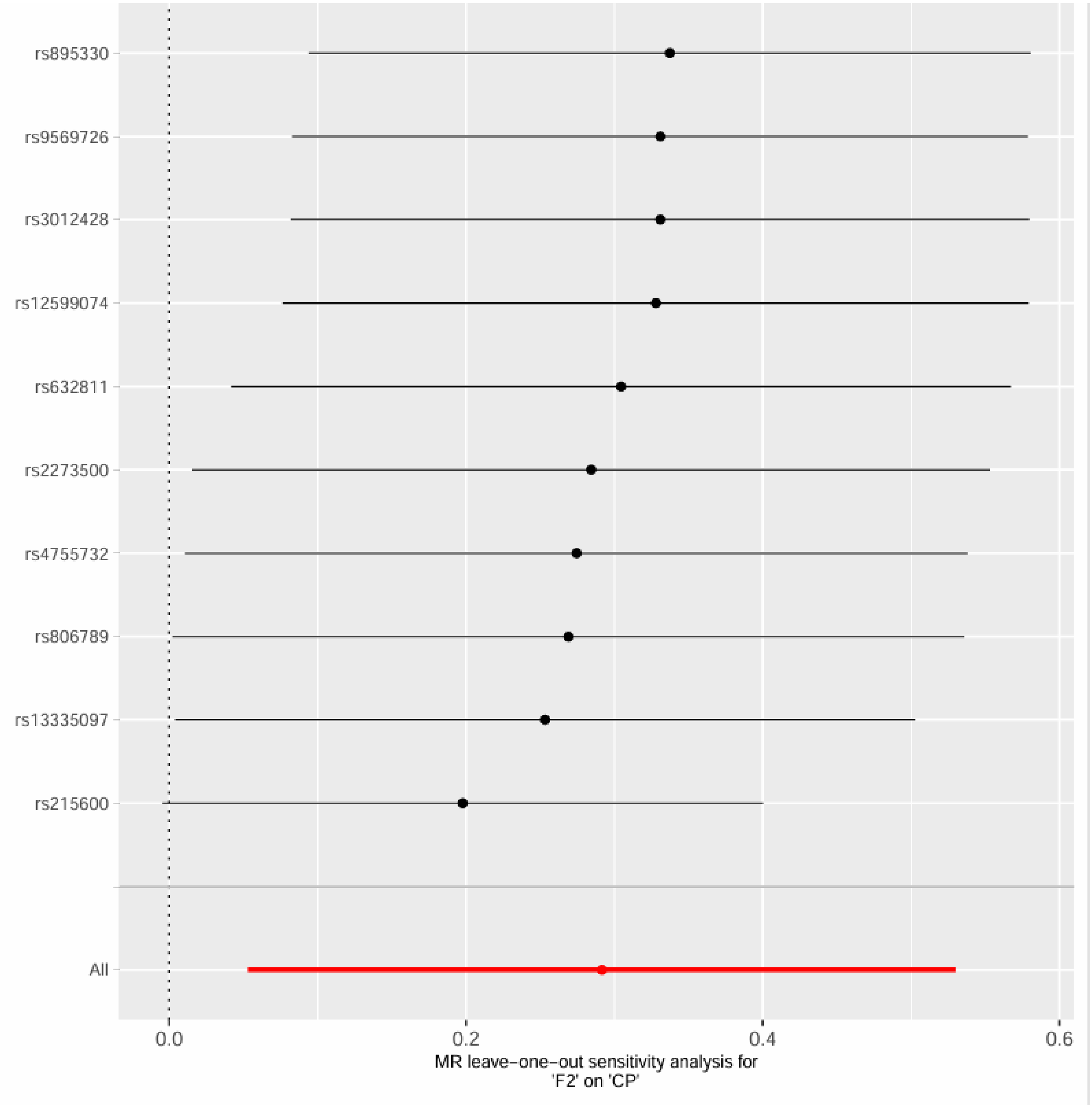

F2 on CP, excluding MHC region

Supplementary Figure 44. Leave-one-out sensitivity analysis for F2 on CP, excluding MHC region.

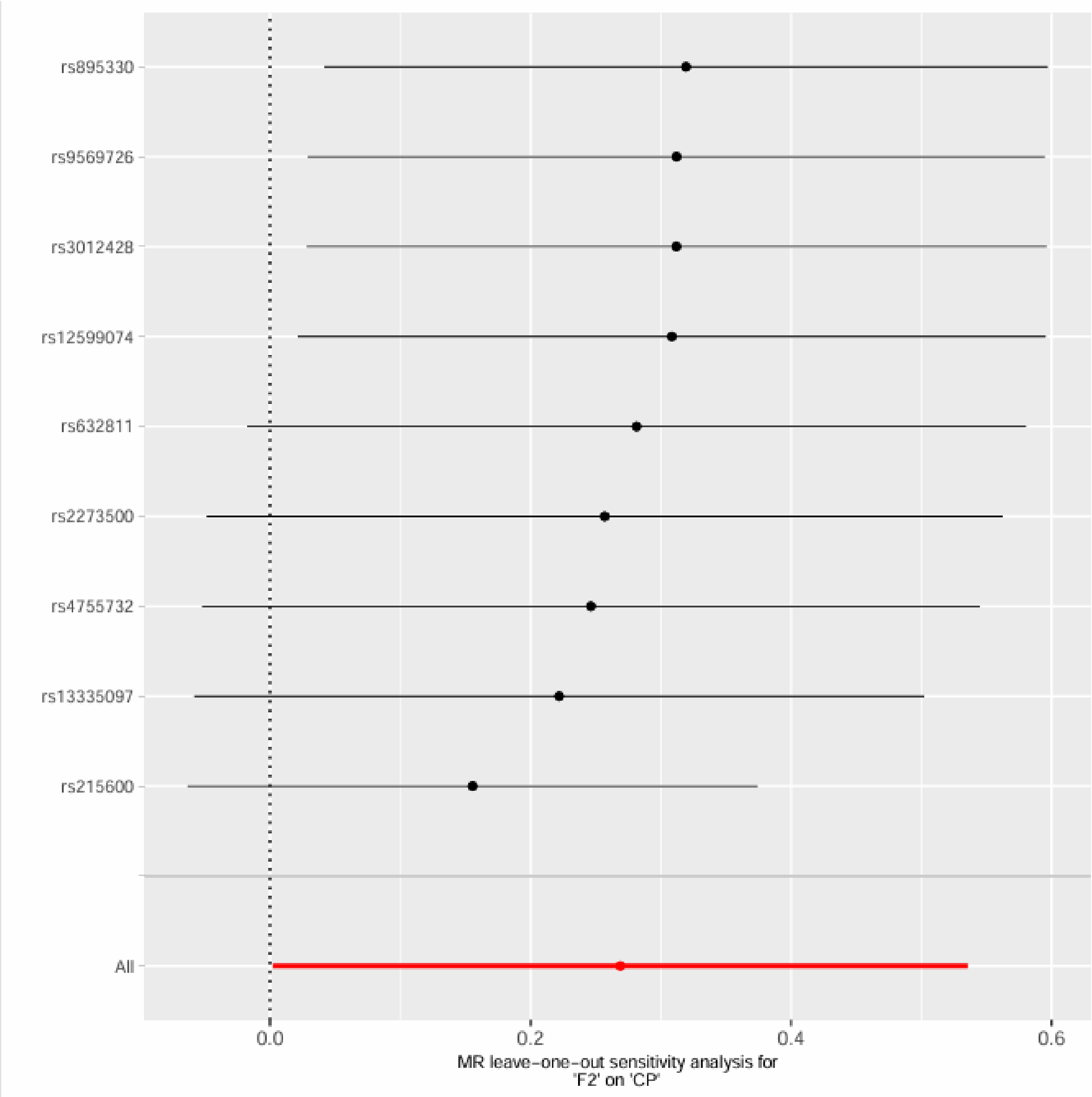

F3 on CP

Supplementary Figure 45. Leave-one-out sensitivity analysis for F3 on CP.

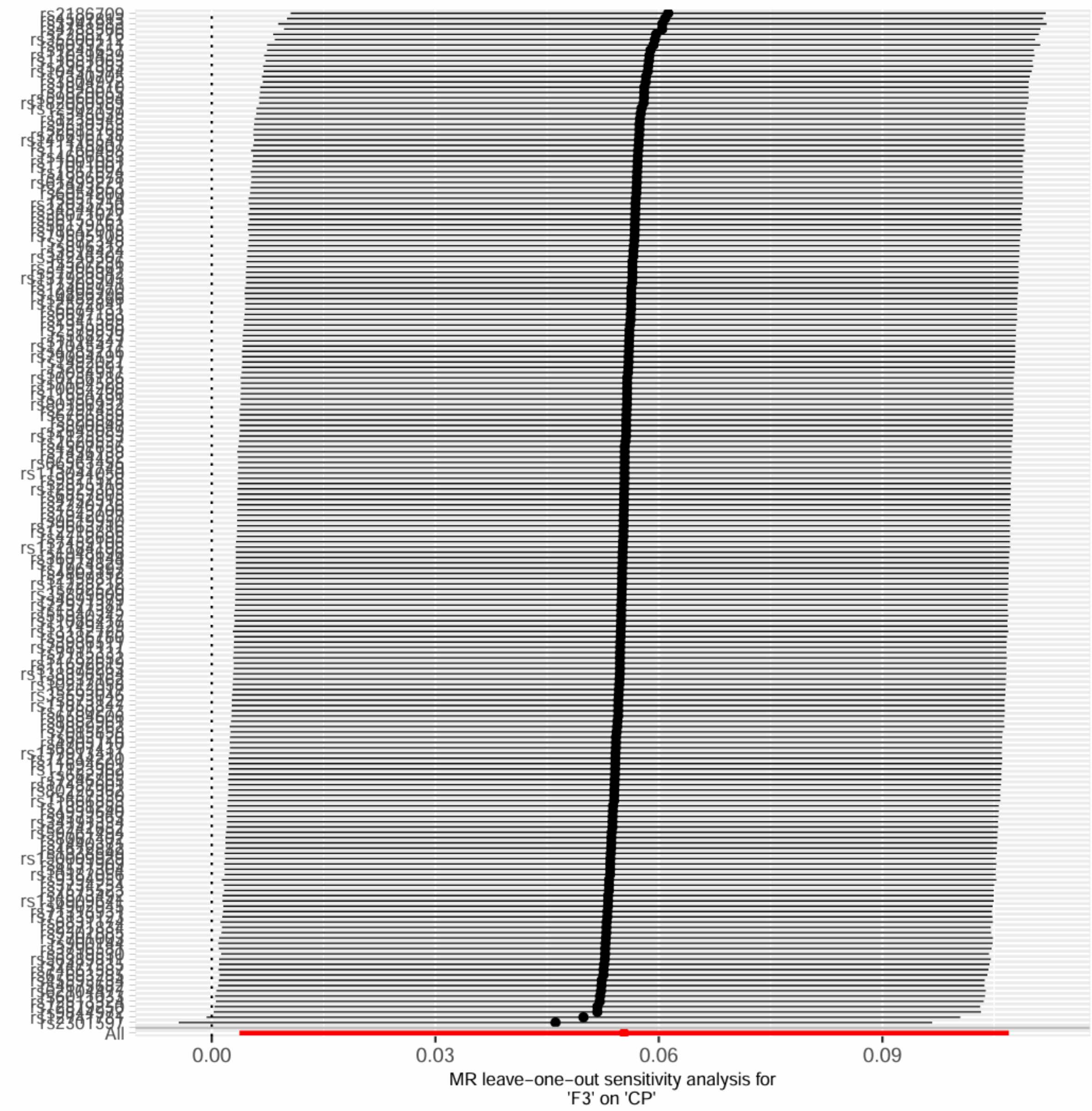

F3 on CP, excluding MHC region

Supplementary Figure 46. Leave-one-out sensitivity analysis for F3 on CP, excluding MHC region.

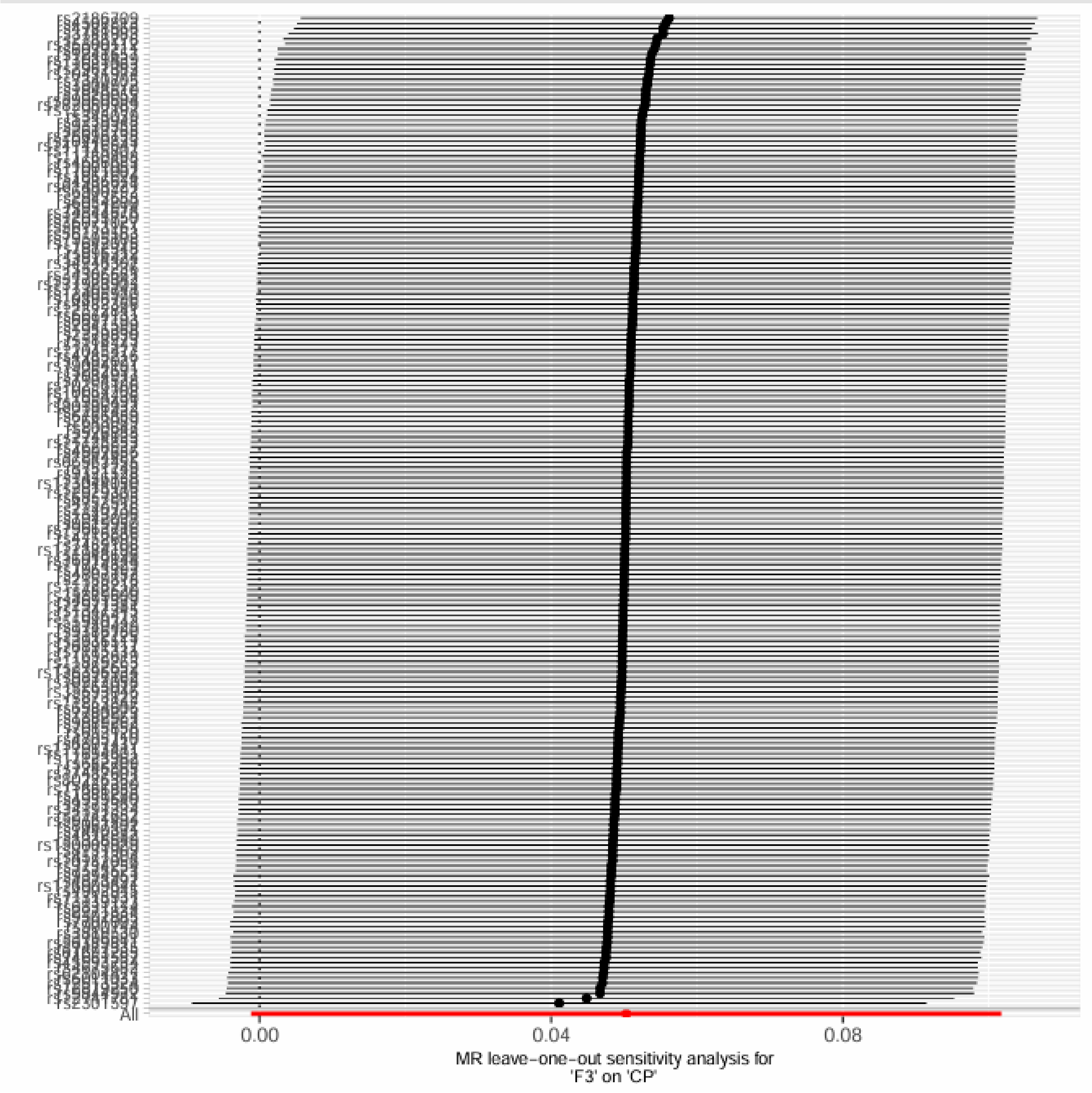

F4 on CP

Supplementary Figure 47. Leave-one-out sensitivity analysis for F4 on CP.

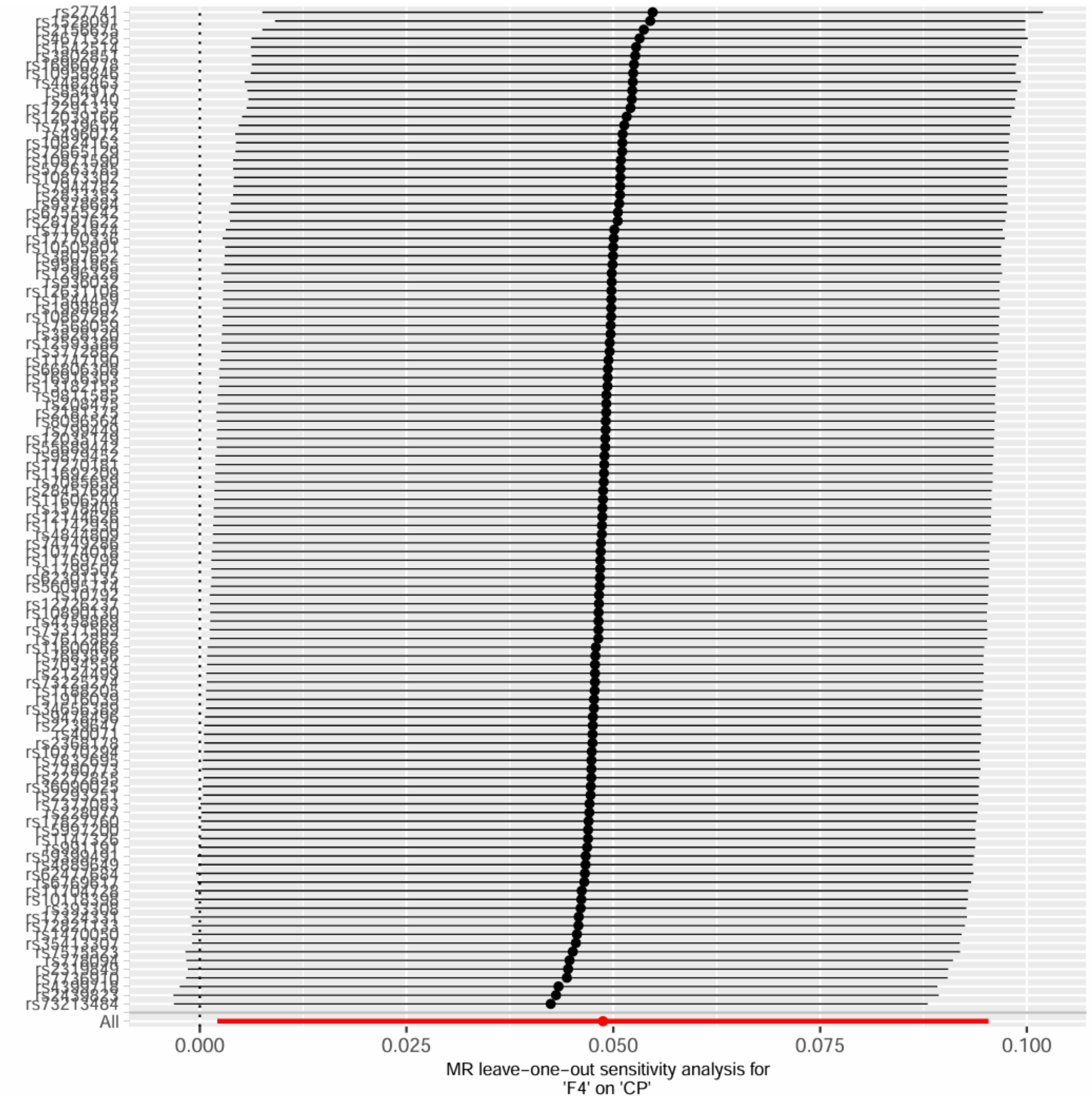

F4 on CP, excluding MHC region

Supplementary Figure 48. Leave-one-out sensitivity analysis for F4 on CP, excluding MHC region.

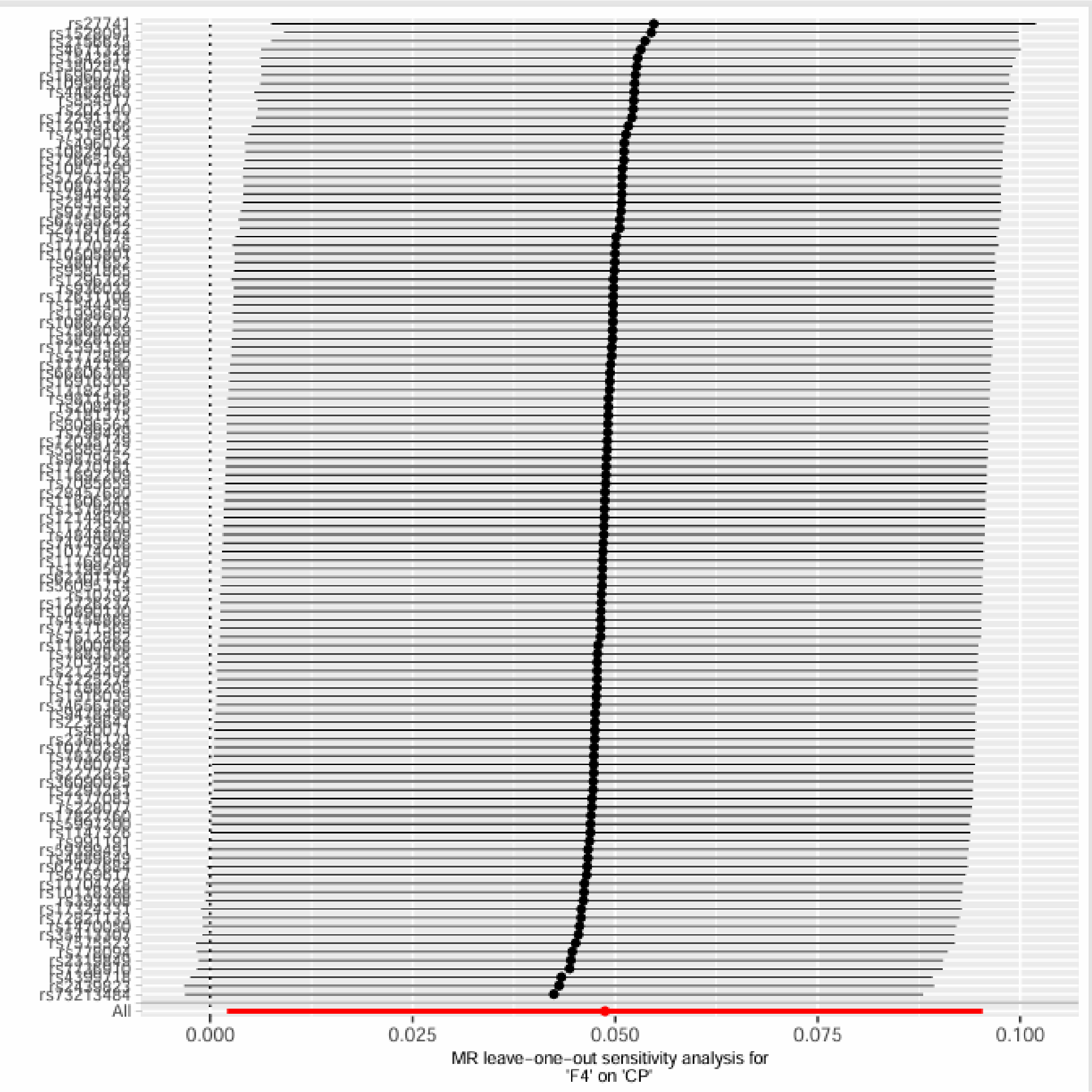

F5 on CP

Supplementary Figure 49. Leave-one-out sensitivity analysis for F5 on CP.

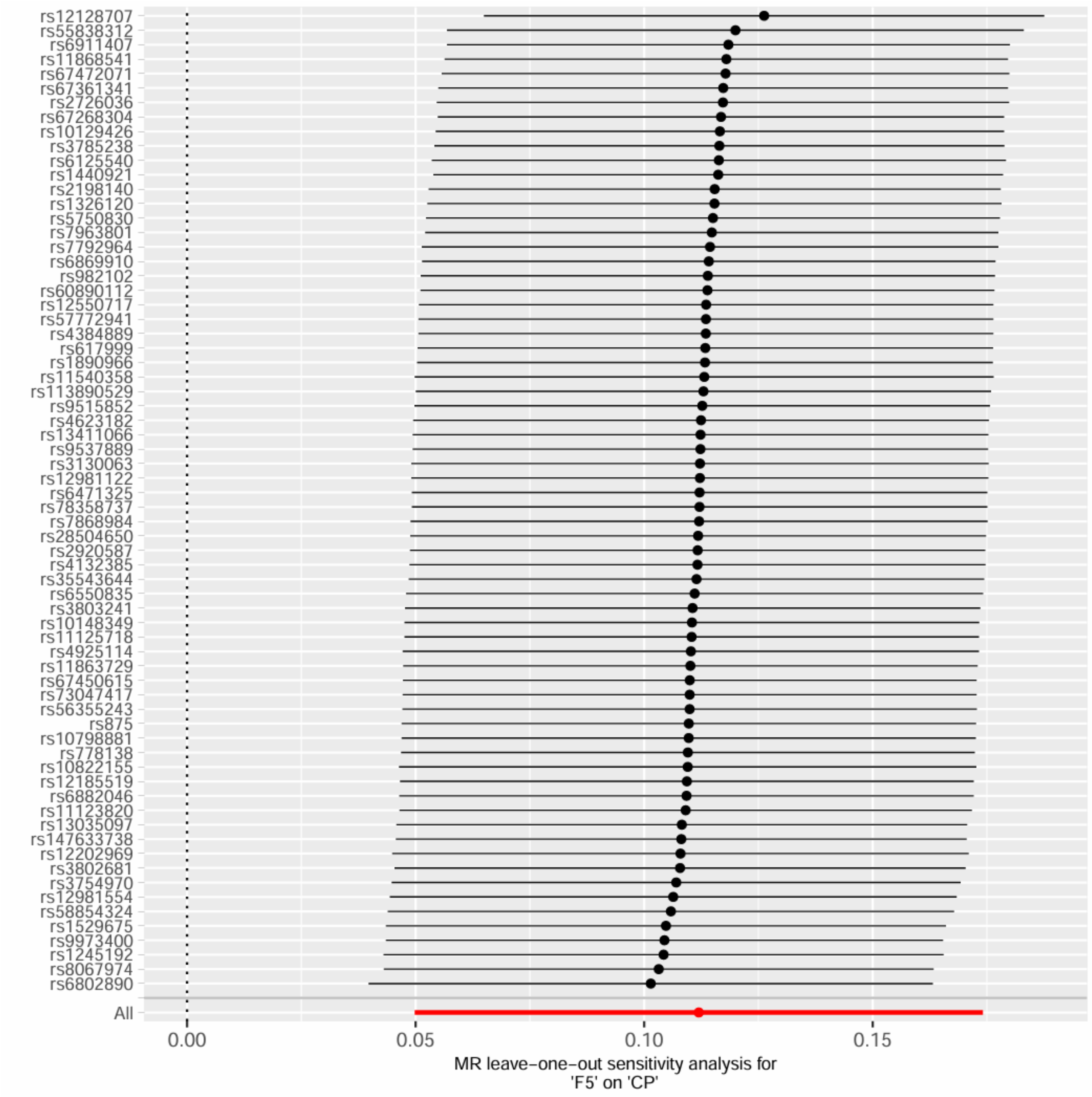


F5 on CP, excluding MHC region

Supplementary Figure 50. Leave-one-out sensitivity analysis for F5 on CP, excluding MHC region.

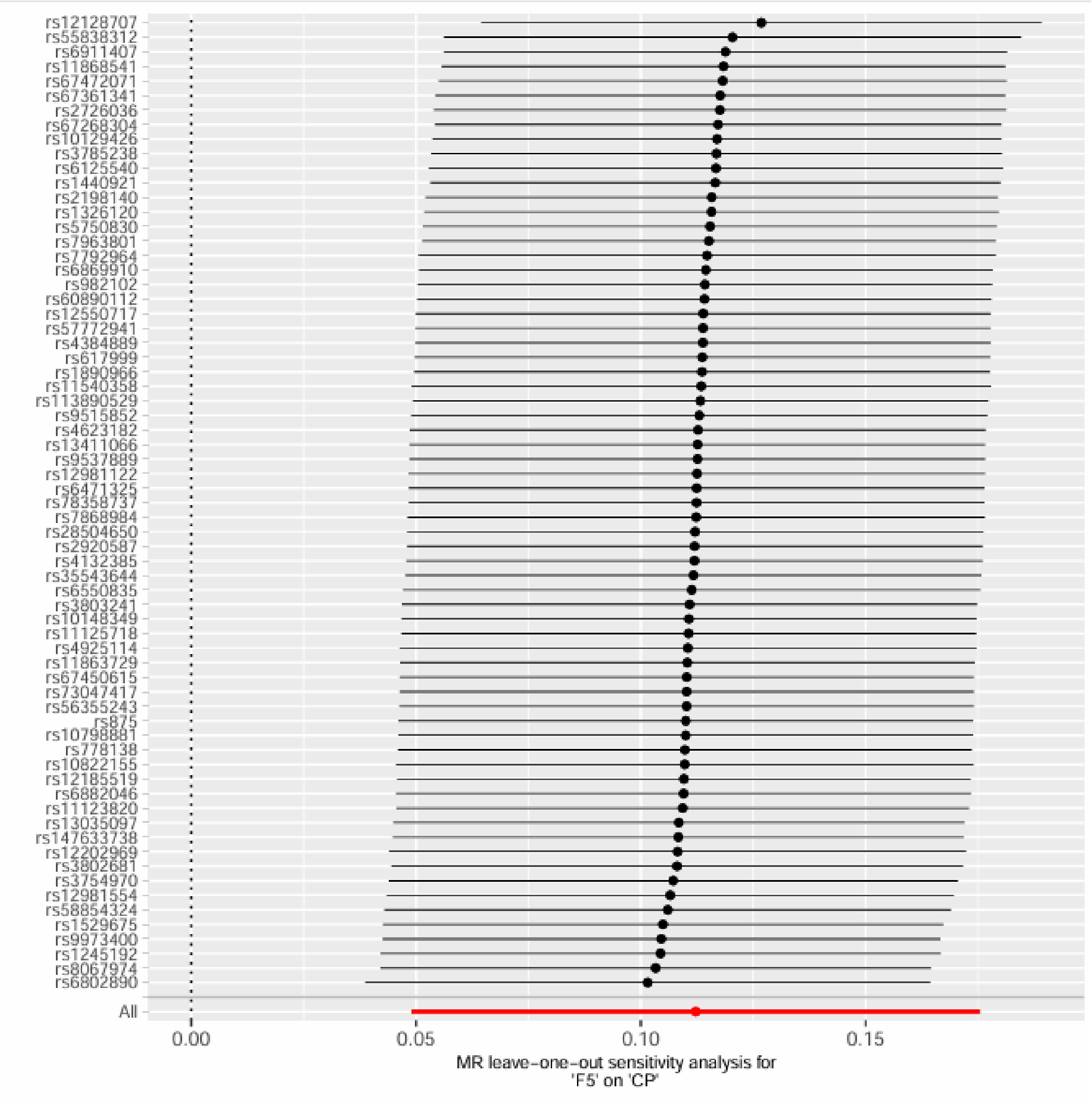
rs12128707
rs55838312
rs6911407
rs11868541
rs67472071
rs67361341
rs2726036
rs67268304
rs10129426
rs3785238
rs6125540
rs1440921
rs2198140
rs1326120
rs5750830
rs7963801
rs7792964
rs6869910
rs982102
rs60890112
rs12550717
rs57772941
rs4384889
rs617999
rs1890966
rs11540358
rs113890529
rs9515852
rs4623182
rs13411066
rs9537889
rs12981122
rs6471325
rs78358737
rs7868984
rs28504650
rs2920587
rs4132385
rs35543644
rs6550835
rs3803241
rs10148349
rs11125718
rs4925114
rs11863729
rs67450615
rs73047417
rs56355243
rs875
rs10798881
rs778138
rs10822155
rs12185519
rs6882046
rs11123820
rs13035097
rs147633738
rs12202969
rs3802681
rs3754970
rs12981554
rs58854324
rs1529675
rs9973400
rs1245192
rs8067974
rs6802890
All
0.00
0.05
0.10
0.15
MR leave-one-out sensitivity analysis for
'F5' on 'CP'

F6 on CP

Supplementary Figure 51. Leave-one-out sensitivity analysis for F6 on CP.

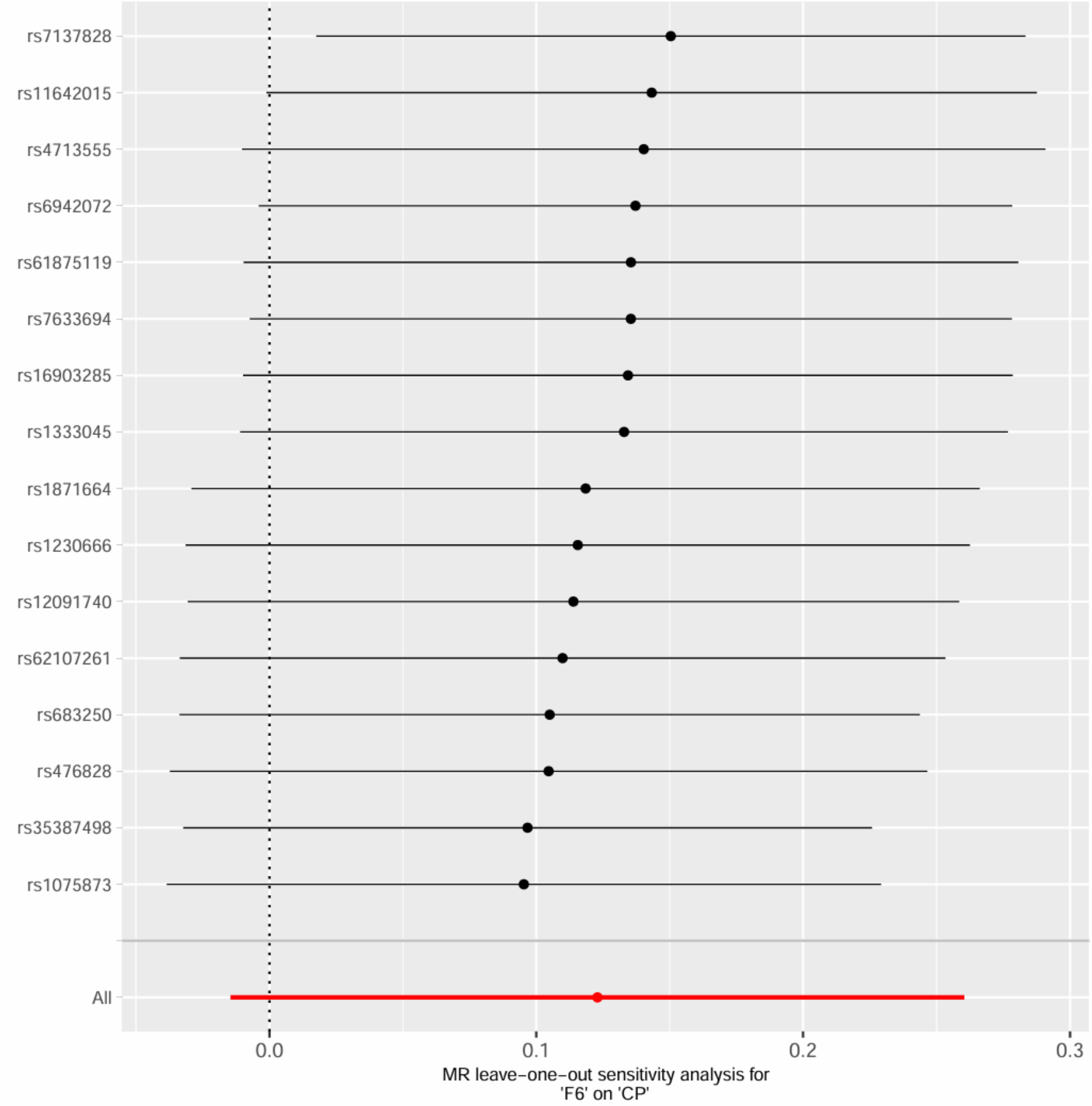

F6 on CP, excluding MHC region

Supplementary Figure 52. Leave-one-out sensitivity analysis for F6 on CP, excluding MHC region.

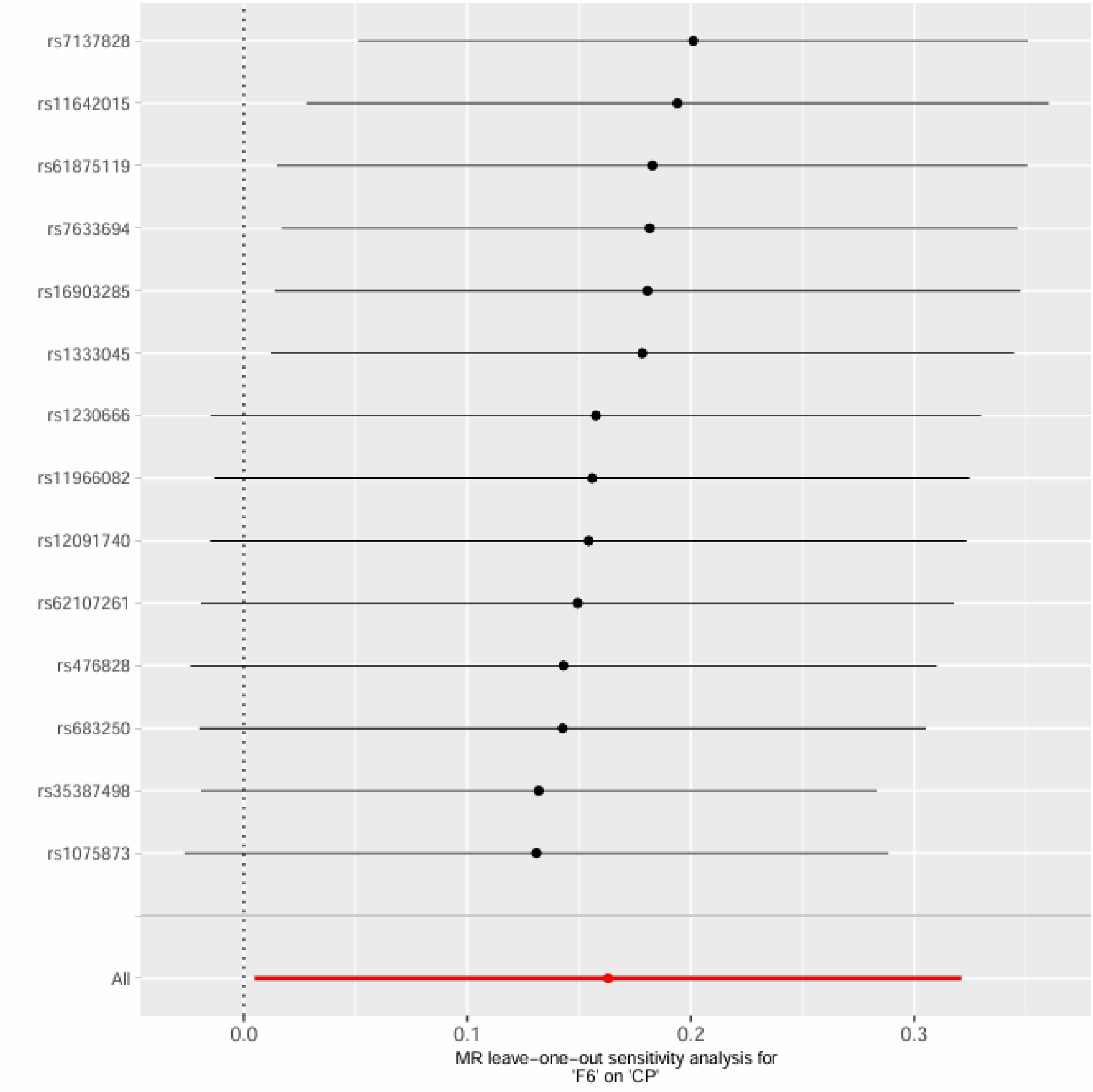

Frailty Exposures on Rheumatoid Arthritis Outcome

FI on RA

Supplementary Figure 53. Leave-one-out sensitivity analysis for FI on RA.

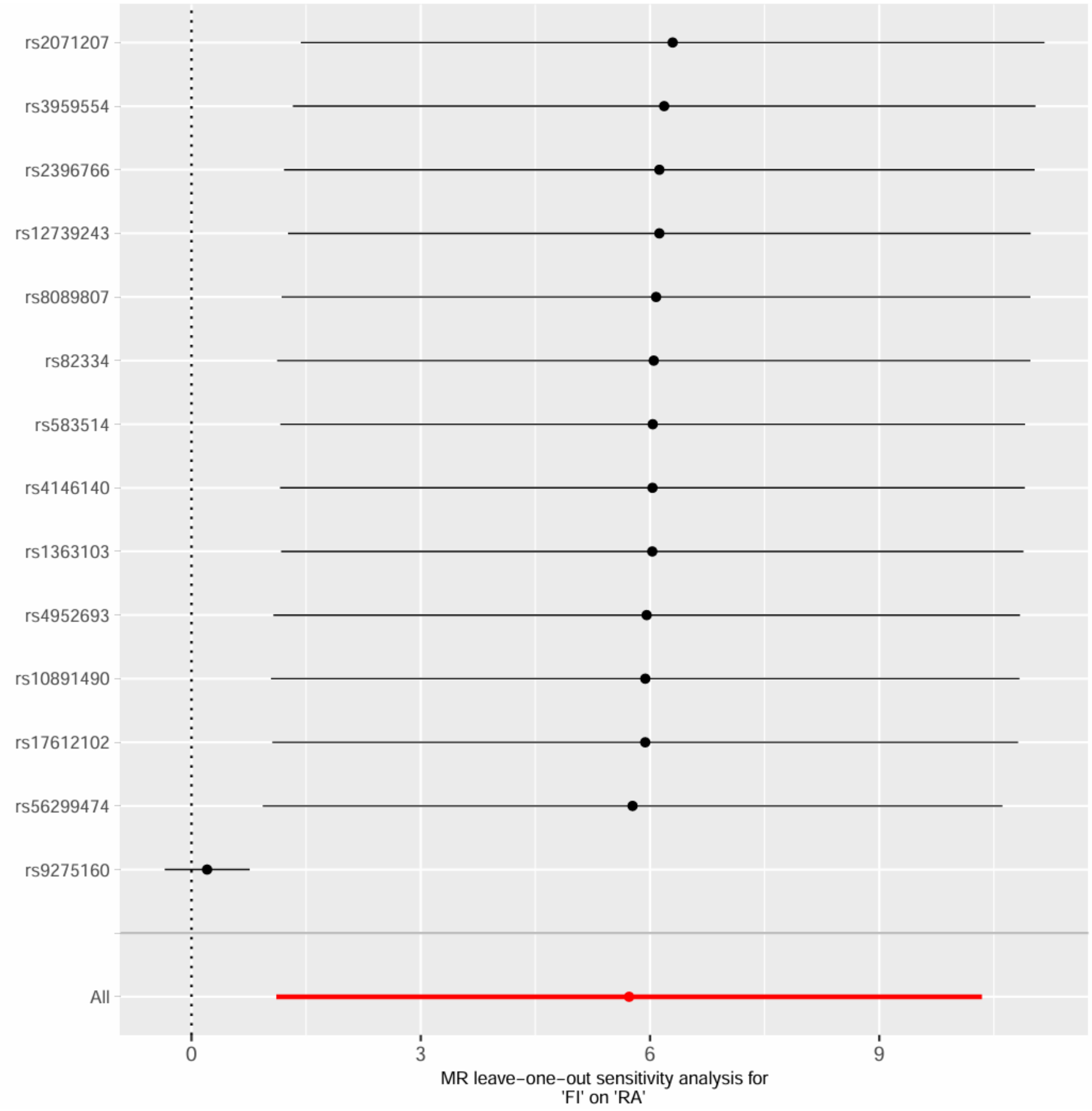

FI on RA, excluding MHC region

Supplementary Figure 54. Leave-one-out sensitivity analysis for FI on RA, excluding MHC region.

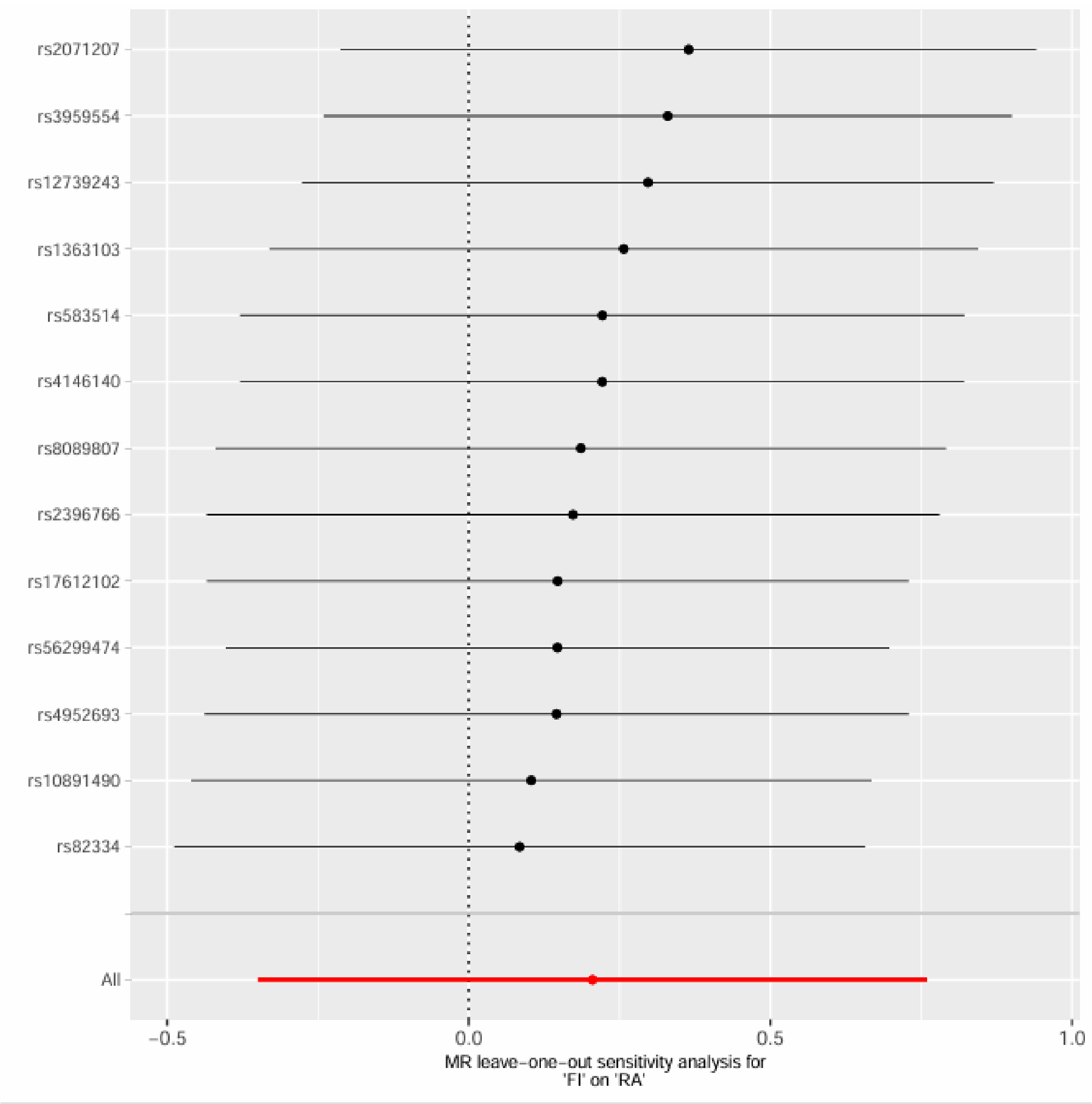

FFS on RA

Supplementary Figure 55. Leave-one-out sensitivity analysis for FFS on RA.

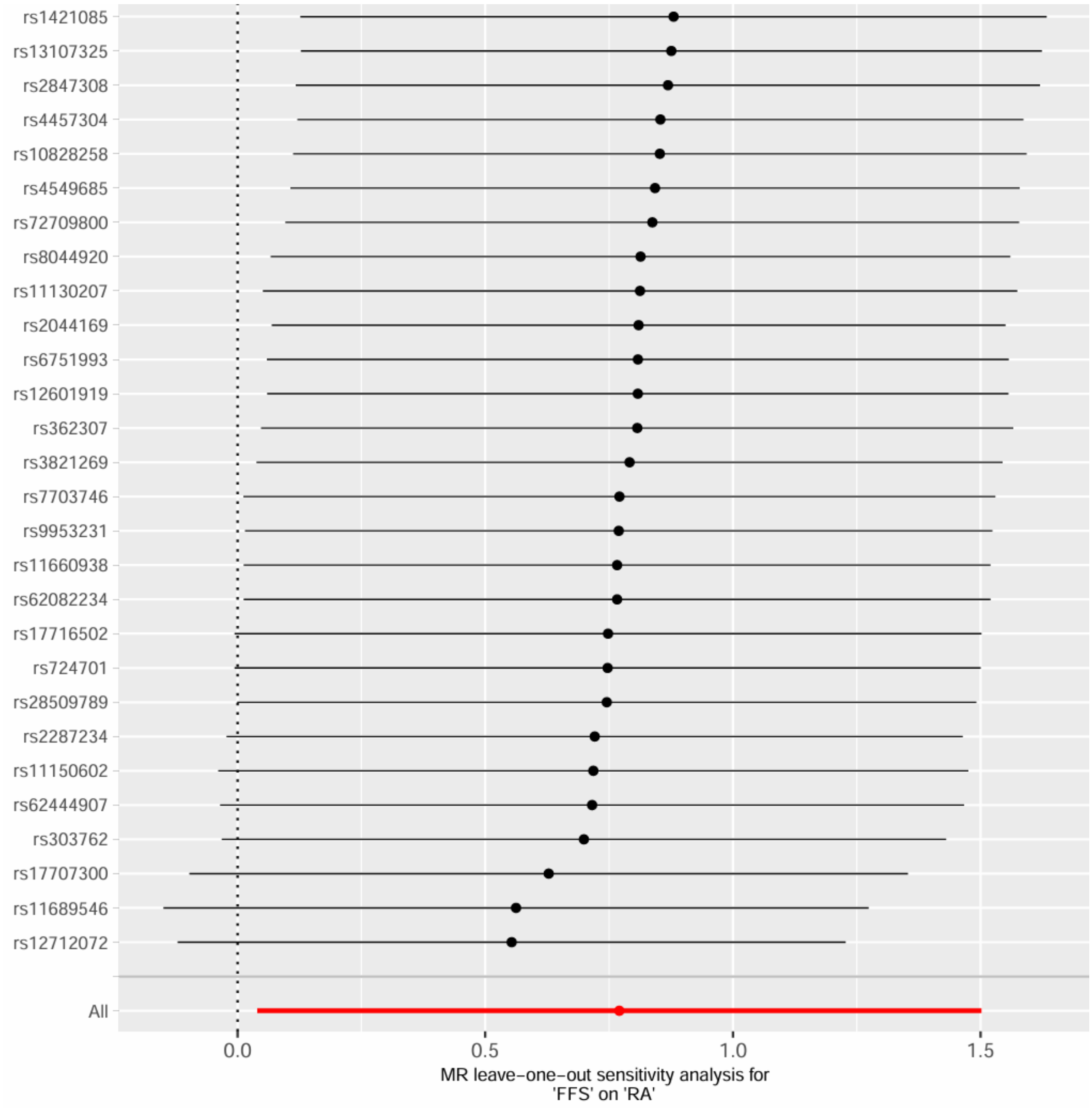

FFS on RA, excluding MHC region

Supplementary Figure 56. Leave-one-out sensitivity analysis for FFS on RA, excluding MHC region.

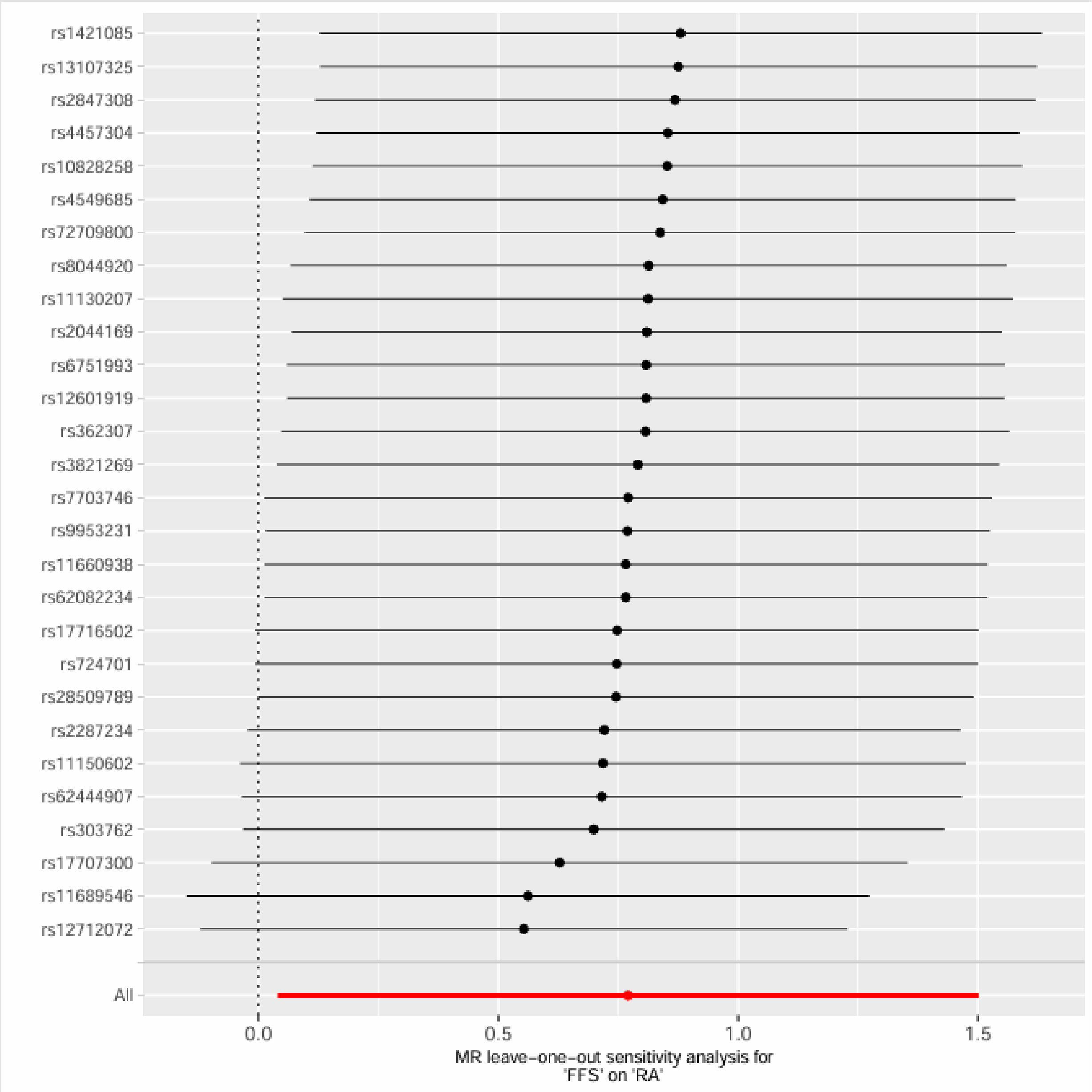

GF on RA

Supplementary Figure 57. Leave-one-out sensitivity analysis for GF on RA.

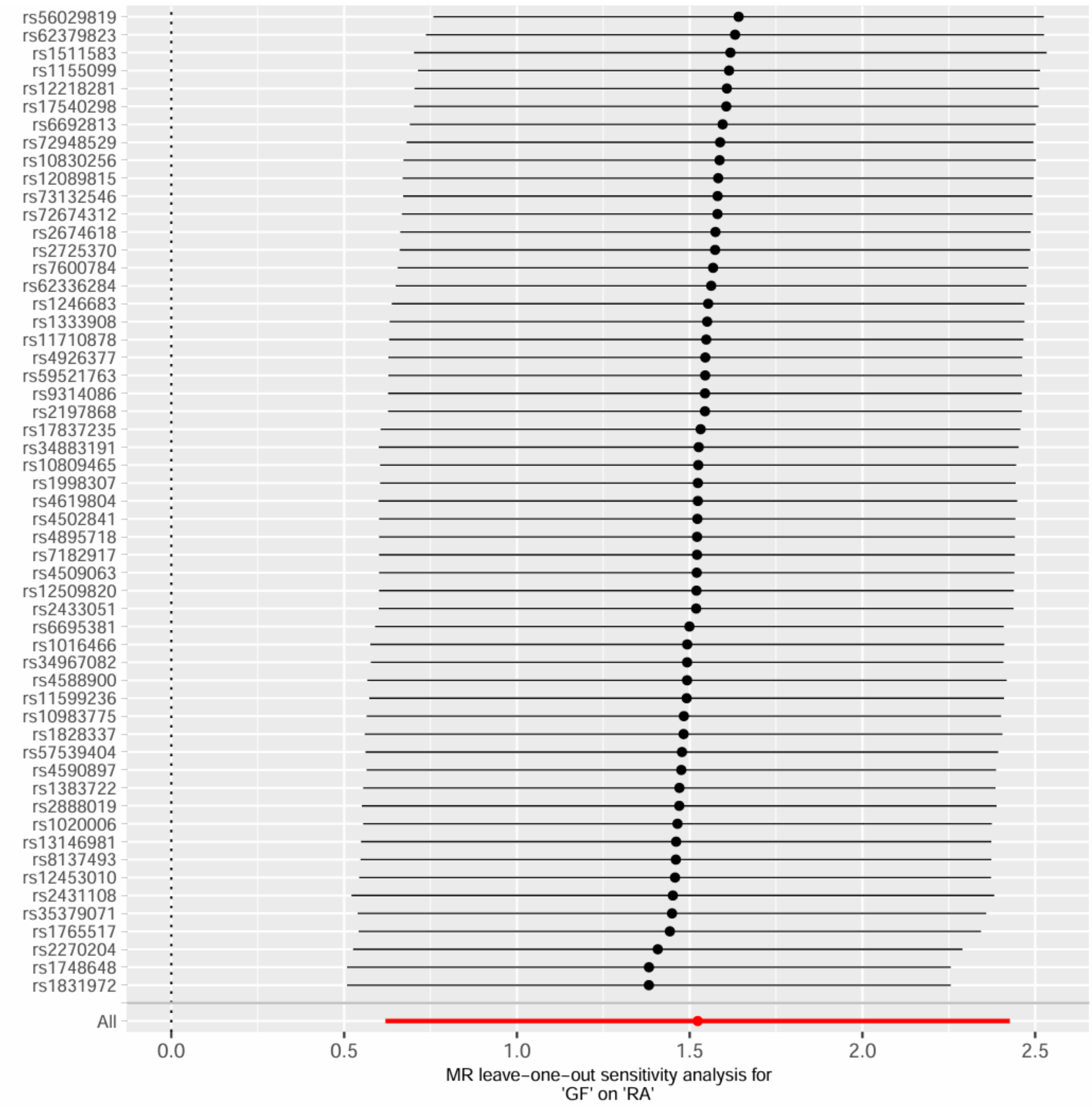

GF on RA, excluding MHC region

Supplementary Figure 58. Leave-one-out sensitivity analysis for GF on RA, excluding MHC region.

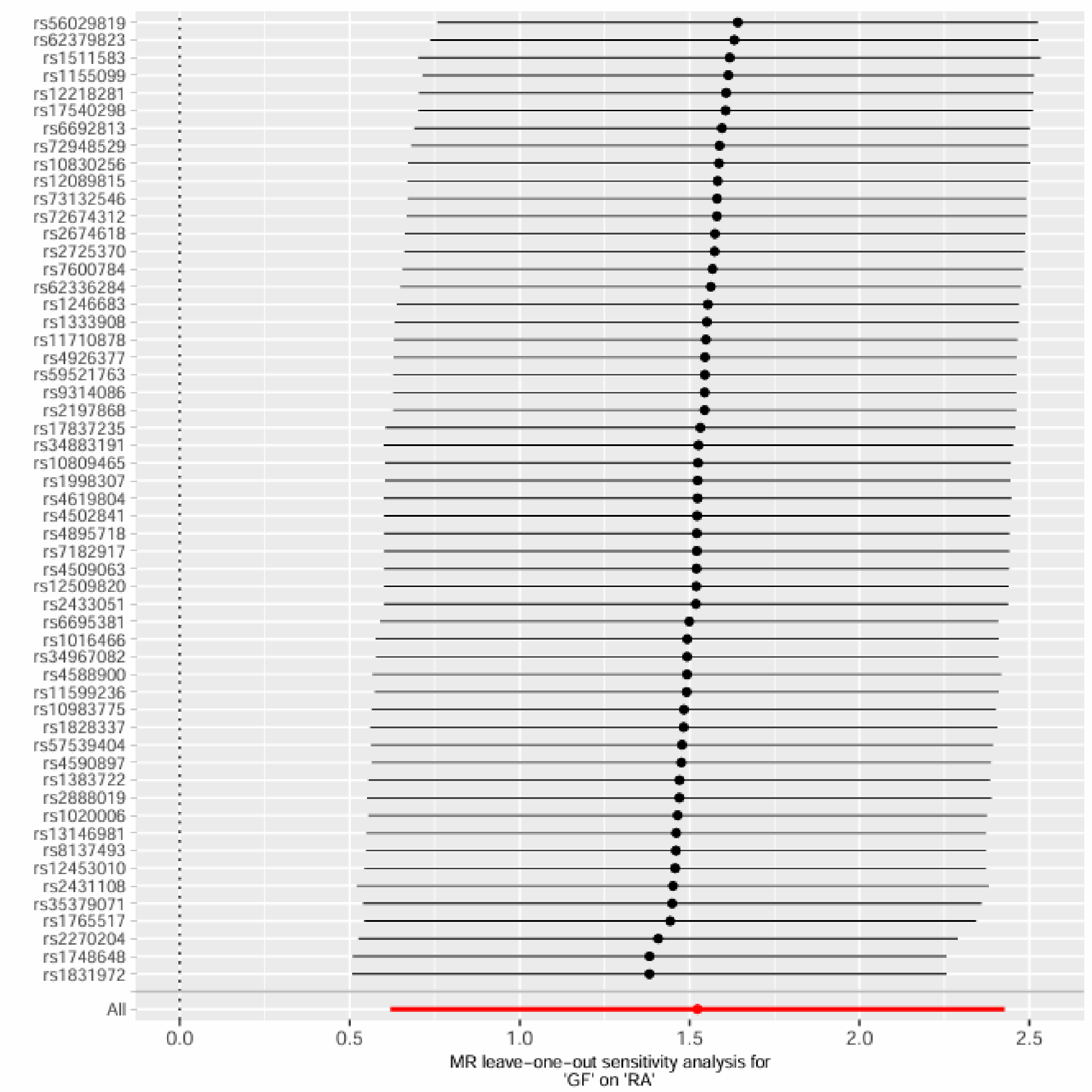

F1 on RA

Supplementary Figure 59. Leave-one-out sensitivity analysis for F1 on RA.

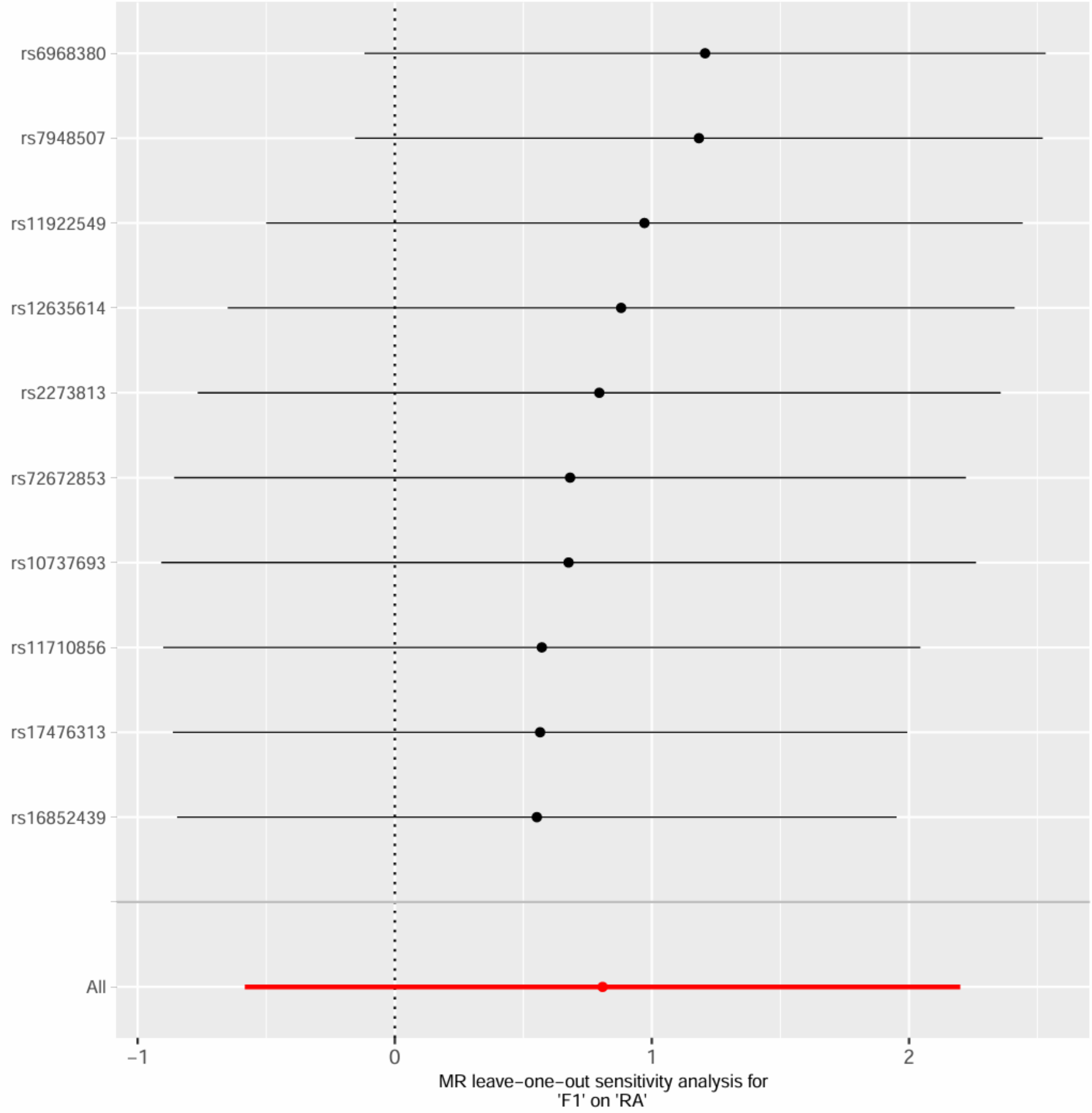


F1 and RA, excluding MHC region

Supplementary Figure 60. Leave-one-out sensitivity analysis for F1 and RA, excluding MHC region.

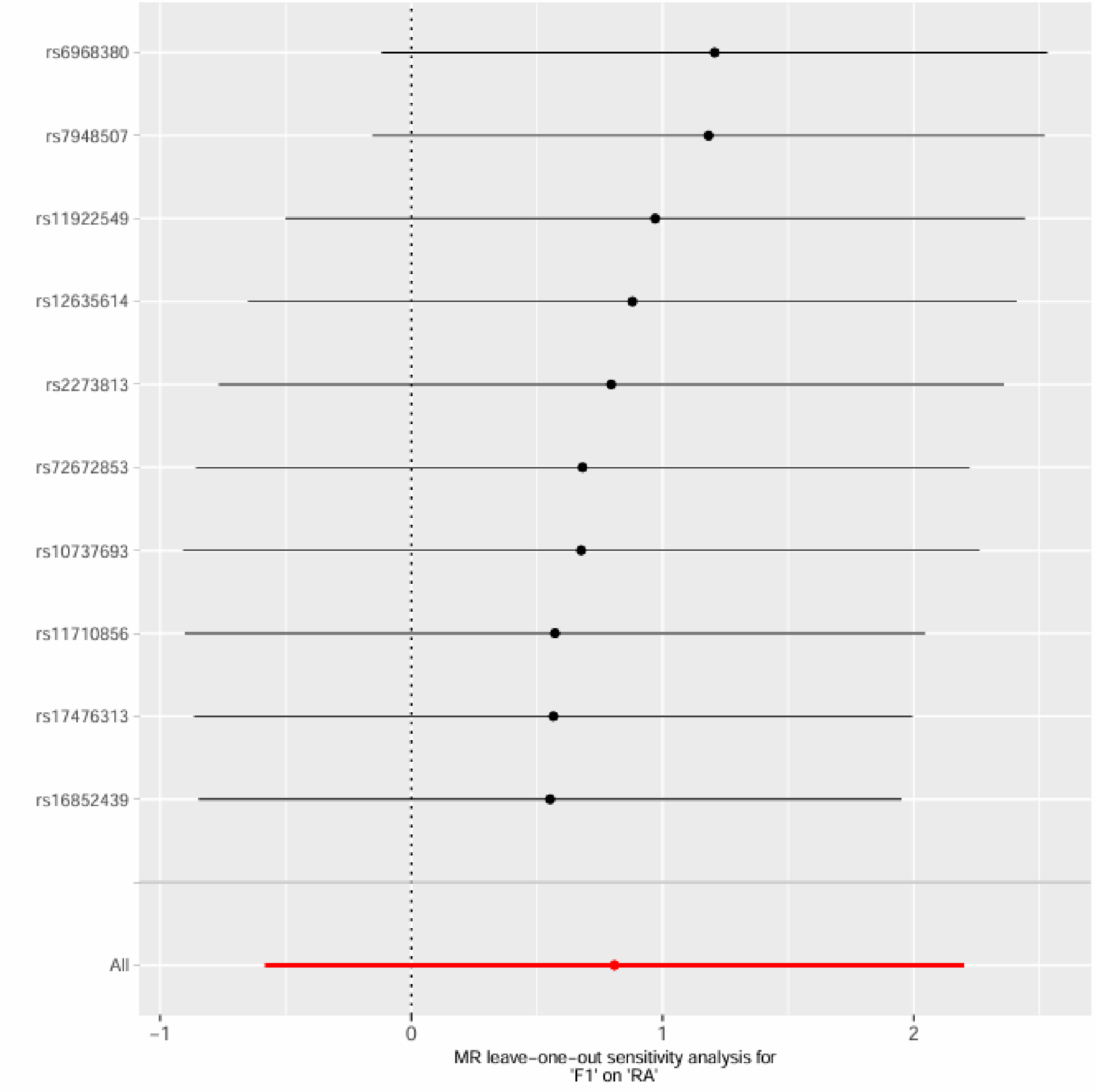
rs6968380
rs7948507
rs11922549
rs12635614
rs2273813
rs72672853
rs10737693
rs11710856
rs17476313
rs16852439
All
-1
0
1
2
MR leave-one-out sensitivity analysis for
'F1' on 'RA'

F2 on RA

Supplementary Figure 61. Leave-one-out sensitivity analysis for F2 on RA.

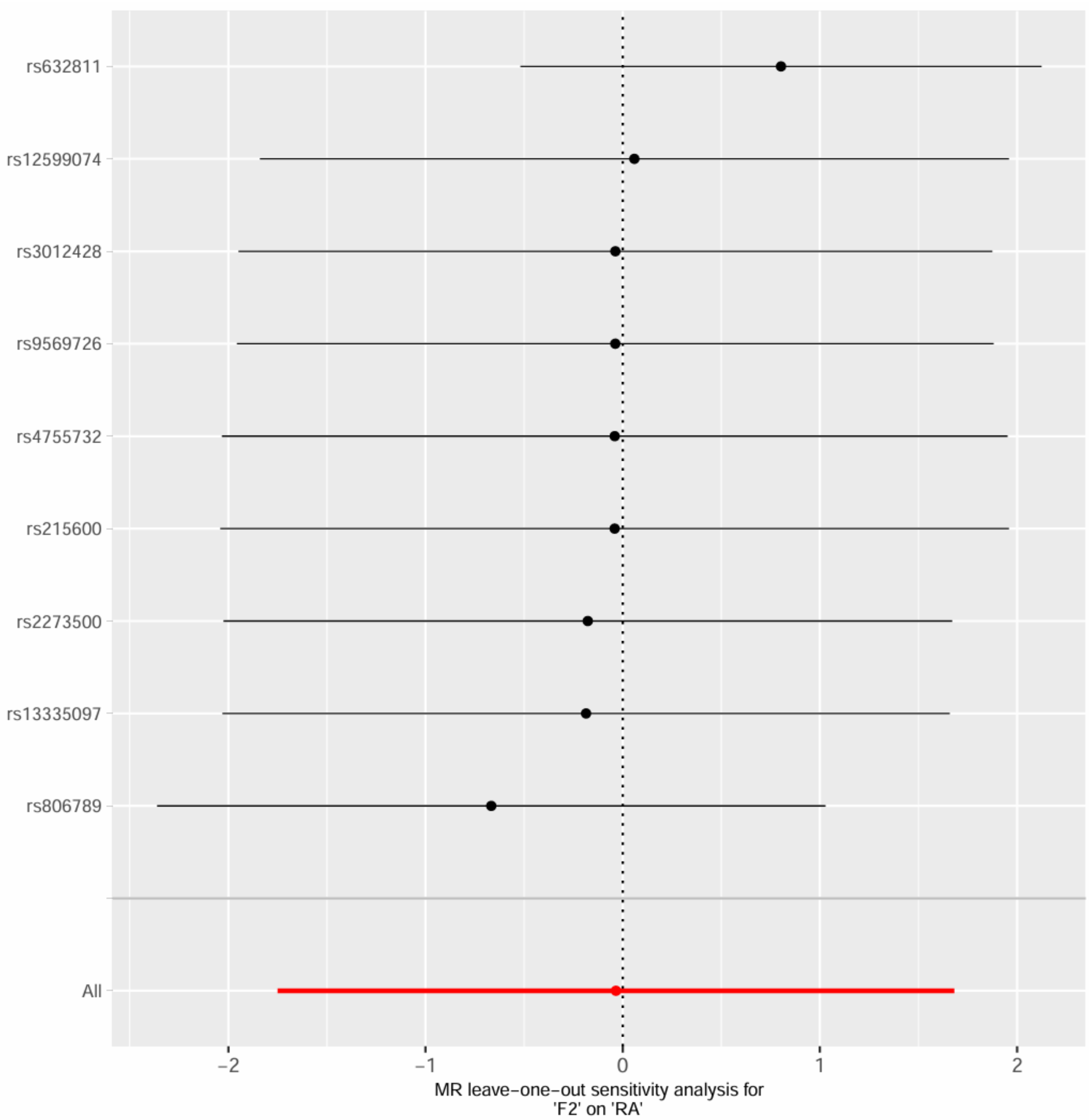


F2 and RA, excluding MHC region

Supplementary Figure 62. Leave-one-out sensitivity analysis for F2 and RA, excluding MHC region.

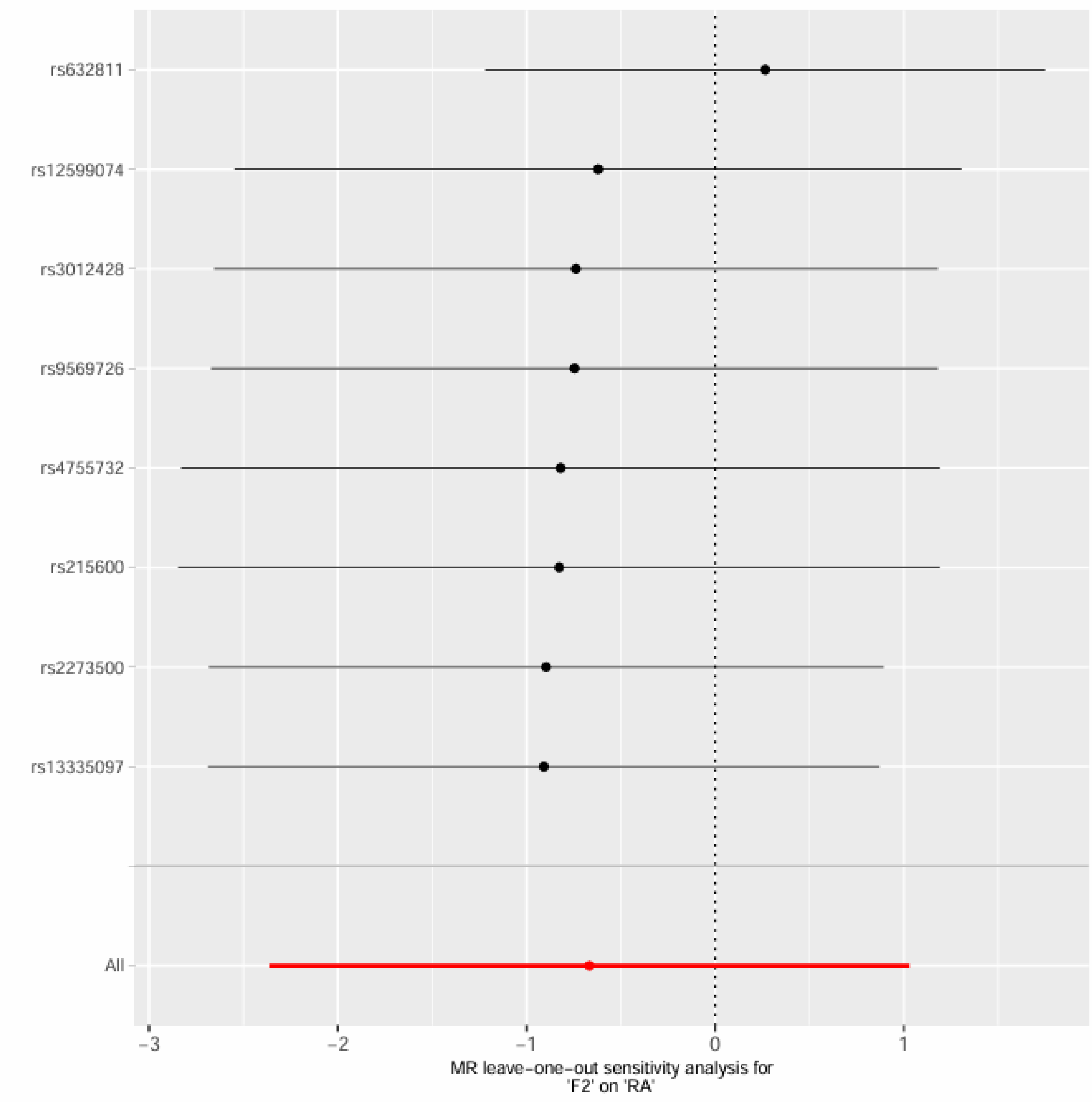

F3 on RA

Supplementary Figure 63. Leave-one-out sensitivity analysis for F3 on RA.

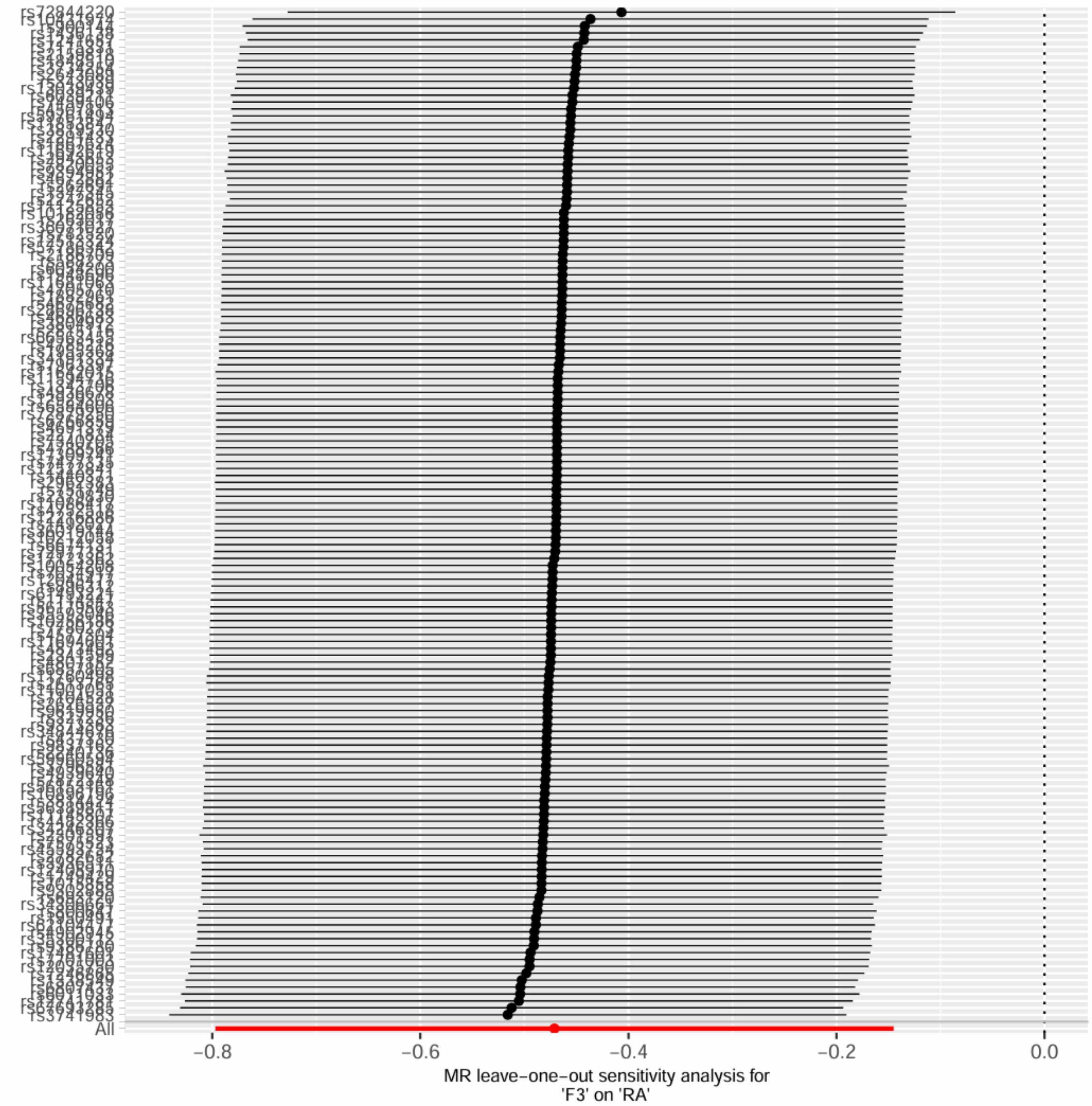

F3 on RA, excluding MHC region

Supplementary Figure 64. Leave-one-out sensitivity analysis for F3 on RA, excluding MHC region.

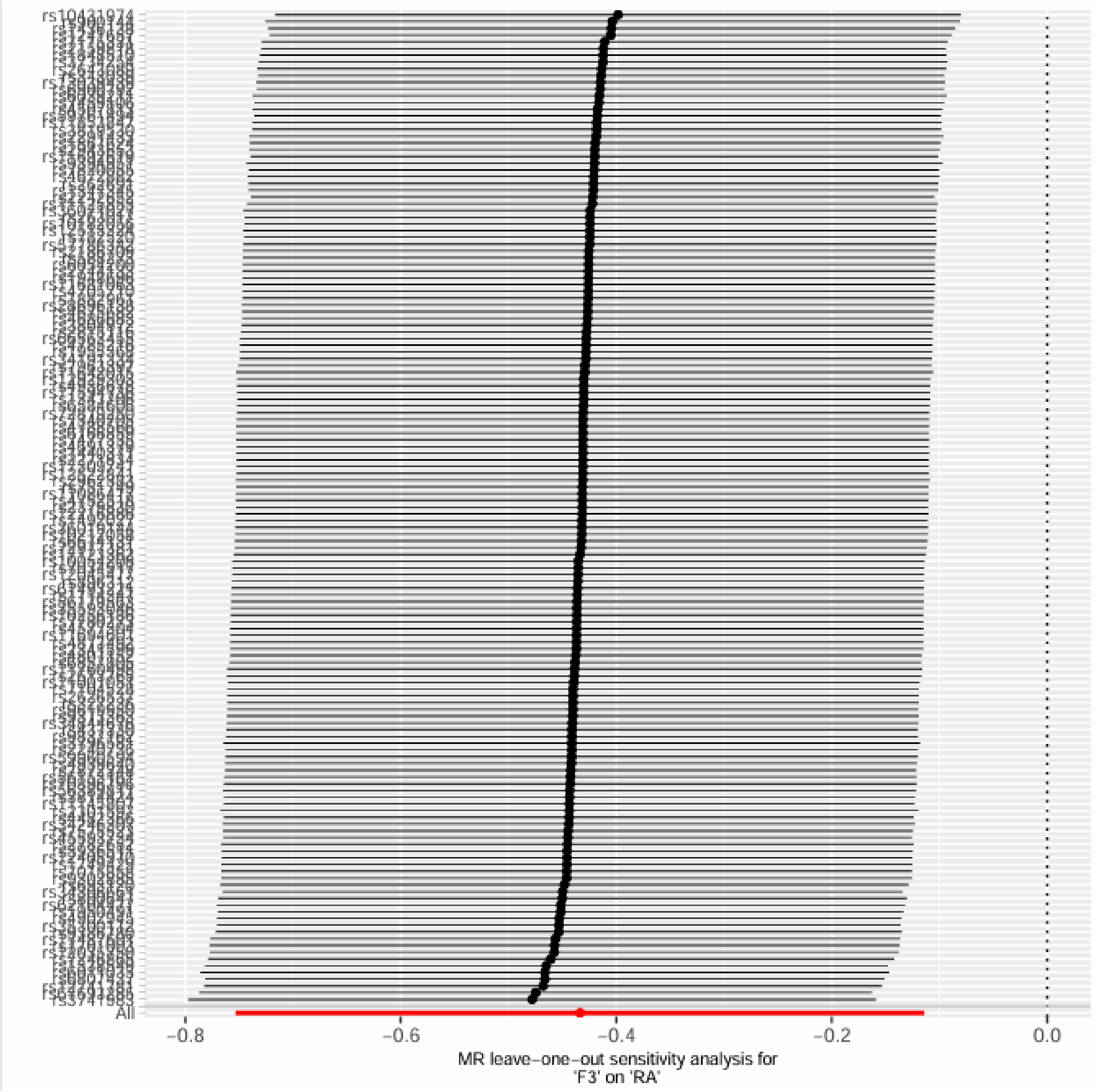

F4 on RA

Supplementary Figure 65. Leave-one-out sensitivity analysis for F4 on RA.

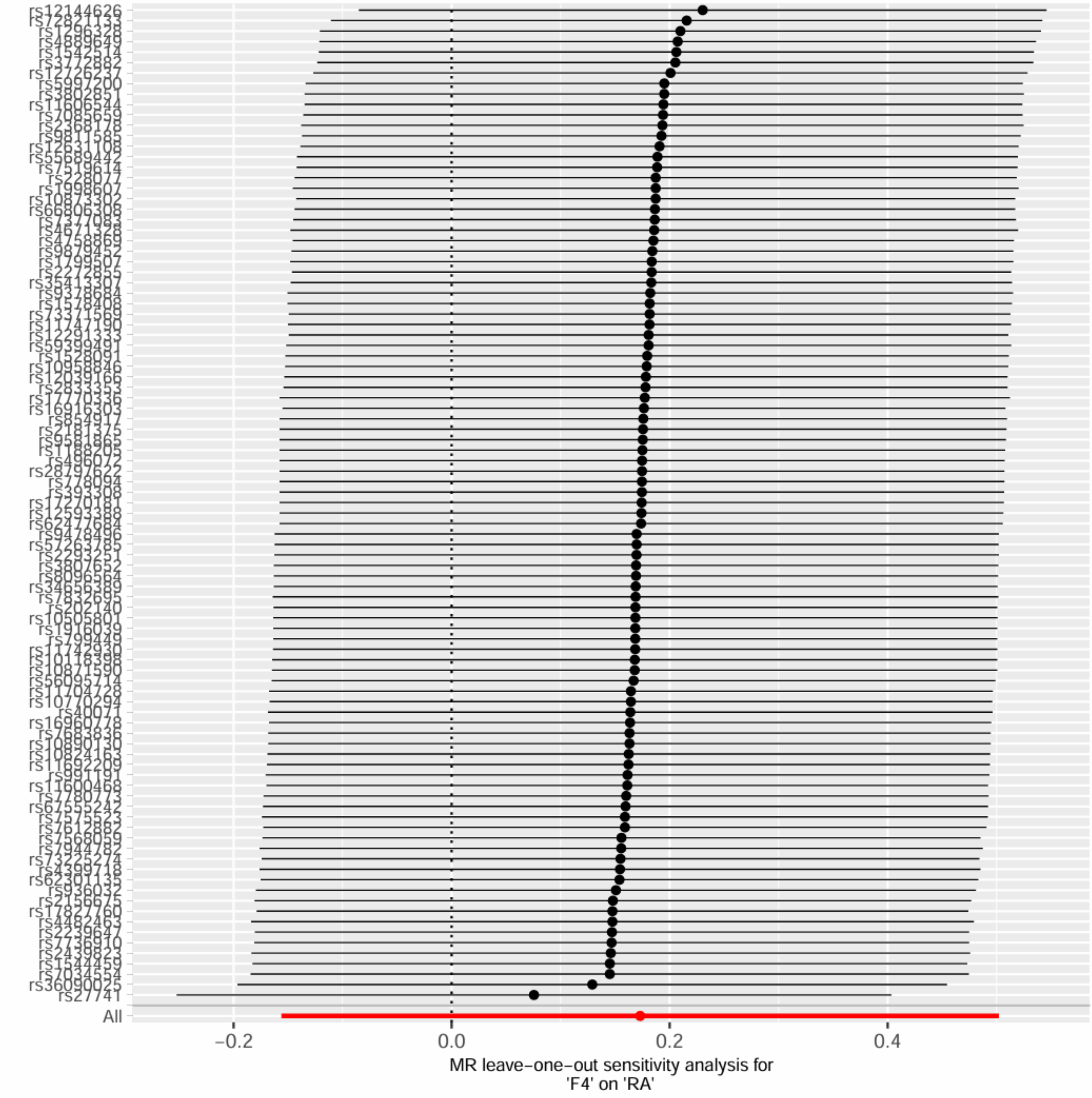

F4 on RA, excluding MHC region

Supplementary Figure 66. Leave-one-out sensitivity analysis for F4 on RA, excluding MHC region.

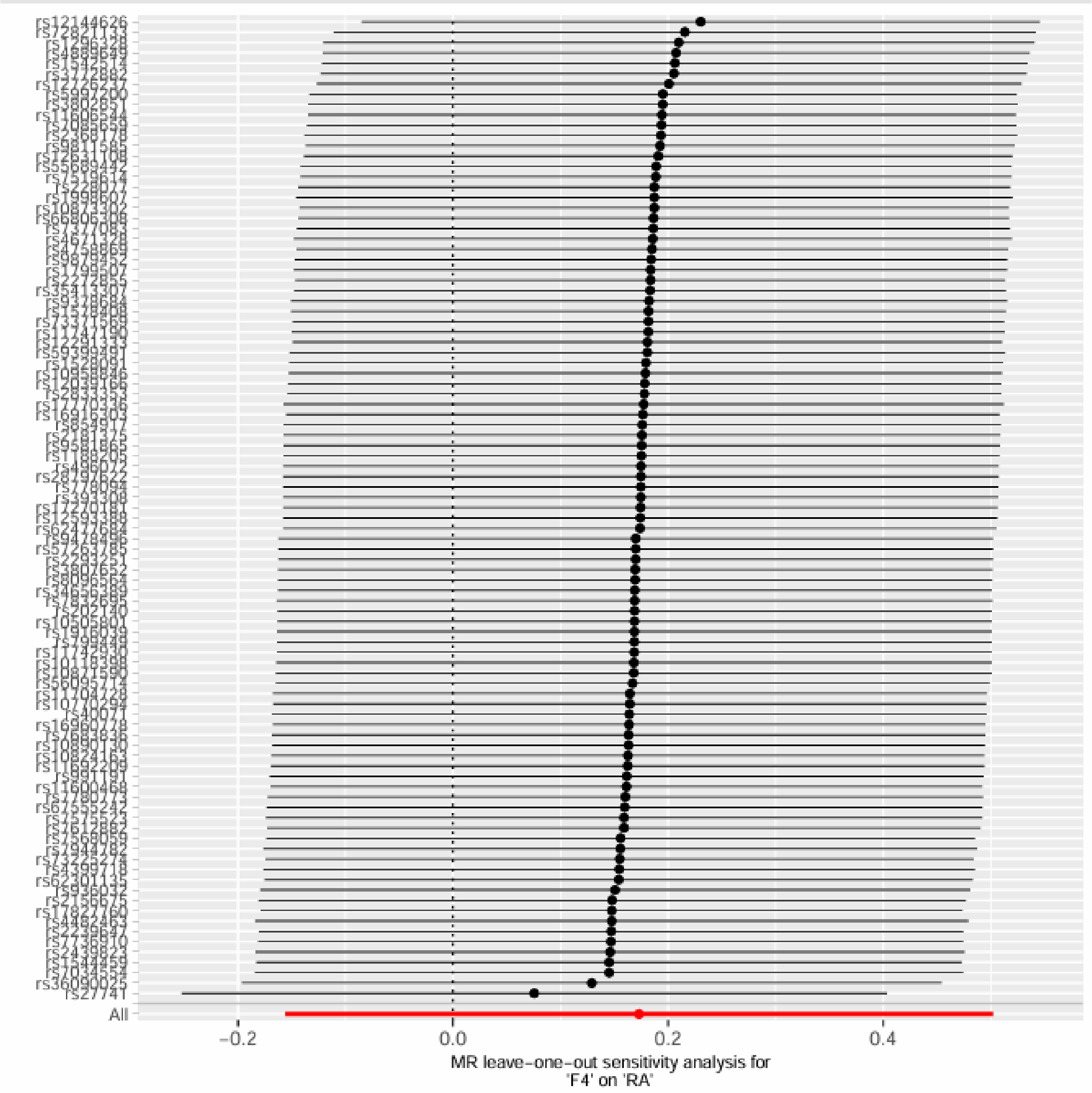

F5 on RA

Supplementary Figure 67. Leave-one-out sensitivity analysis for F5 on RA.

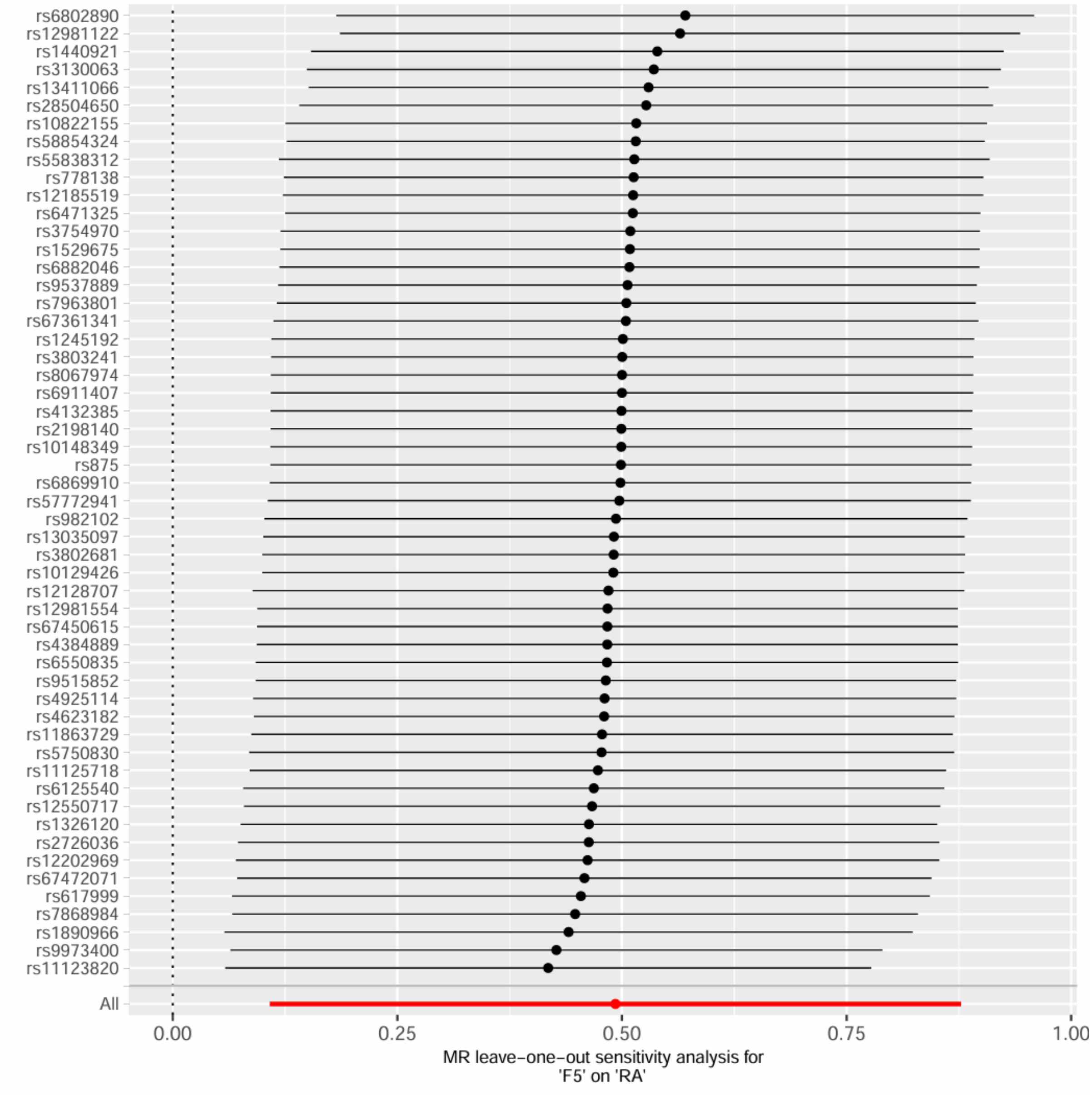

F5 and RA, excluding MHC region

Supplementary Figure 68. Leave-one-out sensitivity analysis for F5 and RA, excluding MHC region.

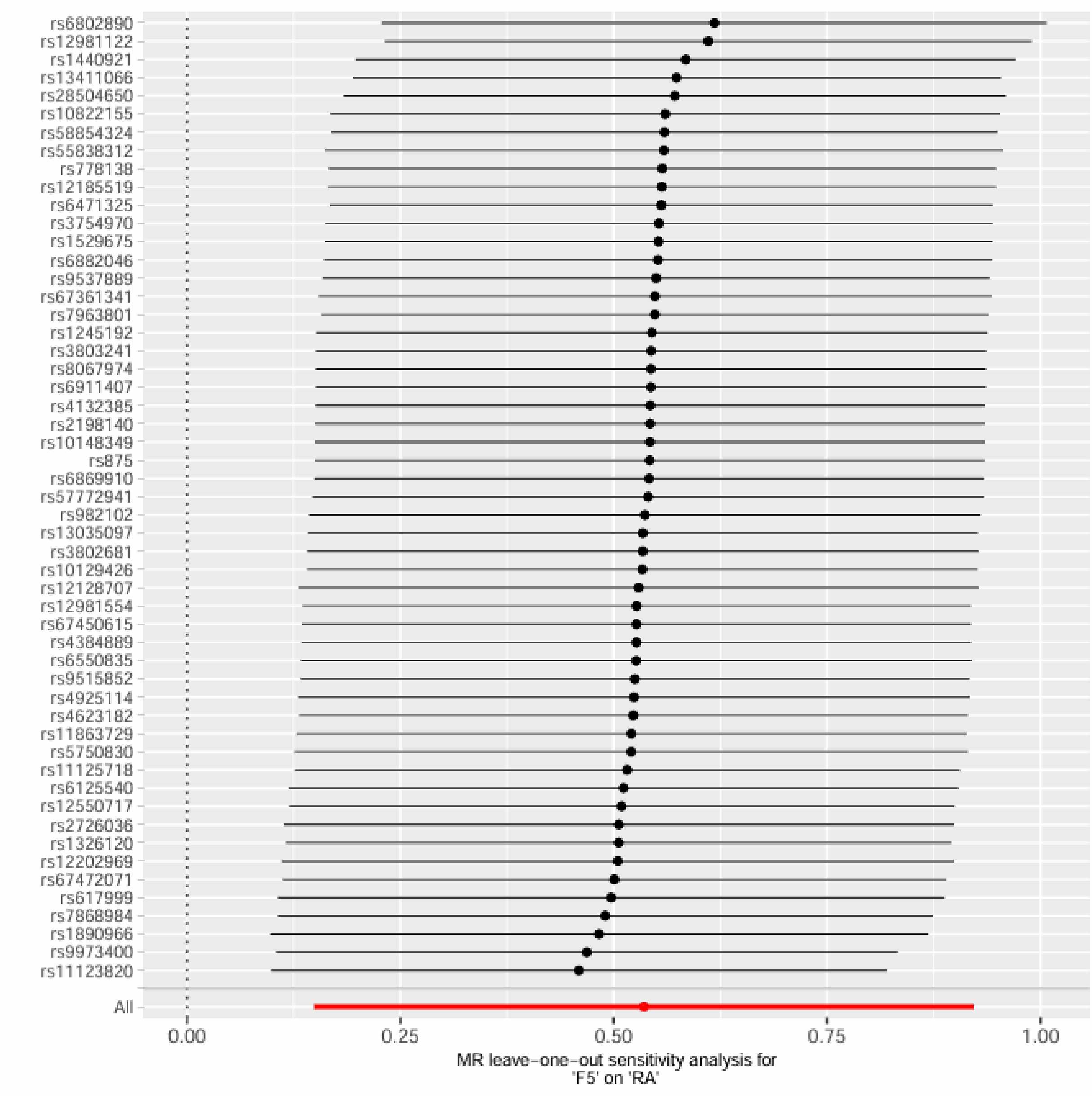

F6 on RA

Supplementary Figure 69. Leave-one-out sensitivity analysis for F6 on RA.

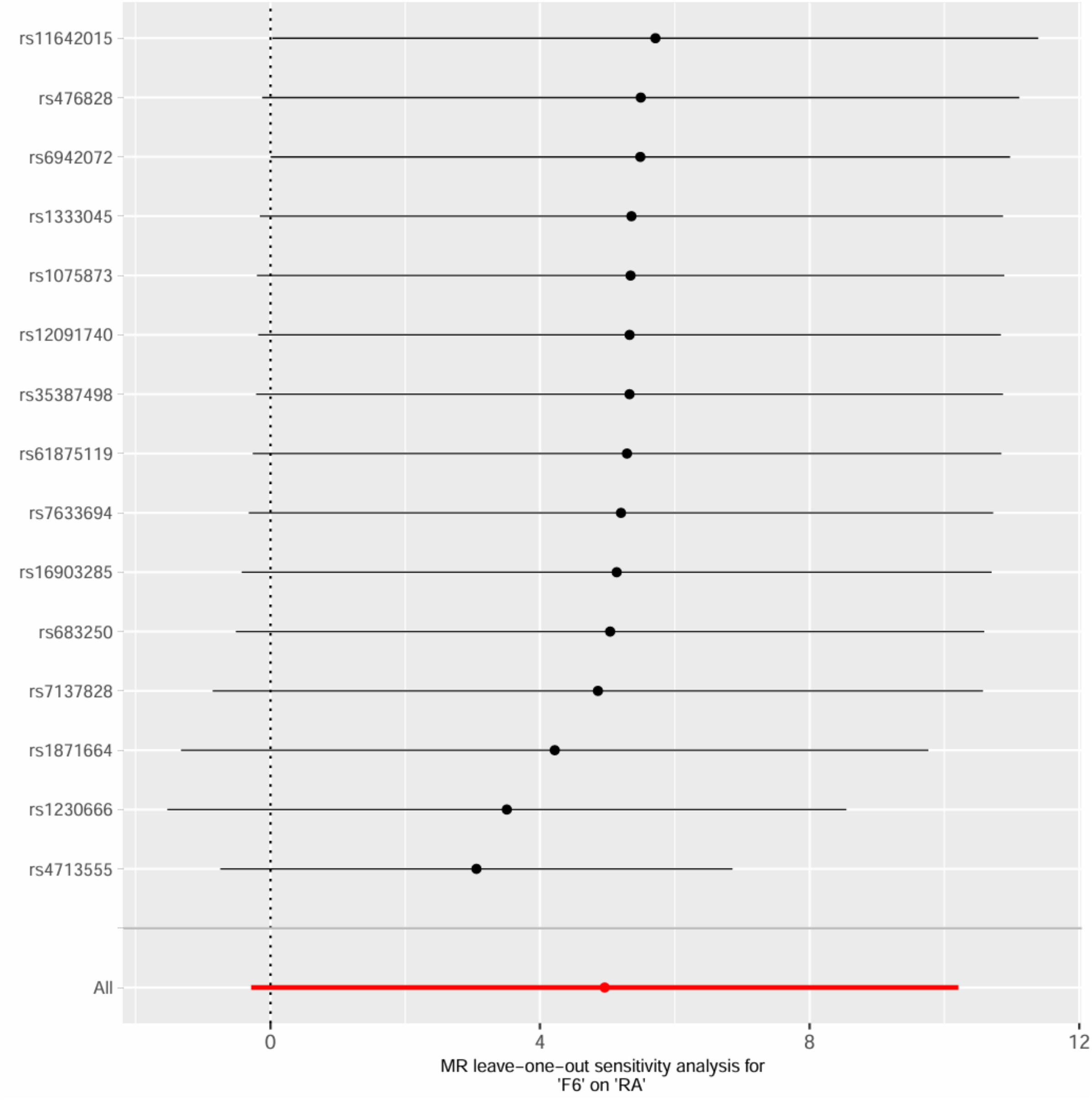

F6 and RA, excluding MHC region

Supplementary Figure 70. Leave-one-out sensitivity analysis for F6 and RA, excluding MHC region.

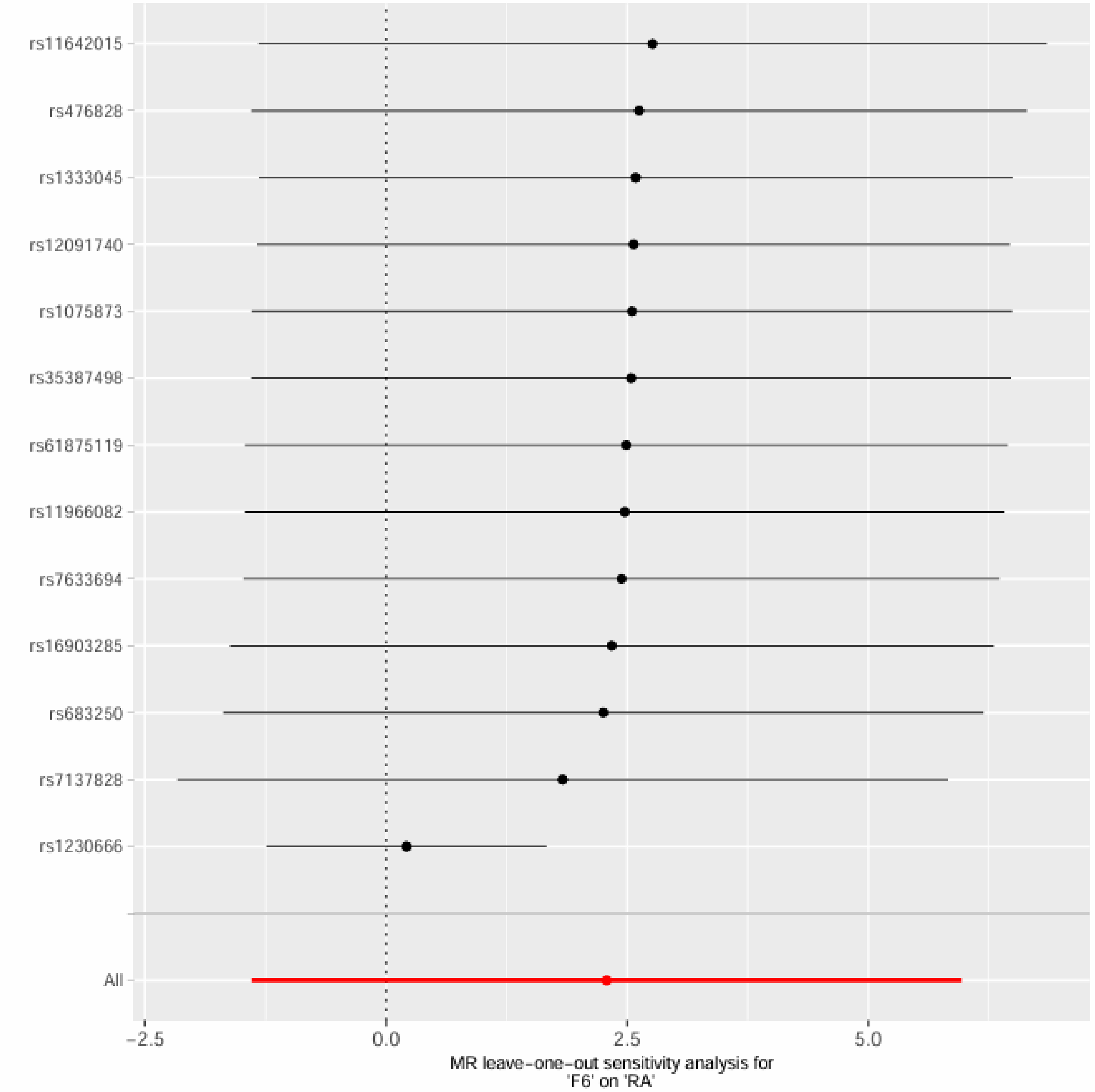

rs11642015
rs476828
rs1333045
rs12091740
rs1075873
rs35387498
rs61875119
rs11966082
rs7633694
rs16903285
rs683250
rs7137828
rs1230666
All
−2.5
0.0
2.5
5.0
MR leave−one−out sensitivity analysis for
'F6' on 'RA'

90% Credible Sets and Gene-Level Plots

Shared 90% credible sets were derived from genomic regions supporting a shared causal variant (PW-GWAS Model 3). The upper panel of each figure summarises genomic regions containing shared credible-set variants, with bar length representing the number of variants in the 90% credible set and colour representing the lead posterior probability of association ($PPA_3$). The lower panel prioritises nearby genes according to the maximum variant-level $PPA_3$ observed within each gene, highlighting candidate genes underlying the shared genetic architecture.

*F1 and CP*

Supplementary Figure 71. Shared 90% credible-set variants and gene-level prioritisation for F1 × chronic pain.

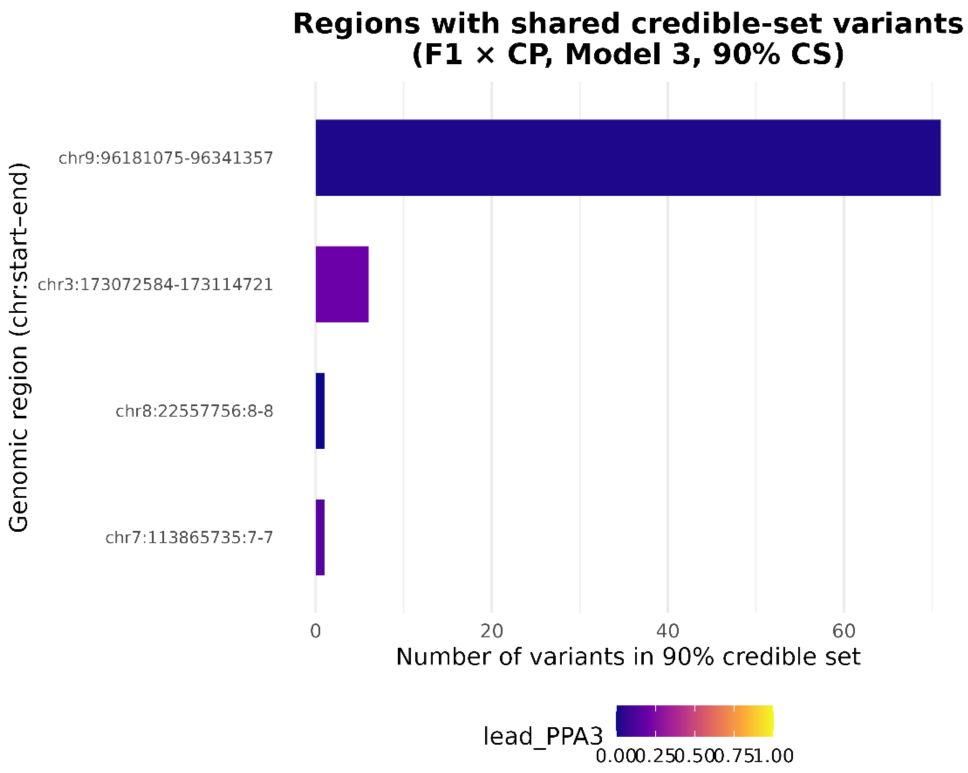

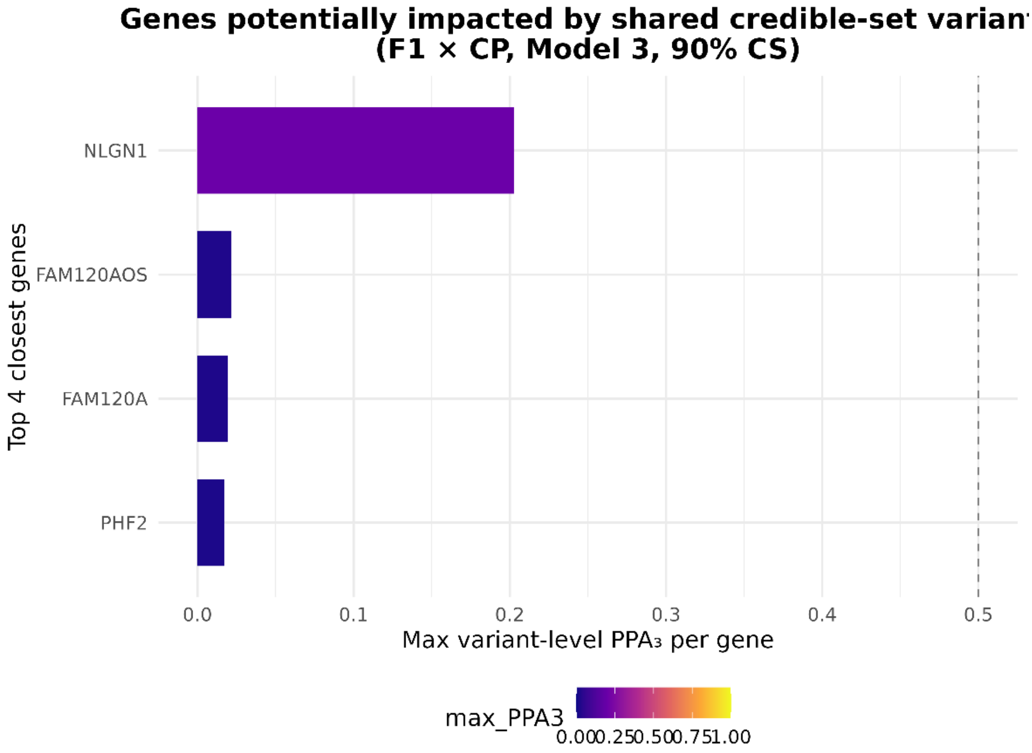


*F2 X CP*

Supplementary Figure 72. Shared 90% credible-set variants and gene-level prioritisation for F2 × chronic pain.

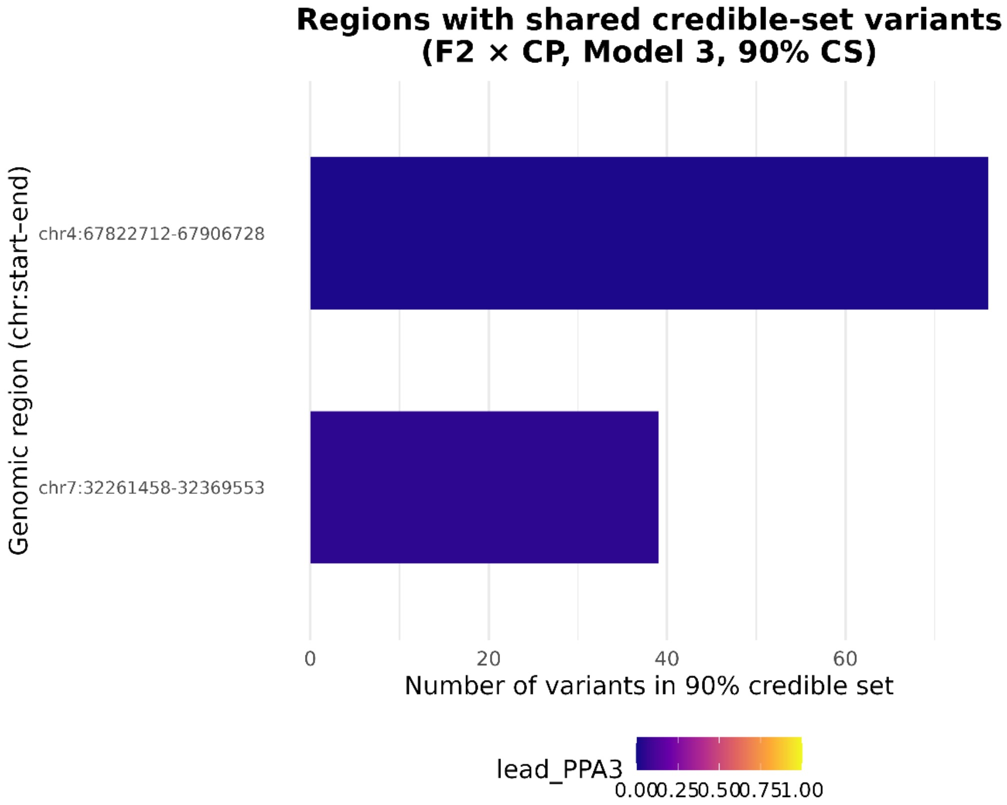

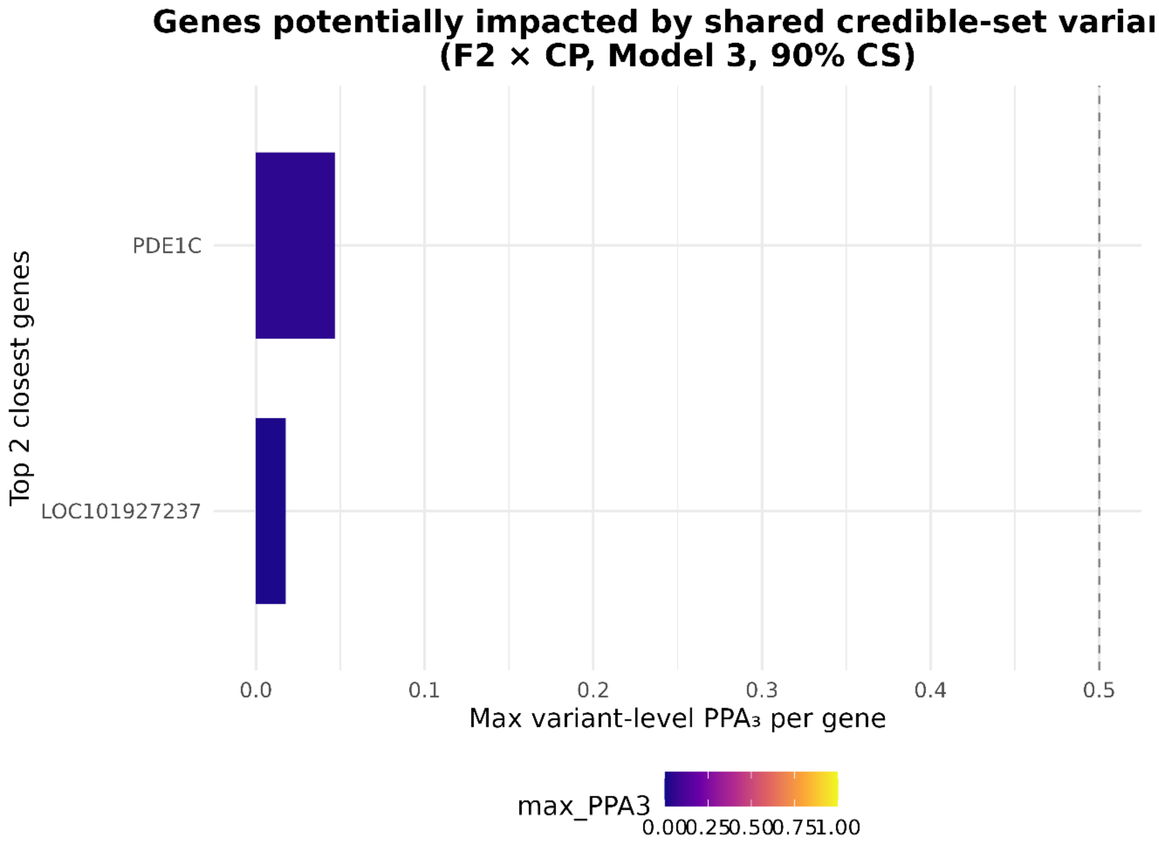


*F3 X CP*

Supplementary Figure 73. Shared 90% credible-set variants and gene-level prioritisation for F3 × chronic pain.

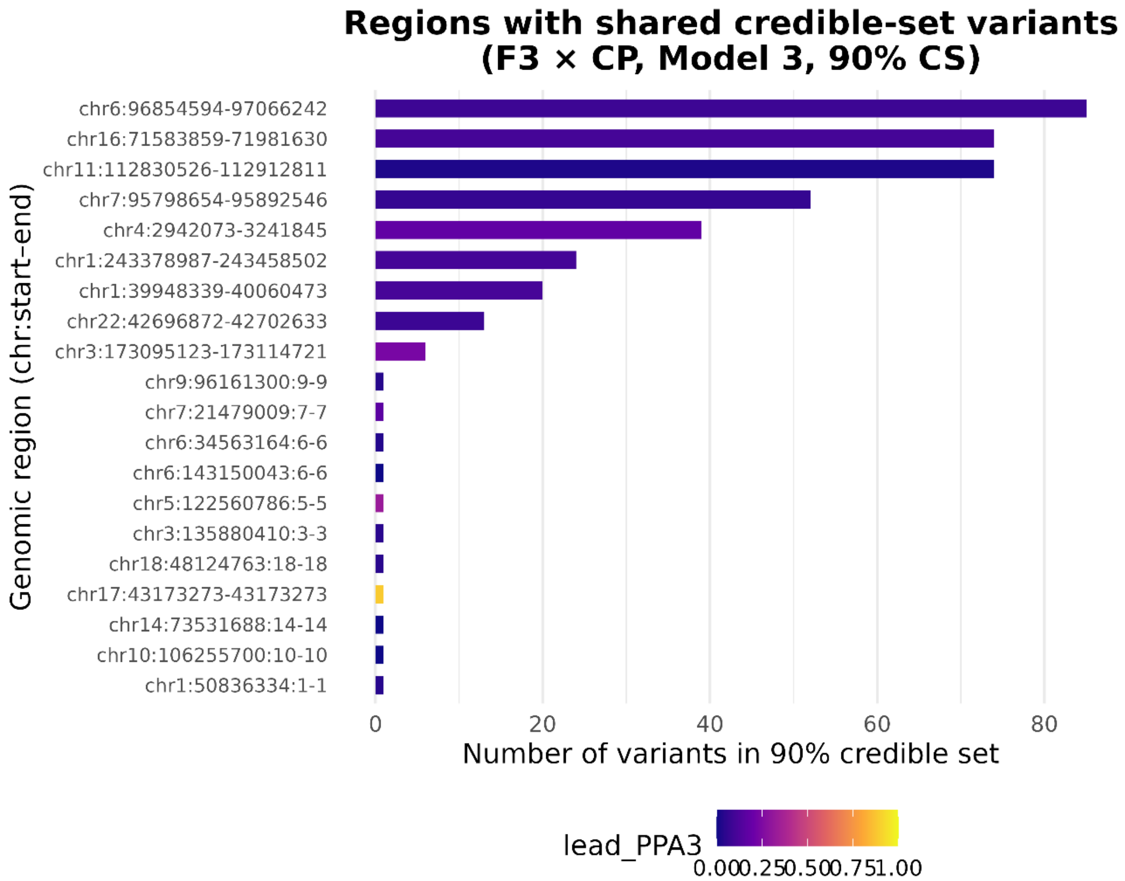
Regions with shared credible-set variants
(F3 × CP, Model 3, 90% CS)
chr6:96854594-97066242
chr16:71583859-71981630
chr11:112830526-112912811
chr7:95798654-95892546
chr4:2942073-3241845
chr1:243378987-243458502
chr1:39948339-40060473
chr22:42696872-42702633
chr3:173095123-173114721
chr9:96161300:9-9
chr7:21479009:7-7
chr6:34563164:6-6
chr6:143150043:6-6
chr5:122560786:5-5
chr3:135880410:3-3
chr18:48124763:18-18
chr17:43173273-43173273
chr14:73531688:14-14
chr10:106255700:10-10
chr1:50836334:1-1
Genomic region (chr:start–end)
0
20
40
60
80
Number of variants in 90% credible set
lead_PPA3
0.00
0.25
0.50
0.75
1.00

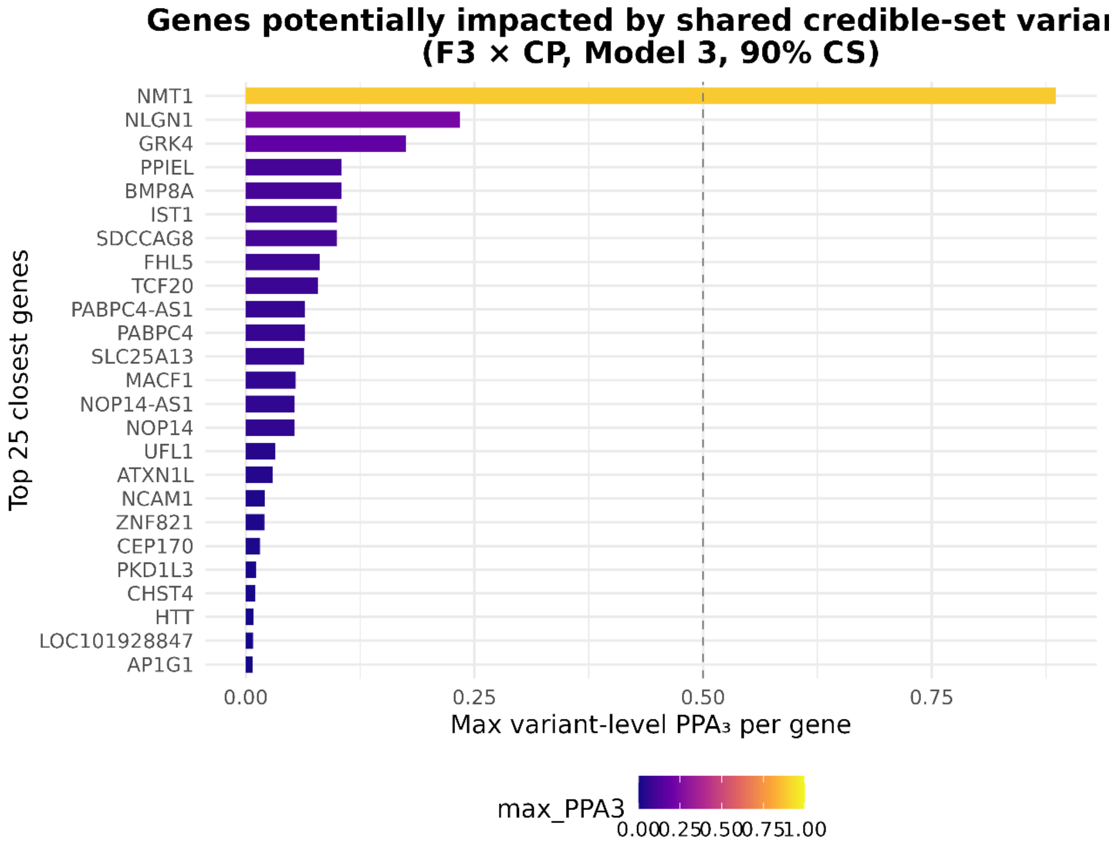
Genes potentially impacted by shared credible-set variai
(F3 × CP, Model 3, 90% CS)
NMT1
NLGN1
GRK4
PPIEL
BMP8A
IST1
SDCCAG8
FHL5
TCF20
PABPC4-AS1
PABPC4
SLC25A13
MACF1
NOP14-AS1
NOP14
UFL1
ATXN1L
NCAM1
ZNF821
CEP170
PKD1L3
CHST4
HTT
LOC101928847
AP1G1
Top 25 closest genes
0.00
0.25
0.50
0.75
Max variant-level PPA₃ per gene
max_PPA3
0.00
0.25
0.50
0.75
1.00

*F4 X CP*

Supplementary Figure 74. Shared 90% credible-set variants and gene-level prioritisation for F4 × chronic pain.

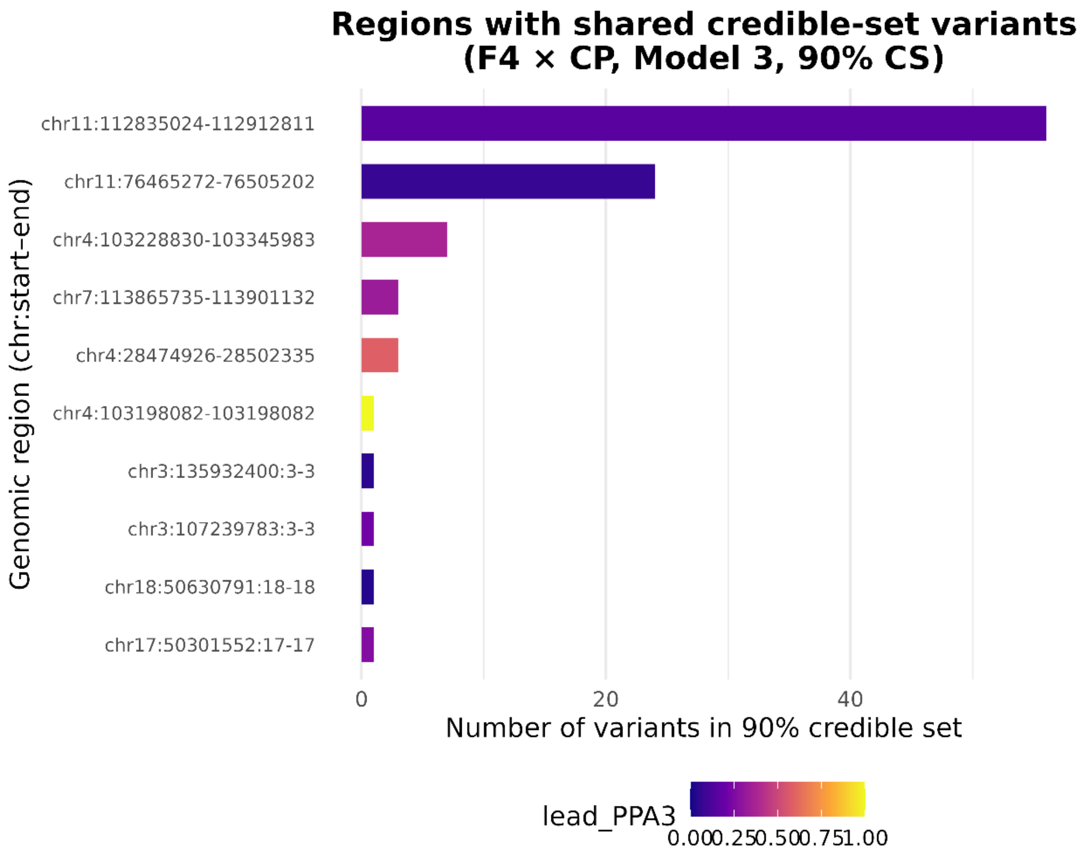

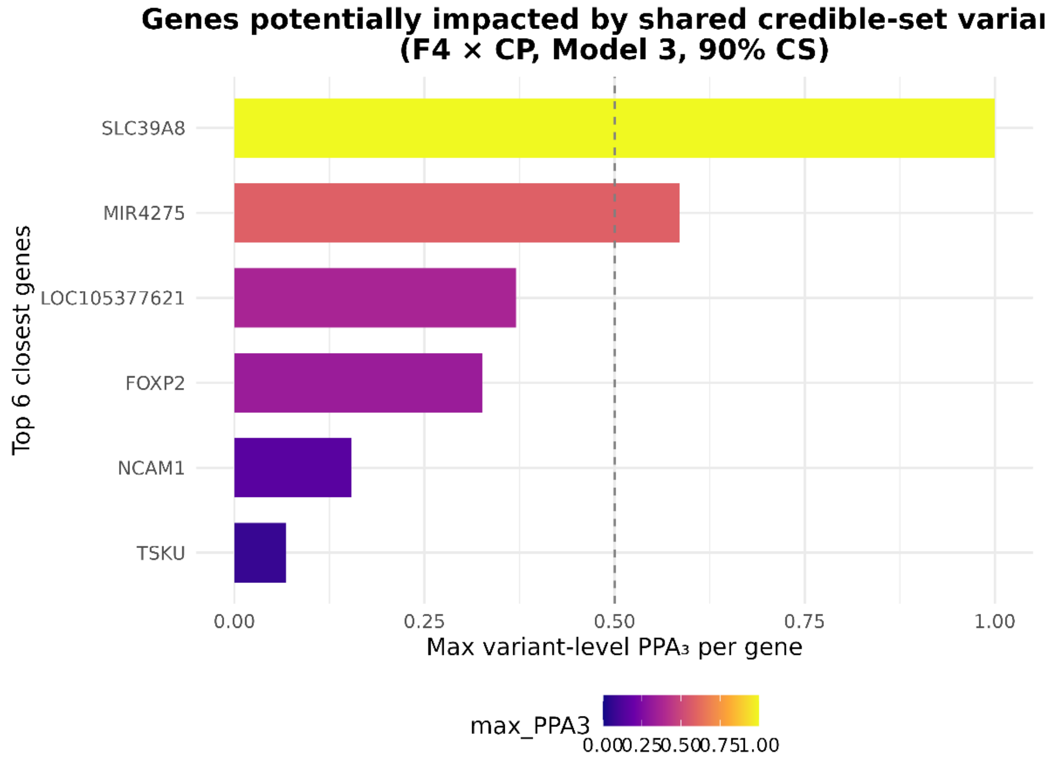


*F5 X CP*

Supplementary Figure 75. Shared 90% credible-set variants and gene-level prioritisation for F5 × chronic pain.

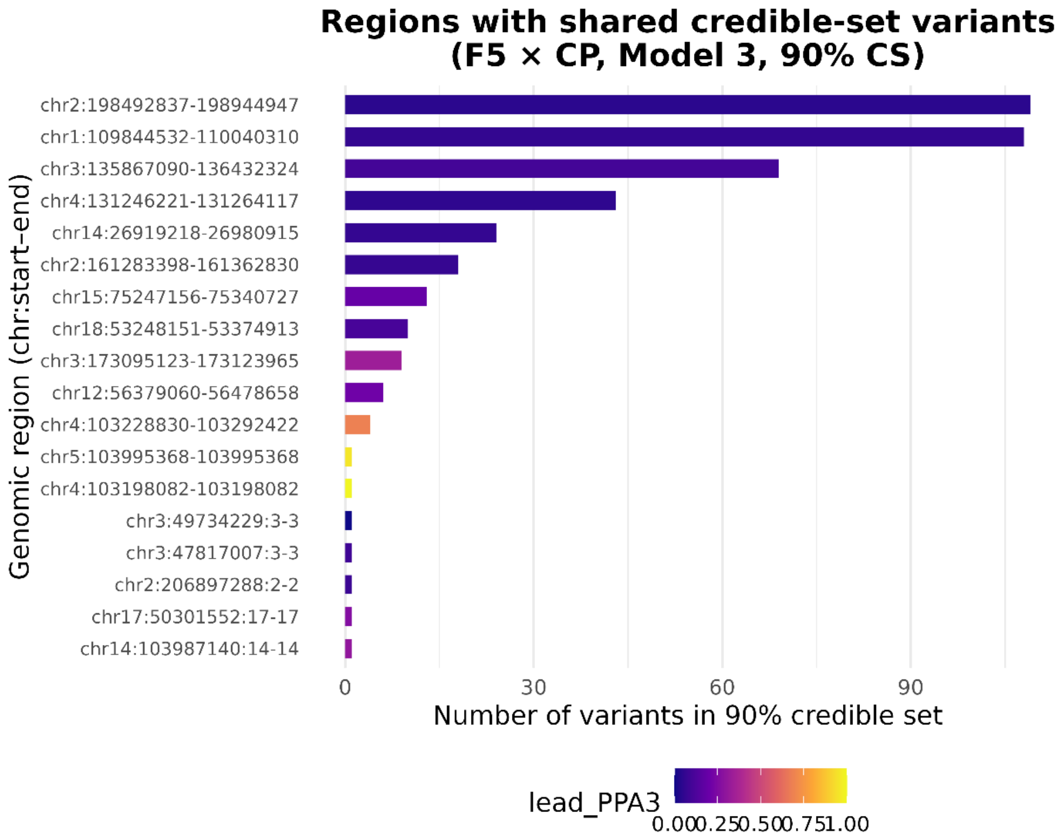
Regions with shared credible-set variants
(F5 × CP, Model 3, 90% CS)
chr2:198492837-198944947
chr1:109844532-110040310
chr3:135867090-136432324
chr4:131246221-131264117
chr14:26919218-26980915
chr2:161283398-161362830
chr15:75247156-75340727
chr18:53248151-53374913
chr3:173095123-173123965
chr12:56379060-56478658
chr4:103228830-103292422
chr5:103995368-103995368
chr4:103198082-103198082
chr3:49734229:3-3
chr3:47817007:3-3
chr2:206897288:2-2
chr17:50301552:17-17
chr14:103987140:14-14
Genomic region (chr:start–end)
0
30
60
90
Number of variants in 90% credible set
lead_PPA3
0.00 0.25 0.50 0.75 1.00

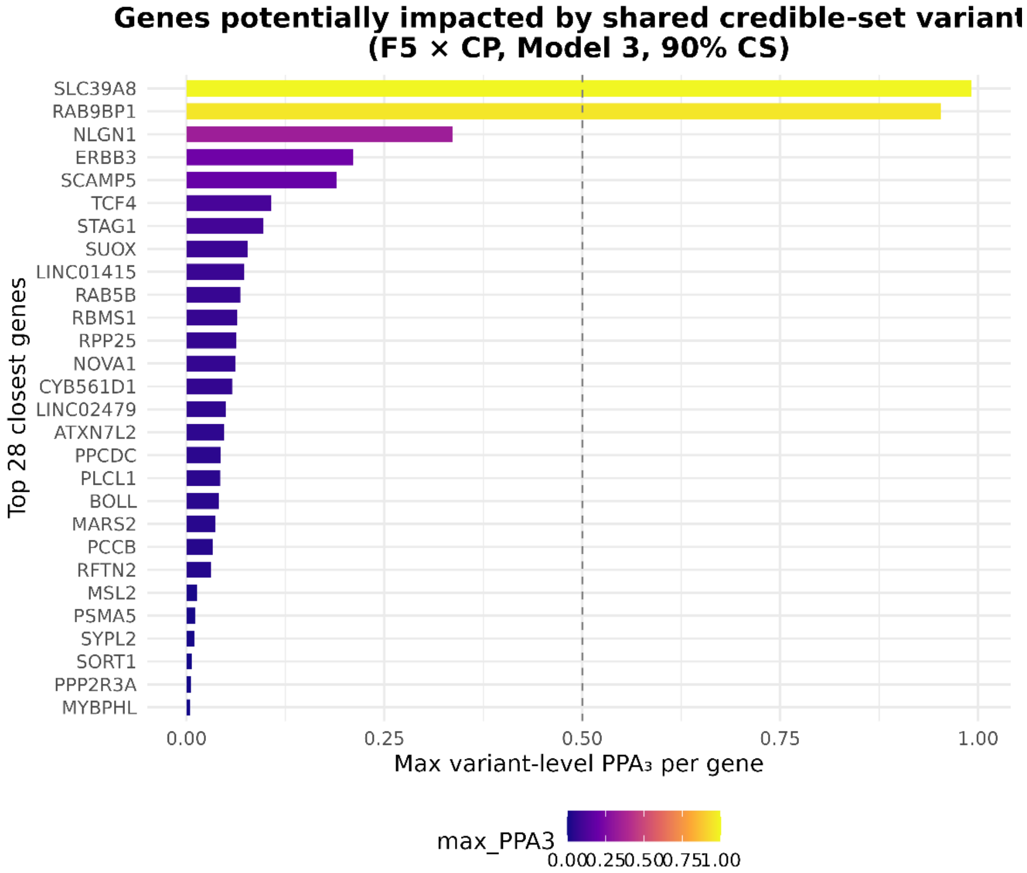
Genes potentially impacted by shared credible-set variant
(F5 × CP, Model 3, 90% CS)
SLC39A8
RAB9BP1
NLGN1
ERBB3
SCAMP5
TCF4
STAG1
SUOX
LINC01415
RAB5B
RBMS1
RPP25
NOVA1
CYB561D1
LINC02479
ATXN7L2
PPCDC
PLCL1
BOLL
MARS2
PCCB
RFTN2
MSL2
PSMA5
SYPL2
SORT1
PPP2R3A
MYBPHL
Top 28 closest genes
0.00
0.25
0.50
0.75
1.00
Max variant-level PPA3 per gene
max_PPA3
0.00 0.25 0.50 0.75 1.00

*F6 X CP*

Supplementary Figure 76. Shared 90% credible-set variants and gene-level prioritisation for F6 × chronic pain.

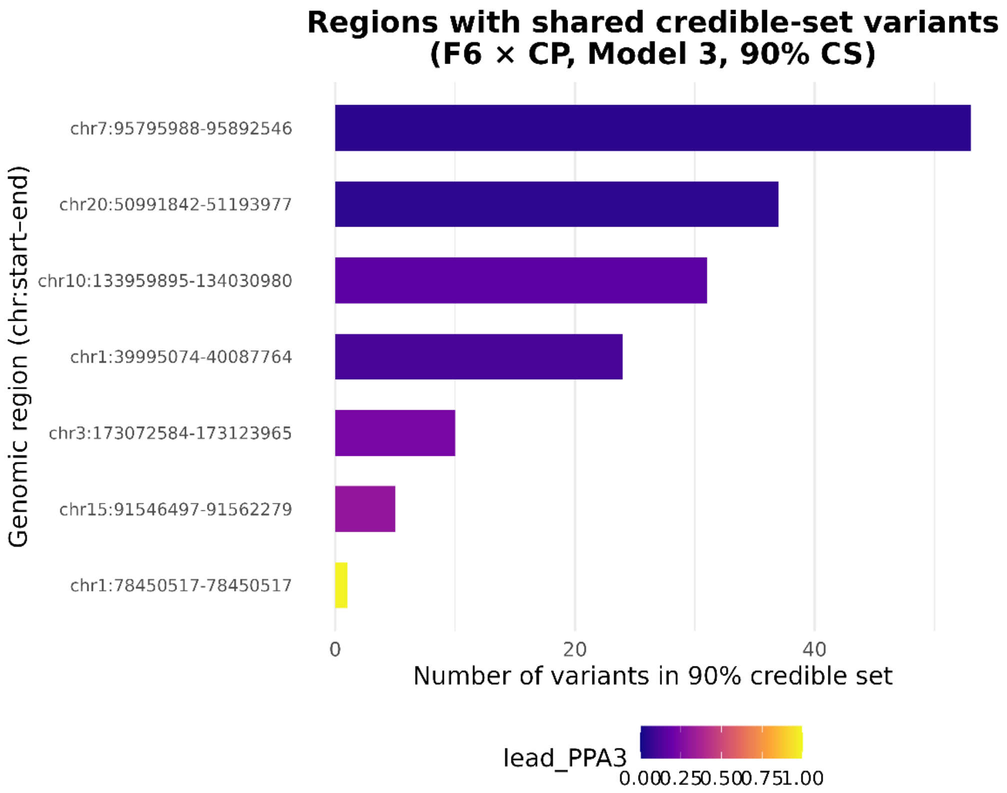


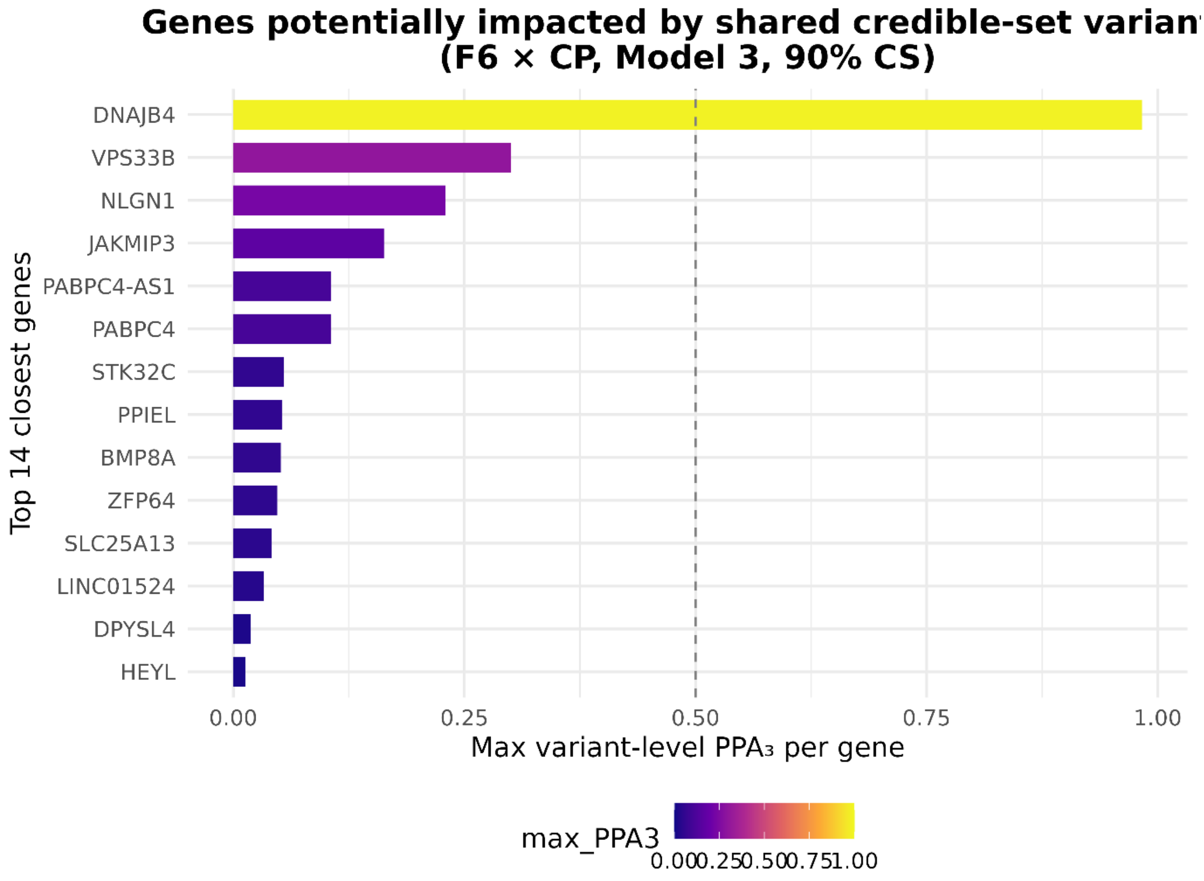

*GF X CP*

Supplementary Figure 77. Shared 90% credible-set variants and gene-level prioritisation for General Frailty × chronic pain.

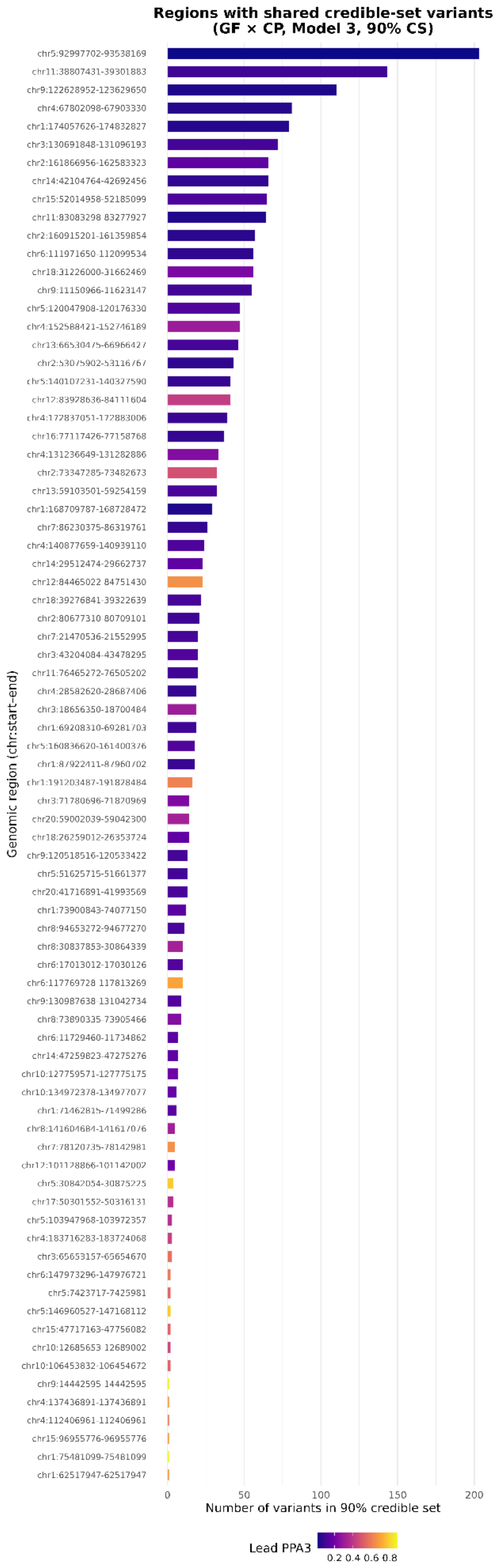
Regions with shared credible-set variants
(GF × CP, Model 3, 90% CS)
Genomic region (chr:start–end)
chr5:92997702-93538169
chr11:38807431-39301883
chr9:122628952-123629650
chr4:67802098-67903330
chr1:174057626-174832827
chr3:130691848-131096193
chr2:161866956-162583323
chr14:42104764-42692456
chr15:52014958-52185099
chr11:83083298 83277927
chr2:160915201-161359854
chr6:111971650-112099534
chr18:31226000-31662469
chr9:11150966-11623147
chr5:120047908-120176330
chr4:152588421-152746189
chr13:66530475-66966427
chr2:53075902-53116767
chr5:140107231-140327590
chr12:83928636-84111604
chr4:172837051-172883006
chr16:77117426-77158768
chr4:131236649-131282886
chr2:73347285-73482673
chr13:59103501-59254159
chr1:168709787-168728472
chr7:86230375-86319761
chr4:140877659-140939110
chr14:29512474-29662737
chr12:84465022 84751430
chr18:39276841-39322639
chr2:80677310 80709101
chr7:21470536-21552995
chr3:43204084-43478295
chr11:76465272-76505202
chr4:28582620-28687406
chr3:18656350-18700484
chr1:69208310-69281703
chr5:160836620-161400376
chr1:87922411-87960702
chr1:191203487-191828484
chr3:71780696-71820969
chr20:59002039-59042300
chr18:26259012-26353724
chr9:120518516-120533422
chr5:51625715-51661377
chr20:41716891-41993569
chr1:73900843-74077150
chr8:94653272-94677270
chr8:30837853-30864339
chr6:17013012-17030126
chr6:117769728 117813269
chr9:130987638-131042734
chr8:73890335-73905466
chr6:11729460-11734862
chr14:47259823-47275276
chr10:127759571-127775175
chr10:134972378-134977077
chr1:71462815-71499286
chr8:141604684-141617076
chr7:78120735-78142981
chr12:101128866-101142002
chr5:30842054-30875225
chr17:50301552-50316131
chr5:103947968-103972357
chr4:183716283-183724068
chr3:65653157-65654670
chr6:147973296-147976721
chr5:7423717-7425981
chr5:146960527-147168112
chr15:47717163-47756082
chr10:12685653-12689002
chr10:106453832-106454672
chr9:14442595-14442595
chr4:137436891-137436891
chr4:112406961-112406961
chr15:96955776-96955776
chr1:75481099-75481099
chr1:62517947-62517947
0
50
100
150
200
Number of variants in 90% credible set
Lead PPA3
0.2 0.4 0.6 0.8

References

1. Taylor AM, Pattie A, Deary IJ. Cohort Profile Update: The Lothian Birth Cohorts of 1921 and 1936. International Journal of Epidemiology. 2018;47(4):1042-r.
2. Houlihan, L. M. et al. Common Variants of Large Effect in F12, KNG1, and HRG Are Associated with Activated Partial Thromboplastin Time. Am. J. Hum. Genet. 86, 626–631. https://doi.org/10.1016/j.ajhg.2010.02.016 (2010).
3. Gadd DA, Hillary RF, McCartney DL, Zaghlool SB, Stevenson AJ, Cheng Y, et al. Epigenetic scores for the circulating proteome as tools for disease prediction. eLife. 2022;11:e71802.
4. Shah S, McRae AF, Marioni RE, Harris SE, Gibson J, Henders AK, et al. Genetic and environmental exposures constrain epigenetic drift over the human life course. Genome Res. 2014;24(11):1725-33.
5. Zhang Q, Marioni RE, Robinson MR, Higham J, Sproul D, Wray NR, et al. Genotype effects contribute to variation in longitudinal methylome patterns in older people. Genome Med. 2018;10(1):75.
6. Lupton MK, Robinson GA, Adam RJ, Rose S, Byrne GJ, Salvado O, et al. A prospective cohort study of prodromal Alzheimer's disease: Prospective Imaging Study of Ageing: Genes, Brain and Behaviour (PISA). NeuroImage: Clinical. 2021;29:102527.
7. Cuellar-Partida G, Springelkamp H, Lucas SEM, Yazar S, Hewitt AW, Iglesias AI, et al. WNT10A exonic variant increases the risk of keratoconus by decreasing corneal thickness. Human Molecular Genetics. 2015;24(17):5060-8.
8. Medland SE, Nyholt DR, Painter JN, McEvoy BP, McRae AF, Zhu G, et al. Common Variants in the Trichohyalin Gene Are Associated with Straight Hair in Europeans. The American Journal of Human Genetics. 2009;85(5):750-5.
9. McCarthy S, Das S, Kretzschmar W, Delaneau O, Wood AR, Teumer A, et al. A reference panel of 64,976 haplotypes for genotype imputation. Nature Genetics. 2016;48(10):1279-83.
10. Min JL, Hemani G, Davey Smith G, Relton C, Suderman M. Meffil: efficient normalization and analysis of very large DNA methylation datasets. Bioinformatics. 2018 Dec 1;34(23):3983-9.
